# The Coupled Stochastic Dynamical System: A Generative Model for Simulating and Forecasting Youth Mental Health Trajectories

**DOI:** 10.64898/2026.07.26.26358943

**Authors:** Nikolaos Koutsouleris, Georgia Turner, David J. Grüning, Nora Penzel, Grace Jacobs, Julia Fietz, Madalina Buciuman, Maria-Fernanda Urquijo, Sergio Mena, Charlotte Fraza, Paris Lalousis, Linda A. Antonucci, Silvia Schneider, Huifang Wang, Viktor Jirsa, Petr Slovak, Amy Orben

## Abstract

Predicting individual mental-health trajectories requires models that capture cross-domain dynamics rather than only statistical associations. We introduce the Coupled Stochastic Dynamical System (CSDS), a forward generative model of interacting digital behaviour, wearable-derived physiology and psychiatric burden. In the multi-year GLOBEM student cohort (N=496), Bayesian inversion recovered four behavioural phenotypes, reproducing 80–83% of cluster targets through distinct trait-vulnerability, physical-activity and affective-response parameter combinations. These phenotypes were primarily differentiated by digital-media overuse, physical activity, sleep and affective burden. Participant-specific inversion then generated virtual twins whose synthetic trajectories preserved observed behavioural traces: hidden Markov models trained on virtual-twin data reproduced state occupancy and transition patterns (*r*=0.84–0.87 and 0.93–0.94). Forecasts outperformed causal mean and persistence baselines for short-horizon magnitude prediction and predicted affective-state changes with 66–78% balanced accuracy over 21 days. These findings establish CSDS as an interpretable framework for simulating individual behavioural dynamics and advancing personalised mental-health modelling.

## 1 Introduction

Mental disorders emerge through the interplay of psychological and neurobiological vulnerabilities, psychiatric state dynamics, daily behavioural patterns, and, increasingly, digital environments^1,2^. Understanding the underlying complex forces through which these trajectories unfold would create new opportunities for the early recognition of mental disorders and for identifying therapeutic windows at which preventive action is most effective. It would also enable interventions that target an individual’s trajectory rather than relying on empirical group-level factors.

Mobile sensing studies have shown that smartphone- and wearable-derived signals carry rich longitudinal information about mental-health trajectories in young people, making continuous and unobtrusive monitoring technically feasible.^3–5^ Machine learning methods have proven well-suited to extract predictive patterns from this data, handling the scale, multimodality, and high temporal resolution of passively sensed time series more flexibly than classical univariate approaches.^6–9^ These methods have demonstrated clinically meaningful predictive accuracy for depressive episodes and symptom severity across independent cohorts.^10^ However, prevailing machine-learning approaches treat smartphone and wearable-derived signals as inputs without modelling the generative process through which digital engagement, psychological states, and psychomotor-vegetative patterns co-evolve. Therefore, a classifier that predicts depressive states from screen time cannot identify which modifiable behavioural dimension is driving these states at a given time point, nor can it simulate the effects of a proposed intervention before delivery.

Generative and dynamical modelling approaches may overcome these limitations by offering more interpretable accounts of mental-health trajectories (Box 1). In computational psychiatry, generative models infer latent system states and person-specific parameters from observed data, supporting mechanistically informed prediction and stratification.^11–13^ Network approaches similarly conceive mental disorders as interacting symptom dynamics, observable using EMA and time-series models.^14–16^ EMA repeatedly samples symptoms, affect and context in daily life, enabling within-person analyses of symptom fluctuations. Autoregressive models and early-warning indicators such as rising autocorrelation and variance have been used to characterise symptom coupling and critical slowing down before transitions into maladaptive or syndromal states.^17,18^

### Box 1 Key concepts in computational modelling of mental-health dynamics

#### Generative model

A model that specifies how observable data could arise from underlying processes and their interactions. Rather than only predicting an outcome from measured variables, a generative model can simulate new trajectories from a given set of model parameters and starting conditions.

#### Mechanistic model

Here, *mechanistic* means that the model explicitly represents hypothesised components and interactions, such as effects linking sleep, digital behaviour, affect and psychiatric burden, rather than allowing their relationships to be determined solely from statistical associations in the data. These mechanisms are theory-encoded hypotheses and should not be interpreted as uniquely established causal mechanisms.

#### State dynamics and state space

A person’s *state* describes their condition at a particular time across relevant dimensions such as affect, behaviour and physiology. The *state space* is the set of possible configurations of these variables, while *state dynamics* describe how an individual moves through this space over time.

#### Attractor and attractor landscape

An *attractor* is a region of state space towards which a dynamical system tends to return after disturbances. In mental health, relatively persistent configurations of symptoms, behaviour and physiology can therefore be conceptualised as attractor states. The *attractor landscape* describes the relative locations and stability of these states and the possible transitions between them.

#### Stochastic dynamics

Human trajectories are governed by deterministic and stochastic forces. In a stochastic dynamical model, systematic tendencies such as persistence, recovery and feedback interact with random fluctuations and unpredictable environmental influences. Repeated simulations from the same person-specific model can therefore produce different but statistically related trajectories.

#### Model inversion

Forward simulation maps model parameters onto predicted observations. *Model inversion* reverses this process: observed data are used to identify parameter configurations capable of reproducing salient features of an individual’s or group’s empirical trajectory. In the present study, inversion identifies ensembles of compatible configurations rather than a uniquely correct parameter set.

#### Virtual twin

A virtual twin is an individualised instance of the generative model whose parameters have been constrained by observations from a particular person. It is not a literal replica of that person, but a model capable of generating trajectories that reproduce selected characteristics of their observed behaviour, mental state and physiology.

#### Counterfactual simulation

Once a virtual twin has been established, selected model inputs or mechanisms can be altered to ask *what might happen under a different scenario –*for example, with changed sleep timing or reduced night-time digital use. Such simulations are conditional on the model and its encoded assumptions; prospective intervention studies are required before their differences can be interpreted as causal treatment effects.

This dynamical-systems perspective has been formalised as an attractor landscape in which psychiatric states are sustained by self-reinforcing feedback loops.^16^ Nonlinear state-space models have reconstructed attractor geometry directly from longitudinal EMA data, while control-theoretic approaches have begun to formalise how inferred dynamics can guide ecological momentary interventions.^19,20^ Despite these advances, data-driven generative approaches retain a structural limitation. They are fitted to symptom data alone, ignoring the digital media behaviours and neurodevelopmental processes that shape mental-health trajectories in adolescence and young adulthood. They also usually reconstruct dynamics without encoding the causal assumptions needed for counterfactual simulation.^19^ Counterfactual simulation embedded in virtual twin designs, i.e., individualised parameterisations of a mechanistic simulator, is a promising route towards personalised interventions: it enables explicit questions about what might happen if e.g. sleep timing changed, if a specific digital behaviour were reduced, or if an intervention were delivered at a different point in the given individual’s evolving attractor landscape.^21–23^

We therefore introduce the Coupled Stochastic Dynamical System (CSDS), a mechanistic forward generative model that simulates the co-evolution of digital engagement, EMA-recorded psychological and psychiatric state trajectories as well as wearable physiology. The CSDS maps clinical, behavioural and neurobiological knowledge onto a set of coupled stochastic oscillatory systems in which identified entities, such as traits, latent syndrome loads, behavioural and physiological channels, interact through prespecified activities, organised so as to produce regular change from a person’s set-up conditions to their observed trajectories.^24^ Because these cross-domain couplings are fixed a priori from established evidence rather than discovered post hoc, the resulting parameters retain a correspondence to putative processes rather than describing an unconstrained fit. Building on the CSDS’ outputs, decoding and forecasting methods such as Hidden Markov Models (HMM) translate inferred attractor states into interpretable, individualised state-space representations supporting risk prediction over days to weeks, and thus tailored interventions.

We tested whether such a theory-encoded generative system can be falsified against real-world longitudinal data by asking whether (a) Bayesian model inversion can recover parameter configurations that reproduce the empirical signatures of distinct behavioural risk profiles, (b) those configurations align with mechanistic accounts derived from clinical and psychometric theory, and (c) model-generated synthetic physiological, psychological and digital behaviour traces can support – alone or combined with observed data – short-term forecasting of clinically relevant state transitions. We first derived multimodal behavioural subtypes from the multi-year GLOBEM student cohort,^5^ which integrates passive sensing, ecological momentary assessment, and survey data. We evaluated whether the CSDS would be able to reproduce observed subtypes using model inversion and tested whether recovered posterior model coefficients could point to distinct behavioural mechanisms underlying the GLOBEM subtypes. Then, we performed individualised model inversion to generate virtual twins for each study participant and tested whether HMM trained on the twins’ synthetic data traces would allow decoding reproducible behavioural states in the observed data. Finally, we assessed the decoders’ performance in predicting short-term behavioural trajectories by training them on synthetic or hybrid data.

## 2 Methods

We first specified the CSDS as a theory-informed generative model independently of the empirical validation data. We then evaluated the model using the GLOBEM cohort of university students (N=496), which combines passive smartphone sensing, wearable-derived measures, ecological momentary assessment and survey data.^5^ The analysis proceeded from empirical phenotype definition and cluster-level model inversion to participant-specific virtual-twin generation, followed by state decoding and out-of-sample forecasting. A CSDS design and analysis overview is provided in Supplementary Figure S1.

### 2.1 Design of the Coupled Stochastic Dynamical System (CSDS)

The CSDS is a multiscale generative model designed to simulate the evolution of multimodal signal patterns, comprising daily psychological and psychopathological experiences, digital behaviours including social media activity, gaming, or conversational AI use, and wearable physiology represented as sleep and daily steps, electrodermal activity, heart rate, and heart rate variability. Conceptually, the CSDS separates four levels of simulation: (i) stable person-level traits, (ii) slowly evolving day-level burden states, (iii) sub-hourly behavioural–physiological allocation processes, and (iv) noisy observation/emission mappings. The dynamics of change across these levels are defined by various mechanisms. At level (i), traits define vulnerability, habit and regulatory parameters. At levels (ii) and (iii), modelled environmental factors perturb dynamic states, while bounded attractor systems generate persistence, tipping, recovery and recurrence. Finally, at level (iv) observation modules convert latent state trajectories into EMA, digital and wearable outputs. Thus, the levels combine to constitute a *structured forward simulation* in which symptoms, behaviour and physiology interact through feedback loops, such as sleep debt increasing next-day vulnerability, nocturnal digital use altering sleep and activity, and psychiatric burden reshaping EMA dynamics and behavioural allocation. Parameters governing the generation of these multimodal signals are organised centrally in a shared library at the level of mechanism families, including trait and habit coordinates, wearable physiology, digital behaviours, psychiatric burden dynamics and EMA emission mappings. Supplementary Methods S1□S6 detail and formalise the outlined CSDS architecture.

The CSDS architecture emerged iteratively by combining long-standing clinical expertise of the first author with theoretical guidance from dimensional models of psychopathology, without exposure to the validation data at any stage. This approach follows the established practice in theory-driven computational psychiatry of using prior clinical and mechanistic knowledge to specify model structure before empirical model inversion (Box 1), with face validity evaluated against clinical phenomenology rather than held-out data.^12,25^ Development began with the core affective and cognitive EMA channels and the formulation of depressive, suicidal, and psychotic burden processes as Ornstein-Uhlenbeck (OU) dynamics coupled to digital behaviour and wearable physiology. The OU formulation was chosen because mean-reverting stochastic processes with trait-anchored set points and state-dependent inertia have well-established validity as models of psychiatric symptom trajectories in high-frequency longitudinal data.^26–28^ The EMA driver infrastructure comprises three parallel first-order autoregressive (AR(1)) latent processes — negative affect, positive affect, and cognitive load — grounded in dimensional models that identify valence and arousal as fundamental to affective experience.^29–31^ In the initial formulation, syndrome-specific burden influenced EMA responses indirectly via the shared AR(1) drivers that fed all channels. Once the CSDS was extended to further psychiatric and behavioural dimensions this indirect projection route became a bottleneck which could not reproduce rich syndrome-specific EMA signatures describing human psychopathological experience: channels indexing distinct syndromes covaried too strongly, and syndrome-typical responses remained anchored to concurrent changes in affective loading. We therefore added direct projections from specific syndrome loads onto the latent utilities of the EMA channels indexing those syndromes, based on syndrome-specific phenomenological descriptions from clinical literature.^14,32^ Each channel utility thus comprises a driver-mediated component and, where clinically warranted, a syndrome-specific component gated by the corresponding load, so that syndrome-typical responding could occur without a general rise in negative affect. This modelling approach is broadly consistent with the Hierarchical Taxonomy of Psychopathology (HiTOP) framework, in which common transdiagnostic liability coexists with more specific syndromal dimensions that carry independent variance.^32,33^ It also aligns with network-analytic findings that syndrome-specific symptom clusters maintain connectivity patterns not reducible to shared negative affect.^15,34^

#### Trait coordinates, transformations, and simulator-wide influence

Each simulated individual is characterised by a vector of trait coordinates in z score ranges [-2.5; +2.5] comprising psychological dimensions, such as neuroticism, self-regulation capacity, reward sensitivity, social need, and resilience, as well as psychiatric pre-dispositions, covering affective-spectrum (anhedonia, hopelessness, bipolarity) and psychosis-spectrum phenotypes (psychosis proneness, cognitive disturbances) (Supplementary Table S1.1a). These are supplemented by domain-specific digital habits, age-referenced physical activity and sleep need, as well as selected exposomal factors, including early-life adversity, peer threat, and cannabis use. For selected traits, additional nonlinear features are derived from the bounded, normalised coordinates to define clinically relevant excess, deficit, or mid-range sensitivity zones that gate syndrome-specific pathways. This reflects the observation that some vulnerability dimensions are not adequately captured by monotonic linear effects, because low and high trait levels can relate to different forms of psychopathology: for reward sensitivity, low responsivity has been linked to autism-spectrum social difficulties,^35^ whereas heightened reactivity has been associated with borderline psychopathology and affective instability;^36^ for self-regulation, low control has been linked to impulsive and externalising behaviours,^37^ whereas excessive control has been associated with rigidity, perfectionism, and compulsive personality features (Supplementary Table S1.1b).^38,39^

**Exposomal processes** are generated as a set of stochastic drivers representing the environmental context of each simulated day (Supplementary Methods S4.1). To keep complexity manageable, we chose to model three key exposomal drivers known to impact youth mental health: (1) early-life adversity was encoded as a person-level constant contributing a stable allostatic load, aligned with evidence that adverse childhood experiences can become biologically embedded, with enduring effects on stress-regulatory systems.^40,41^ Its downstream influence on psychopathology was moderated by psychological vulnerability traits such as neuroticism, known to amplify the impact of stress on anxiety- and depression-related outcomes;^42^ (2) peer victimisation was generated as a daily stochastic event process whose base probability depended on school/work attendance, and whose intensity was amplified by psychosis burden through a perception-salience bias term, consistent with evidence for threat hypersensitivity in paranoid ideation;^43^ (3) Cannabis exposure was modelled as a stochastic daily use event whose probability depended on cannabis habit strength, current depressive and externalising load, and early-life adversity.^44,45^ Acute exposure was then converted into a dose-potency-weighted salience signal that fed into psychiatric pathways, most prominently psychosis, mania, and externalising reactivity.^46,47^ A composite allostatic load integrates early-life adversity, persistent peer threat, and cannabis salience as a slow OU process that feeds directly into psychiatric burden updating.

**Day-level psychological and psychiatric processes** comprise depression, suicidality, externalising, psychotic, obsessive-compulsive and eating-disorder loads as bounded stochastic attractor systems with trait-linked plateaus, context-sensitive forcing, and syndrome-specific regulation. For depression, psychosis, OCD and eating disorder, hysteretic on/off thresholds generate persistence and asymmetric recovery consistent with stability-landscape accounts of psychopathology.^16^ We model brief psychotic bursts as arising from a common daily hazard onto which threshold proximity, exposomal stress, and psychosis-proneness converge. Once the tipping point is reached, a burst adds a bounded transient amplitude over 2–5 days followed by mild kindling, capturing brief limited intermittent psychotic symptoms (BLIPS) in ultra-high-risk presentations (Supplementary Methods S3.3).^48,49^ Suicidal bursts are modelled analogously as short-lived crisis excursions, consistent with evidence that suicidal thinking can fluctuate markedly over hours and may intensify near tipping points.^50,51^ Bipolar activation is handled separately by a manic oscillator with sleep-irregularity forcing, episode-duration decay, and recurrence-dependent kindling that lowers the entry threshold across episodes.^52^ Mixed affective burden is derived as an interaction term of current manic and depressive load, consistent with the conceptualisation of mixed states as concurrent opposite-pole activation.^53^ Emotional-instability dynamics are represented separately by a signed borderline personality mood-pole oscillator, reflecting the rapid switching and affective-valence instability characteristic of this syndrome.^54^ Fast residual channels, including threat, social evaluation, perseveration, appearance and addictive cue salience, are computed from prior-day EMA deviations after subtraction of a global distress proxy and feed selectively into the slower syndrome pathways (Supplementary Methods S2 and S4.2).

EMA observations are then generated at six prompts per day from the within-day AR(1) latent processes indexing negative affect, cognitive load and positive affect (Supplementary Methods S3.5, S3.7 and S4.5). EMS signals’ inertia and volatility are modulated by resilience, syndrome state and instability profile,^17,31^ together with additive direct syndrome-, trait- and item-specific terms that preserve variance not explained by the shared latent substrate.^15^ The integration of stable trait coordinates with latent affective, cognitive and syndrome-specific processes yields distinguishable temporal signatures, including higher inertia and lower volatility in depression, very high persistence in OCD, and lower inertia with higher volatility in emotionally unstable profiles.^55,56^

#### Behavioural–physiological time-budget generation

Behavioural activity and wearable physiology are generated as a coupled, lagged, sub-hourly time budget system (Supplementary Methods S3.6, S4.3). The wearable module first proposes a physiologically plausible planned sleep schedule from a two-force competition between sleep need and wake drive: sleep need combines an age-dependent normative target, a person-specific sleep-need trait and prior sleep debt, with curfew and intervention effects modifying sleep opportunity; realised physical activity subsequently feeds back into sleep duration and quality. Wake drive integrates low self-regulation, reward sensitivity, neuroticism, entrenched digital habits, strain, manic activation, borderline personality-related arousal, externalising load, weekend structure and excess prior-night digital use. Depressive load enters this competition bidirectionally: anhedonic withdrawal increases sleep pull, whereas an anhedonia-gated endogenous depression factor shortens sleep duration and quality, consistent with altered sleep architecture, reduced rapid eye movement (REM) latency and early-morning-awakening physiology in depressive disorders.^57–59^ Mania shifts sleep phase and increases sleep variability, consistent with circadian phase disturbance in bipolar disorder,^60,61^ whereas cannabis withdrawal contributes to insomnia-like sleep-onset delay.^62,63^ The behavioural module then treats this wearable-derived schedule as the planned sleep constraint, but can modify realised sleep through per-night digital binge episodes. These episodes delay realised sleep onset, open late-night wake bins and inject nocturnal digital volume; their effect on sleep loss depends on next-day anchoring, so weak-anchor days can partly preserve sleep duration through delayed wake-up. Realised sleep duration, sleep debt, nocturnal digital use and total platform exposure are then written forward to shape next-day sleep pressure, attendance, physical activity, physiological dysrhythmia and behavioural allocation. Thus, sleep is both a physiological state variable and a behavioural time-budget constraint (Supplementary Methods S4.3, S4.4).

#### Hierarchical behavioural allocation and wearable signal generation

Within each five-minute interval, behaviour is allocated under a closed time-budget constraint (Supplementary Methods S3.6, S4.4): sleep defines the unavailable part of the bin, and the remaining wake budget is partitioned across residual “other” activity, in-person activity and total digital engagement. Structured school or weekday obligations and a minimum “other”-activity floor first reserve part of this wake budget; digital and in-person claims then compete over the residual budget, with school attendance represented as a sticky regime informed by clinical models of absenteeism and school refusal.^64^ In-person behaviour is modelled as an active social claim, while total digital engagement is influenced by habit-gated reward, loneliness, strain, sleep pressure, age-modulated self-regulation,^65^ and problematic-internet-use mechanisms such as compulsive use and online affect regulation.^66^ Additional clinical-state terms capture depressive withdrawal or passive digital escape, psychosis-related checking, OCD-related stereotypy, manic or borderline activation, and borderline interpersonal-affective instability.^67^ Within the allocated digital budget, time is probabilistically distributed across social media, gaming and chatbot use based on habitual preferences, with cross-domain inhibition, state-dependent stochasticity and switching noise when channel preferences are weakly differentiated.

Wearable signals are generated from the same latent and behavioural state space (Supplementary Methods S4.3): physical activity (steps) follows a state-modulated autoregressive activity process with burden-, mania-, eating-disorder- and lagged digital-displacement effects; HRV, heart rate and electrodermal activity are generated from baselines influenced by traits and modified by sleep debt, strain, psychiatric load, physical activity and dysrhythmic state, with manic, externalising and borderline arousal contributing to sympathetic and electrodermal bursts.^67,68^ A hysteretic physiological dysrhythmia oscillator integrates sleep debt, nocturnal use, digital dysrhythmia, psychiatric load, regulatory vulnerability, previous-day platform/chat exposure and in-person protection into phase diffusion and circadian amplitude collapse. The combined module emits in-person, digital and residual other activity, as well as realised sleep, HRV, heart rate and electrodermal activity (Supplementary Methods S4.4).

### 2.2 Derivation of empirical phenotype targets and CSDS validation

The timeseries of the GLOBEM sample (N=496) were converted into CSDS-compatible phenotype summaries using a predeclared mapping procedure (Supplementary Methods S7). Daily digital and wearable measures and EMA assessments were transformed to the corresponding simulator scales, including total and night-time platform use, sleep duration, steps, stress, positive affect, and depression burden. Participants were assigned to four clusters by k-means clustering of 24 standardised CSDS-facing level and longitudinal features (Supplementary Methods S8). The number of clusters was determined empirically through clinically meaningful interpretation which distinguished between heavy digital users with low physical activity, reduced sleep, but compensated mental health parameters (CL1); heavy digital users with increased depressive burden, low physical activity and poor sleep (CL2); individuals with relatively social media use, good physical activity and sleep (CL3); and an affectively burdened group with preserved sleep and physical activity and relatively low smartphone use (CL4).

For each cluster, empirical target specifications were generated directly from the participant-level phenotype distributions (Supplementary Methods S9). Specifications consisted of (a) *hard targets* defining admissible ranges that simulated phenotypes were required to reproduce during model inversion: CSDS parameter configurations generating values outside these ranges were treated as target violations and were progressively excluded from the accepted particle ensemble; (b) *soft targets* contributing continuously to the goodness-of-fit score without acting as strict admissibility criteria. Hard acceptance corridors were defined for seven directly mapped level phenotypes: total platform use, night-time use, sleep duration, daily steps, stress, positive affect, and depression burden. Weighted soft targets represented within-participant variability, threshold probabilities, selected digital autocorrelations, an approximate rumination proxy, and the between-participant dispersion and lower- and upper-tail structure of key EMA and behavioural measures. Target specification combined empirical cluster distributions with target-specific proportional bands and admissible bounds; correlation-type targets were defined in Fisher-(z) space. Because mania, obsessive-compulsive symptoms and psychotic symptoms were not assessed in GLOBEM and high prevalence of these symptoms cannot be expected in a non-clinical student cohort,^69,70^ we additionally specified low-burden reference targets for these channels across all clusters. Simulations producing greater burden on these unmeasured dimensions received a poorer fit to the target profile, but were not automatically excluded during model inversion. Similarly, although suicidal ideation is assessed as part of the BDI-II, the respective item-level responses were not available in the released data. Therefore, no suicidality target could be specified. The low suicidality constraint was relaxed for Clusters 2 and 4, whose depressive and hopelessness burden was substantially elevated. Retaining low-burden specifications would have induced compensatory distortion of the depression- and hopelessness-related parameters in these clusters.

For the generation of subject-specific virtual-twins, target specifications were generated from each selected participant’s own observed summaries using participant-level uncertainty intervals. Mapping rules, target weights and corridor formulas are provided in the Supplementary Methods S9.

### 2.3 Cluster- and participant-level model inversion

We used Approximate Bayesian Computation with Sequential Monte Carlo-style (ABC-SMC) proposal refinement to invert the CSDS at two levels. First, cluster-level inversion identified parameter configurations that reproduced the empirical target specification of each GLOBEM cluster (Supplementary Methods S10.1). Second, participant-level inversion generated virtual twins whose simulated phenotypes matched the longitudinal target specification of an individual participant (Supplementary Methods S10.2). Because the implementation used sequential rejection with perturbation-based proposal refinement, the resulting particle clouds were interpreted as ensembles of empirically compatible parameter configurations rather than formally weighted posterior samples. Acceptance was determined solely by the target specification at every stage (see below), so proposal refinement affected sampling efficiency and coverage, but not which configurations were admissible.

For cluster-level inversion, the validator automatically constructed the search space from the CSDS parameter library by retaining parameters that were active in the relevant simulation pathways and eligible for fitting, while excluding fixed, intervention-specific, explicitly blocked, or inactive parameters. This resulted in 1644 parameters defining the search space for the subsequent ABC-SMC-guided model inversion. Stage 0 evaluated 7,500 Latin-hypercube proposals, with each proposal generating an 80-day simulated population of 30 individuals. A cloud of 50 particles was refined over four scheduled stages—Stage 0 and three subsequent proposal stages—followed by final hard-target evaluation. The initial acceptance threshold, *ε*_0_, was defined by the 70th percentile of the finite Stage-0 distances. At each subsequent stage, *ε* was set to the 30th percentile of the preceding particle cloud and constrained not to increase. New proposals were generated using a principal-component-based multivariate perturbation kernel.

Participant-specific virtual-twin inversion used the same target-based acceptance logic, but individual longitudinal target corridors and simulation durations matched to each participant’s observation period. For each participant, 10,000 Stage-0 candidates were evaluated, and a target cloud of 50 particles was refined over seven scheduled stages. The initial epsilon was again the 70th percentile of Stage-0 distances; subsequent stages used the 50th, 45th, 35th, 30th, and 25th percentiles of the preceding cloud, with the 25th percentile retained for the final refinement stage. Simulation averaging increased from 7 to 15 repetitions across stages. Accepted cluster- and participant-level particles from previous runs informed proposal generation through a transfer bank with decreasing influence during the first 3 stages, but all acceptance decisions remained based on full CSDS simulations evaluated against the current participant’s target specification.

Hard phenotype targets acted as admissibility constraints. Among candidates with similar hard-target violations, fit was differentiated using a continuous distance combining the largest corridor-normalized hard-target deviation with a weighted soft-target distance. Surrogate regressors using Support Vector Regression were used only to rank enlarged candidate pools before simulation and did not determine acceptance. Parent selection was additionally weighted toward sparsely populated regions of a learned phenotype embedding to maintain diversity in the contracting parameter space. Model fit was quantified from the final hard-target pass rate, weighted soft-target coverage and proximity, epsilon compression, parameter-space contraction, and the reduction in mean raw distance from the unselected Stage-0 proposals to the final particle cloud.

### 2.4 Parameter-space audit

To characterise the particle ensembles recovered by the four cluster-level inversions, the 200 retained particles (50 per cluster specification) were expanded to 2,200 and re-simulated with the CSDS particle auditor (Supplementary Methods S10.3). Mechanistic attractor signatures were extracted from each simulated trajectory, comprising symptom-anchor severity, volatility, variance, skewness, lag-1 autocorrelation, reversion and recovery-speed proxies, unpredictable residual fluctuation, modal structure, and cross-domain coupling slopes linking digital behaviour, sleep, activity and symptom-anchor change, together with digital-sensitivity summaries and exposure–symptom lag descriptors. Particle-level signatures were averaged across repetitions and compared across the four ensembles. This tested whether ensembles fitted to different phenotype corridors occupied separable regions of the simulator’s latent dynamical and coupling space, indicating that the CSDS reproduced each phenotype through distinct mechanistic configurations rather than through variation in observed means alone. In a complementary analysis, the raw θ-parameter values of the expanded particles were tested individually for association with ensemble membership, and parameters were ranked by effect size *η*² to identify which regions of the simulator’s parameter space differentiated the four ensembles.

### 2.5 HMM-based state decoding

We used CSDS virtual-twin simulations as a synthetic-trained latent-state model for decoding daily behavioural, EMA, and wearable-derived time series from the GLOBEM clusters (Supplementary Methods S11). In each cluster, participants were ranked by proximity to the cluster centroid and the 50 closest were retained, keeping participant-level inversion computationally tractable while ensuring that each subsample comprised prototypical phenotype representatives. For every participant, 20 repeated simulations were generated from each of the 50 accepted particles, yielding 1,000 participant-specific synthetic trajectories. Separate participant-specific decoders were fitted to these dense virtual-twin trajectories and subsequently applied to the corresponding observed time series. To place the synthetic training and observed application data in the same feature reference frame, channel means and standard deviations were estimated exclusively from the participant-specific dense synthetic training trajectories and applied to both synthetic and observed sequences. Observed data therefore contributed neither to feature-scaling estimation nor to fitting the latent-state models.

The decoder representation combined standardised channel levels with lagged changes. For dense synthetic trajectories and daily passive or wearable channels, changes were calculated relative to the preceding day. For irregularly sampled observed EMA channels, changes were calculated relative to the immediately preceding available assessment and therefore represented variable multi-day intervals. EMA and passive or digitally assessed channels received equal domain weights. Observed time points were retained on their original temporal grid and were not imputed or deleted solely because individual channels were missing. We applied prespecified availability criteria to ensure that state assignments were supported by sufficient contemporaneous information while retaining lower-frequency EMA measurements (Supplementary Methods S11.2). State assignment was permitted only when at least three channels met their availability thresholds within a causal seven-day rolling window. The required availability was 50% for the passive sensing channels and positive affect and 25% for low-frequency EMA channels. Missing channels within otherwise supported time points were marginalized in the likelihood. Unsupported time points retained missing state labels. As an additional temporal regularisation criterion, decoded state segments were required to persist for at least two days, suppressing isolated one-day assignments without imposing an explicit state-duration model.

State models used mixed emissions, with diagonal-Gaussian likelihoods for continuous level and change features and categorical likelihoods for configured ordinal EMA channels. We evaluated two participant-specific latent-state decoders (Supplementary Methods S11.3). The first was a conventional sticky HMM, in which additional prior mass on self-transitions encouraged temporal persistence. The second was a native sticky autoregressive hidden Markov model (AR-HMM), which additionally incorporated state-specific first-order autoregressive dependence within the observation model. In the native AR-HMM, autoregressive dynamics contributed directly to latent-state inference. This model was distinct from the conventional sticky HMM with a separate horizon-specific autoregressive forecasting head used subsequently in the forecasting analysis.

The number of latent states in each participant-specific HMM was selected from *K = 4*–8 by particle-grouped cross-validation. Participant-specific states were subsequently aligned within each cluster to an expandable registry using Spearman correlations between dense-simulation state fingerprints. States with a best-match correlation below *p = 0.50* were added as new registry entries (Supplementary Methods 11.4 and S11.5). Consequently, the final cluster-level registry could contain more states than any individual participant-specific HMM. Model selection assessed reproducibility of state occupancy, off-diagonal transition structure, and multivariate state-fingerprint geometry while requiring a minimum state occupancy of 5%. The smallest state solution with a reproducibility score within 0.02 of the best-performing candidates was selected. Participant-specific state solutions were subsequently aligned to an expandable registry-based taxonomy using correlations between dense-simulation state fingerprints, enabling corresponding behavioural states to be compared across participants.

We then assessed how well the latent-state structure learned from the synthetic virtual-twin trajectories transferred to the observed data (Supplementary Methods S11.6). Because synthetic and observed trajectories were not temporally synchronized, this comparison did not test point-by-point agreement. Instead, it quantified structural overlap between the synthetic reference model and the observed decoding at three levels. State-occupancy correlation measured whether the same latent states occurred with similar relative frequencies in synthetic and observed data. Transition correlation measured whether the observed sequence reproduced the synthetic pattern of persistence and switching between states, based on the correspondence of the estimated state-transition matrices. State-fingerprint correlation measured whether corresponding states retained similar multivariate behavioural, EMA, and wearable-channel profiles when identified in the observed data. Together, these metrics assessed whether the synthetic decoder recovered in the observed time series a comparable repertoire of states, a comparable temporal organization of state transitions, and comparable state-specific phenotypic signatures.

### 2.6 Forecasting

We evaluated whether CSDS-generated virtual-twin time series could support forecasting of future behavioural and symptom trajectories using a causal rolling-origin design (Supplementary Methods S12). For each participant, forecasts were generated repeatedly at successive time points during the observed time series. At each forecast time point (anchor), the models were provided only with data collected up to and including that day (the causal prefix). Subsequent observations were withheld and used only to evaluate predictions at 1, 3, 7, 14, and 21 days.

Models were evaluated under two training regimes. First, in the synthetic regime, all latent-state and forecasting parameters were estimated exclusively from virtual-twin-generated time series. The participant’s observed history up to the forecast time point, the causal observed history, then passed through their fixed synthetic model to estimate the probabilities of the participant occupying each latent behavioural state. These state probabilities were then used to generate channel-specific forecasts. For fixed models with autoregressive components, the most recent observed level and recent change were additionally used as the starting conditions for the forecast. Second, in the hybrid regime, the synthetic model and its latent-state identities were retained, but emission distributions, state-transition probabilities, and, where applicable, autoregressive parameters were updated from the observed data using regularised shrinkage estimates. All non-adaptation parameters and eligible forecast timepoints were held constant between synthetic and hybrid comparisons.

The forecasting performance of the three decoding architectures described above were compared against each other and versus the null model comparators which were the *causal prefix mean* defined as the mean of all finite observations for the participant and channel available up to and including the forecast origin, as well as the *persistence* model which predicted the target based the latest finite observed value. Performance was evaluated separately for each target channel and summarized across channels using unweighted, domain-balanced averages. The pooled total-variation coefficient 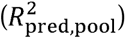 quantified prediction error relative to the total observed variation across all participants in a cluster and all eligible forecast time points. It therefore reflected both the models’ ability to distinguish participants with different average behavioural or symptom levels and their ability to reproduce temporal variation within participants. Furthermore, we used the within-subject skill to quantify how much the forecast reduces prediction error relative to the participant’s causal prefix-mean baseline, using the same eligible participant–channel–anchor observations at each horizon. Positive values indicate improvement over the baseline, zero equal performance, and negative values poorer performance.

Prediction of future change was assessed by comparing observed and predicted deviations from the participant- and channel-specific mean of the causal observed history available at each forecast time point. Directional accuracy quantified whether the predicted and observed deviations had the same sign, indicating agreement as to whether the future value was above or below the preceding individual mean. Systematic temporal tracking was additionally evaluated by comparing participant-specific slopes fitted to observed and predicted deviations across forecast time points. Percentile 95% confidence intervals were obtained using 5,000 subject-block bootstrap resamples, thereby retaining the dependence among repeated forecasts from the same participant.

Forecasting architectures, training regimes, and forecast horizons were compared separately for each target using factorial repeated-measures analyses of variance with the within-participant factors decoder architecture, training regime, and forecast horizon. Analyses were restricted to participants with complete data across the respective factorial design. Significant effects were decomposed using paired comparisons, with Holm correction for multiple testing.

The possibility of indirect information leakage between full-time series-derived target specifications used for virtual twin training and the downstream forecasting models trained on the synthetic data of these virtual twins was examined in samples of 20 participants per cluster selected for recording length and completeness (Supplementary Methods S10.4). Target corridors were derived independently from the complete observed time series and the first temporal half. To assess temporal stability between the half-series and full-series specifications we quantified the relative displacement between target corridors for hard and soft targets, separately, taking the weights of the latter into account at the levels of the full and the cluster-specific samples (Supplementary Table S10.4a). We also performed the hard target analysis by clusters x channels (Supplementary Table S10.4b).

To determine whether any downstream decoding or forecasting performance depended on information contained in the withheld second half of the observed series, we additionally performed a prospective split-half validation in clusters 1 and 2. Virtual twins were generated using only target specifications derived from the first temporal half of each participant’s data, while synthetic trajectories were simulated over the complete observed time span. The resulting split-half VTs were then subjected to the identical latent-state decoding and causal forecasting pipeline used for the corresponding full-series VTs. Comparisons were performed on matched participants using identical decoder settings, forecast horizons, target channels, quality-control criteria and eligible forecast observations, thereby isolating the effect of restricting virtual-twin calibration to the first half of the longitudinal series.

## 3 Results

### 3.1 GLOBEM clustering results

The four-cluster solution was organized across two principal biopsychosocial dimensions involving digital-media exposure, sleep and physical activity, as well as affective burden and psychosocial resources (Figures 1 and 2; Table 1). Specifically, total platform and night-time media use were elevated in Cluster 1 participants compared to Clusters 3 and 4, measuring 6.61 h/day and 2.62 h/day, respectively, while daily steps were low (5,343 steps). In contrast to Cluster 2, however, Cluster 1 showed substantially lower weekly stress, depression, trait anxiety, and perceived stress, together with higher positive affect, flourishing, mindfulness, resilience, and social support. In contrast, Cluster 2 participants showed highest overall affective and psychosocial burden amongst all clusters as measured by weekly stress (PSS-4) and depression (BDI-II, CES-D) scores, as well as higher trait anxiety, perceived stress, physical-symptom burden, loneliness, and neuroticism (Table 1). Concurrently, these individuals showed the lowest positive affect, flourishing, mindfulness, resilience, and social support. Total platform (6.00 h/day) and night-time media (2.51 h/day) were similar to Cluster 1, while physical activity was lowest (5,175 steps/day).

**Figure 1.**
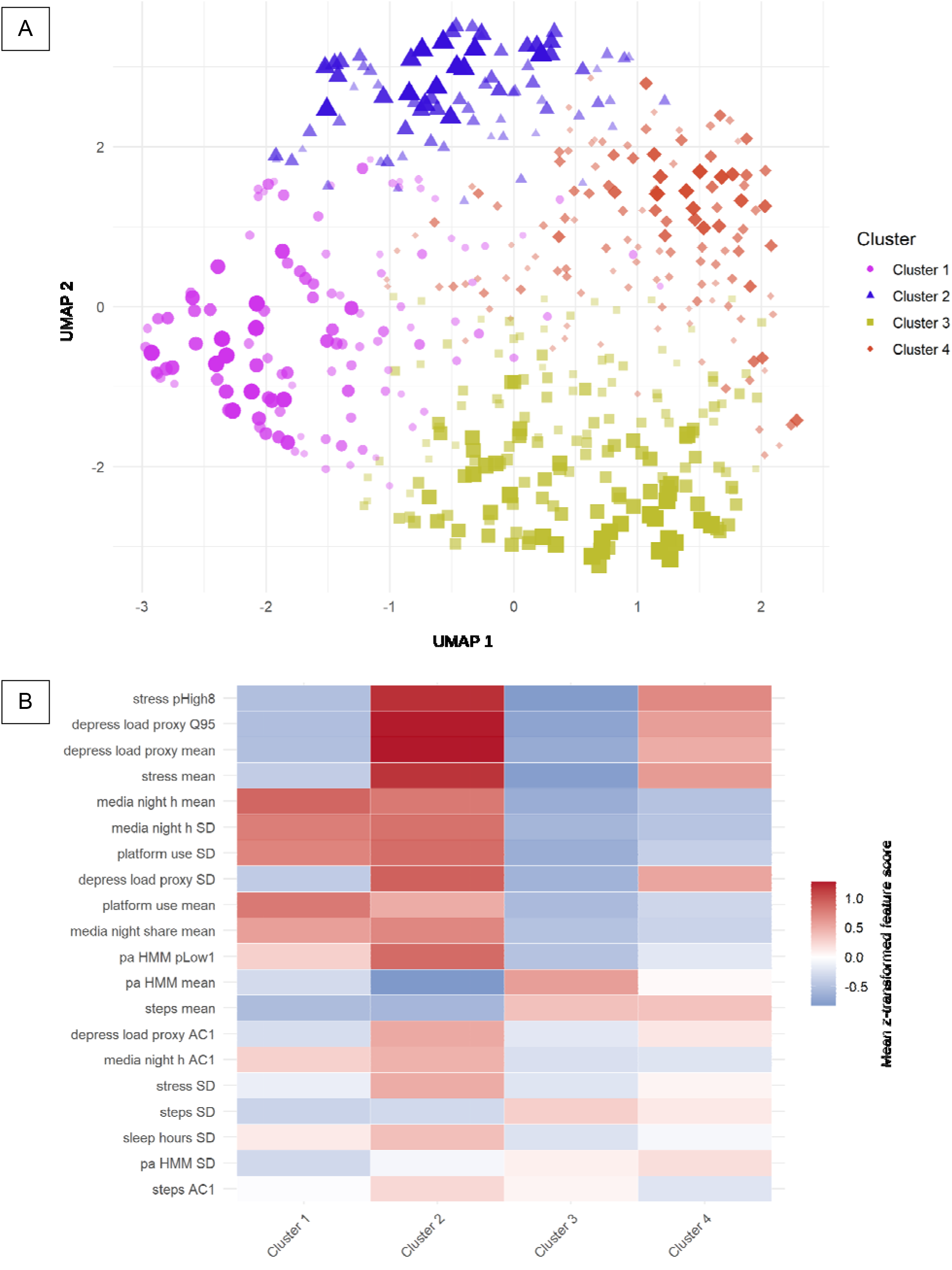
Four-cluster solution in the GLOBEM sample and cluster-specific multimodal feature profiles. **(A)** UMAP projection of the selected four-cluster solution. Each point represents one participant and is coloured/shaped by assigned cluster. Point size and opacity are scaled by the centroid margin, such that larger and more opaque points indicate participants whose assigned cluster is more clearly separated from alternative cluster assignments, whereas smaller and fainter points indicate subjects closer to cluster boundaries. The embedding shows a structured arrangement of the four clusters with limited central overlap and more compact peripheral cores. **(B)** Heatmap of the top varying standardized cluster-centre features across clusters. Columns represent clusters and rows represent features ranked by their between-cluster variation in cluster-center standardized values. Warmer colours indicate above-average z-score values and cooler colours indicate below-average values relative to the full sample. *Feature abbreviations*: *mean*, participant-level mean; *SD*, within-participant standard deviation; *AC1*, lag-1 autocorrelation; *Q95*, empirical 95th percentile; *pHigh8*, proportion of observations above the predefined high-threshold; *pLow1*, proportion of observations below the predefined low-threshold.

**Figure 2.**
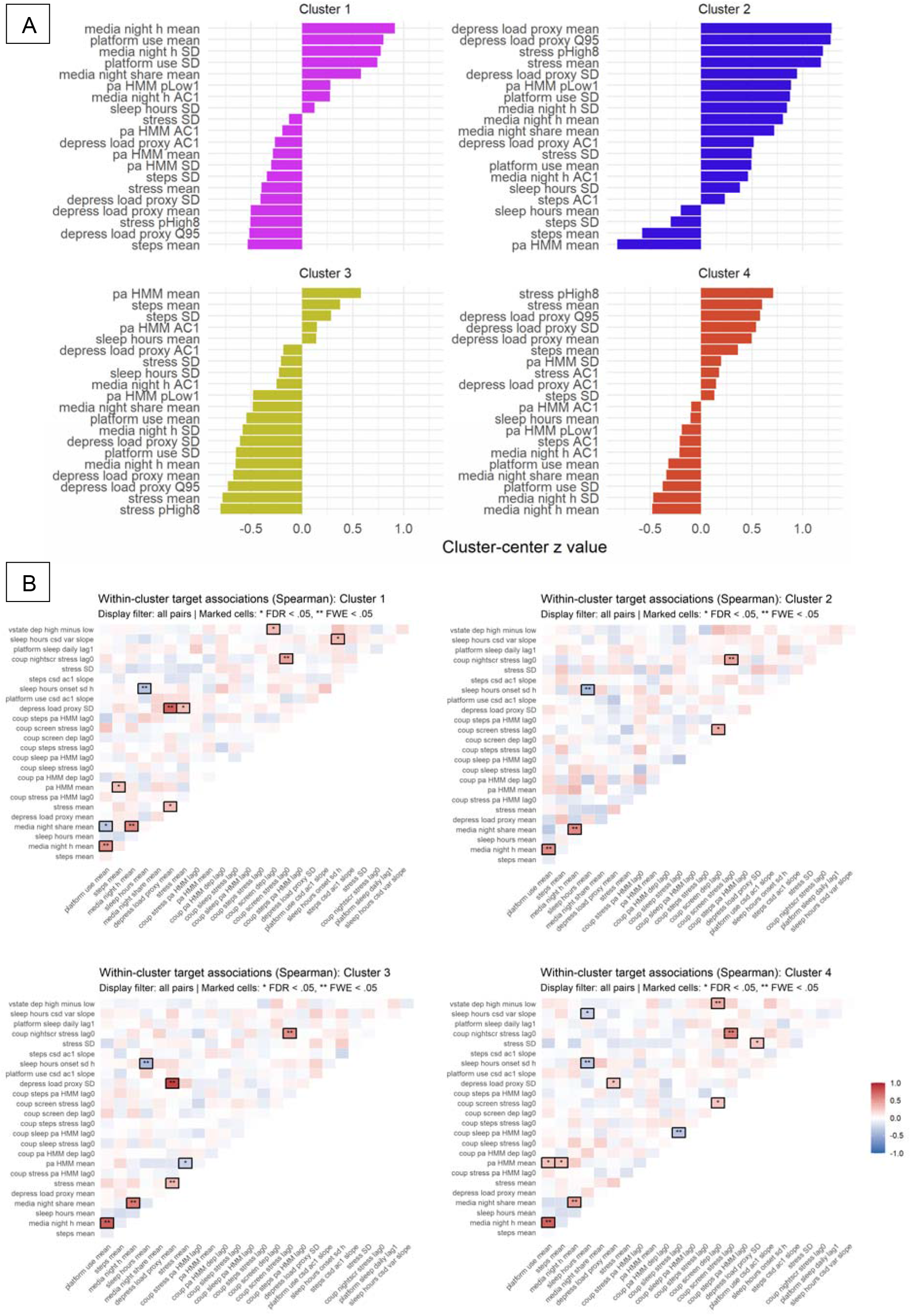
Top standardised feature deviations by cluster and within-cluster feature association maps. **(A)** Bar plots show the strongest positive and negative deviations of each cluster centroid from the overall sample mean, expressed as z values and ranked by absolute magnitude. These profiles highlight the dominant clinical, sleep, media-use, affective, and behavioural features characterising each cluster. **(B)** Within-cluster heat maps show pairwise Spearman correlations among target features; red indicates positive and blue negative associations. Marked cells denote statistically supported associations after multiple-comparison correction, with stronger marking indicating FWE-level significance. Together, the panels summarise both the marginal feature profile of each cluster and its internal cross-feature dependency structure.

**Table 1.** Cluster-wise descriptive profile of participants, summarising core digital-use, wearable-derived sleep and activity, weekly symptom, and baseline psychosocial measures. Data are shown as mean (SD) within each cluster, with cluster sample sizes reported in the first row. Between-cluster differences were assessed using one-way ANOVA tests, with Benjamini-Hochberg false discovery rate correction applied across the variables.

| Measure | Cluster 1 | Cluster 2 | Cluster 3 | Cluster 4 | df | F | P | P<sub>FDR</sub> |
| --- | --- | --- | --- | --- | --- | --- | --- | --- |
| N | 116 | 82 | 166 | 132 |  |  |  |  |
| Stress, PSS-4 weekly (0-16) | 5.32 (1.58) | 9.14 (1.85) | 4.39 (1.68) | 7.74 (1.29) | 3, 492 | 216.90 | <.0001 | <.0001 |
| Night-time media use (h/day) | 2.62 (1.01) | 2.51 (0.98) | 1.02 (0.45) | 1.20 (0.43) | 3, 491 | 168.93 | <.0001 | <.0001 |
| Platform use SD (h/day) | 2.39 (0.59) | 2.48 (0.66) | 1.47 (0.41) | 1.65 (0.38) | 3, 492 | 126.21 | <.0001 | <.0001 |
| Night-time media use SD (h/day) | 1.53 (0.58) | 1.58 (0.63) | 0.667 (0.393) | 0.741 (0.374) | 3, 491 | 124.54 | <.0001 | <.0001 |
| Depression, BDI-II (0-63) | 9.09 (6.01) | 22.41 (8.95) | 7.36 (5.17) | 15.86 (7.96) | 3, 481 | 105.02 | <.0001 | <.0001 |
| Depression, CES-D (0-30) | 6.74 (3.64) | 13.65 (4.66) | 5.46 (3.45) | 10.90 (4.42) | 3, 489 | 100.92 | <.0001 | <.0001 |
| Trait anxiety, STAI (20-80) | 40.83 (9.15) | 55.50 (9.04) | 38.64 (8.78) | 49.84 (9.15) | 3, 488 | 85.19 | <.0001 | <.0001 |
| Perceived stress, PSS (0-56) | 24.73 (6.29) | 34.00 (6.35) | 22.79 (6.74) | 30.81 (5.90) | 3,489 | 77.61 | <.0001 | <.0001 |
| Platform use (h/day) | 6.61 (2.01) | 6.00 (2.04) | 3.91 (1.32) | 4.36 (1.43) | 3,492 | 77.05 | <.0001 | <.0001 |
| Night-time media share (0-1) | 0.413 (0.168) | 0.433 (0.161) | 0.262 (0.079) | 0.282 (0.079) | 3,491 | 61.96 | <.0001 | <.0001 |
| Positive affect, PANAS-positive (0-20) | 5.68 (4.00) | 3.61 (2.62) | 9.03 (3.52) | 6.92 (2.93) | 3,492 | 53.16 | <.0001 | <.0001 |
| Daily steps | 5342.7 (3360.3) | 5174.6 (2530.9) | 8762.6 (3578.3) | 8723.6 (3564.2) | 3,492 | 42.09 | <.0001 | <.0001 |
| Flourishing, FS-PWB (8-56) | 44.51 (6.62) | 37.62 (9.07) | 46.72 (5.85) | 40.70 (7.30) | 3,489 | 37.79 | <.0001 | <.0001 |
| Mindfulness, MAAS (1-6) | 4.19 (0.83) | 3.39 (0.85) | 4.17 (0.79) | 3.51 (0.80) | 3,489 | 31.39 | <.0001 | <.0001 |
| Physical symptoms, CHIPS (0-132) | 13.68 (11.14) | 28.42 (19.11) | 12.21 (10.58) | 23.22 (18.28) | 3,485 | 30.75 | <.0001 | <.0001 |
| Loneliness, UCLA-10 (10-40) | 20.87 (4.73) | 24.94 (5.96) | 20.05 (5.02) | 24.30 (5.00) | 3,487 | 27.35 | <.0001 | <.0001 |
| Active bout duration (min) | 2.53 (0.69) | 2.54 (0.59) | 3.08 (0.67) | 3.05 (0.61) | 3,492 | 27.27 | <.0001 | <.0001 |
| Resilience, BRS (1-5) | 3.54 (0.74) | 2.89 (0.77) | 3.58 (0.67) | 3.05 (0.68) | 3,489 | 27.14 | <.0001 | <.0001 |
| Belonging, SocialFit (17-119) | 77.83 (7.20) | 73.12 (9.56) | 79.37 (7.02) | 74.34 (8.34) | 3,488 | 16.58 | <.0001 | <.0001 |
| Daily steps SD | 1510.9 (788.2) | 1549.3 (771.3) | 2036.0 (841.0) | 1907.7 (786.6) | 3,492 | 13.24 | <.0001 | <.0001 |
| Gratitude, GQ-6 (6-42) | 34.50 (5.07) | 32.90 (5.51) | 36.18 (4.33) | 33.46 (5.27) | 3,489 | 11.00 | <.0001 | <.0001 |
| Social support, 2waySSS (0-1) | 0.781 (0.142) | 0.717 (0.169) | 0.805 (0.124) | 0.725 (0.167) | 3,488 | 10.34 | <.0001 | <.0001 |
| Neuroticism, BFI-10 (1-10) | 6.25 (1.85) | 7.33 (2.01) | 6.02 (1.98) | 6.77 (1.88) | 3,488 | 9.95 | <.0001 | <.0001 |
| Conscientiousness, BFI-10 (1-10) | 6.83 (1.51) | 6.54 (1.69) | 7.55 (1.75) | 7.00 (1.81) | 3,488 | 7.99 | <.0001 | <.0001 |
| Sleep duration SD (h) | 0.791 (0.472) | 0.894 (0.394) | 0.651 (0.360) | 0.718 (0.351) | 3,492 | 7.87 | <.0001 | .0001 |
| Weekend routine similarity (0-1) | 0.709 (0.191) | 0.666 (0.196) | 0.631 (0.188) | 0.589 (0.179) | 3,434 | 7.72 | <.0001 | .0001 |
| Circadian routine similarity (0-1) | 0.689 (0.205) | 0.653 (0.202) | 0.612 (0.198) | 0.567 (0.185) | 3,434 | 7.46 | .0001 | .0001 |
| Wake time (min after midnight; 0-1440) | 547.9 (104.9) | 548.0 (103.8) | 510.3 (98.9) | 523.9 (74.3) | 3,492 | 4.87 | .0024 | .0028 |
| Reappraisal, ERQ (1-7) | 4.98 (0.86) | 4.71 (1.01) | 4.99 (0.93) | 4.65 (1.10) | 3,489 | 4.33 | .0050 | .0057 |
| Alcohol problems, BYAACQ (0-24) | 1.75 (2.61) | 2.77 (3.23) | 1.70 (2.57) | 2.98 (4.62) | 3,276 | 2.87 | .0366 | .0403 |
| Sleep duration (h) | 7.06 (0.91) | 6.83 (0.93) | 7.13 (0.82) | 6.92 (0.87) | 3,492 | 2.80 | .0395 | .0421 |
| Suppression, ERQ (1-7) | 4.08 (1.12) | 4.38 (1.25) | 4.10 (1.12) | 4.33 (1.14) | 3,489 | 2.07 | .1039 | .1071 |
| Bedtime (min after midnight; 0-1440) | 1425.9 (111.8) | 1404.3 (120.2) | 1413.4 (91.3) | 1426.5 (79.7) | 3,492 | 1.21 | .3056 | .3056 |

**Abbreviations:** 2waySSS, 2-Way Social Support Scale; BDI-II, Beck Depression Inventory-II; BFI-10, Big Five Inventory-10; BRS, Brief Resilience Scale; BYAACQ, Brief Young Adult Alcohol Consequences Questionnaire; CES-D, Center for Epidemiologic Studies Depression Scale; CHIPS, Cohen–Hoberman Inventory of Physical Symptoms; ERQ, Emotion Regulation Questionnaire; FS-PWB, Flourishing Scale / Psychological Well-Being Scale; GQ-6, Gratitude Questionnaire-6; MAAS, Mindful Attention Awareness Scale; PANAS-positive, positive affect subscale of the Positive and Negative Affect Schedule; PSS-4, Perceived Stress Scale-4 items version; SD, standard deviation; SocialFit, Sense of Social and Academic Fit Scale; STAI, State–Trait Anxiety Inventory for Adults; UCLA-10, 10-item short-form UCLA Loneliness Scale. Time-use variables are reported in hours per day where indicated; active bout duration is reported in minutes; bedtime and wake time are expressed as minutes after midnight; routine similarity and night-time media share are unitless indices scaled from 0 to 1. Instrument names and score ranges follow the GLOBEM v1.1 documentation.

Cluster 3 was characterized by the lowest stress, affective burden and digital exposure, and highest physical activity. Participants in this cluster showed the lowest trait anxiety, physical symptoms, loneliness, and neuroticism scores, together with the highest positive affect, flourishing, resilience, belonging, gratitude, social support, and conscientiousness. Platform use averaged 3.91 h/day, night-time media use 1.02 h/day, and daily steps 8,763. Cluster 3 also had the longest active bouts and the lowest sleep-duration variability. Cluster 4 combined moderate stress, loneliness, and affective burden with relatively low digital exposure and high physical activity. Positive affect and psychosocial-resource measures generally occupied an intermediate position. Platform use averaged 4.36 h/day and night-time media use 1.20 h/day, whereas daily steps were high at 8,724 and active-bout duration was similar to Cluster 3.

Sleep-duration differences were small in absolute terms, ranging from 6.83h in Cluster 2 to 7.13h in Cluster 3, despite clearer differences in sleep variability and temporal organization (Table 1). Clusters 1 and 2 showed substantially greater variability in platform and night-time media use than Clusters 3 and 4. Conversely, Clusters 3 and 4 showed longer active bouts, and greater day-to-day step variability. Weekend and circadian routine-similarity indices followed a different ordering, with the highest values in Cluster 1 and the lowest in Cluster 4.

The dynamics analysis identified a smaller set of cluster differences after multiple-comparison correction (Table 2). Bedtime variability was greatest in Cluster 2 and lowest in Cluster 3. Night-time media persistence was higher in Clusters 1 and 2 than in Clusters 3 and 4, while persistence of depression burden was highest in Cluster 2. Daily step counts were strongly persistent in all four clusters, with AC(1) values between 0.776 and 0.829; Cluster 4 showed the strongest day-of-week regularity, whereas Cluster 2 showed the weakest. Positive-affect persistence was highest in Clusters 2 and 3, while stress persistence was highest in Clusters 2 and 4. The daily-step critical-slowing-down slope also differed across clusters, although the absolute differences were small.

**Table 2.** Cluster-specific within- and cross-domain dynamics profiles based on CSDS-aligned features. The table summarizes temporal regularity, persistence, early-warning dynamics, and cross-domain coupling across sleep–circadian, digital-behavioural, activity, stress, affective, and depression-related signals. Within-domain dynamics include circular variability in bedtime, day-of-week regularity of steps, and first-order autocorrelation, AC(1). AC(1) quantifies short-term persistence from one observation to the next, with higher positive values indicating stronger carry-over. Critical-slowing-down indices are reported as slopes in AC(1) or variance over time, capturing whether signals become progressively more persistent or unstable. Cross-domain dynamics are expressed as lag-0 Pearson correlations between contemporaneous domains, for example stress–positive affect, sleep–stress, platform-use–depression, and night-time-media–stress coupling. The lagged metric, platform-use[t] → sleep[t+1], captures whether daily platform use is associated with next-day sleep variation. Positive affect is abbreviated as PA; all coupling coefficients are scaled from −1 to 1 unless otherwise indicated.

| Measure | Cluster 1 | Cluster 2 | Cluster 3 | Cluster 4 | df | F | P | P<sub>FDR</sub> |
| --- | --- | --- | --- | --- | --- | --- | --- | --- |
| Bedtime circular SD (h) | 1.72 (1.10) | 2.19 (1.08) | 1.39 (0.77) | 1.54 (0.69) | 3, 492 | 15.52 | .0000 | **.0000** |
| Night-time media use AC(1) [-1,1] | 0.207 (0.226) | 0.246 (0.229) | 0.097 (0.186) | 0.105 (0.178) | 3, 490 | 15.30 | .0000 | **.0000** |
| Depression-load proxy AC(1), weekly [-1,1] | -0.050 (0.306) | 0.221 (0.363) | -0.017 (0.345) | 0.096 (0.341) | 3, 417 | 11.72 | .0000 | **.0000** |
| Steps day-of-week regularity [0,1] | 0.028 (0.022) | 0.022 (0.025) | 0.027 (0.024) | 0.036 (0.029) | 3, 492 | 5.85 | .0006 | **.0031** |
| Daily steps AC(1) [-1,1] | 0.799 (0.116) | 0.829 (0.092) | 0.809 (0.116) | 0.776 (0.139) | 3, 492 | 3.72 | .0114 | **.0417** |
| Positive affect AC(1), weekly [-1,1] | 0.076 (0.280) | 0.166 (0.273) | 0.167 (0.256) | 0.103 (0.264) | 3, 488 | 3.60 | .0135 | **.0417** |
| Stress AC(1), weekly [-1,1] | 0.009 (0.339) | 0.099 (0.359) | -0.003 (0.341) | 0.110 (0.363) | 3, 485 | 3.58 | .0139 | **.0417** |
| Daily steps CSD AC(1) slope (per day) | -0.004 (0.006) | -0.002 (0.005) | -0.004 (0.006) | -0.004 (0.006) | 3, 492 | 3.48 | .0160 | **.0420** |
| Stress-PA coupling r, lag 0 [-1,1] | -0.138 (0.383) | -0.211 (0.390) | -0.079 (0.350) | -0.169 (0.366) | 3, 483 | 2.73 | .0435 | .1015 |
| Steps-PA coupling r, lag 0 [-1,1] | 0.021 (0.283) | 0.065 (0.320) | -0.009 (0.295) | 0.071 (0.298) | 3, 486 | 2.14 | .0943 | .1850 |
| Sleep-PA coupling r, lag 0 [-1,1] | 0.009 (0.272) | -0.009 (0.253) | 0.048 (0.268) | -0.031 (0.309) | 3, 486 | 2.12 | .0969 | .1850 |
| Platform-use/depression coupling r, lag 0 [-1,1] | -0.019 (0.367) | 0.010 (0.382) | 0.045 (0.328) | 0.083 (0.335) | 3, 476 | 1.90 | .1282 | .2244 |
| Night-time-media/stress coupling r, lag 0 [-1,1] | 0.003 (0.381) | -0.045 (0.408) | 0.041 (0.365) | -0.059 (0.407) | 3, 452 | 1.74 | .1584 | .2559 |
| PA-depression coupling r, lag 0 [-1,1] | -0.092 (0.408) | -0.182 (0.375) | -0.085 (0.356) | -0.129 (0.389) | 3, 424 | 1.27 | .2849 | .4274 |
| Platform use AC(1) [-1,1] | 0.177 (0.157) | 0.203 (0.164) | 0.170 (0.158) | 0.182 (0.178) | 3, 492 | 0.75 | .5233 | .6543 |
| Sleep-stress coupling r, lag 0 [-1,1] | -0.049 (0.381) | -0.089 (0.429) | -0.009 (0.375) | -0.033 (0.437) | 3, 486 | 0.75 | .5246 | .6543 |
| Steps-stress coupling r, lag 0 [-1,1] | -0.046 (0.423) | 0.034 (0.417) | -0.023 (0.377) | -0.039 (0.411) | 3, 486 | 0.74 | .5297 | .6543 |
| Platform-use/stress coupling r, lag 0 [-1,1] | 0.035 (0.330) | 0.008 (0.389) | 0.047 (0.355) | -0.003 (0.402) | 3, 489 | 0.54 | .6565 | .7659 |
| Platform use CSD AC(1) slope (per day) | -0.001 (0.005) | -0.001 (0.006) | -0.000 (0.005) | 0.000 (0.005) | 3, 491 | 0.46 | .7089 | .7835 |
| Sleep duration CSD variance slope [h<sup>2</sup> / day] | 0.009 (0.022) | 0.010 (0.027) | 0.007 (0.023) | 0.009 (0.024) | 3, 492 | 0.31 | .8194 | .8604 |
| Platform[t] -> sleep[t+1] daily coupling r [-1,1] | 0.001 (0.173) | -0.003 (0.177) | 0.002 (0.163) | -0.011 (0.162) | 3, 492 | 0.16 | .9242 | .9242 |

None of the tested cross-domain coupling measures remained significant after false-discovery-rate correction (Table 2). Figure 2B nevertheless showed that the distribution of corrected within-cluster feature associations differed across clusters, indicating cluster-specific covariance structures superimposed on the more prominent differences in mean levels and within-domain temporal dynamics.

### 3.2 Cluster-level model inversion

Inverting the CSDS against each of the four cluster specifications yielded particle clouds that reproduced the cluster-level phenotype targets with high fidelity (Table 3). Weighted target coverage ranged from 80.0% to 83.3% and target proximity from 91.3% to 94.6%, and the ABC-SMC threshold contracted substantially in each inversion process (*1 — ε*_final_ */ε₀* = 0.969–0.980). At the individual level, inversion of the 50 virtual-twin target specifications per cluster was less complete than cluster-level inversion. Mean target coverage ranged from 56.0% to 66.8%, and mean target proximity from 75.7% to 84.2%, with the highest values observed for Cluster 3.

**Table 3.** ABC-SMC process diagnostic describing the posterior fitting quality of the CSDS to the four cluster-level specifications and 50 single-subject target specification per cluster. Target coverage is the weighted percentage of target corridors reached by the accepted-particle summary; target proximity is the weighted mean closeness to target corridors, with full credit for values inside bounds and distance-based penalties outside bounds. Weighting used the soft target weights defined in cluster target specifications. ε-compression is defined as 1 — *ε_final_*/*ε*_0_, indicating relative tightening of the ABC threshold.

| **Metric** | **Cluster 1** | **Cluster 2** | **Cluster 3** | **Cluster 4** |
| --- | --- | --- | --- | --- |
| *Cluster-level ABC-SMC* |  |  |  |  |
| Target coverage [%] | 80.0 | 80.0 | 83.3 | 83.3 |
| Target proximity [%] | 91.3 | 94.1 | 94.6 | 94.2 |
| $1 - \epsilon_{final}/\epsilon_0$ ratio [0,1] | 0.969 | 0.979 | 0.972 | 0.980 |
| *Single-subject (VT-based) ABC-SMC* |  |  |  |  |
| Target coverage [mean% (SD)] | 59.3 (13.8) | 56.0 (12.3) | 66.8 (11.1) | 56.9 (10.0) |
| Target proximity [mean % (SD)] | 77.6 (11.5) | 75.7 (10.3) | 84.2 (8.9) | 77.1 (10.4) |
| $\epsilon_0/\epsilon_{final}$ ratio [0,1] [mean (SD)] | 0.785 (0.103) | 0.739 (0.101) | 0.802 (0.112) | 0.699 (0.144) |

Projection of the cluster-specific particle ensembles onto the first three principal components of the CSDS parameter space revealed a structured and partially separable organisation (Figure 3A). Separation was clearest for Cluster 2, whereas Clusters 1, 3, and 4 retained partial overlap. The first principal component (PC) explained 21.1% of the variance and was dominated by the severity and instability characteristics indexing affective dysregulation dynamics. PC1 primarily distinguished the high-burden Cluster 2 from the lower-burden Clusters 1 and 3, with Cluster 4 occupying an intermediate region. PC2 explained 5.8% of the variance and was associated with coupling between digital exposure, particularly night-time media and total platform use, and subsequent changes in affective and cognitive dysregulation and recovery processes. It differentiated the high-digital Clusters 1 and 2 from the lower-digital Clusters 3 and 4. PC3 explained a further 4.5% and represented a smaller mixed dimension involving mean night-time media use and dysthymic, psychotic-symptom, circadian, and depression-related persistence metrics.

**Figure 3.**
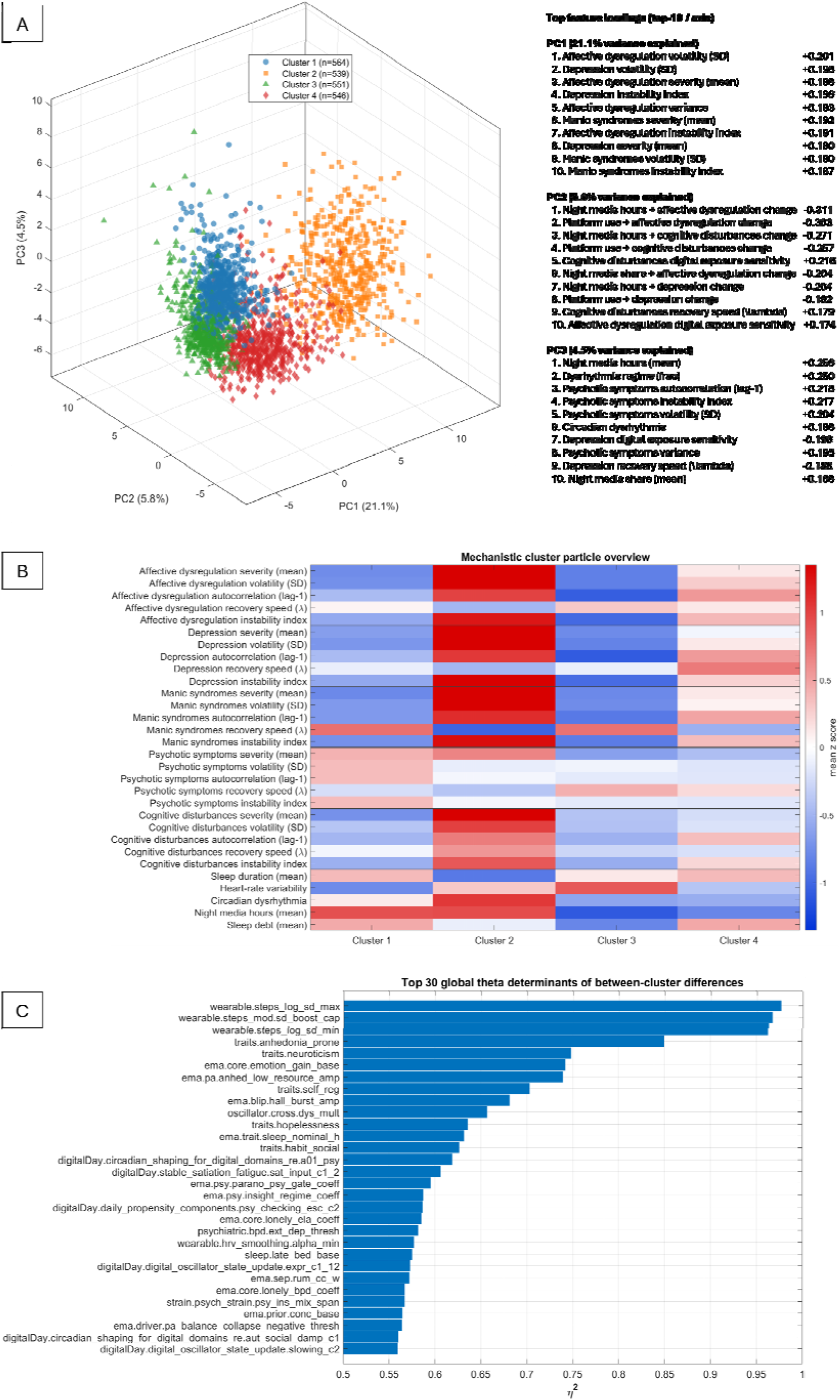
In CSDS feature space: PCA embedding and mechanistic differences of the particle clouds fitted to the four GLOBEM cluster specifications. **(A)** Three-dimensional PCA embedding of accepted particle clouds grown by means of barycentric expansion and coloured by inferred cluster phenotype, with the right-hand table listing the top feature loadings associated with PC1–PC3. **(B)** Cluster overview heat map showing standardised feature deviations across the four particle clusters, highlighting their attractor-based metrics that separate them in the CSDS feature space. **(C)** Global theta-determinant ranking, showing simulator parameters with the strongest overall association with particle cluster separation, quantified by *η*². Mechanistic roles of ranked parameters are explained in Supplementary Table S10.3.

The cluster-level mechanistic profiles supported these axis-level patterns (Figure 3B) and mirrored empirical cluster differences. Cluster 2 showed the strongest elevations across affective-dysregulation, depression, and manic-syndrome severity, volatility, autocorrelation, and instability metrics, together with high night-time media exposure. Cluster 3 showed the opposite configuration, while Cluster 4 displayed intermediate affective and depression-related values, with digital-exposure measures similarly expressed as Cluster 3. Cluster 1 was characterized by high night-time media exposure but comparatively low affective-dysregulation and depression-related severity and instability.

The global *θ*-parameter analysis identified three prominent groups of simulator parameters associated with particle-cluster separation (Figure 3C, Supplementary Table S10.3). The strongest effects involved the generation of day-to-day physical-activity variability. These included the lower and upper bounds on the effective log-scale variability of daily steps (wearable.steps_log_sd_min and wearable.steps_log_sd_max) and the cap on state-dependent volatility amplification (wearable.steps_mod.sd_boost_cap), which limits the combined increases in step variability arising from manic, mixed, psychotic, externalising, and other state-related mechanisms. These three parameters showed the largest cluster membership associations, with *η*^2^ ranging between 0.95 and 0.97.

The second group comprised baseline trait parameters, such as traits.anhedonia_prone, which encodes baseline susceptibility to anhedonic and depressive processes, traits.neuroticism indexing emotional reactivity, and traits.self_reg representing regulatory capacity. In the simulator, these traits enter multiple downstream mechanisms, including depression sensitivity, strain feedback, recovery and mean-reversion processes, affective variability, and behavioural regulation. Their high ranking therefore reflected between-cluster differences in trait-level vulnerability and regulatory capacity rather than variation in a single output channel.

A third group involved the generation of affective response emissions: ema.core.emotion_gain_base defines the baseline gain by which latent affective dynamics are translated into EMA affect utilities, with additional amplification under low resilience. ema.pa.anhed_low_resource_amp controls the additional reduction of simulated positive affect when depressive or anhedonic liability coincides with low sleep, activity, or social resources. Together with the trait parameters, these coefficients link latent affective vulnerability to the amplitude and expression of the simulated EMA phenotype. Overall, the posterior parameter differences mapped the empirical between-cluster contrasts in Tables 1 and 2 onto distinct combinations of CSDS activity, trait-vulnerability, and affective-response mechanisms.

### 3.3 Decoding analysis

Across GLOBEM clusters, median subject-level correlations indicated high concordance between the behavioural state architecture of the synthetic and observed signals, ranging from *r*=0.836 to *r*=0.870, while occupancy total-variation distance ranged from 0.199 to 0.276 (Table 4; Figure 4). Transition-matrix correlations ranged from 0.930 to 0.943 and Jensen–Shannon divergence from 0.070 to 0.097 bits. The fingerprint signal level correlations ranged between 0.793 to 0.855, indicating a strong alignment of mean signal levels of synthetic and observed states. Correlations of difference features were lower (0.465–0.587), likely reflecting the mismatch in measurement cadence and variability between synthetic (daily) vs. observed EMA channels spanning 3–7 days.

**Figure 4.**
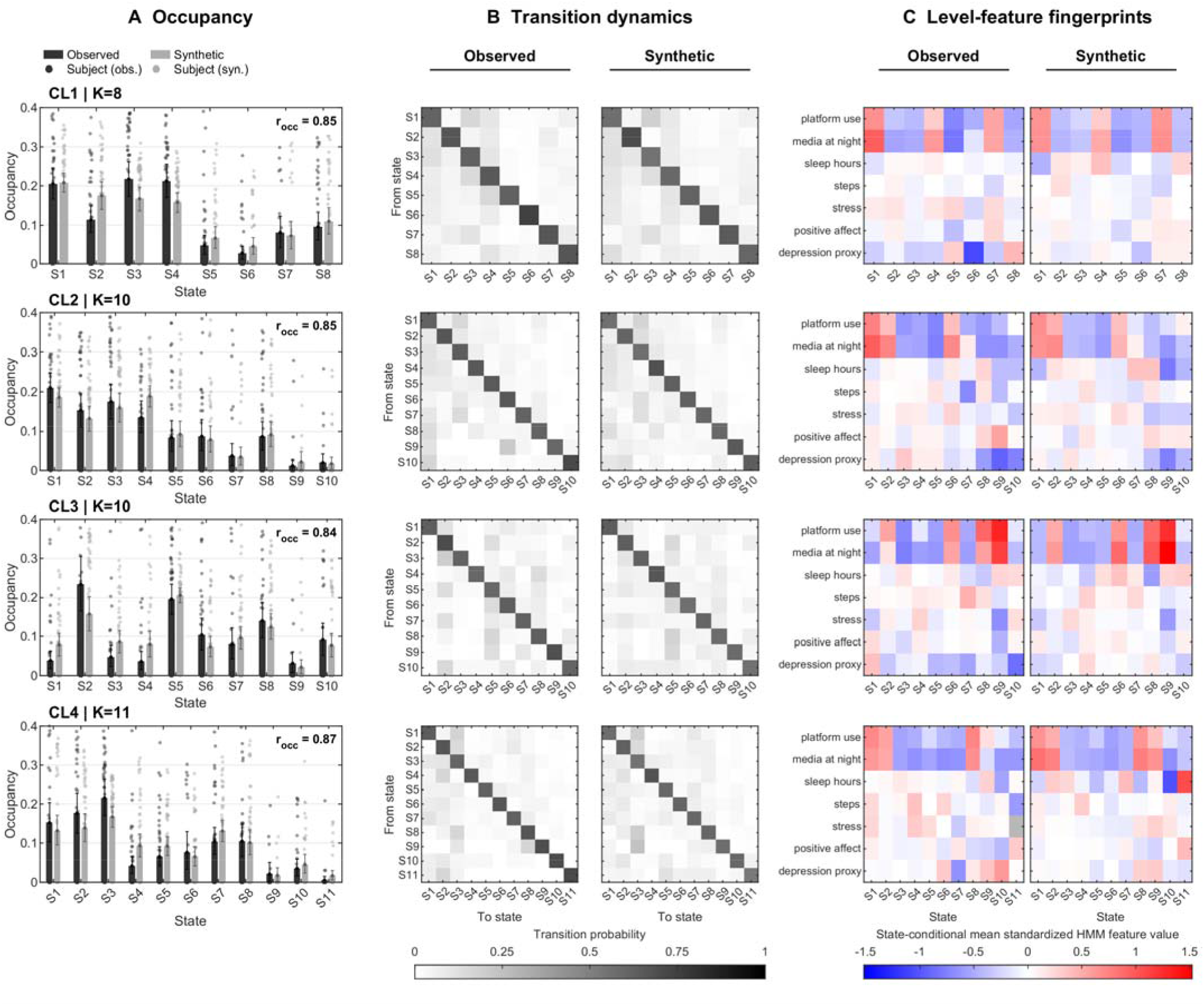
Concordance of VT-derived synthetic and observed sticky-HMM state structure across the four GLOBEM clusters. **(A)** Equal-subject-weighted mean state occupancy for observed sequences and subject-specific virtual-twin ensembles. Bars show cluster means, points show subject-level values and error bars show percentile subject-bootstrap 95% confidence intervals. *r_occ_* denotes the median subject-level correlation between observed and synthetic occupancy vectors. **(B)** Equal-subject-weighted observed and synthetic transition matrices, with rows denoting origin states and columns destination states; all matrices use a common 0–1 probability scale. **(C)** Equal-subject-weighted state fingerprints for the seven level features, expressed as state-conditional means on the synthetic-reference standardized HMM-feature scale. States were aligned within clusters; identically numbered states do not imply cross-cluster homology. CL1 comprised K=8 states, CL2 and CL3 K=10 states and CL4 K=11 states; n=50 subjects per cluster.

**Table 4.**
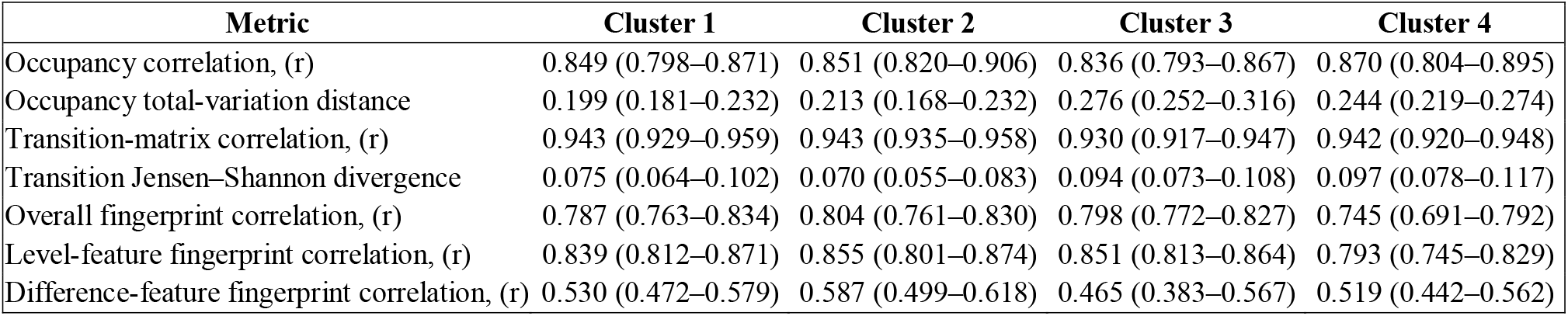
Concordance between observed and VT-derived synthetic state representations across GLOBEM clusters. Values are median subject-level estimates with bootstrap-derived 95%-CIs. Higher correlations indicate greater concordance; lower total-variation distance and Jensen–Shannon divergence indicate smaller discrepancies. Jensen–Shannon divergence is expressed in bits.

| Metric | Cluster 1 | Cluster 2 | Cluster 3 | Cluster 4 |
| --- | --- | --- | --- | --- |
| Occupancy correlation, (r) | 0.849 (0.798–0.871) | 0.851 (0.820–0.906) | 0.836 (0.793–0.867) | 0.870 (0.804–0.895) |
| Occupancy total-variation distance | 0.199 (0.181–0.232) | 0.213 (0.168–0.232) | 0.276 (0.252–0.316) | 0.244 (0.219–0.274) |
| Transition-matrix correlation, (r) | 0.943 (0.929–0.959) | 0.943 (0.935–0.958) | 0.930 (0.917–0.947) | 0.942 (0.920–0.948) |
| Transition Jensen–Shannon divergence | 0.075 (0.064–0.102) | 0.070 (0.055–0.083) | 0.094 (0.073–0.108) | 0.097 (0.078–0.117) |
| Overall fingerprint correlation, (r) | 0.787 (0.763–0.834) | 0.804 (0.761–0.830) | 0.798 (0.772–0.827) | 0.745 (0.691–0.792) |
| Level-feature fingerprint correlation, (r) | 0.839 (0.812–0.871) | 0.855 (0.801–0.874) | 0.851 (0.813–0.864) | 0.793 (0.745–0.829) |
| Difference-feature fingerprint correlation, (r) | 0.530 (0.472–0.579) | 0.587 (0.499–0.618) | 0.465 (0.383–0.567) | 0.519 (0.442–0.562) |

Because Figure 4C expresses state profiles in the synthetic-reference HMM feature space, observed raw-channel levels were additionally referenced to a common pooled four-cluster distribution (Supplementary Figure S11.5a). This recapitulated the phenotype structure: Cluster 1 combined high digital use and low activity with low affective burden; Cluster 2 showed similarly high digital use but markedly greater stress and depression; Cluster 3 combined low digital exposure with high activity and low affective burden; and Cluster 4 showed low digital exposure with intermediate affective burden.

### 3.4 Forecasting future behavioural dynamics

Across the three forecasting architectures, models containing autoregressive components showed the highest short-horizon performance. At *h = 1*, pooled 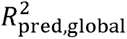 across the 28 cluster-by-channel estimates was 0.450 for the synthetic AR-HMM and 0.446 for its hybrid counterpart, compared with 0.392 and 0.393 for the stickyHMM with autoregressive heads, and 0.311 and 0.413 for the synthetic and hybrid stickyHMM, respectively. By *h = 21*, median values across the six architecture-by-regime combinations converged to a narrower range of 0.353–0.367. Target-stratified analyses of the within-subject skill identified significant decoder-by-horizon interactions for daily steps, sleep duration, and night time platform use in all four clusters. These interactions were strongest for daily steps, ranging from F(8,360)=8.13, *P*_Holm_=7.69×10^-8^, 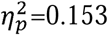, in Cluster 1 to F(8,344)=70.21, *P*_Holm_=2.04×10^-65^, 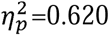, in Cluster 3. Only two individual synthetic–hybrid contrasts survived Holm correction, indicating little advantage of supplementing synthetic with observed training data (Supplementary Tables S12.11a–b).

For the synthetic AR-HMM, 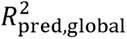 declined mainly between the 1- and 3-day horizons and changed less thereafter (Supplementary Tables S12.6a). Cluster-level channel means ranged from 0.406 to 0.550 at *h = 1* and from 0.322 to 0.458 at *h = 21*, remaining above both causal comparator curves (Figure 5A). Calibration was closest for steps and sleep channels, whereas digital-use showed broader dispersion and EMA-derived stress, positive affect and depression had more restricted distributions (Figure 5E).

**Figure 5.**
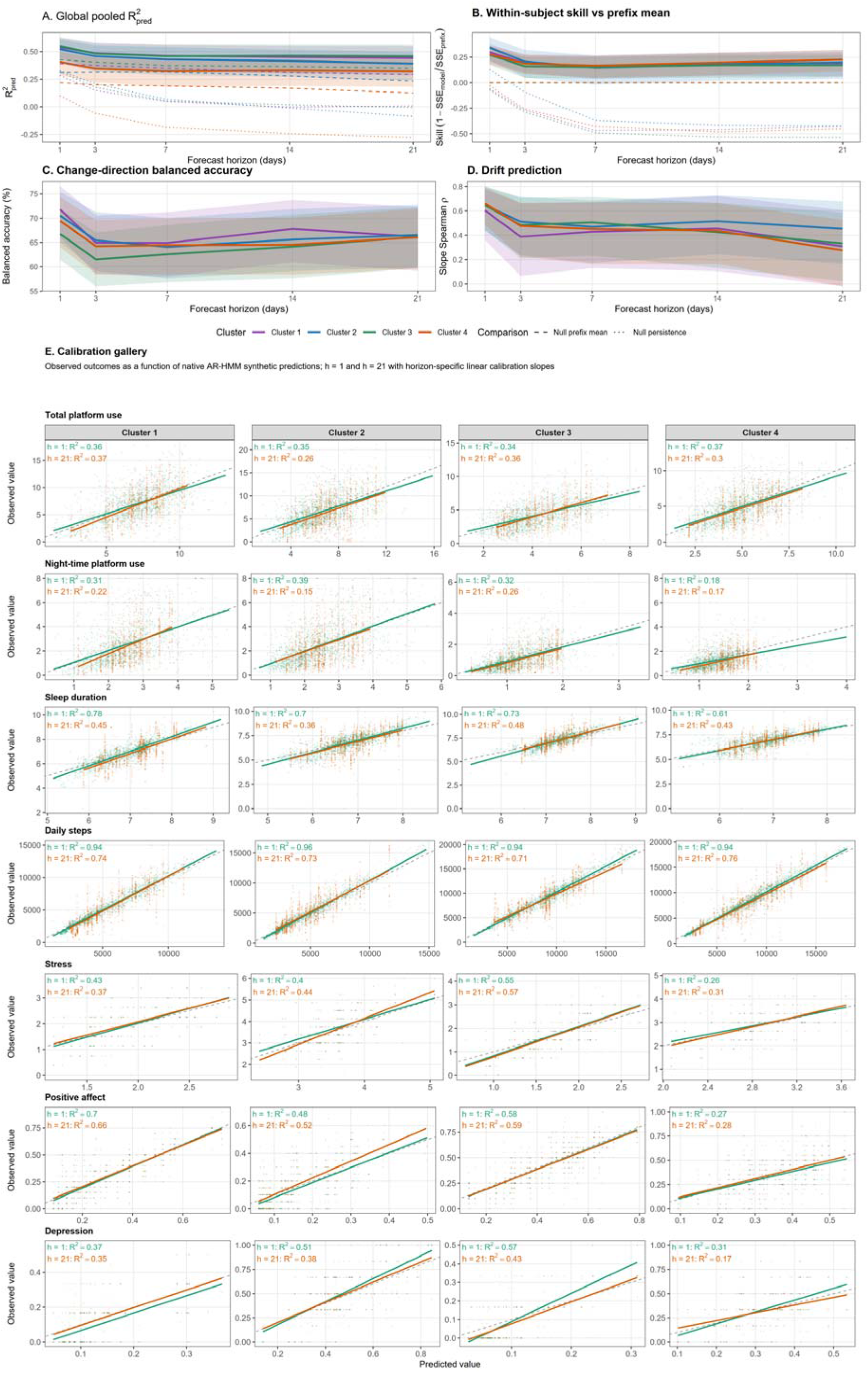
Forecast performance, within-subject skill, drift tracking, and calibration of the synthetic-trained native AR-HMM. Solid lines in panels A–D show the native AR-HMM trained on synthetic virtual-twin time series; dashed and dotted lines show the subject-specific causal prefix-mean and persistence null models, respectively. The “prefix” denotes the observed data available to the model from the start of the monitoring period up to and including the forecast origin (anchor). Panels A–D were calculated separately at each forecast horizon using all evaluable observations at that horizon. Model and causal prefix-mean predictions were evaluated on identical within-horizon support for the skill and change-based analyses. Channel-specific 95% confidence intervals were estimated by 5000 subject-block bootstrap resampling iterations. **(A)** Global pooled predictive 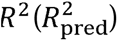 across forecast horizons of 1, 3, 7, 14, and 21 days. **(B)** Within-subject skill relative to the causal subject-specific prefix-mean baseline, defined as 1 – *SSE_model_*/*SSE_prefix_*. A value of zero indicates performance equal to the prefix-mean baseline, positive values indicate improvement, negative values poorer performance vs. the prefix-mean comparator. The dotted line shows persistence skill relative to the same prefix-mean baseline. **(C)** Balanced accuracy for prediction of the direction of change and **(D)** Spearman correlation between predicted and observed subject-level drift slopes. In panels **A–D**, lines denote clus-ter-specific channel means and shaded ribbons summarize the corresponding channel-specific 95% confidence limits. Cluster 1 is shown in purple, Cluster 2 in blue, Cluster 3 in dark green, and Cluster 4 in orange. **(E)** Calibration plots for 1-day and 21-day native AR-HMM forecasts. Rows represent behavioural channels and columns represent clusters. Each point represents an evaluable subject–anchor forecast in the channel’s native units, with predicted values on the x-axis and observed values on the y-axis. Green and orange lines show horizon-specific linear calibration fits for *h* = 1and *h* = 21, respectively; the grey dashed line denotes perfect calibration. Annotations report the calibration *R*^2^ for each horizon.

Within-subject forecasting was evaluated as skill relative to each participant’s causal prefix mean. After inclusion of all seven channels, channel-balanced mean skill at *h = 1* ranged from 0.280 (0.191–0.361) in Cluster 4 to 0.362 (0.241–0.462) in Cluster 2. It decreased to 0.161–0.207 at *h = 3* and 0.149–0.168 at *h = 7*, before stabilising or partially recovering to 0.172–0.198 at *h = 14* and 0.173–0.227 at *h = 21*. Confidence intervals remained above zero throughout, except for Cluster 2 at *h = 7* (Figure 5B; Supplementary Table S12.6b). Thus, the dominant horizon effect was an early loss of exact within-person magnitude accuracy rather than progressive deterioration across the complete 21-day interval.

Channel-resolved results revealed four predictability profiles (Supplementary Table S12.6b). Daily steps showed the highest short-horizon skill, ranging from 0.658 to 0.750 at *h = 1*, which declined to 0.116–0.302 at *h = 7*, but subsequently recovered to 0.270–0.432 at *h = 21*. Sleep-duration skill was 0.326–0.435 at *h = 1* but generally approached zero thereafter, except in Cluster 4, where it remained positive and increased from 0.240 at *h = 7* to 0.330 at *h = 21*. Stress, positive affect and depression showed comparatively stable skill: their respective ranges were 0.232–0.394, 0.174–0.258 and 0.217–0.354 at *h = 1*, and 0.214–0.372, 0.202–0.280 and 0.228–0.375 at *h = 21*. Total platform-use skill remained close to zero, while night-time platform use was forecastable in Clusters 1 and 2 and showed limited skill in Clusters 3 and 4. Predictability was therefore governed by target channel rather than by GLOBEM cluster.

Change-direction balanced accuracy exceeded chance across all clusters and horizons, ranging from 66.8–71.8% at *h = 1* and 66.1–66.6% at *h = 21* (Figure 5C; Supplementary Table S12.7a). Predicted and observed participant-level slow drift slopes were likewise positively correlated, with cluster means of *p = 0.603*–0.661 at *h = 1*and *p = 0.276*–0.453 at *h = 21* (Figure 5D; Supplementary Tables S12.8a–b). Channel-resolved analyses showed that daily steps combined high short-horizon magnitude and drift accuracy with a non-monotonic longer-horizon skill profile, whereas the sparse affective and clinical channels retained moderate skill and comparatively stable direction and drift information across horizons. Depression provided the clearest metric dissociation: magnitude skill was moderate, but directional accuracy reached 71.2–80.0% in Clusters 1 and 2. Supplementary Figures S12.9a–b show corresponding observed trajectories, point forecasts and directional changes for the centroid-closest participant in each cluster.

Split-half timeseries validation did not support that decoding or forecasting results were driven by indirect leakage from full-series VT calibration (Supplementary Tables and Figures S13). Across clusters (n=80 participants), hard target anchors showed variable, channel-specific displacements between first-half and full-series specifications (median participant shift -0.44%), although 15.3% of full-series anchors fell outside the corresponding first-half corridors (Supplementary Table S13a). Weighted soft targets were less stable, particularly within-subject dispersion measures (25.1%). Channel-specific instability was greatest for daily steps (43.8%) and night-time platform use (30.0%) and lowest for sleep (2.5%) and stress (5.0%; Supplementary Table S13b). Decoding and forecasting validation using the split-half timeseries-optimised VTs of clusters 1 and 2 participants reproduced observed state occupancy and transition dynamics almost as well as full-series VTs: 98.5%, 95.8%, 91.1%, 85.2% and 81.6% of full-series causal forecasting skill was retained at 1-, 3-, 7-, 14- and 21-day horizons, respectively, arguing against full-series target construction as the principal source of forecasting performance (Supplementary Table S13c, Figure S13).

## 4 Discussion

Smartphones, wearables and EMA enable scalable measurement of health trajectories in daily life, raising the prospect of preventive strategies that detect hazardous mental-health states before they occur.^71,72^ Yet digital phenotyping remains dominated by descriptive associations and data-driven prediction,^73,74^ motivating interpretable dynamical frameworks that model how psychological, behavioural and physiological processes co-evolve.^16^

Against this background, the principal methodological novelty of the present study lies in integrating clinically encoded cross-domain dynamics, empirical Bayesian inversion, individualised virtual-twin generation, temporal state validation, and out-of-sample forecasting within one naturalistic behavioural modelling framework. Using this framework, we observed that Bayesian inversion produced posterior particle ensembles that closely recovered the four distinct youth mental health phenotypes by identifying plausible combinations of digital-use, physical-activity, trait-vulnerability and affective-response parameters within the simulator’s mechanistic “universe”. This cluster-level results establish that the observed behavioural phenotypes map onto separable regions of the CSDS parameter space, providing the mechanistic scaffold within which the subsequent participant-level virtual-twin inversions individualize longitudinal dynamics. When these participant-specific posterior configurations were used to generate virtual-twin trajectories, hidden Markov models trained on these data closely reproduced the observed multi-domain behavioural patterns, with high concordance in state occupancy, transition structure, and particularly level-feature fingerprints. In the out-of-sample forecasting, models trained on synthetic trajectories generally outperformed causal null models in predicting unseen observations, with little added value of the observed data for improving prediction beyond synthetic AR(1)-augmented model performance (AR-HMM).

In the empirical data analysis step, we observed substantial psychosocial and behavioural heterogeneity in the GLOBEM student population. Study participants could be meaningfully grouped into four behavioural subtypes along two partially separable dimensions: high digital engagement accompanied by physical inactivity, and affective burden paralleled by reduced psychosocial resources and functioning. Their partial independence was most clearly revealed by the paired cluster contrasts: Clusters 1 and 2 shared high platform and night-time media use and comparatively low activity, yet differed markedly in depressive, stress-related, and psychosocial burden, whereas Clusters 3 and 4 shared lower digital exposure and higher activity but likewise separated along the affective-burden dimension. This pattern is consistent with evidence that average associations between digital technology use and mental health are generally small and heterogeneous across individuals, such that digital exposure alone is an insufficient proxy for psychopathology, while lower physical activity shows a more consistent, although not necessarily causal association with depression risk.^75–77^ Importantly for the subsequent generative analyses, the empirical structure was expressed more strongly in mean levels, variability, and selected within-domain persistence measures than in cross-domain temporal couplings, none of which remained significant after false-discovery-rate correction. This aligns with evidence that complex dynamic indices often add limited discriminative information beyond levels and variability.^78^

These clustering results also defined the empirical constraints against which the CSDS could be subjected to a non-trivial reality test: could a single theory-encoded forward simulator identify cluster-specific parameter configurations that simultaneously satisfied the prespecified digital, wearable, EMA, and burden targets characterising each phenotype? Successful recovery of all four multi-domain phenotypes by distinct posterior particle ensembles would demonstrate that the CSDS mechanistic parameter space was sufficiently expressive to generate the major empirical configurations present in the GLOBEM data. The Bayesian inversion results met this criterion: cluster-specific posterior occupied distinct regions of mechanistic feature space, with Cluster 2 particles showing the most prominent separation and highest positional variability. Cluster separation was associated with three clinically coherent parameter families: physical-activity variability, trait vulnerability and self-regulation, and affective-response mechanisms governing EMA expression. The strong discriminative role of these parameters jointly mirrored the empirical clustering findings stratifying participants along physical and digital activity as well as affective-burden dimensions. Thus, the CSDS recovered the four phenotypes through distinct multi-mechanistic configurations rather than variation along a single latent severity axis, supporting the notion that complex phenotype modelling requires coupled multi-domain oscillatory systems approaches.^14,16,20,79^

Based on the cluster-level reproducibility of the empirical phenotype space, the decoding analysis asked whether this generative correspondence extended to the temporal organisation of behaviour within individual participants. Hidden Markov models have been increasingly used in longitudinal psychiatric and digital phenotyping to represent multivariate observations as recurrent within-person states, with occupancy and transition dynamics revealing intraindividual depressive phenotypes, mood instability, and digitally measured behavioural modes.^80–82^ These applications, however, generally derive latent states directly from observed time series; to our knowledge, no study has actually tested whether a separately parameterised mechanistic simulator can generate a temporal architecture closely corresponding to complex human behavioural data. This represents a bottleneck for the validation and construct validity of psychiatric dynamical models, which must be overcome to develop actionable tools for intervention planning.^16,83^

In the present study, models trained on participant-specific virtual-twin trajectories decoded observed sequences with closely corresponding state occupancy, transition organisation, and multi-domain fingerprints, particularly for the levels defining each state. The CSDS therefore reproduced more than the static targets used for inversion: its individualised forward dynamics generated a recurrent state architecture that had not itself been imposed as an inversion constraint. We observed a lower difference-feature concordance (Supplementary Figure S11.6) which may reflect a mismatch in the temporal expression of change rather than low empirical relevance: observed EMA differences integrated changes across multi-day assessment intervals, whereas synthetic differences primarily represented daily emissions. Taken together, the findings of the decoding experiment extend HMM-based state discovery from descriptive temporal phenotyping towards an individual-level test of generative validity. We may cautiously interpret these states as reproducible, model-dependent biobehavioural macrostates rather than uniquely identifiable biological entities: their boundaries reflect the selected measurements, observation cadence, and state-model assumptions,^82^ although their recurrence and synthetic-to-observed correspondence support their utility as clinically interpretable summaries of the temporal organisation of human behaviour.

Based on the recoverability of the observed state architecture, the final hypothesis test asked whether the locked HMM models could extrapolate from the inferred state at a given forecast anchor to temporally held-out multi-modal trajectories. Previous work demonstrated substantial predictability of future human behaviour, mood and health using observed smartphone, wearable, and EMA histories over intervals ranging from the following day to several weeks.^84–86^ More recently, nonlinear state-space models have improved forecasting of multivariate momentary psychological states, supporting the value of explicitly modelling temporal dynamics rather than relying solely on static predictors.^87^ These approaches nevertheless depend on substantial person-specific observed data and reveal considerable heterogeneity in the models and features that perform best for each individual. Within this context, the central finding of our study was that models trained on virtual-twin trajectories generally outperformed the causal prefix-mean and persistence comparators. Strikingly, supplementing synthetic training with observed-prefix data produced little systematic improvement. This suggests that the CSDS trajectories preserved much of the person- and channel-specific temporal structure needed for forecasting and could provide a scalable source of training data when real longitudinal records are short, sparse, costly, or ethically challenging to acquire.^88^

Our study showed that predictability was strongly channel- and horizon-dependent: autoregressive information was most useful close to the forecast anchor whereas exact within-person change magnitudes deteriorated rapidly. In contrast, clinically relevant information about future levels, direction of change, and broader drift remained more stable, resembling the scale-dependent predictability of weather systems, in which fine-grained future states lose forecast skill earlier than slower, larger-scale structure.^89^ The CSDS therefore captured slower state organisation and directional tendencies more reliably than the magnitude of behavioural changes for which forecasting performance may reach a natural ceiling given the probabilistic explosion of behavioural state paths interacting with generally unpredictable future innovations (Supplementary Table S12.12). This is consistent with entropy-based evidence that human behavioural trajectories are highly structured but remain subject to intrinsic limits on predictability.^84,90^ Pending prospective validation in independent cohorts, the results of our study support synthetic trajectories as a data-efficient training resource for establishing predictive models of multi-domain mental health trajectories.

Several limitations qualify these findings. The CSDS is not a comprehensive representation of human behaviour, but a modular and deliberately tractable approximation of interacting world–behaviour–biology processes. Its components compress complex physiological, psychological, and environmental mechanisms into interpretable dynamical rules, while some processes remain simplified, indirect, or absent. The recovered posterior configurations therefore demonstrate generative sufficiency within the current CSDS mechanistic space, but do not uniquely identify the mechanisms underlying the GLOBEM phenotypes or exclude alternative architectures capable of generating similar observations. The mechanistic audit should accordingly be interpreted as identifying discriminative parameter families rather than causal determinants.

The results may also depend on the present cohort, clustering solution, target specification, priors, and observation processes. The four-cluster solution was selected for clinical interpretability rather than by a quantitative stability criterion, and a different cluster number or feature set could yield alternative phenotypes and correspondingly different inversions; our downstream results are conditional on this partition. The GLOBEM sample further comprises university students, which bounds the age range, the digital-behaviour regime, and, most consequentially, the severity distribution of the observed symptom channels. Also, unequal sampling cadence across dense digital and wearable streams and more sparsely measured EMA channels may have contributed to differences in state concordance and predictability. The HMM-derived macrostates likewise remain conditional on the selected channels, emission assumptions, state number, and alignment procedure.

Furthermore, several syndrome-specific CSDS subsystems could not be validated in this study. Mania, obsessive–compulsive and psychosis channels were unassessed in GLOBEM and are of low general-population prevalence;^69,70^ they were held to fixed low-burden corridors across clusters, contributing no discriminative signal. Suicidal ideation was assessed within the BDI-II, but item-level data are unreleased; a structural corridor was substituted, specified a priori as more permissive for the high-burden Clusters 2 and 4 to avoid distorting depression-related parameters. Borderline-instability, eating-disorder and physiological outputs carried no targets. Their generative fidelity therefore remains untested, and the validated scope of this study is confined to the affective, digital-behavioural and wearable channels the GLOBEM cohort expressed.

A potential source of “forecasting optimism” in our results is indirect information leakage from full-series-derived target corridors into the inferred VTs, their simulated trajectories, and consequently the downstream HMMs used for decoding and forecasting. The split-half validation results argue against this being a major explanation: although restricting calibration to the first half produced displaced ABC-SMC posteriors, latent-state occupancy and transition structure were largely preserved, and causal forecasting retained 98.5%, 95.8%, 91.1%, 85.2%, and 81.6% of full-series skill at 1-, 3-, 7-, 14-, and 21-day horizons, respectively (Supplementary Tables S13a-c; Figure S13). The increasing loss at longer horizons is more consistent with less stable target estimation from shorter records, particularly for sparsely sampled EMA measures, than with leakage driving the original forecasting performance. Nonetheless, prospective validation in independent cohorts remains necessary.

These limitations define a concrete development pathway. The modular CSDS architecture permits individual components to be refined or replaced as more accurate physiological, neurobiological, and behavioural models of specific world–behaviour–biology interactions become available. Counterfactual perturbations, subsystem ablations, and comparison with competing mechanistic architectures should determine which mechanisms are indispensable and which can be compensated by alternative pathways, thereby progressively constraining a minimal sufficient mechanistic subspace. Two validation priorities follow most directly from the limitations above. First, clinical cohorts are needed to invert the psychosis, mania, suicidality, obsessive–compulsive, and eating-disorder subsystems against real burden rather than pinning them low, and to validate the physiological outputs, including heart-rate variability and electrodermal activity, that the present sample could not constrain. Second, prospective studies should implement the rolling pre-screening-and-inversion procedure, locking the inversion and forecasting pipelines on a leading window before application to the genuinely unseen new data, and testing predicted responses to naturally occurring or experimental perturbations. This would develop the CSDS from an empirically grounded generative platform towards an iteratively validated framework for counterfactual experimentation and intervention planning.

In summary, the CSDS provides a unified forward-generative framework for reconstructing, decoding, and forecasting multi-domain mental-health trajectories. Its coupled dynamics reproduced distinct empirical GLOBEM phenotypes through coherent combinations of activity, vulnerability-regulating, and affective-response mechanisms, while participant-specific virtual twins generated state architectures that transferred meaningfully to observed behaviour. Synthetic trajectories also supported temporal out-of-sample forecasting, indicating their potential as a scalable source of individualised training data. These findings establish the CSDS as a falsifiable mechanistic platform and define a clear path towards refinement through ablation, counterfactual experimentation, and prospective validation in independent cohorts.

## Supporting information

Supplementary Methods and Results

## Data Availability

I included the following in the manuscript:
All data produced in the present study are available upon reasonable request to the authors

https://the-globem.github.io/datasets/overview

## 5 Data availability

All data produced in the present study are available upon reasonable request to the authors.

## 6 Acknowledgements

The authors acknowledge the use of OpenAI’s ChatGPT (5.1-5.4) and Anthropic’s Claude Opus 4.5 as interactive research and coding assistants during this project. They supported the auditing, documentation and validation of analysis code, suggested sensitivity analyses, and generated summaries of the intermediate and final results. They helped the authors in the preparation of code for figure and table generation, as well as linguistic editing of the manuscript. All scientific hypotheses, methodological decisions, analyses, verification of numerical results, interpretation of findings and final manuscript content remained under the full responsibility and control of the authors.

