## Supplementary Methods and Results for "The Coupled Stochastic Dynamical System: A Generative Model for Simulating and Forecasting Youth Mental Health Trajectories"

Supplementary Material

### S1. CSDS: scope, temporal resolution and simulation sequence

#### S1.1 Model scope and state hierarchy

The Coupled Stochastic Dynamical System (CSDS) is a forward simulator of interacting psychological, psychiatric, behavioural, digital and physiological processes. A simulated individual is defined by fixed demographic and trait characteristics, persistent day-level latent states, within-day behavioural and physiological trajectories, and noisy observation mappings.

The current implementation distinguishes four temporal levels:

1. **Subject-fixed characteristics**, including age, sex at birth, psychological traits, psychiatric predispositions, digital habits and individual sleep need.
2. **Slow day-level states**, including strain, psychiatric burden, environmental (exposomal) burden, sleep pressure, behavioural habits, learned regulatory skills and physiological and digital dysrhythmia states.
3. **Within-day processes**, including sleep–wake allocation, physical activity, autonomic physiology, in-person activity and digital engagement in five-minute bins.
4. **Observation processes**, including six EMA assessments per day and daily summaries of digital and wearable-derived signals.

The within-day simulation step was five minutes, yielding 288 bins per simulated day. EMA prompt times were sampled independently for each day as six ordered times within the interval from 08:00 to 22:00. Psychiatric burden, strain, exposomal states, habits and other slow variables were updated once per day.

#### S1.2 Trait and demographic initialization

Raw psychological and psychiatric traits were represented on a bounded population z-score scale from (-2.5) to (+2.5). The principal trait coordinates were neuroticism, reward sensitivity, self-regulation, social need, resilience, psychosis proneness, cognitive-disturbance proneness, anhedonia proneness, hopelessness, bipolarity proneness, and social-media, gaming and conversational-AI habits (Supplementary Table S1.1a). Individual sleep need was represented directly in hours.

Most raw trait coordinates were transformed to bounded ∈ [0,1] liability variables using the logistic function. Psychiatric predispositions used the same transformation with pathway-specific midpoint shifts where required. The model additionally derived excess and deficit zones for traits with non-monotonic effects (Supplementary Table S1.1b). For a bounded trait $x$, an upper-tail excess was defined as:

$$E\left( x;t,r \right)=\frac{\max\left( x-t,0 \right)}{r}$$

and a lower-tail deficit as:

$$D\left( x;t,r \right)=\min\left( \frac{\max\left( t-x,0 \right)}{r},1 \right)$$

where $t$ was the activation threshold and $r$ the scaling range. These transformations distinguished, for example, low-self-regulation impulsivity from high-self-regulation rigidity, and low-reward-sensitivity withdrawal from high-reward-sensitivity instability.

Adaptive mid-range effects were represented by the inverted-U transformation:

$$U\left( x \right)=4x\left( 1-x \right)$$

Age-dependent prefrontal maturation was represented by a logistic maturation function. The resulting maturation factor scaled the proportion of dispositional self-regulation available to the individual at the simulated age.

#### S1.3 Persistent states

The CSDS evolves from one simulated day to the next through a persistent state vector. A persistent state is any internal quantity whose value at the end of day $d$ is carried forward and used when generating the signals of the next day $d+1$. Persistent states therefore differ from subject-fixed traits, which do not change during a simulation, and from transient within-day outputs, which are recalculated for every five-minute bin or EMA prompt and are not necessarily retained once the day has ended.

Let $\mathbf{S}_{d}$denote the complete persistent state available at the start of day $d$. The next-day state is generated as $F\left( \mathbf{S}_{d},\mathbf{T},\mathbf{X}_{d},\boldsymbol{\varepsilon}_{d} \right)$, where $\mathbf{T}$ contains fixed traits and demographic characteristics, $\mathbf{X}_{d}$ contains the current day’s simulated behavioural, physiological, exposomal and EMA outputs, and $\boldsymbol{\varepsilon}_{d}$ contains stochastic innovations. The state vector gives the simulator memory: two individuals with the same fixed traits can develop different future trajectories because their carried states differ.

The persistent state comprised five functional groups.

**Accumulated regulatory and physiological context.** Global strain, sleep pressure, sleep debt, recovery state, allostatic load and related regulatory variables summarised the accumulated impact of preceding days. These quantities linked recent experience to future dynamics. For example, insufficient sleep increased next-day sleep pressure and psychiatric vulnerability, whereas recovery and regulatory resources accelerated return towards lower-burden states.

**Psychiatric burden and episode memory.** The current depressive, psychotic, manic, suicidal, externalising, obsessive–compulsive and eating-disorder loads were retained across days. For processes with episode-like dynamics, the state also stored regime indicators, time spent in the current regime, burst amplitudes, burst durations, refractory states and basin-memory variables. These variables allowed clinically relevant states to persist after the original trigger had subsided and permitted episode entry, maintenance and recovery to follow different trajectories.

**Dynamical oscillator coordinates.** The physiological and digital dysrhythmia systems retained their current activation, phase, amplitude and regime state. Carrying these coordinates forward allowed circadian and behavioural dysregulation to evolve continuously rather than being redrawn independently each day. They influenced subsequent sleep timing, digital allocation, autonomic physiology and vulnerability to psychiatric burden.

**Learned habits and regulatory skills.** Digital habits, intervention-related skills, behavioural plasticity variables and adherence-related quantities were updated gradually and retained. These states represented experience-dependent change over longer periods. Repeated engagement in a digital behaviour could strengthen the corresponding habit, whereas sustained behavioural regulation or intervention exposure could increase the relevant skill state.

**Lagged behavioural and observational summaries.** Selected previous-day outputs were stored because they entered explicitly into next-day mechanisms. These included prior platform use, night-time digital use, in-person activity, sleep duration, sleep debt, physical activity and the final or aggregated EMA driver states. They implemented directional lagged pathways such as night-time digital use affecting subsequent sleep pressure, prior sleep loss affecting next-day activity and burden, and previous EMA deviations contributing to fast residual psychiatric channels.

Not every quantity generated during a simulated day was added to the persistent state. High-resolution digital, wearable and EMA outputs were retained as observations, but only those summaries required by subsequent-day update equations were carried forward. This distinction kept the state vector limited to variables that mediated temporal dependence across days.

#### S1.4 Daily update sequence

For each simulated day (d), the model proceeded as follows:

1. **Assembly of the day-level context.** Fixed traits and demographic characteristics were combined with the persistent state carried from day $d-1$. This context included the current psychiatric burdens, strain, sleep pressure, allostatic load, digital and physiological dysrhythmia states, learned habits and regulatory skills, and the behavioural and EMA summaries retained from the preceding day.
2. **Generation of exposomal inputs**. Daily environmental inputs were generated, including peer-threat events and cannabis exposure where enabled. These were combined with the subject-fixed early-life-adversity term and the preceding allostatic-load state to update the current exposomal context.
3. **Generation of planned sleep, activity and physiological state**. The wearable module generated the planned sleep schedule from individual sleep need, accumulated sleep debt, strain, psychiatric burden, prior digital behaviour and circadian context. It also generated the current physical-activity trajectory, physiological dysrhythmia state and autonomic signals.
4. **Allocation of waking time and digital behaviour**. Using the planned sleep schedule as an initial constraint, the behavioural module allocated each five-minute bin across sleep, residual other activity, in-person activity and digital engagement. Digital engagement was subsequently distributed across social-media, gaming and conversational-AI channels. Night-time digital binge processes could delay planned sleep onset and open additional waking bins.
5. **Determination of realised sleep and recovery**. The sleep schedule modified by digital behaviour was written back as the realised sleep onset and duration. Realised sleep was then used to calculate sleep loss, sleep debt, recovery and the physiological context carried into subsequent updates.
6. **Update of slow psychiatric and regulatory states**. Psychiatric burdens, regime states, burst processes, fast residual channels and related regulatory variables were updated. These updates used the previous day’s EMA observations, lagged digital, sleep, activity and in-person summaries, the current exposomal context, and the persistent psychiatric and regulatory state.
7. **Generation of current-day EMA observations**. Six EMA assessments were generated from the updated psychiatric burdens, within-day affective and cognitive drivers, current digital and in-person activity, sleep and physiological context, and the current exposomal state.
8. **Construction of the next-day persistent state**. The model updated global strain, allostatic load, behavioural habits, regulatory-skill variables, oscillator coordinates and the selected digital, wearable and EMA summaries required for day $d+1$. High-resolution within-day observations were retained in the simulation output, but only variables entering a subsequent-day equation were added to the persistent state vector.

This order defined the temporal direction of the principal feedback pathways. Previous-day behaviour, sleep and EMA states influenced the current psychiatric update; current psychiatric and physiological states shaped current EMA observations; and realised current-day digital behaviour and sleep were carried forward to influence subsequent strain, burden, activity and behavioural allocation. The study configuration can therefore be summarized as:

$\text{persistent state}\text{ }d\to\text{exposomal and physiological context}\text{ }d\to\text{behavioural allocation and realised sleep}\text{ }d\to\text{psychiatric-state update}\text{ }d\to\text{EMA observations}\text{ }d\to\text{persistent state}\text{ }d+1$.

### S2. Fast residual processes

To preserve rapid, content-specific perturbations that could be obscured by a broader distress state, the CSDS maintained five short-memory residual channels: threat, social evaluation, perseveration, appearance, and addictive cue. For each channel, the relevant prior-day EMA deviations were adjusted for a global distress component dominated by strain and sleep disruption. The remaining channel-specific signal was then carried forward with exponential decay and entered selectively into the slower psychiatric or behavioural processes to which it was mechanistically relevant. This design reflects hierarchical models of psychopathology in which broad transdiagnostic liability coexists with narrower symptom dimensions, while retaining the short-term variability and temporal dependence observed in intensive longitudinal assessments of psychological states (Trull et al. 2015; Kotov et al. 2017).

### S3. Shared dynamical primitives

#### S3.1 Bounded stochastic mean reversion

Most daily burden states were implemented as bounded, state-dependent, mean-reverting stochastic Ornstein–Uhlenbeck processes. This modelling approach was motivated by previous applications of OU models to intensive longitudinal affective and psychotic experience sampling data, in which psychological dynamics are characterised by an individual attractor, a regulatory parameter governing return towards that attractor, and stochastic variability around it (Oravecz et al. 2011). OU models have been applied to psychiatric experience-sampling data to relate psychosis-spectrum experiences to affective attractor position, regulation and variability (Nowak et al. 2023). In the case of the CSDS, we define process $m$ as:

$x_{m}\left( d+1 \right)=\mathrm{clip}_{\left[ 0,1 \right]} \left[ x_{m}\left( d \right)+\kappa_{m}\left( d \right)\left\{ \mu_{m}\left( d \right)-x_{m}\left( d \right) \right\}+\sigma_{m}\left( d \right)\varepsilon_{m}\left( d \right) \right],$where $\varepsilon_{m}\left( d \right)\sim\mathcal{N}\left( 0,1 \right)$.

The attractor target $\mu_{m}\left( d \right)$, recovery rate $\kappa_{m}\left( d \right)$, and innovation amplitude $\sigma_{m}\left( d \right)$ were influenced by relevant traits, exposures, behavioural context, regulatory resources, psychiatric regimes and intervention variables. This common form was instantiated differently for depression, psychosis, suicidality, externalising, obsessive–compulsive and eating-disorder burden. The global strain process used an analogous bounded stochastic transition but incorporated direct feedback from EMA stress and rumination, sleep pressure, digital overuse, in-person activity, circadian disruption and psychiatric burden.

#### S3.2 Hysteretic regime switching

Psychosis, depression, obsessive–compulsive burden and eating-disorder burden were represented as potentially self-maintaining dynamical states with separate entry and recovery thresholds. This hysteretic formulation was motivated by transdiagnostic dynamical-systems accounts of psychiatric disorders and by disorder-specific models describing bistability, attractor persistence, or reinforcing feedback in depression, psychosis, obsessive–compulsive disorder, and eating disorders (Scheffer et al. 2024; Loh et al. 2007; Hosenfeld et al. 2015; Cramer et al. 2016; Rolls et al. 2008; Troscianko and Leon 2020). Physiological dysrhythmia used an analogous hysteretic switch, consistent with physiological sleep–wake models in which mutual inhibition produces bistable sleep and wake states and different transition conditions in the two directions (Phillips and Robinson 2007).

For a latent load $x\left( d \right)$, the regime indicator $r\left( d \right)$ was updated according to $r\left( d \right)=1\quad\text{when}\quad x\left( d \right)>T_{\mathrm{on}}$, and remained active until $x\left( d \right)<T_{\mathrm{off}}$, with $T_{\mathrm{off}}=T_{\mathrm{on}}-H$, where $H>0$ was the hysteresis width. Entry and recovery therefore occurred at different state values. Thresholds and hysteresis widths could depend on trait liability and current environmental or regulatory context.

#### S3.3 Burst processes and basin memory

Psychosis and suicidality involve short-lived high-amplitude burst processes in addition to their slow dynamics (Fusar-Poli et al. 2017; Kleiman et al. 2017; Coppersmith et al. 2023). We modelled the daily burst probability in these processes as a function of the current burden, proximity to the relevant threshold, trait proneness, acute stressors and fast residual channels. Once initiated, a burst contributed an additional transient amplitude for several days. Persistent basin-memory variables were updated alongside the burden processes and contributed additional state retention. Burst amplitude and duration were carried explicitly in the psychiatric state and were also available to the EMA observation model.

#### S3.4 Exponentially persistent auxiliary states

Slow exposures, habits and short-memory residual channels were updated using exponential smoothing. For an input $u\left( d \right)$ with time constant $\tau$,

$\alpha=1-\exp\left( -\frac{1}{\tau} \right)$,	$q\left( d+1 \right)=\left( 1-\alpha\right)q\left( d \right)+\alpha u\left( d \right)$.

This form was used for peer threat, allostatic load, cannabis habit and several behavioural carry-over processes. Fast EMA-derived residual channels used the same principle with an approximately ten-hour decay time.

#### S3.5 Within-day autoregressive drivers

The CSDS uses three latent autoregressive drivers, negative affect, positive affect and cognitive load, to represent the shared, rapidly fluctuating component of EMA observations over the day course. This low-dimensional layer was introduced based on the assumption that EMA signals do not result from entirely independent stochastic processes but from shared covariance of momentary affective or cognitive states interacting with more specific, but slower psychopathological and physiological processes (Merz and Roesch 2011). Negative and positive affect were modelled a separate drivers based on findings showing that they represent partially independent dimensions rather than opposite ends of a single continuum (Rush and Hofer 2014; Watson et al. 1988). Cognitive load was introduced to capture transient occupation of information-processing capacity by sleep loss, strain, contextual demands and psychopathological preoccupation, and therefore provided a common fluctuating substrate for concentration difficulty and related cognitively demanding experiences (Lim and Dinges 2010; Shields et al. 2016; Zetsche et al. 2018).

These three drivers receive projections from slower, syndrome-specific psychiatric burden processes. Depression, psychosis, obsessive–compulsive burden, eating-disorder burden and other psychiatric states evolved mainly across days and could alter the mean input, persistence and volatility of the within-day drivers. Digital behaviour, sleep, physiological state, global strain and current context also contributed to the prompt-level driver inputs. The drivers therefore integrated rapidly changing influences from several domains, but they did not mediate all effects of the slower states.

EMA observations were generated through two parallel routes. First, each EMA channel received a weighted contribution from one or more of the shared negative-affect, positive-affect and cognitive-load drivers. Second, selected channels received direct projections from the relevant psychiatric burdens, traits, digital context and physiological state. This dual-pathway architecture allowed broad transdiagnostic co-fluctuation among EMA observations while preserving syndrome-specific manifestations that could not be reduced to general affect or cognitive load. In parallel, the psychiatric burdens also acted directly on digital behaviour, sleep, activity and autonomic physiology, whose outputs subsequently fed back into later psychiatric and EMA dynamics.

Specifically, for prompt $p$, the three drivers produced a three-dimensional latent state which was updated as the sum of autoregressive carry-over from previous inputs $\boldsymbol{\rho}_{d}\odot\mathbf{z}_{d,p-1}+\mathbf{g}_{d,p}$, deterministic contextual input $\mathbf{g}_{d,p}$and scaled statistical innovation $\boldsymbol{\sigma}_{d}\odot\boldsymbol{\epsilon}_{d,p}$, where

$\mathbf{z}_{d,p}\boldsymbol{=}\boldsymbol{\rho}_{d}\odot\mathbf{z}_{d,p-1}+\mathbf{g}_{d,p}+\boldsymbol{\sigma}_{d}\odot\boldsymbol{\epsilon}_{d,p},$ where $\boldsymbol{\epsilon}_{d,p}\sim\mathcal{N}\left( \mathbf{0},\mathbf{I} \right)$, and $\mathbf{z}_{d,p}=\left[ \begin{matrix} {NA}_{d,p} \\ {PA}_{d,p} \\ {CL}_{d,p} \end{matrix} \right]$

The final prompt state of day $d-1$initialized the first prompt of day $d$. Driver inertia and innovation amplitude were modified by resilience, regulatory skill and psychiatric state. Depressive burden increased negative-affect inertia, consistent with EMA evidence linking depressive symptoms and depressive disorders to stronger affective carry-over (Koval et al. 2013; Hawes and Klein 2024) and reduced innovation amplitude. Emotional-instability processes reduced inertia and increased innovation amplitude to operationalise the rapid and pronounced affective shifts observed in borderline personality disorder (Santangelo et al. 2014). Psychosis, obsessive–compulsive burden and eating-disorder burden were assigned distinct inertia and innovation signatures. EMA studies support altered affect dynamics in psychosis and obsessive–compulsive disorder and diagnostically heterogeneous affect dynamics across eating disorders (Nowak et al. 2023; Williams-Kerver et al. 2020; So et al. 2023), but they do not validate the exact coefficient directions used in the CSDS. At the current development stage of the CSDS, these coefficient modifications should therefore be understood as clinical experience-informed model specifications rather than direct empirical estimates.

After each update, the latent-driver vector was passed through a scaled hyperbolic-tangent transformation. This acted as a smooth saturation function: it was approximately linear around the centre of the latent space but progressively compressed extreme values, preventing unstable autoregressive accumulation and maintaining numerically well-behaved inputs to the EMA observation functions.

#### S3.6 Time-budget conservation

We implemented a wake-budget model that represents a very simplified formulation of young people’s sleep-wake behaviour (see S4.3). Within every five-minute bin $b$, the digital-behaviour module enforced: $s_{d,b}+w_{d,b}=1$, where $s_{d,b}$ was the sleep fraction and $w_{d,b}$ the available waking fraction. Waking time was partitioned as: $o_{d,b}+i_{d,b}+g_{d,b}=w_{d,b}$, where $o_{d,b}$ was residual other activity, $i_{d,b}$ in-person activity and $g_{d,b}$ total digital engagement. Digital engagement was further decomposed as:

$$g_{d,b}=g_{d,b}^{social}+g_{d,b}^{gaming}+g_{d,b}^{chatbot}$$

Structured obligations and a minimum residual-activity floor were allocated before in-person and digital claims. Digital habits initially displaced residual other activity; competition with in-person activity occurred only after the available residual budget was reduced.

The relative allocation of total digital engagement across social media, gaming and conversational AI was generated using a habit- and state-dependent softmax:

$$\pi_{c,d,b}=\frac{\exp\left( \mathcal{l}_{c,d,b}/T_{d,b} \right)}{\sum_{c'} \exp\left( \mathcal{l}_{c',d,b}/T_{d,b} \right)}$$

$$g_{d,b}^{c}=g_{d,b}\pi_{c,d,b}$$

The logits $\mathcal{l}_{c,d,b}$ incorporated stable habits, current utilities, trait and syndrome effects, cross-domain inhibition and stochastic switching. The temperature $T_{d,b}$ increased under strain, psychiatric burden, sleep pressure and dysrhythmia.

#### S3.7 Bounded observation mapping

Prompt-level EMA utilities were converted to bounded 0–6 responses using:

$$y_{c,d,p}=\mathrm{clip} \left[ 0,6 \right]\left[ 6\left\{ 1+\exp\left( -\frac{u_{c,d,p}+b_{c}}{s_{c}} \right) \right\}^{-1}+e_{c,d,p} \right]$$

where $b_{c}$, $s_{c}$ and the observation-noise distribution were channel-specific. Each utility combined the shared latent drivers $\mathbf{z}_{d,p}$ with direct channel-specific terms:

$$u_{c,d,p}=a_{c}+f_{c}\left( \mathbf{z}_{d,p} \right)+\mathcal{l}_{c}\left( \mathbf{L}_{d} \right)+q_{c}\left( \mathbf{C}_{d,p} \right).$$

Here, $\mathbf{L}_{d}$ contained the slow psychiatric loads and $\mathbf{C}_{d,p}$ contained prompt-level behavioural, physiological, trait and contextual variables. The direct-load terms allowed syndrome-specific channels to retain variation not mediated by the shared negative-affect, cognitive-load and positive-affect drivers. The functions $f_{c}$, $\mathcal{l}_{c}$, and $q_{c}$ denote the channel-specific mappings from the shared latent drivers, slow psychiatric loads, and prompt-level contextual variables, respectively. These mappings may include linear coefficients, nonlinear transformations, interactions, and trait-dependent gates.

### S4. Study-relevant module instantiations

#### S4.1 Exposomal and global-strain context

The exposomal module generated three principal non-digital influences, selected for their known adverse impact on multiple mental health dimensions:

1. **Early-life adversity** was represented as a subject-fixed bounded exposure (Kessler et al. 2010). Its current psychiatric effect was residualised against neuroticism, self-regulation and resilience to reduce duplication between adversity and the stable trait layer.
2. **Peer victimisation** was generated as a stochastic daily event. Event probability depended on school-day exposure and current externalising and depressive burden (Reijntjes et al. 2010, 2011). Event intensity was amplified by psychosis and depression-related threat sensitivity and entered an exponentially persistent peer-threat state (Wolke et al. 2014; Reininghaus et al. 2016).
3. **Cannabis exposure** could be disabled or generated as binary or continuous daily use. Use probability depended on baseline exposure probability, accumulated cannabis habit, depressive and externalising burden and early-life adversity (Griffith-Lendering et al. 2011; Hines et al. 2023). Dose and potency were combined through a nonlinear salience transformation. Cannabis habit was updated as an exponential average of prior use.

A persistent allostatic-load state integrated early-life adversity, peer threat and cannabis salience (McEwen 2007):

$$A_{d}=\mathrm{clip}_{\left[ 0,1 \right]} \left[ \left( 1-\alpha_{A} \right)A_{d-1}+\alpha_{A}\mathrm{clip}_{\left[ 0,1 \right]} \left( w_{E}E_{i}+w_{T}T_{d}+w_{C}C_{d} \right) \right],\quad\quad\alpha_{A}=1-\exp\left( -\frac{1}{\tau_{A}} \right)$$

Here, $E_{i}$is early-life adversity, $T_{d}$is persistent peer threat, and $C_{d}$ is cannabis salience. The exposomal module additionally provided sleep-penalty and avoidance-bias signals.

Global strain was updated at the end of each day. Its principal inputs were deviations in EMA stress and rumination, excess digital use, sleep pressure, circadian misalignment, low in-person activity and excess psychiatric burden. Resilience, self-regulation, behavioural skill, recovery and intervention support increased damping or reduced the effective strain attractor:

$$G_{d+1}=G_{d}-\delta_{d}\left( G_{d}-b_{d} \right)+\gamma_{d}F_{d}+\Psi_{d}-R_{d}+\sigma_{d}\varepsilon_{d},\quad\quad\varepsilon_{d}\sim\mathcal{N}\left( 0,1 \right)$$

Here, $F_{d}$ summarizes the behavioural and experiential feedback terms, $\Psi_{d}$ is excess psychiatric burden, $R_{d}$is recovery-related relief, $\delta_{d}$is effective damping, and $b_{d}$ is the trait-, skill- and intervention-modulated strain attractor, which was not used in the current work. The resulting strain state $G$ entered subsequent psychiatric, digital, sleep and EMA dynamics.

#### S4.2 Psychiatric burden and fast residual processes

The psychiatric layer maintained seven primary non-negative loads: psychosis, depression, mania, suicidality, externalising burden, obsessive–compulsive burden, eating-disorder burden. Psychosis, depression, suicidality, externalising, obsessive–compulsive and eating-disorder burden were based on the bounded mean-reverting form defined in S3.1. Specifically the following modelling approaches were implemented:

- **Depressive burden** combined a trait-linked plateau with strain, anhedonia and hopelessness liability, sleep disruption, social withdrawal, low positive activity, fast perseveration and social-evaluation channels, and cross-pathway input from other psychiatric processes. A hysteretic regime represented sustained depressive episodes. An episode-fatigue state accumulated during active episodes and gradually lowered the depressive attractor, permitting endogenous episode termination and a subsequent refractory interval.
- **Suicidality** was represented as a chronic bounded liability with an additional acute burst process. Inputs included depressive and hopelessness burden, social pain, regulatory failure, externalising activation and crisis-related support.
- **Mania** was represented by a distinct episodic oscillator in which sleep and circadian disruption, cannabis exposure and recurrence-related sensitisation increased episode-entry probability, after which activation was transiently self-maintained and gradually decayed with episode duration. Mixed affective burden was calculated from concurrent manic and depressive activation.
- **Psychosis** forcing included psychosis and cognitive-disturbance proneness, peer threat, cannabis salience, sleep and physiological dysrhythmia, fast threat and social-evaluation signals, and regulatory or intervention terms. Psychosis also included threshold-dependent brief limited intermittent psychotic-symptom-like bursts.
- **Externalising burden** was continuously updated without a discrete regime. Reward sensitivity, low self-regulation, acute contextual burden, addictive-cue residuals and cannabis exposure increased its attractor and volatility.
- **Obsessive–compulsive burden** combined high-regulation rigidity, perseveration, stress and compulsive-control processes.
- **Eating-disorder burden** combined reward-sensitivity deficit, appearance-related residuals, developmental and sex-related gates, restrictive processes and corresponding intervention terms.
- A signed **emotional-instability** mood-pole process represented alternating activated/externalising and depressive/crash states. Its amplitude depended on upper-tail reward sensitivity, low resilience, neuroticism, social-evaluation perturbations, adversity and cannabis exposure. This state altered affective volatility, digital and in-person behaviour, autonomic arousal and selected EMA channels but was not counted as an additional non-negative burden.

Five fast residual channels linked preceding-day content-specific deviations to subsequent slow states. The **threat** channel captured excess irritability, stress and arousal after removing shared distress and cognitive-load effects, and entered psychosis and externalising pathways. **Social evaluation** represented loneliness and social pain beyond general distress and influenced depressive and emotional-instability pathways. **Perseveration** captured rumination that persisted beyond concurrent cognitive load and contributed to depressive and obsessive–compulsive burden. **Appearance** represented non-negative excess in appearance-comparison concerns and entered eating-disorder forcing. **Addictive cue** captured domain-specific digital-use salience beyond the person’s overall digital exposure and influenced externalising burden and subsequent digital-use propensity.

The channels decayed exponentially over an approximately ten-hour timescale, allowing brief content-specific experiences to affect later psychiatric dynamics without becoming persistent burden states themselves.

#### S4.3 Sleep, physical activity and autonomic physiology

The wearable module first generated a planned sleep schedule. Sleep–wake dynamics were based on Borbély’s two-process model of sleep regulation (Borbély 1982), in which sleep–wake-dependent homeostatic pressure (Process $S$) interacts with circadian sleep–wake propensity (Process $C$). Although originally proposed in 1982, this framework remains a central systems-level account of human sleep regulation which has been recently to include mechanistic neuronal driving forces (Borbély 2022).

Wake drive incorporated low self-regulation, reward sensitivity, strain, digital habits, prior night-time digital exposure, psychiatric burden, mania, emotional instability, externalising activation, weekend structure and cannabis withdrawal. Depressive burden entered through partially distinct withdrawal and endogenous-physiology pathways. The planned onset was additionally shifted by developmental chronotype and social-jetlag effects. Furthermore, the digital module could subsequently delay planned onset through a nightly binge state. The resulting realised sleep onset and duration replaced the planned duration in the daily wearable output. Sleep quality and sleep debt were calculated from realised sleep and the current physiological and psychiatric context.

A physiological dysrhythmia oscillator was embedded in the CSDS that integrates trait vulnerability, sleep debt, strain, psychiatric burden, prior digital dysrhythmia, previous-day digital exposure, night-time digital exposure, in-person protection and regulatory capacity. This construct was motivated by evidence that sleep and circadian disruption impair autonomic regulation and that altered diurnal physiological rhythms are associated with transdiagnostic psychopathology and mood instability (Alvares et al. 2016; Grimaldi et al. 2016; Carr et al. 2018; Meyer et al. 2024). Night-time digital exposure and regulatory capacity were included based on studies linking evening light-emitting-device use to circadian phase delay and HRV to neurovisceral self-regulation, respectively (Thayer et al. 2009; Chang et al. 2015). The dysrhythmia oscillator involves hysteretic entry and recovery thresholds, phase diffusion and state-dependent amplitude as phenomenological representations of transitions between regulated and dysrhythmic physiological states. Its outputs affect heart-rate variability, heart rate and electrodermal activity and enter subsequent digital and psychiatric dynamics and thereby fed back indirectly into later sleep.

Daily steps were generated from a lognormal, autoregressive activity process with pull towards an individual activity anchor. Strain, depressive burden, anhedonic psychomotor suppression, psychosis, eating-disorder-related compulsive activity, mania, emotional instability and prescribed physical activity modified the daily total (Stults-Kolehmainen and Sinha 2014; Vancampfort et al. 2017; Dittmer et al. 2018; Krane-Gartiser et al. 2014; Bizzozero-Peroni et al. 2024). Previous-day platform use and night-time concentration reduced steps through a lagged time-displacement pathway, with in-person activity and regulatory resources providing a partially protected time-displacement pathway (Serrano-Sanchez et al. 2011; Kushlev and Leitao 2020; Lin et al. 2023; He et al. 2026). The daily step total was distributed across five-minute bins using bout-shaped activity kernels. Heart rate, heart-rate variability and electrodermal activity were generated at the same temporal resolution from circadian phase, activity, sleep debt, strain, psychiatric state, trait-linked physiology and dysrhythmia. The wearable module returned realised sleep summaries, steps, heart rate, electrodermal activity, heart-rate variability, physical-activity dose and end-of-day oscillator states.

#### S4.4 Digital behaviour and in-person activity

The digital module generated total digital engagement, its allocation across platform domains, in-person activity, school attendance and nocturnal binge behaviour under the conserved waking-time budget defined in Section S3.6.

The CSDS maintains a slow digital-engagement dysregulation oscillator separately from the physiological dysrhythmia oscillator described in Section S4.3. The two processes were coupled across days: physiological dysrhythmia increased vulnerability to dysregulated digital engagement, whereas digital dysrhythmia and night-time platform exposure contributed to subsequent physiological dysregulation. The input structure of the digital oscillator was informed by process models in which problematic digital engagement emerges through interactions among reward sensitivity, affective and psychiatric states, executive control, coping motives and gradually acquired habits (Brand et al. 2019; Wood and Rünger 2016). The oscillator integrated self-regulatory capacity, reward sensitivity, social need, global strain ($G$, see S4.1), sleep pressure, psychiatric burden and accumulated digital habits. A self-regulation-dependent threshold governed entry into the dysregulated regime. Increasing dysregulation prolonged recovery, increased phase diffusion and attenuated the amplitude of the daily digital-engagement rhythm. The oscillator and its threshold dynamics were phenomenological CSDS implementations rather than empirically estimated clinical mechanisms.

A sticky school attendance state represented structured weekday obligations. Sleep loss, psychiatric burden, withdrawal and loss of daily structure increased attendance fragility, whereas regulatory and intervention support were modelled to increase attendance probability. This construction was supported by evidence linking insufficient or disturbed sleep and depressive symptoms to school non-attendance in adolescence (Hysing et al. 2015; Finning et al. 2019). Attendance constrained the time available for digital activity during the corresponding bins.

Within the waking-time budget, the in-person claim depended on social need, recent in-person activity, psychiatric burden, digital withdrawal, emotional-instability state and structured obligations. The digital claim combined habit-gated volitional engagement with a clinical escape component driven particularly by depressive withdrawal and psychosis-related checking. This distinction was informed by theoretical models differentiating habitual and reward-driven engagement from compensatory internet use aimed at escaping adverse affect or unmet offline needs (Kardefelt-Winther 2014; Brand et al. 2019). Its temporal distribution was modified by circadian engagement profiles, chronotype, school attendance, post-school rebound and stochastic day-level variation.

The nightly binge process described in Section S4.3 could extend waking time beyond planned sleep onset and add further night-time digital exposure. Binge probability, duration and intensity depended on digital habits, prior bingeing, sleep debt, regulatory capacity and psychiatric state. In addition to our clinical experience, this mechanism was supported by studies associating bedtime and night-time digital-media use with delayed sleep onset, shorter sleep duration and poorer sleep quality in children and adolescents (Hale and Guan 2015; Carter et al. 2016; Woods and Scott 2016).

Total digital engagement was allocated across social-media, gaming and conversational-AI channels using the softmax mechanism defined in Section S3.6. Channel-specific logits incorporated the corresponding habits, reward sensitivity, social need, psychiatric effects and cross-domain inhibition. Realised use was subsequently written forward into slowly adapting channel-specific habit states, reflecting the gradual strengthening of behaviours repeated in recurring contexts (Wood and Rünger 2016). The softmax allocation and cross-domain competition were computational mechanisms used to represent competition among alternative digital activities rather than empirically estimated choice equations.

The digital outputs used in the present study were:

- platform hours: $G_{d}=\sum_{b} g_{d,b}\Delta_{t}$
- night-time platform hours: $G_{d}^{\mathcal{N}}=\sum_{b\in\mathcal{N}} g_{d,b}\Delta t$, and
- night-time share: $R_{d}^{\mathcal{N}}=\frac{G_{d}^{\mathcal{N}}}{G_{d}}$,

where $\mathcal{N}$ comprised bins outside the 06:00–22:00 daytime interval.

#### S4.5 EMA observations and validator-facing psychiatric proxies

The current implementation generated 20 prompt-level 0–6 EMA channels: stress; rumination; loneliness; concentration difficulty; irritability; digital urge; perceived digital benefit; paranoia; hallucinatory experiences; insight; anhedonia; hopelessness; suicidality; appearance comparison; compulsive checking; rigidity; restrictive-eating urge; emotional reactivity; positive affect; elevated mood.

Utilities for the core affective and cognitive channels were based primarily on the negative-affect, cognitive-load and positive-affect drivers. Syndrome-specific channels additionally received direct projections from the corresponding burden states and trait gates. For example, compulsive checking received direct obsessive–compulsive and rigid-overcontrol contributions, consistent with evidence for impaired goal-directed control and excessive habitual responding in obsessive–compulsive disorder (Gillan et al. 2011). Restrictive-eating urge received eating-disorder, developmental and sex-related contributions, reflecting the developmental concentration and marked sex difference observed for restrictive eating disorders (Treasure et al. 2020). Hallucination-like experiences received psychosis-state and transient burst contributions, reflecting evidence that positive psychotic experiences can fluctuate over short timescales and may occur as brief intermittent episodes (Fusar-Poli et al. 2017; Bell et al. 2024).

The positive-affect proxy used for empirical validation of the CSDS was constructed as a simulator-side analogue of the PANAS positive-affect measure collected in the GLOBEM weekly EMA surveys. PANAS treats positive affect as a distinct dimensional score, and the GLOBEM release provided a positive-affect subscale on a 0–20 scale (Watson et al. 1988; Xu et al. 2023). The simulator proxy was the normalised daily mean of the six prompt-level positive-affect observations:

$$\mathrm{PA}_{d}^{\mathrm{proxy}}=\frac{1}{6}\left( \frac{1}{6}\sum_{p=1}^{6} y_{\mathrm{PA},d,p} \right)$$

Equivalently, this was the mean prompt-level positive-affect score divided by its maximum value of 6. During validation, the observed GLOBEM PANAS-positive score was mapped from its released 0–20 scale into the corresponding simulator emission range.

The GLOBEM depression target was derived preferentially from the depression subscale of the PHQ-4, an ultra-brief validated measure containing separate two-item depression and anxiety dimensions (Kroenke et al. 2009; Xu et al. 2023). Because the CSDS did not reproduce the PHQ-4 questionnaire items directly, its daily depression proxy was constructed as a broader internalising emission from anhedonia, hopelessness, rumination and irritability:

$$D_{d}^{\mathrm{raw}}=\frac{1}{4}\sum_{c\in\mathcal{D}} \left( \frac{1}{6}\sum_{p=1}^{6} \frac{y_{c,d,p}}{6} \right),\quad\mathcal{D}=\{\mathrm{anhedonia},\mathrm{hopelessness},\mathrm{rumination},\mathrm{irritability}\}$$

This composite was then rendered onto the bounded validator-facing scale:

$$D_{d}^{\mathrm{proxy}}=\mathrm{clip}_{\left[ 0,1 \right]} \left[ g_{D}\left( D_{d}^{\mathrm{raw}}-b_{D} \right) \right]$$

where $g_{D}$and $b_{D}$were fixed gain and offset parameters in the parameter library. The proxy was therefore designed to reproduce the distributional characteristics of the GLOBEM PHQ-4 depression target while retaining the richer symptom dynamics available within the simulator. Where the PHQ-4 depression subscale was unavailable in GLOBEM, the extraction pipeline used the total PHQ-4 score and then the available weekly depression measure as predefined fallbacks.

The EMA output structure also retained the day-level psychiatric loads, mixed burden, emotional-instability state, burst states, digital summaries and positive-affect latent summaries for diagnostics and downstream analysis.

### S5. Empirical constraint status in the present study

Not every implemented CSDS component could be validated against the empirical time series provided by the GLOBEM dataset (Xu et al. 2023). The empirical role of each principal process or output was classified as directly constrained, indirectly constrained, structurally safety-constrained, used in downstream state modelling, or generated but not evaluated.

| **CSDS component or output** | **Role in the present study** |
| --- | --- |
| Total platform use | Direct hard cluster- and participant-level target; used in decoding and forecasting |
| Night-time platform use | Direct hard cluster- and participant-level target; used in decoding and forecasting |
| Night-time platform-use share | Direct hard target |
| Realised sleep duration | Direct hard target; used in decoding and forecasting |
| Daily steps | Direct hard target; used in decoding and forecasting |
| EMA stress | Direct hard level target; additional soft dispersion and threshold targets; used in decoding and forecasting |
| Positive-affect proxy | Direct hard level target; additional soft dispersion and low-tail targets; used in decoding and forecasting |
| Depression proxy | Direct hard level target; additional soft dispersion and tail-probability targets; used in decoding and forecasting |
| Digital and wearable within-person SDs | Weighted soft targets |
| Total and night-time platform-use autocorrelation | Weighted soft targets |
| Rumination proxy | Approximate weighted soft target |
| Between-participant SDs of selected participant means | Cluster-level soft targets only |
| Q10 and Q90 of stress, positive affect and depression-proxy participant means | Cluster-level soft targets only |
| Psychosis-load mean | Fixed structural safety corridor |
| Mania-load mean | Fixed structural safety corridor |
| Obsessive–compulsive-load mean | Fixed structural safety corridor |
| Suicidal-load mean | Fixed structural safety corridor |
| Stable traits and digital habits | Constrained through cluster- or participant-specific prior ranges rather than phenotype targets |
| Global strain, sleep pressure and dysrhythmia states | Indirectly constrained through their effects on observed target channels |
| Social-media, gaming and conversational-AI decomposition | Generated but not directly constrained because GLOBEM did not provide corresponding app-semantic target channels |
| In-person activity | Generated and used internally; not directly constrained |
| Heart rate, heart-rate variability and electrodermal activity | Generated but not empirically evaluated in this study |
| Remaining EMA channels | Generated and used to define internal psychiatric dynamics; not directly validated individually |
| Cross-domain coupling targets | Calculated diagnostically but excluded from the scored ABC-SMC target specification |
| Critical-slowing-down and phase-regulation biomarkers | Calculated for clustering or diagnostics but excluded from scored ABC-SMC fitting |
| Intervention modules | Implemented but disabled during the observational validation simulations |

The inversion therefore tested whether the complete coupled architecture could reproduce the observed digital, sleep, activity and EMA target geometry while maintaining low burden in psychiatric dimensions not represented by the intended cluster phenotype. It did not constitute separate empirical validation of every internal state, syndrome pathway or physiological output.

### S6. Parameterization, implementation and reproducibility

#### S6.1 Parameter library

All simulator coefficients were defined centrally in a library which contains more than 2,400 explicit parameter fields, including active coefficients, fixed rendering constants, configuration flags, compatibility aliases and intervention parameters. ABC-SMC inversion sampled only the eligible active subset remaining after fixed, blocked, intervention-specific and inactive paths had been removed.

The principal families in the CSDS model parameter library were:

| **Parameter family** | **Content** |
| --- | --- |
| P.trait, P.traits | Trait distributions, logistic transforms, excess and deficit zones, sleep need and digital habits |
| P.population, P.demog, P.sexEffects | Age, sex-at-birth, gender and developmental moderation |
| P.expo | Early-life adversity, peer threat, cannabis exposure, allostatic load and buffering |
| P.sleep | Sleep need, sleep pressure, rebound, chronotype, social jetlag and realised-sleep bounds |
| P.wearable | Steps, HRV, heart rate, electrodermal activity, sleep and physical-activity effects |
| P.oscillator | Physiological and digital dysrhythmia geometry |
| P.digital, P.digitalDay | Digital propensity, time-budget allocation, night-time binge, school attendance and domain allocation |
| P.psychiatric | Psychiatric plateaus, forcing, recovery, volatility, tipping thresholds, bursts and regime dynamics |
| P.fast | Short-memory residual-channel construction |
| P.ema | Within-day latent drivers, channel utilities, observation mappings and validator-facing proxies |
| P.strain | Global strain feedback, damping, noise and psychiatric-load contributions |
| P.habit, P.skill, P.plastic | Behavioural habit and regulatory-skill learning |
| P.intervention, P.modules | Intervention adherence and mechanism-specific effects |
| P.sim | Time resolution, initial conditions, scenario effects and orchestration constants |

Parameters were expressed in the natural units of their associated subsystem whenever possible: hours for sleep and daily behavioural exposure, steps per day for physical activity, five-minute fractions for within-bin allocation, 0–1 for bounded loads and probabilities, and 0–6 for EMA observations.

#### S6.2 Authoritative implementation modules

The forward simulation was implemented in the following principal functions:

| **Function** | **Responsibility** |
| --- | --- |
| runSimulation.m | Initializes the subject, maintains persistent states, executes the daily module order and returns the complete simulation |
| initTraits.m | Performs all trait, developmental and demographic transformations |
| generateExposomalDay.m | Generates daily adversity, peer-threat, cannabis and allostatic inputs |
| generateWearableDay.m | Generates planned sleep, steps, dysrhythmia and autonomic physiology |
| generateDigitalDay.m | Performs time-budget-constrained digital and in-person allocation and determines realised sleep |
| updatePsychiatricState.m | Updates psychiatric burdens, regimes, bursts, fast residuals and related persistent states |
| generateEMADay.m | Generates the within-day latent drivers and prompt-level EMA observations |
| loadSimParams.m | Defines the complete parameter library |
| expoTemplate.m, wearableTemplate.m, digitalTemplate.m, emaTemplate.m | Define the output schema of each simulation module |

The daily result returned by the simulator contained the strain trajectory, daily exposomal structures, five-minute wearable and digital outputs, prompt-level EMA outputs, fixed trait values, structural-functioning states and intervention diagnostics.

#### S6.3 Randomness and repeated simulation

Stochasticity entered through trait sampling, initial demographic sampling, day-level burden innovations, exposomal events, oscillator transitions, digital allocation, night-time binge episodes, activity generation, autonomic signals and EMA observation noise. The validator assigned reproducible simulation seeds to individual proposals and repetitions. The exposomal generator additionally used a deterministic day-indexed random-number substream derived from its configured seed and the simulation time step.

Because one parameter configuration could generate different stochastic trajectories, cluster-level and participant-level ABC-SMC evaluated configurations over repeated simulations and compared aggregated phenotype summaries with the corresponding target specification. The number of repetitions and simulation days used at each inversion level are reported in the Bayesian inversion section.

### S7 CSDS-compatible feature space

GLOBEM observations from the INS-W_2, INS-W_3 and INS-W_4 study waves were transformed into a CSDS-compatible phenotype space defined a priori by the simulator and validator architecture. The primary phenotype families comprised digital behaviour, including total platform use and night-time media share; wearable-derived physical activity and sleep duration; and weekly survey-derived stress, positive-affect and depression measures. Repeated observations were summarised for each participant using level descriptors and, where sufficiently supported by the available data, dynamic descriptors such as within-person standard deviation, lag-1 dependence, tail-event frequency and temporal-regularity measures.

Baseline psychosocial and psychometric measures were not included in the primary clustering feature set and were not used as ABC-SMC target constraints. They were retained for group-level description and post hoc interpretation of the resulting clusters.

Participants were first restricted to the three study waves used for cluster discovery. Within each study wave, channel-specific missingness was calculated relative to the largest number of observations available for that channel among participants in the same wave:

$$m_{i,c}=1-\frac{n_{i,c}}{\max_{j\in s\left( i \right)} n_{j,c}}$$

Participants were excluded when this missingness metric exceeded 50% in any of the following six core channels: total platform use, sleep duration, daily steps, perceived stress, positive affect or depression. Thus, a participant was retained only when the number of available observations in every core channel was at least half of the corresponding within-study maximum.

Model inversion was performed on derived phenotype measures rather than on the original heterogeneous GLOBEM variables for two related reasons. First, GLOBEM and the CSDS represented related constructs using different instruments, scales, sampling schedules and levels of abstraction. Model inversion therefore required a concept-level mapping into quantities that could be computed comparably from both the observed data and the simulator. Second, reducing the longitudinal observations to a predefined set of participant-level phenotype descriptors substantially reduced the dimensionality and computational cost of ABC-SMC evaluation.

Directly corresponding digital and wearable measures were harmonised primarily through unit and scale transformations. Survey-derived affective measures were mapped to related CSDS-facing proxies, including mappings between PANAS positive affect and the CSDS positive-affect proxy and between PHQ-4 depression and the CSDS depression proxy. These mappings represented alignment at the construct level rather than item-level reproduction of the original questionnaires. Partial and approximate mappings were explicitly labelled and distinguished from directly observed simulator outputs (see S5).

### S8 Clustering strategy and manual four-cluster selection

The clustering matrix comprised 24 participant-level features. Eight features represented phenotype levels: total platform use, night-time platform use, night-time platform-use share, sleep duration, steps, stress, positive affect, and depression burden. Sixteen features represented within-participant dispersion, persistence, or tail behaviour: the standard deviation and lag-1 autocorrelation of total platform use; the standard deviation and lag-1 autocorrelation of night-time platform use; sleep-duration standard deviation; step-count standard deviation and lag-1 autocorrelation; stress standard deviation, lag-1 autocorrelation, and high-stress proportion; positive-affect standard deviation, lag-1 autocorrelation, and low-positive-affect proportion; and depression-burden standard deviation, lag-1 autocorrelation, and 95th percentile.

Missing values remaining in the clustering matrix were replaced separately for each feature by the median of that feature across the retained participants. Each feature was subsequently standardised to zero mean and unit variance. Values that remained undefined after standardisation, for example because of zero feature variance, were set to zero.

Participants were assigned to four clusters using k-means clustering with Euclidean distance in the standardised feature space. The number of clusters was fixed at $k=4$ based on the clinical interpretability of the factor solution (see **Figure 1**, main manuscript). Cluster assignments were used to calculate cluster-specific participant summaries and to generate the corresponding validator target specifications.

### S9 Construction of validator target specifications

A predeclared mapping registry was constructed to link participant-level GLOBEM summaries to phenotype paths returned by the CSDS validator. For each candidate target, the registry specified the source metric, CSDS phenotype path, mapping class, hard or soft role, soft-target weight, proportional corridor factor, admissible lower and upper bounds, target class, estimation level, and derivation rule. The same registry was used to generate the cluster-level and participant-specific target-specification files.

Digital platform use was calculated from daily total screen-unlock duration and expressed in hours. Night-time digital use was approximated as screen-unlock duration between 00:00 and 06:00 plus one third of the 18:00–24:00 evening interval, thereby approximating the CSDS interval from 22:00 to 06:00. Night-time share was calculated as night-time digital use divided by total platform use. Stress summaries were linearly transformed from the observed PSS-4 range of 0–16 to the CSDS range of 0–6. Positive-affect summaries were linearly transformed from the PANAS-positive range of 0–20 to the CSDS proxy range of 0–1. The depression proxy was retained on a 0–1 burden scale using the PHQ-4 depression subscale divided by 6, with the available BDI-II summary data divided by 63 as fallback for the participant-level mean and dispersion summaries, if PHQ-4 data were not available. Stress and positive-affect standard deviations were transformed by the corresponding linear scale factors. To define within-subject dynamic targets the autocorrelation AR(1) of total platform use and night-time platform use were employed.

#### S9.1 Cluster-level target specifications

For cluster $k$ and target metric $j$, the target centre for non-correlation metrics was the arithmetic mean of the participant-level CSDS-compatible summaries:

$$\mu_{k,j}=\frac{1}{n_{k,j}}\sum_{i=1}^{n_{k,j}} x_{i,k,j}$$

A target-specific proportional corridor was first constructed around this centre. For a non-zero centre, its half-width was:

$h_{kj}=\max\left( f_{j}\left| \mu_{k,j} \right|a_{j} \right)$,

where $f_{j}$ was the prespecified proportional corridor factor and $a_{j}$ was the minimum absolute half-width. Proportional corridor factors are reported in Supplementary Table S9.1. The proportional interval was:

$P_{k,j}=\left[ \max\left( B_{j}^{L},\mu_{k,j}-h_{k,j} \right),\min\left( B_{j}^{U},\mu_{k,j}+h_{k,j} \right) \right]$,

where $B_{j}^{L}$ and $B_{j}^{U}$ were the admissible bounds of the target. For centres numerically equal to zero, the proportional factor itself was used as the candidate half-width, subject to the same absolute-width floor and admissible bounds.

The 25^th^ and 75^th^ percentiles of the participant-level values were also calculated:

$Q_{k,j}^{25}=Q_{0.25}\left( x_{1,k,j},\ldots,x_{n_{k,j}k,j} \right)$,
$Q_{k,j}^{75}=Q_{0.75}\left( x_{1,k,j},\ldots,x_{n_{k,j}k,j} \right)$.

When a non-degenerate empirical interquartile range was available, the final target corridor was the bounded outer envelope of the proportional interval and empirical interquartile range:

$L_{k,j}=\max\left[ B_{j}^{L},\min\left( Q_{k,j}^{25},P_{k,j}^{L} \right) \right]$,
$U_{k,j}=\min\left[ B_{j}^{U},\max\left( Q_{k,j}^{75},P_{k,j}^{U} \right) \right]$.

Thus, the corridor was not narrower than either the prescribed proportional band or the central 50% of the observed participant distribution.

Correlation-like targets were constructed in Fisher-(z) space. For participant-level correlations $r_{i,k,j}$,
$z_{i,k,j}=\mathrm{atanh} \left( r_{i,k,j} \right)$, and the cluster centre was:

$\bar{z}_{k,j}=\frac{1}{n_{k,j}}\sum_{i} z_{i,k,j},\quad\quad c_{kj}=\tanh\left( \bar{z}_{k,j} \right)$.

The Fisher-(z) half-width was:

$$h_{z,kj,}=\min\left[ 0.50,\max\left( 0.10,t_{n_{k,j}-1,0.84}s_{z,k,j} \right) \right]$$

corresponding to a central 68% between-subject interval with lower and upper half-width limits of 0.10 and 0.50 on the Fisher-(z) scale. The resulting corridor was:

$$L_{k,j}=\max\left[ B_{j}^{L},\tanh\left( \bar{z}_{k,j}-h_{z,k,j} \right) \right],$$

$$U_{k,j}=\min\left[ B_{j}^{U},\tanh\left( \bar{z}_{k,j}+h_{z,kj} \right) \right].$$

The conjunctive hard-target set was defined before cluster-specific corridor generation, i.e. violation of one corridor led to particle rejection. Corridors were computed for each cluster and comprised the means of total platform use, night-time platform use, night-time platform-use share, sleep duration, daily steps, stress, positive affect, and depression burden. All remaining empirical targets were written as weighted soft constraints.

Soft empirical targets included threshold probabilities, within-participant standard deviations, the two retained digital autocorrelation measures, and a composite rumination proxy. Depression-tail geometry was represented by the participant-level fractions of observations at or below 0.10 and at or above 0.30 on the 0–1 depression-proxy scale. The rumination proxy was calculated as the equal-weighted mean of depression burden, stress, low positive affect, low mindfulness, expressive suppression, maladaptive coping, and neuroticism after transformation to compatible component scales, and was mapped to the CSDS 0–6 rumination range.

Between-subject heterogeneity was represented only in the cluster-level specifications. For each cluster, the SD across participant-level means was calculated for total platform use, night-time platform use, sleep duration, stress, positive affect, and depression burden:

$s_{k,j}=\mathrm{SD} i\left( x,i,k,j \right)$.

A 95% non-parametric bootstrap confidence interval for $s_{k,j}$ was obtained from 200 participant-level bootstrap samples. The final SD corridor was the bounded outer envelope of this bootstrap interval and a proportional interval with half-width ($0.25*s_{k,j}$).

For stress, positive affect, and depression burden, the 10^th^ and 90^th^ percentiles of the participant-level means were additionally used as lower- and upper-tail shape anchors:

$q_{k,j}^{10}=Q_{0.10}\left( x_{1,k,j},\ldots,x_{n_{k,j}k,j} \right)$,
$q_{k,j}^{90}=Q_{0.90}\left( x_{1,k,j},\ldots,x_{n_{k,j}k,j} \right)$.

For each quantile, 200 participant-level bootstrap samples were generated. The 10th and 90th percentiles of the resulting bootstrap distribution defined the bootstrap interval. The final corridor was the bounded outer envelope of this interval and a proportional band with half-width equal to 20% of the absolute quantile value.

Four fixed structural safety corridors were appended to the specifications of clusters 1 and 3: suicidal-load mean ([0,0.08]), psychosis-load mean ([0,0.10]), mania-load mean ([0,0.12]), and obsessive–compulsive-load mean ([0,0.25]). These were entered as soft safety targets with weight 0.85 and were not estimated from GLOBEM. For clusters 2 and 4, the suicidal-load mean corridor was [0,0.30] with weight 0.45, and [0,0.20] with weight 0.45, respectively.

The resulting target specification was exported programmatically for each cluster. The attached Cluster-1 specification contained eight hard targets and 30 weighted soft targets, including six between-subject SD corridors, six EMA Q10/Q90 shape corridors, four structural safety constraints, and the empirical level, dispersion, threshold, and digital-autocorrelation targets.

#### S9.2 Participant-specific target specifications

Participant-specific target specifications were generated for the 50 participants nearest to the cluster centroid within each of the four clusters. Centroid distance was calculated in the standardised feature space used for clustering. The same scored mapping registry as in the cluster-level analysis was applied, but the target centre was the selected participant’s own observed CSDS-compatible summary rather than a cluster aggregate.

For non-correlation targets, an initial proportional corridor was generated around the participant-level value $x_{i,j}$. The proportional half-width was 10% for level targets, 15% for within-participant dispersion and threshold-probability targets, and 20% for within-participant dynamic targets. All intervals were clipped to the target-specific admissible bounds.

A minimum corridor width was then imposed to avoid degenerate intervals. The minimum width was 10% of the effective target span for level and threshold targets, 8% for dispersion targets, and 5% for dynamic targets. These minimum fractions were multiplied by 1.25 for approximate, partial, proxy-based, weekly-resolution, or non-externally-calibrated mappings. The effective span was the full mapped range for fixed-scale EMA variables, 1 for variables bounded to ([0,1]), at least 2,000 steps for step targets, and at least one unit for sleep and digital-use targets.

For individual correlation-like targets, a two-sided 95% Wald interval was calculated in Fisher-(z) space:
$z_{i,j}=\mathrm{atanh} \left( r_{i,j} \right)$,
$\mathrm{SE} \left( z_{i,j} \right)\frac{1}{\sqrt{\max\left( n_{i,j}-3,1 \right)}}$,
$L_{i,j}=\tanh\left[ z_{i,j}-1.96,\mathrm{SE} \left( z_{i,j} \right) \right]$,
$U_{i,j}=\tanh\left[ z_{i,j}+1.96,\mathrm{SE} \left( z_{i,j} \right) \right]$.

For the retained autocorrelation targets, $n_{ij}$ was the participant’s number of weekly stress observations, with a lower computational floor of four observations. The maximum correlation half-width was 0.35, increased to 0.40 for approximate or partial dynamic mappings. The resulting interval was clipped to ([-1,1]).

Cross-channel coupling targets were absent from the final participant-specific files because they had already been removed from the scored target registry. The export algorithm nevertheless required at least six valid pairs for lag-0 coupling targets and eight valid date-compatible pairs for lag-1 targets when such targets were enabled. Between-subject SD and Q10/Q90 targets were omitted from all participant-specific specifications. The four fixed structural safety corridors were carried over unchanged.

Baseline trait information was used to construct participant-specific ABC-SMC prior ranges rather than phenotype targets. Neuroticism, depression-related anhedonia propensity, self-regulation, resilience, and social need were mapped to the simulator trait coordinate system and assigned symmetric half-widths of 1.00, 0.85, 0.85, 0.75, and 1.25, respectively, clipped to the population range ([-2.5,2.5]). Social-media, gaming, and chat habit parameters retained the full population range ([-2.5,2.5]). These trait ranges were appended to the participant target-specification manifest and read separately by the virtual-twin validator.

### S10 Bayesian inversion and validation framework

#### S10.1 Cluster-level ABC-SMC inversion

Cluster-level CSDS inversion was performed separately for each of the four GLOBEM cluster target specifications. Phenotype fit was evaluated against hard target corridors and weighted soft targets contained in each cluster specification. A candidate failed the active hard gate when at least one hard target fell outside its admissible corridor, whereas deviations from soft-target corridors contributed continuously to the fitting distance.

The active parameter space was constructed automatically from the eligible CSDS parameter paths. Fixed quantities, intervention-specific parameters, blocked namespaces, and parameter paths identified as inactive in the simulation pathway were excluded. For each cluster, the ABC-SMC procedure produced a cloud of 50 parameter configurations (particles). Six scheduled stages were performed: an initial Stage 0 followed by five sequential refinement stages. A separate final hard-target evaluation was conducted after completion of the scheduled stages. At Stage 0, 7,500 parameter configurations were generated by Latin-hypercube sampling from the constrained prior ranges. Each parameter configuration was evaluated using two independent simulation repetitions. Each repetition simulated a population of 30 individuals over 80 days, and the resulting time series were aggregated into the validator phenotype representation before comparison with the empirical cluster specification.

Each simulated configuration produced a hard-gated evaluation distance and a continuous raw distance. The raw distance used for surrogate fitting and proposal guidance was:

$$d_{hard,max}+0.35, d_{\mathrm{soft}}$$

where $d_{hard,max}$was the largest corridor-width-normalised overshoot across the hard targets and $d_{\mathrm{soft}}$ was the weighted mean interval-normalised violation across the soft targets. The maximum hard-target violation determined the primary distance contribution, while the soft-target term differentiated configurations with similar hard-target fit.

The initial tolerance, $\epsilon_{0}$, was defined as the 70^th^ percentile of the finite Stage-0 evaluation distances. The five subsequent refinement stages used $\epsilon$quantiles of 0.45, 0.35, 0.275, 0.225, and 0.20, respectively. Monotonic tightening was enforced so that a scheduled epsilon could not exceed that of the preceding stage.

New parameter configurations were generated by perturbing parent particles from the preceding stage using a principal-component-based multivariate kernel in prior-normalised parameter space. Principal components explaining 90% of the particle-cloud variance were retained, up to a maximum of 100 components. The kernel used a scale factor of 1.5, a minimum marginal kernel fraction of 0.01, and ridge regularisation of 10^-6^.

A linear support-vector regression surrogate was fitted to predict the continuous raw distance from the parameter vector. The surrogate was considered usable when its validation rank correlation exceeded 0.10. At the five refinement stages, enlarged candidate pools with pre-screening factors of 16, 24, 32, 40, and 48 were ranked by the surrogate before full simulation. To preserve exploration outside the surrogate-preferred region, fractions of 0.25, 0.20, 0.15, 0.10, and 0.05 of the simulated candidate pool, respectively, were selected independently of the surrogate ranking. The surrogate was retrained after each stage using accumulated simulation results, with the retained training reservoir limited to 10,000 configurations.

Parameter-level importance derived from the support-vector surrogate was additionally used to scale the perturbation kernel by parameter dimension. Dimensions more strongly associated with phenotype fit received greater relative perturbation, whereas less informative dimensions were attenuated. Relative dimension scaling was bounded below at 0.15 and applied with an exponent of 1.

Parent-particle selection was weighted to preserve diversity in the simulated phenotype space. A multilayer perceptron was trained to predict the continuous raw fit distance from the simulated phenotype summaries. When soft-target paths were available, these defined the phenotype feature vector; non-finite feature values were mean-imputed during normalization. The prediction target was transformed as $\log\left( 1+d_{\mathrm{raw}} \right)$. The model contained two fully connected hidden layers with 64 and 32 units and rectified-linear activations and was trained using Adam for 300 epochs with a learning rate of (10^-3^), mini-batches of 64, and a 15% validation split. A minimum of 50 simulated configurations was required for model fitting, and the model was used for diversity weighting when its validation Spearman correlation with the simulator-derived distance was at least 0.25.

The activations of the second hidden layer provided a 32-dimensional embedding of the simulated phenotype space. Retained particles were projected into this embedding, and local density was estimated using the distance to the ten nearest neighbours. Parent-selection probabilities were proportional to inverse local density with an exponent of 1, thereby increasing the probability of selecting particles from sparsely represented phenotype regions. A uniform weighting component of 0.05 ensured that every retained particle maintained a non-zero probability of selection. The phenotype model was updated after refinement stages using accumulated simulator evaluations when sufficient training data were available.

Surrogate ranking, parameter-specific kernel scaling, and phenotype-diversity weighting influenced candidate generation but not acceptance. Every candidate retained by the ABC-SMC procedure was evaluated through a complete CSDS simulation and assessed against the active hard and soft target specification.

If fewer than 25 Stage-0 configurations passed the complete hard-target specification, hard targets were ranked according to their failure frequency and the most restrictive targets could temporarily be represented as soft constraints with a weight of 2.0. This temporary geometry was used to maintain a viable particle cloud during refinement. Restoration of the complete hard-target geometry was attempted at the end of subsequent stages and required at least 25 of the 50 particles to pass the restored specification. Final hard-decisioning was conducted after completion of the six scheduled stages.

No donor-derived proposals were used during cluster-level inversion. The accepted cluster-level particle clouds were, however, saved to a transfer bank for use as proposal donors during subsequent participant-specific virtual-twin inversion.

The final outputs comprised the retained parameter configurations, final hard-target acceptance status, stage-wise epsilon and acceptance trajectories, weighted soft-target coverage and proximity, parameter-cloud contraction, influential parameter dimensions, and target-specific residual violations. Let ($\phi_{ij}$) denote the realised value of soft target $j$ for final particle $i$, and let ${[L}_{j},U_{j}]$ denote the target corridor. The location of the final particle cloud for target $j$ was defined as $c_{j}=\mathrm{median} i\left( \phi_{\mathrm{ij}} \right)$, or as the weighted median when an aligned particle-weight vector was available. For each evaluable soft target, the in-corridor indicator was $I_{j}=\mathbf{1}\left( L_{j}\leq c_{j}\leq U_{j} \right)$and the distance of the particle-cloud location beyond the nearest corridor boundary was $\delta_{j}=\max\left( L_{j}-c_{j},c_{j}-U_{j},0 \right)$.

The target-specific proximity score was then defined as:

$p_{j}=\max\left[ 0, 1-\frac{\delta_{j}}{U_{j}-L_{j}} \right]$.

Thus, $p_{j}=1$ when the particle-cloud location was inside the target corridor, decreased linearly with the width-normalised distance outside the corridor, and was bounded below at zero. Let $w_{j}$ denote the prespecified soft-target weight and:

$\tilde{w}_{j}=\frac{w_{j}}{\sum_{k\in\mathcal{J}} w_{k}}$,

where $\mathcal{J}$ is the set of evaluable soft targets with finite positive weights. Weighted soft-target coverage and proximity were calculated as:

$C_{\mathrm{target}}=\sum_{j\in\mathcal{J}} \tilde{w}_{j}I_{j}$ and $P_{\mathrm{target}}=\sum_{j\in\mathcal{J}} \tilde{w}_{j}p_{j}$,

respectively. Coverage therefore quantified the weighted proportion of soft-target corridors containing the final particle-cloud location, whereas proximity also retained information about the width-normalised magnitude of residual corridor violations.

Parameter-cloud contraction was evaluated separately for each parameter dimension. Let ($\theta^{\left( 0 \right)}i,j$ denote parameter $j$ in the unselected finite Stage-0 proposal cloud and $\theta^{\left( F \right)}i,j$ its value among the final hard-accepted particles. The contraction ratio for parameter (j) was:

$$r_{j}=\frac{\mathrm{SD} i\left( \theta^{\left( F \right)}i,j \right)}{\mathrm{SD} i\left( \theta^{\left( 0 \right)}i,j \right)}$$

For log-uniform parameters, both standard deviations were calculated after logarithmic transformation. The overall contraction summary was:

$$\overline{r}=\frac{1}{\left| \mathcal{R} \right|}\sum_{j\in\mathcal{R}} r_{j},$$

where $\mathcal{R}$ contained parameter dimensions identified by the analysis as having shifted in marginal location or spread during ABC-SMC refinement. If this relevance set was empty, all dimensions with a finite, non-zero Stage-0 standard deviation were used. The numbers of strongly and weakly contracted dimensions were additionally reported as:

$N_{\mathrm{strong}}=\sum_{j\in\mathcal{R}} \mathbf{1}\left( r_{j}<0.50 \right)$ and $N_{\mathrm{weak}}=\sum_{j\in\mathcal{R}} \mathbf{1}\left( r_{j}>0.80 \right)$

Improvement in continuous phenotype fit was quantified by comparing the mean raw distance of the final scheduled particle cloud with the mean raw distance across all finite Stage-0 proposals before epsilon selection:

$$G_{\mathrm{fit}}=1-\frac{\overline{d}_{raw,final}}{\overline{d}_{raw,Stage,0}}$$

Together, these measures quantified hard-target feasibility, soft-target boundary compliance, narrowing of the mechanistic parameter cloud, and improvement in the continuous phenotype-fitting objective over the initial search landscape.

To illustrate the outcome of the cluster-level inversion process, we visualised the generated time-series for the particles with the smallest fit distance in each GLOBEM cluster (Supplementary Figures S9.1a-d).

#### S10.2 Participant-level virtual-twin inversion and bank transfer

Subject-specific virtual twins were generated from individual target specifications derived by applying the same mapping logic to each observed participant. These subject-level specifications used the participant's own longitudinal summaries and trait-prior information where available. Individual inversion is more difficult than cluster-level inversion because the target corridors are narrower and subject trajectories may be atypical relative to the cluster centroid.

To improve search efficiency, the transfer-VT implementation used accepted particles and stage-bank information from prior group-level inversions as donor material. Donor selection was based on target-descriptor similarity and masked target-path overlap; exact target-geometry identity was treated only as a bonus, not as a compatibility requirement. At Stage 0, a configurable fraction of the design could be replaced by donor-derived proposals; in later stages, candidate pools mixed donor, local-posterior and fresh prior proposals, with donor influence decaying across early stages and a prior stream retained to reduce collapse toward donor archetypes. Critically, transfer modified only proposal generation and warm starts. Acceptance remained subject-specific and simulator-based, using the individual target specification. This design is consistent with digital-twin logic in precision medicine, where mechanistic simulations are constrained by individual data but should remain falsifiable against the target phenotype.

#### S10.3 Parameter-space audit of expanded cluster-level particle clouds

After fitting the four cluster specifications, the 50 accepted particles per cluster yielded 200 retained cluster-level particles. We examined whether the four resulting particle ensembles were distinguished by latent mechanisms organised in parameter families such as regulatory capacity, digital allocation, sleep-dysrhythmia sensitivity, strain coupling, or psychiatric tipping thresholds. To this end, the retained particle clouds were expanded from 200 to 2,200 particles and evaluated with the CSDS particle auditor. This step tested whether the CSDS could produce a mechanistically informative manifold in which cluster-level phenotypes mapped onto separable regions of the simulator’s latent dynamical and coupling space, rather than merely onto different means of observed variables.

The particle auditor re-simulated particles from the expanded cloud and extracted mechanistic signatures from the simulated trajectories. The audited features included symptom-anchor severity, volatility, variance, skewness, lag-1 autocorrelation, reversion or recovery-speed proxies, unpredictable residual fluctuation, modal structure, and cross-domain coupling slopes linking digital behaviour, sleep, activity, and symptom-anchor change. Digital-sensitivity summaries and exposure–symptom lag descriptors were also computed. Particle-level signatures were averaged across repetitions and then used for mechanistic profiling and cross-cluster comparison. The 30 highest-ranked parameters, with their effect sizes, dominant-ensemble contrasts, and functional roles in the CSDS, are listed in Supplementary Table S10.3.

### S11 Virtual-twin-based latent-state decoding

#### S11.1 Analytical design and synthetic training data

The decoding analysis examined whether participant-specific CSDS virtual-twin simulations contained a latent-state structure that could be transferred to the corresponding observed GLOBEM time series. Fifty observed participants were analysed within each GLOBEM cluster. For every participant, 20 repeated simulations were generated from each of 50 accepted virtual-twin particles, yielding 1,000 participant-specific synthetic trajectories. Separate latent-state decoders were trained for each participant on these synthetic trajectories and subsequently applied to the corresponding observed behavioural, ecological momentary assessment (EMA), and wearable-derived time series.

The primary analysis used the dense synthetic simulation output to retain the information contained in the continuously simulated level trajectories. Channel means and standard deviations were estimated exclusively from each participant’s dense synthetic training trajectories and applied to both the synthetic and observed sequences:

$$z_{t,d}=\frac{x_{t,d}-\mu_{d}^{\mathrm{syn}}}{\sigma_{d}^{\mathrm{syn}}}$$

Observed data therefore contributed neither to estimating the feature-scaling parameters nor to fitting the latent-state models.

#### S11.2 Decoder features and handling of irregular observations

The decoder representation combined standardized channel levels with lagged change features. For dense synthetic trajectories and daily passive or wearable-derived channels, changes were calculated relative to the preceding day. For irregularly sampled observed EMA channels, the change at an assessment was calculated relative to the immediately preceding available assessment and assigned to the day of the later assessment. Accordingly, observed PANAS changes generally spanned 3–4 days, whereas PHQ-4 and stress changes generally spanned approximately one week. These changes were not divided by the elapsed interval. EMA and passive or wearable-derived domains received equal feature weights in the primary analysis.

Observed time points were retained on their original daily grid. The adequacy of recent measurement support was assessed separately for each channel using a causal seven-day rolling window containing the current and preceding days. Most channels were considered supported when observations were available on at least 50% of this window. The threshold was reduced to 25% for lower-frequency EMA channels and was 50% for positive affect. A time point was eligible for state assignments when at least three channels met their respective support thresholds and at least one current raw-channel value was available.

Contiguous eligible time points formed assignable temporal blocks. Individual missing features within an otherwise assignable time point were accommodated by omitting their contribution from the emission likelihood rather than imputing their values. Unsupported time points were retained on the temporal grid but received no latent-state assignment.

#### S11.3 Mixed-emission latent-state models

Two participant-specific state models were examined. Both used mixed emissions: continuous level and change features were represented by state-specific diagonal-Gaussian distributions, while configured ordinal EMA variables were represented by state-specific categorical distributions with smoothed category probabilities. The sticky hidden Markov model assumed a first-order Markov sequence of latent states. Temporal persistence was encouraged by assigning additional prior mass to self-transitions before normalising the transition probabilities.

We also used a sticky autoregressive HMM (AR-HMM) to extend the observation model by including state-specific first-order autoregressive dynamics. Its latent states therefore differed not only in their characteristic multivariate levels but also in how current observations depended on preceding observations. This model was distinct from the conventional sticky HMM with a separate autoregressive forecasting head used later in the forecasting analysis: in the AR-HMM, autoregressive dynamics contributed directly to latent-state estimation.

Model parameters were estimated from the dense synthetic trajectories using expectation–maximisation. Continuous emission parameters were updated from posterior-weighted Gaussian moments, ordinal probabilities from posterior-weighted category counts, and transition probabilities from posterior transition counts.

#### S11.4 Selection of the number of states

For each participant and decoder family, the number of states was selected from $K=4,\ldots,8$ using particle-grouped cross-validation. All repeated simulations generated from the same accepted particle were assigned jointly to either the training or validation partition, thereby preventing closely related trajectories from appearing in both partitions. Three cross-validation repetitions were performed using 60% of the particle groups for training. At most ten simulated trajectories per particle group were included in the state-number selection procedure.

For each candidate $K$, independently fitted training and validation solutions were aligned and compared in terms of:

1. The relative occupancy of corresponding states;
2. The off-diagonal transition structure describing switching between states; and
3. The geometry of the multivariate state fingerprints.

Candidate solutions were required to maintain a minimum state occupancy of 5% in both partitions. The final state number was the smallest $K$ whose mean reproducibility score was within 0.02 of the best valid candidate, thereby favouring the least complex solution with essentially equivalent reproducibility.

#### S11.5 State-path decoding and cross-participant state alignment

Observed assignable blocks were decoded using the most likely Viterbi state path. A minimum state-segment duration of two days was imposed using a duration-constrained Viterbi recursion with strict boundary handling. This constraint reduced isolated one-day state assignments but did not introduce an explicit state-duration distribution.

Because a separate state model was fitted for every participant, local state labels were subsequently aligned to an expandable cross-participant state registry. Alignment was based on Spearman correlations between state fingerprints estimated from the dense synthetic trajectories. A local state was assigned to an existing registry state when its fingerprint correlation was at least 0.50. Otherwise, it could provisionally define a new registry state. Registry states were retained when they occurred in at least three participant-specific virtual-twin models and in at least 10% of models and had a mean occupancy of at least 2%. This procedure enabled structurally corresponding states to be compared across participants without requiring every participant to express the same state repertoire.

An interpretation of the cluster-specific state taxonomies can be found in **Supplementary Table S11.5**. States are visualised using raw channel-value levels in **Supplementary Figure S11.5a** and using VT-normalized HMM fingerprints in **Supplementary Figure S11.5b**.

#### S11.6 Assessment of synthetic-to-observed state transfer

Decoder validity was assessed by determining whether the state structure learned from the synthetic trajectories was recovered when the decoder was applied to the observed data. Synthetic and observed trajectories were independent realizations and were not temporally synchronized; consequently, this analysis did not test point-by-point correspondence. Instead, structural transfer was evaluated at three complementary levels.

State-occupancy correlation quantified whether corresponding latent states occurred with similar relative frequencies in the synthetic and observed sequences. Transition correlation quantified whether the observed sequence reproduced the synthetic organization of state persistence and switching, based on the correspondence between their transition matrices. State-fingerprint correlation quantified whether corresponding states retained similar multivariate profiles across behavioural, EMA, and wearable-derived features. State fingerprints were defined by the mean standardized feature values within each decoded state.

Together, these measures assessed whether the synthetic decoder recovered in the observed data a comparable state repertoire, a comparable temporal organization of those states, and comparable state-specific phenotypic profiles. Additional distance-based diagnostics for occupancy and transition matrices were retained as secondary quality-control measures.

### S12 Causal forecasting of behavioural and symptom trajectories

#### S12.1 Rolling-origin forecast design

Forecast validation used a causal rolling-origin design. For each participant, forecasts were generated repeatedly at successive eligible time points in the observed time series. At a forecast time point (t), only observations collected up to and including (t) were available to the model. Later observations were withheld and used exclusively to evaluate predictions at horizons $h\in1, 3, 7, 14, 21 \text{days}.$

Forecast generation began once 21 aligned calendar days of observed history were available. This criterion refers to the retained daily grid rather than requiring 21 fully observed or state-assignable days. Unsupported observations within the available history were retained as non-informative time points, allowing the state process to evolve across periods of missing observation support.

All forecasting architectures and training regimes used the same standardized decoder features, support masks, raw target values, forecast horizons, and eligible forecast time points. No feature construction or standardisation was repeated within the forecasting component.

#### S12.2 Synthetic and hybrid forecasting regimes

In the synthetic regime, latent-state, emission, transition, and forecasting parameters were estimated exclusively from the participant-specific virtual-twin trajectories and remained fixed during application to the observed data. At every forecast time point, the participant’s observed history up to that day was passed through the fixed synthetic decoder to estimate the current probability distribution over latent states. Thus, the observed history individualized the starting state of the forecast without contributing to model-parameter estimation. For forecasting models with autoregressive components, the most recent usable observed level and change additionally provided the initial numerical conditions.

In the hybrid regime, the number and identities of the synthetic latent states were retained. The causally available observed history was used to calibrate the synthetic emission distributions, transition probabilities and, where applicable, autoregressive coefficients. Calibration used state-posterior-weighted observed estimates that were shrunk towards their synthetic counterparts:

$$\hat{\theta}_{hyb}=\frac{w_{syn}\hat{\theta}_{syn}+N_{eff}\hat{\theta}_{obs}}{w_{syn}+N_{eff}}$$

where $w_{syn}$ is the synthetic prior weight and $N_{eff}$ is the effective observed sample size for the relevant state, channel, transition row, or regression cell. Parameters were adapted only when the observed history met the pre-specified minimum effective sample size; otherwise, the corresponding synthetic parameter was retained.

In the production analysis, synthetic prior weights of 5 were used for emission, transition, and autoregressive adaptation. Emission and transition updates required an effective observed sample size of at least 3, as did observed-prefix adaptation of autoregressive coefficients. A variance floor of (10^-4^) was applied during Gaussian emission adaptation. All settings unrelated to this adaptation were held constant between the paired synthetic and hybrid models.

#### S12.3 Forecasting architectures

##### S12.3.1 Sticky HMM state-emission forecast

For the conventional sticky HMM, the filtered state-probability vector at forecast time $t$, $\boldsymbol{\pi}_{t}$, was propagated through the state-transition matrix $A$:

$\boldsymbol{\pi}_{t+h}=\boldsymbol{\pi}_{t}A^{h}$.

For channel $d$, the forecast was the probability-weighted expectation of the state-specific raw-channel means:

$\hat{y}_{t+h,d}=\sum_{k=1}^{K} \pi_{t+h,k}\mu_{k,d}^{\mathrm{raw}}$.

For ordinal targets, state-specific expected category values were used. In the hybrid regime, the emission distributions and transition probabilities were first calibrated from the available observed history and then used in the same state-propagation procedure.

##### S12.3.2 Sticky HMM with a state-conditioned autoregressive forecasting head

The second architecture retained the conventional sticky HMM as the latent-state decoder and added a direct, horizon-specific autoregressive forecasting head. For each state $k$, horizon $h$, and output channel $d$, the dense synthetic trajectories were used to estimate

$$y_{d}\left( t+h \right)=\beta_{0,k,h,d}+\beta_{1,k,h,d}y_{d}\left( t \right)+\beta_{2,k,h,d}\Delta y_{d}\left( t \right)+\varepsilon_{k,h,d}$$

The predictors were the current channel value and its most recent available change. The regressions were estimated by ridge regression with penalty ($\lambda$=0.01) and required an effective synthetic sample size of at least 10. Soft probabilities of the latent state at the future target time were used as regression weights. During forecasting, state-specific predictions were combined according to the propagated future-state probabilities.

In the synthetic regime, the regression coefficients remained fixed at their synthetic estimates. In the hybrid regime, corresponding regressions were estimated from the causally available observed history when sufficient support existed and were shrunk towards the synthetic coefficients. Because these regressions were horizon-specific, adaptation at longer horizons became available only when the observed history contained sufficient predictor–outcome pairs for that horizon.

##### S12.3.3 Native sticky AR-HMM forecast

The native sticky AR-HMM represented state-specific autoregressive dynamics within the latent-state observation model itself. Multi-step forecasts were generated recursively by jointly propagating the future state distribution and the state-specific autoregressive expectations. Autoregressive slopes were constrained to an absolute magnitude below 0.98 to limit unstable extrapolation.

A current observed channel value was used as the autoregressive starting value only when it corresponded to the forecast time point and was no more than 0.5 days from it. Otherwise, the probability-weighted state mean was used as the starting value. In the hybrid regime, the emission, transition, and state-specific autoregressive parameters were calibrated from the observed history while retaining the synthetic state identities.

The native AR-HMM and the sticky-HMM autoregressive head therefore represented distinct modelling strategies. In the native AR-HMM, temporal dependence was part of both state inference and forecasting. In the autoregressive-head model, latent states were first inferred by the conventional sticky HMM, and the separate regression layer was used only to forecast future channel values.

#### S12.4 Causal reference forecasts

Model predictions were compared with two causal reference forecasts constructed on the same participant, channel, forecast time point, and horizon grid. The observed-history-mean forecast predicted that the future value would equal the mean of all finite observations available for that participant and channel up to the forecast time point. The persistence forecast predicted that the latest finite observed value would remain unchanged.

A participant-specific mean calculated from the complete observed time series was retained only as a deliberately non-causal diagnostic reference. It was used to estimate how much apparent forecasting performance could be explained by prior knowledge of each participant’s overall level and was excluded from the primary causal model comparisons and inferential tests. Null-relative skill was calculated from matched squared errors: $\mathrm{Skill}_{\mathrm{null}}=1-\frac{\mathrm{SSE}_{\mathrm{model}}}{\mathrm{SSE}_{\mathrm{null}}}$. Positive values indicated lower squared prediction error than the reference forecast.

#### S12.5 Monitoring-period-matched skill on common evaluation support

To determine whether the widening advantage of the decoders over the causal monitoring-period-mean null at longer horizons reflected genuine capture of subject-specific drift rather than horizon-dependent attrition of evaluable series, skill was recomputed on a fixed set of evaluation points common to all horizons. For each decoder, target channel and cluster, the set of participant–evaluation-point keys jointly evaluable by both the decoder and the monitoring-period-mean null was intersected across every available horizon, retaining only evaluation points present at all horizons. On this horizon-invariant support, forecasts were matched to the monitoring-period-mean null at the identical participant, channel, evaluation point and horizon, and skill was defined as 1 − SSE_model_/SSE_null_. Because the evaluated origins and monitoring-period lengths were thereby held constant across horizons, any residual horizon dependence of skill could not be attributed to shortening of the evaluable monitoring period. Results are reported per channel and as a channel-balanced overview (**Supplementary Table S12.5**).

#### S12.6 Global and within-participant forecast performance

Performance was first calculated separately for each cluster, target channel, decoder architecture, training regime, and forecast horizon. Channel-balanced overview estimates were subsequently obtained as unweighted means of the valid channel-specific estimates.

Let $y_{s,a}$ and $\hat{y}_{s,a}$ denote the observed and predicted values for participant $s$ at eligible forecast time point $a$, for a fixed target channel and forecast horizon. The global total-variation prediction coefficient was:

$R_{pred,global}^{2}=1-\frac{\sum_{s} \sum_{a} \left( y_{s,a}-\hat{y}_{s,a} \right)^{2}}{\sum_{s} \sum_{a} \left( y_{s,a}-{\bar{y}.}_{\cdot\cdot} \right)^{2}}$.

This coefficient evaluated prediction error relative to the total variation across participants and forecast time points. It therefore reflected both differences in average level between participants and temporal variation within participants; because it was based on prediction error rather than squared correlation, it could take negative values.

Prediction of within-participant temporal variation was assessed using forecast skill relative to a causal participant-specific prefix-mean baseline. At each forecast origin, the baseline predicted the target from the mean of the participant’s observations available up to that time. Let $\hat{y}_{s,a}^{\mathrm{PM}}$ denote this prefix-mean prediction, and let $\mathcal{E}_{h}$ denote the set of participant–anchor pairs for which both the evaluated model and the prefix-mean baseline produced valid forecasts at horizon $h$. Within-participant forecast skill was defined as:

$$\mathrm{Skill}_{\mathrm{within},h}=1-\frac{\sum\left( s,a \right)\in\mathcal{E}_{h}\left( y_{s,a}-\hat{y_{s,a}} \right)^{2}}{\sum\left( s,a \right)\in\mathcal{E}_{h}\left( y_{s,a}-\hat{y}_{s,a}^{\mathrm{PM}} \right)^{2}}$$

Thus, $\mathrm{Skill}_{\mathrm{within},h}>0$ indicated that the forecasting model reduced squared prediction error relative to the participant’s causal prefix mean, $\mathrm{Skill}_{\mathrm{within},h}=0$ indicated equal performance, and $\mathrm{Skill}_{\mathrm{within},h}>0$ indicated poorer performance than the prefix-mean baseline. Because the denominator was the error of a participant-specific causal baseline, the measure quantified predictive information beyond the participant’s previously observed average level.

Channel-specific skill estimates were obtained by pooling squared errors over the matched participant–anchor observations. For comparisons across forecast horizons and model configurations, calculations were restricted to common eligible anchor support. Cross-channel averages with 95%-CIs of $R_{pred,pool}^{2}$ and $\mathrm{Skill}_{\mathrm{within},h}$were depicted in **Figure 5A** and **5B** of the main manuscript. Channel-specific plots are displayed cluster-wise in **Supplementary Figures 12.6a** and **12.6b**.

#### S12.7 Prediction of change relative to the observed history

To evaluate future change rather than only future level, each model forecast was matched to the observed-history-mean forecast for the same participant, channel, forecast time point, and horizon. Observed and predicted deviations were defined as:

$$\Delta_{s,a,h,d}^{obs}=y_{s,a+h,d}\bar{y}_{s,a,d}^{\mathrm{history}},$$

$$\Delta_{s,a,h,d}^{pred}=\hat{y}_{s,a+h,d}\bar{y}_{s,a,d}^{\mathrm{history}}.$$

Change-direction balanced accuracy was reported to assess models’ performance in correctly predicting future values above and below the preceding participant-specific mean, indicating whether the observed and predicted deviations had the same sign (**Supplementary Table S12.7a**). Additionally, correspondence between observed and predicted deviations was summarized using Spearman correlations (**Supplementary Table S12.7b**). Cross-channel averages with 95%-CIs of change-direction balanced accuracy are shown in **Figure 5C** of the main manuscript. Channel-specific plots are displayed cluster-wise in **Supplementary Figure 12.7**.

#### S12.8 Tracking of longer-term temporal slopes (drifts)

For every participant, target channel, forecasting model, training regime, and horizon, observed and predicted deviations from the causal history mean were separately regressed on forecast time using ordinary least squares. At least three valid observations at three distinct forecast time points were required. The resulting participant-specific observed and predicted slopes were compared across participants using Spearman correlation and direction agreement (**Supplementary Tables S12.8a** and **S12.8b**).

#### S12.9 Bootstrap confidence intervals

Uncertainty was quantified using 5000 subject-block bootstrap resamples with a fixed random-number seed recorded in the analysis script. Participants were sampled with replacement, and all forecast time points, channels, horizons, and repeated observations belonging to a selected participant were sampled together. This preserved the dependence among repeated forecasts from the same participant.

The relevant statistic was recomputed in every bootstrap sample. Percentile 95% confidence intervals were defined by the 2.5^th^ and 97.5^th^ percentiles of the valid bootstrap estimates. Subject-block confidence intervals were calculated for $R_{pred, global}^{2}$, the within-participant skill metric, balanced directional accuracy, deviation correlations, slow drift correlations, and slope-sign agreements.

#### S12.10 Qualitative visualisation of forecasting results

The quantitative analysis of models’ forecasting performance was supplemented by a visualisation of the observed vs. predicted time series of the four GLOBEM individuals that were most closely positioned to the four cluster centroids. Absolute and change predictions for the next-day and 21-day horizons provided by the hybrid AR-HMM forecaster were overlaid on the observed signals of the four study participants in **Supplementary Figures S12.10a** and **S12.10b**.

#### S12.11 Statistical comparison of forecasting models

Formal model comparisons were performed separately for each target channel and for participant-local $R_{\mathrm{pred}}^{2}$ and NRMSE. A fully within-participant factorial repeated-measures analysis of variance included the factors:

$\text{decoder architecture} \left( 3 \text{levels} \right)\times\text{training regime}\text{ }\left( 2 \text{levels} \right)\times\text{forecast horizon} \left( 5 \text{levels} \right)$.

The decoder levels were the sticky HMM, sticky HMM with autoregressive forecasting head, and native sticky AR-HMM. Training regime comprised synthetic and hybrid models, and forecast horizon comprised 1, 3, 7, 14, and 21 days. Main effects and all two- and three-way interactions were evaluated.

Each target-specific analysis included only participants with finite metric values in every decoder-by-regime-by-horizon cell, resulting in a balanced complete-case repeated-measures design. F statistics were computed from within-participant projection matrices. Conventional $\eta^{2}$ and partial $\eta^{2}$ were reported as effect-size measures.

Significant effects were decomposed using paired two-sided comparisons of decoder architectures within each training-regime-by-horizon cell and of training regimes within each decoder-by-horizon cell. The mean and median paired differences and standardized paired effect size $d_{z}$ were reported. Holm adjustment was applied to the corresponding families of omnibus and post hoc tests.

Horizon-specific Friedman tests and paired signed-rank comparisons were retained as distribution-free sensitivity analyses. The channel-balanced pooled overview metrics were reported descriptively with bootstrap confidence intervals; the current analysis did not fit a separate inferential model to this global channel-balanced endpoint.

#### S12.12 Virtual-twin oracle decomposition of AR-HMM forecast attenuation

All HMM implementations produced increasingly attenuated point forecasts at longer forecast horizons. We therefore conducted a diagnostic oracle analysis of the AR-HMM using held-out virtual-twin trajectories to determine whether this attenuation resulted predominantly from uncertainty in the future latent-state sequence, recursive propagation of the autoregressive component, an interaction between these mechanisms, or residual limitations that remained after both sources of uncertainty were removed.

Oracle forecasts were used exclusively for mechanism analysis. Because they used information from the future held-out trajectory, they were not included in the primary prospective forecasting comparisons and were not applied to the observed GLOBEM trajectories.

##### S12.12.1 Audit design

The audit was performed for the Cluster 2 subject-specific native AR-HMMs. The selected number of behavioural states was retained for each participant, but the AR-HMM was refitted within each audit repetition using only the designated training virtual twins.

Virtual-twin trajectories were partitioned by accepted-particle group. Repeated simulations generated from the same accepted particle shared the same cross-validation fold and were assigned jointly to either the training or held-out partition. This prevented closely related trajectories from occurring on both sides of the split. Eighty per cent of particle groups were used for fitting and the remaining groups for held-out evaluation.

Forecasts were issued after a minimum 35-day prefix at horizons of 1, 3, 7, 14, and 21 days. Mechanism classification was based primarily on the 7-, 14-, and 21-day horizons. Participants were added sequentially, with interim analyses after 5, 7, 9, 11, and 13 participants. The final audit included 13 participants, with the participant treated as the inferential unit.

##### S12.12.2 Standard AR-HMM forecast

For participant $i$, target channel $c$, state $k$, forecast origin $t$, and future step $u$, the state-specific AR(1) prediction was:

$$\hat{y}_{i,t+u,c,k}=a_{i,k,c}+b_{i,k,c}l_{i,t+u-1,c,k}$$

The autoregressive slopes were bounded according to:

$$\left| b_{i,k,c} \right|\leq0.98$$

In the standard forecast, the future state probabilities were propagated from the causally filtered state distribution through the fitted transition matrix. Each state-specific trajectory recursively reused its own preceding prediction:

$$\hat{y}_{i,t+u,c,k}^{\left( \mathrm{std} \right)}=a_{i,k,c}+b_{i,k,c}\hat{y}_{i,t+u-1,c,k}^{\left( \mathrm{std} \right)}$$

The final point forecast was the probability-weighted expectation across states:

$$\hat{y}_{i,t+u\mid t,c}^{\left( \mathrm{std} \right)}=\sum_{k=1}^{K} \hat{p}_{i,t+u,k}\hat{y}_{i,t+u,c,k}^{\left( \mathrm{std} \right)}$$

where $\hat{p}_{i,t+u,k}$denotes the transition-propagated probability of state $k$.

##### S12.12.3 Counterfactual oracle conditions

Four matched forecasting conditions were evaluated. All conditions used the same fitted AR-HMM parameters, held-out trajectory, forecast origin, target observation, and horizon. They differed only in the future state or lag information supplied to the forecasting recursion.

##### S12.12.4 Future-state oracle

The future-state oracle replaced the transition-propagated future state distribution with the full-sequence Viterbi state obtained by applying the fitted AR-HMM to the complete held-out trajectory. Let $z_{i,t+u}^{*}$denote the corresponding future Viterbi state. Its one-hot state distribution was:

$$p_{i,t+u,k}^{\left( S \right)}=\mathbf{1}\left( k=z_{i,t+u}^{*} \right)$$

The forecast was propagated recursively along this future state path:

$$\hat{y}_{i,t+u\mid t,c}^{\left( S \right)}=a_{i,z_{i,t+u}^{*},c}+b_{i,z_{i,t+u}^{*},c}\hat{y}_{i,t+u-1\mid t,c}^{\left( S \right)}$$

This condition removed uncertainty about future AR-HMM regime occupancy while retaining recursive autoregressive prediction. The Viterbi path represented the fitted AR-HMM’s own full-sequence state partition. It was not interpreted as a ground-truth mechanistic state sequence.

##### S12.12.5 Future-lag oracle

The future-lag oracle retained the standard transition-propagated future state probabilities but supplied the actually realised preceding value from the held-out trajectory as the autoregressive lag:

$$\hat{y}_{i,t+u\mid t,c}^{\left( L \right)}=\sum_{k=1}^{K} \hat{p}_{i,t+u,k}\left( a_{i,k,c}+b_{i,k,c}y_{i,t+u-1,c} \right)$$

Because the contemporaneous value at the forecast origin was already causally available to the standard model, the lag oracle could provide additional information only beyond the first forecast step. This condition removed recursive lag contraction and propagation of preceding point-prediction errors while retaining uncertainty about future state occupancy.

##### S12.12.6 Combined state-and-lag oracle

The combined oracle supplied both the future Viterbi state and the realised preceding value:

$$\hat{y}_{i,t+u\mid t,c}^{\left( SL \right)}=a_{i,z_{i,t+u}^{*},c}+b_{i,z_{i,t+u}^{*},c}y_{i,t+u-1,c}$$

This was the strongest oracle condition available without modifying the fitted state-specific emission or autoregressive parameters.

##### S12.12.4 Oracle gains

For forecast mode $m$, mean-squared prediction error was calculated within each participant, split repetition, channel, and horizon:

$$\mathrm{MSE}_{i,r,c,h}^{\left( m \right)}=\frac{1}{n_{i,r,c,h}}\sum_{t\in\mathcal{A}_{i,r,c,h}} \left( y_{i,t+h,c}-\hat{y}_{i,t+h\mid t,c}^{\left( m \right)} \right)^{2}$$

Here, $\mathcal{A}_{i,r,c,h}$denotes the common set of valid forecast origins.

The relative oracle gain was:

$$G_{i,r,c,h}^{\left( m \right)}=\frac{\mathrm{MSE}_{i,r,c,h}^{\left( \mathrm{std} \right)}-\mathrm{MSE}_{i,r,c,h}^{\left( m \right)}}{\mathrm{MSE}_{i,r,c,h}^{\left( \mathrm{std} \right)}}$$

Positive values indicated lower prediction error than the standard AR-HMM forecast. The additional gain obtained by combining both forms of oracle information was:

$$J_{i,r,c,h}=G_{i,r,c,h}^{\left( SL \right)}-\max\left( G_{i,r,c,h}^{\left( S \right)},G_{i,r,c,h}^{\left( L \right)} \right)$$

A positive $J$ indicated that neither future-state knowledge nor future-lag knowledge alone captured the complete recoverable improvement.

##### S12.12.5 Forecast attenuation

Attenuation was evaluated using excursions from the causally available prefix mean:

$$\Delta y_{i,t,h,c}=y_{i,t+h,c}-\bar{y}_{i,t,c}^{\left( \mathrm{prefix} \right)}\hat{\Delta y}_{i,t,h,c}^{\left( m \right)}=\hat{y}_{i,t+h\mid t,c}^{\left( m \right)}-\bar{y}_{i,t,c}^{\left( \mathrm{prefix} \right)}$$

The primary attenuation measure compared the dispersion of predicted and observed excursions:

$$A_{i,r,c,h}^{\left( m \right)}=\frac{\mathrm{SD}\left( \hat{\Delta y}_{i,t,h,c}^{\left( m \right)} \right)}{\mathrm{SD}\left( \Delta y_{i,t,h,c} \right)}$$

Values below one indicated attenuated forecast excursions, whereas values near one indicated matched forecast and observed dispersion. IQR-based and mean-absolute-excursion ratios were calculated as robustness measures.

##### S12.12.6 Aggregation and mechanism classification

Oracle gains were first calculated within participant × split × channel × horizon cells. For each participant and channel, gains were aggregated by taking the median across the primary 7-, 14-, and 21-day horizons and subsequently across split repetitions. Cohort summaries were calculated as medians across participants. 95%-CIs were estimated from 1,000 participant-level bootstrap resamples.

A relative MSE improvement of 0.15 was considered meaningful, and a difference of 0.10 between the state- and lag-oracle gains was considered evidence of dominance.

Channels were classified as state-uncertainty dominant when the state-oracle gain was meaningful and exceeded the lag-oracle gain by at least 0.10, and as recursive-AR dominant when the reverse pattern was observed. A mixed state-AR classification was assigned when both components contributed materially to the combined gain. A mixed synergistic classification was assigned when the combined oracle produced meaningful improvement although neither single oracle independently reached the meaningful-gain threshold. Channels were classified as showing an oracle-resistant residual limitation when the combined oracle produced little improvement or when substantial attenuation remained despite both oracle inputs. Remaining cases were classified as inconclusive.

The oracle-resistant category was not interpreted as pure observation noise. It could also reflect limitations of the AR-HMM state partition, state-conditional autoregressive or emission structure, simulator-to-decoder mismatch, or unrepresented exogenous influences.

A strong future-state oracle gain indicated that uncertainty in future regime occupancy contributed materially to prediction attenuation. A strong future-lag oracle gain indicated that recursive autoregressive contraction or propagation of preceding point-prediction errors was the principal source. Additional improvement under the combined oracle indicated interaction between state-path and autoregressive uncertainty.

### S13 Information Leakage and VT Robustness Analysis

Although forecasting was performed causally using only observations available up to each forecast origin, participant-specific virtual-twin target specifications were derived from the complete longitudinal series. Future observations could therefore indirectly influence the anchors and tolerance corridors used for virtual-twin generation. We assessed this potential leakage pathway by recomputing target specifications using only observations available before a prespecified temporal cutoff and comparing them with the corresponding full-series specifications.

For each eligible participant, the calendar span was divided at its temporal midpoint. Eligibility required at least 56 total observation days, at least 28 days after the cutoff, at least 40 passive-sensing days and at least six EMA measurements. The balanced analysis included 20 participants from each of the four clusters, selected using a within-cluster quality score combining observation-span length, passive-data density and EMA density.

First-half and full-series targets were paired by study, participant, cluster and target metric. For each pair, we calculated the signed and absolute relative anchor displacement, the first-half to full-series corridor-width ratio, corridor-midpoint displacement relative to the full-series corridor width, and the Jaccard interval overlap. The primary leakage indicator was the corridor-breach rate, defined as the proportion of full-series anchors falling outside the corresponding first-half corridor. Analyses focused primarily on the hard level targets used to constrain individual model inversion. Low-weight soft dispersion, threshold and simple autocorrelation targets were examined separately using their ABC-SMC weights. Fixed safety priors were excluded from weighted summaries.

The signed relative anchor displacement was defined as:

$$r_{i,j}=\frac{a_{i,j}^{\mathrm{half}}-a_{i,j}^{\mathrm{full}}}{\max\left( \left| a_{i,j}^{\mathrm{full}} \right|,\epsilon\right)}$$

where $a_{i,j}^{\mathrm{half}}$and $a_{i,j}^{\mathrm{full}}$denote the first-half and full-series anchors for participant $i$and target $j$, and $\epsilon$prevents division by zero. Group-level shifts were evaluated for the complete sample and separately within each cluster using Wilcoxon signed-rank tests, which account for both the direction and ranked magnitude of paired changes, complemented by exact binomial sign tests based on participant-level summaries. Uncertainty was quantified using 5,000 participant-level bootstrap samples, stratified by cluster and retaining all targets belonging to each sampled participant. Statistical tests were considered diagnostic and therefore not corrected for multiple comparisons. Results are presented in **Supplementary Tables S13a** and **S13b**.

To determine whether restricting target construction to the first temporal half materially altered the inferred virtual twins or their downstream dynamical properties, we additionally repeated participant-level VT inversion for the selected participants in clusters 1 and 2 using only the corresponding first-half target specifications. Split-half VTs and their matched full-series counterparts were simulated over the complete observed time span so that subsequent decoding and forecasting analyses operated on synthetic trajectories of identical duration. Downstream analyses used the same participants, observed data, channel definitions and analysis settings within each cluster.

Differences between the split-half and full-series VT posteriors were quantified using the particle-audit procedures described above (Supplementary Methods S10.3). For each participant, the 50 accepted particles from both inversions were compared across the 1,644 parameter paths common to the two posterior ensembles. We assessed correspondence of posterior medians and dispersion, standardized shifts in parameter medians, and separation of the two particle populations relative to their within-cloud nearest-neighbour distances. These analyses evaluated whether the reduction in calibration information resulted in materially different admissible parameter ensembles.

Functional preservation was subsequently assessed using the latent-state decoding framework described in Supplementary Methods S11. Split-half and full-series VT trajectories were analysed using identical sticky AR-HMM production settings, and synthetic-to-observed correspondence was compared for state occupancy, transition structure and multivariate state fingerprints. The comparison therefore tested whether differences in the inferred VT parameter ensembles propagated into differences in the latent behavioural-state organization recovered from the observed data.

Finally, the same matched split-half and full-series VTs were evaluated using the causal rolling-origin forecasting framework described in Supplementary Methods S12. Forecasts were generated at 1-, 3-, 7-, 14- and 21-day horizons using identical target channels, model architecture, causal observed histories, null comparators and quality-control criteria. The primary comparison used forecast skill relative to the participant-specific causal prefix mean on matched eligible forecast observations. To quantify functional preservation, split-half forecast skill was expressed relative to the corresponding full-series VT skill within each cluster and horizon. Synthetic and hybrid-online regimes were examined separately. Because the split-half and full-series analyses used the same observed trajectories and, after quality control, the same eligible participants within each cluster, differences in forecasting performance primarily reflected differences in the VT calibration and synthetic training data rather than changes in the downstream evaluation set.

Posterior, decoding and forecasting comparisons are summarized in Supplementary Table S13c and Supplementary Figure S13.

### Supplementary Tables

**Supplementary Table S1.1a. Trait coordinates and physiological anchors.** Trait coordinates are randomly sampled at the start of the model inversion. Z-scaled coordinates are clamped to [−2.5, 2.5]. Sleep need is sampled in hours, and the age-referenced activity anchor in measured in steps/day. Traits are transformed using $\sigma(x) = 1/(1 + e^{-x})$.

| **Coordinate** | **Reference mean** | **Sampling SD** | **Support / bounds** | **Canonical transform** | **Representative uses and evidence** |
| --- | --- | --- | --- | --- | --- |
| **PERSONALITY DIMENSIONS** | | | | | |
| neuroticism | 0 | 1.0 | [−2.5, 2.5] (z) | nn01 = σ(z) | Stress, rumination and negative-affect sensitivity; strain, sleep and HRV initialization; psychiatric vulnerability; BPD composite; raw-z EMA and loneliness gains |
| reward_sens | 0 | 1.0 | [−2.5, 2.5] (z) | rs01 = σ(z) | Reward valuation, positive affect and urges; digital allocation/night use; externalizing and mania; high-, low- and mid-range reward features |
| self_reg | 0 | 1.0 | [−2.5, 2.5] (z) | sr01 = σ(z) | Regulatory capacity; strain, sleep and digital allocation; behavioural plasticity/intervention response; PFC-scaled and zone features |
| social_need | 0 | 1.0 | [−2.5, 2.5] (z) | sn01 = σ(z) | Loneliness and social-discrepancy sensitivity; social, gaming and in-person allocation; protective composite; raw-z loneliness/EMA social gain |
| resilience | 0 | 1.0 | [−2.5, 2.5] (z) | r01 = σ(z) | Broad buffering of strain and psychiatric vulnerability; recovery, sleep and HRV; learning/plasticity; adherence and intervention impact |
| **PSYCHIATRIC PREDISPOSITIONS** | | | | | |
| psychosis_prone | 0 | 1.0 | [−2.5, 2.5] (z) | pp01 = σ(1.2 × (z − 1.5)) | Psychosis initialization, vulnerability and attractor dynamics; paranoia, hallucination and insight; digital and wearable psychosis effects |
| cog_disturbance | 0 | 1.0 | [−2.5, 2.5] (z) | cd01 = σ(1.2 × (z − 1.5)) | Psychosis plateau/susceptibility; concentration and cognitive-load channels; BLIP/hallucination amplification; digital temporal instability |
| anhedonia_prone | 0 | 1.0 | [−2.5, 2.5] (z) | ap01 = σ(1.2 × z) | Depressive vulnerability and attractor dynamics; anhedonia; reward/social withdrawal; sleep, psychomotor and physical-activity effects |
| hopelessness | 0 | 1.0 | [−2.5, 2.5] (z) | hp01 = σ(1.2 × z) | Depressive vulnerability and persistence; hopelessness/hope; suicidal-load and suicidality channels; strain feedback |
| bipolarity_prone | 0 | 1.0 | [−2.5, 2.5] (z) | bp01 = σ(1.2 × z) | Mania and mixed-state attractors; positive-affect and cognitive initialization; cannabis-sensitive mania; wearable psychomotor patterning |
| **DIGITAL HABIT COORDINATES** | | | | | |
| habit_social | 0 | 1.0 | [−2.5, 2.5] (z) | habit_social01 = σ(z) | Initial social-platform habit state and social-use mean; adaptive habit dynamics; platform allocation and domain-specific drive |
| habit_game | 0 | 1.0 | [−2.5, 2.5] (z) | habit_game01 = σ(z) | Initial gaming habit state and gaming-use mean; adaptive habit dynamics; platform allocation and domain-specific drive |
| habit_chat | 0 | 1.0 | [−2.5, 2.5] (z) | habit_chat01 = σ(z) | Initial chat habit state and chat-use mean; adaptive habit dynamics; chatbot/messaging allocation |
| **PHYSIOLOGICAL ANCHORS (NOT z-SCALED TRAITS)** | | | | | |
| sleep_need | 8.5 h | 1.4 h | [6.0, 10.0] h | sleep_need01 = (h − 6)/4 | Person-specific sleep-duration baseline; curfew-sleep bonus; circadian phase brake; wearable sleep generation and indirect sleep-debt dynamics Evidence: Direct—age-specific duration recommendations [S01, S02]; Expert-coded—8.5 h mean, 1.4 h SD, bounds and transform. |
| steps_anchor_z | — | — | [−3, 3] (z); optional | steps_anchor = exp(μlog(age) + zσlog(age)) | Age-referenced daily-step anchor and wearable activity level; absent when not supplied, in which case the legacy parameter-library anchor is used |

**Supplementary Table S1.1b. Nonlinear and demographic-derived transformations of trait coordinates.** The listed subject-fixed features are computed from bounded coordinates, age and sex at birth, using thresholds defined in P.trait.zone.* and demographic parameters in P.demog.* of the CSDS Parameter Library. The hinge features are exactly zero outside their active zones; the inverted-U features peak at the adaptive middle. CSDS modules also construct context- and state-dependent quantities such as skill-augmented self-regulation, liability-specific excess terms and effective-resilience composites. Literature supports the broad low/high phenotype rationale while the exact hinges, quadratic inverted-U functions, age curves, binary sex gate and composite weights are expert-informed CSDS encodings based on clinical experience and calibration rather than published clinical cut-offs.

| **Derived feature** | **Source** | **Type** | **Definition** | **Active range** | **Representative uses and evidence** |
| --- | --- | --- | --- | --- | --- |
| **SELF-REGULATION ZONES** | | | | | |
| sr01_excess | self_reg | Upper hinge | max(sr01 − 0.75, 0) / 0.25 | sr01 > 0.75 | OCD/checking and repetitive-routine channels, usually gated by current OCD load; sleep regularity and autonomic/activity effects |
| sr01_rigid | self_reg | Upper hinge, capped | min(max(sr01 − 0.65, 0) / 0.35, 1) | sr01 > 0.65 | OCD initialization, plateau and sensitivity; checking/rigidity symptoms; reduced digital and behavioural variability |
| sr01_impul | self_reg | Lower hinge, capped | min(max(0.35 − sr01, 0) / 0.50, 1) | sr01 < 0.35 | Externalizing and impulsive-checking susceptibility; negative-affect/urge amplification; late-night digital disinhibition |
| **REWARD-SENSITIVITY ZONES** | | | | | |
| rs01_excess | reward_sens | Upper hinge | max(rs01 − 0.70, 0) / 0.30 | rs01 > 0.70 | BPD/social-evaluation reactivity and volatility; digital pressure/social bias; mania forcing and reward-sensitivity × cannabis interaction |
| rs01_hyper | reward_sens | Upper hinge | max(rs01 − 0.60, 0) / 0.40 | rs01 > 0.60 | Externalizing plateau, cue-reactive forcing and susceptibility in updatePsychiatricState; no direct digital-generator consumer |
| rs01_hypo | reward_sens | Lower hinge, capped | min(max(0.35 − rs01, 0) / 0.35, 1) | rs01 < 0.35 | Eating-disorder vulnerability; reduced digital pressure; autism-like social damping and gaming preference |
| **INVERTED-U FEATURES** | | | | | |
| sr01_eff_u | self_reg | Inverted-U | 4 × sr01 × (1 − sr01) | peak at sr01 = 0.5 | Residual self-regulatory contribution to weekday school/structured-schedule attendance in the digital generator |
| rs01_social | reward_sens | Inverted-U | 4 × rs01 × (1 − rs01) | peak at rs01 = 0.5 | Social-versus-gaming propensity, daily social allocation and in-person social claim |
| **DEVELOPMENTAL AND DEMOGRAPHIC FEATURES** | | | | | |
| pfc_maturity | age | Sigmoid | σ((age − 22.0) / 3.0) | monotone in age | Developmental maturity and psychosis susceptibility through the complementary pfc_immaturity term |
| pfc01 | age | Floored sigmoid | 0.20 + 0.80 × pfc_maturity | [0.20, 1.0] | Developmental regulatory-capacity scale; digital control and cognitive-disturbance × PFC-immaturity pathways |
| sr01_pfc | self_reg × age | Product | sr01 × pfc01 | — | Available regulatory capacity across digital, wearable and psychiatric-state modules |
| sr01_eff_u_pfc | self_reg × age | Inverted-U | 4 × sr01_pfc × (1 − sr01_pfc) | peak at sr01_pfc = 0.5 | Buffers negative-affect and cognitive-load EMA drivers |
| phase_age_peak | age | Inverted-U | 4 × pfc_maturity × (1 − pfc_maturity) | peak at age 22 | Developmental contribution to circadian phase-delay pressure in wearable sleep generation |
| adolesc_peak | age | Gaussian | exp(−(age − 16)² / (2 × 2.5²)) | peak at age 16 | Developmental amplification of sleep/psychiatric volatility and ED, appearance-comparison, externalizing and social-evaluation pathways |
| is_female | sex at birth | Binary | 1 if sex_at_birth begins with ‘F’; otherwise 0 | {0, 1} | Eating-disorder load and symptoms, appearance comparison, digital bias, autonomic/activity effects and small suicidality modulation |
| **COMPOSITE** | | | | | |
| bpd_instability_base | reward_sens × resilience × neuroticism | Product composite | rs01_excess × (1 − r01) × (0.50 + 0.50 × nn01) | — | Static fallback for disproportionate emotional reactivity. updatePsychiatricState recomputes the full composite with R_eff replacing r01 and exposes the dynamic BPD amplitude to generators |

**Supplementary Table S9.1. Proportional corridor factors and soft-target weights.** *f_j_* is the prespecified proportional corridor factor used to construct the proportional interval around each cluster target centre (Supplementary Methods 9.1). For non-correlation targets the final corridor is the bounded outer envelope of this interval and the empirical interquartile range of participant-level values, so the realised corridor may be wider than *f_j_* alone implies. Entries marked n/a do not use a proportional band: within-subject autocorrelation targets are constructed as between-subject prediction intervals on Fisher-z-transformed participant-level correlations, and structural safety priors are fixed corridors not estimated from GLOBEM. Weights apply to soft targets only.

| **CSDS target path** | **Source metric** | **Role** | ***f_j_*** | **Weight** |
| --- | --- | --- | --- | --- |
| **HARD TARGETS (conjunctive)** | | | | |
| digital.platform_use.mean | csds_platform_use_mean | hard | 0.15 | — |
| digital.media_night_h.mean | csds_media_night_h_mean | hard | 0.18 | — |
| digital.media_night_share.mean | csds_media_night_share_mean | hard | 0.30 | — |
| wearable.sleep_hours.mean | csds_sleep_hours_mean | hard | 0.20 | — |
| wearable.steps.mean | csds_steps_mean | hard | 0.20 | — |
| ema.stress.mean | csds_stress_mean | hard | 0.15 | — |
| ema.pa_proxy.mean | csds_pa_hmm_mean | hard | 0.40 | — |
| ema.depress_proxy.mean | csds_depress_load_proxy_mean | hard | 0.20 | — |
| **SOFT — WITHIN-SUBJECT DISPERSION** | | | | |
| digital.platform_use.sd | csds_platform_use_sd | soft | 0.25 | 0.45 |
| digital.media_night_h.sd | csds_media_night_h_sd | soft | 0.30 | 0.45 |
| wearable.sleep_hours.sd | csds_sleep_hours_sd | soft | 0.25 | 0.30 |
| wearable.steps.sd | csds_steps_sd | soft | 0.25 | 0.45 |
| ema.stress.sd | csds_stress_sd | soft | 0.20 | 0.55 |
| ema.pa_proxy.sd | csds_pa_hmm_sd | soft | 0.20 | 0.45 |
| ema.depress_proxy.sd | csds_depress_load_proxy_sd | soft | 0.25 | 0.55 |
| **SOFT — THRESHOLD / EXTREME** | | | | |
| ema.stress.pHigh3 | csds_stress_pHigh8 | soft | 0.25 | 0.45 |
| ema.pa_proxy.pLow1 | csds_pa_hmm_pLow1 | soft | 0.25 | 0.40 |
| ema.depress_proxy.pLow_01 | csds_depress_load_proxy_pLow_01 | soft | 0.25 | 0.65 |
| ema.depress_proxy.pHigh_03 | csds_depress_load_proxy_pHigh_03 | soft | 0.25 | 0.55 |
| **SOFT — COMPOSITE LEVEL** | | | | |
| ema.rumin.mean | csds_rumin_proxy_mean | soft | 0.25 | 0.45 |
| **SOFT — WITHIN-SUBJECT DYNAMICS (Fisher-z path; no proportional band)** | | | | |
| digital.platform_use.ac1 | csds_platform_use_ac1 | soft | n/a | 0.35 |
| digital.media_night_h.ac1 | csds_media_night_h_ac1 | soft | n/a | 0.35 |
| **SOFT — BETWEEN-SUBJECT DISPERSION** | | | | |
| between.digital.platform_use.mean_sd | between_csds_platform_use_mean_sd | soft | 0.25 | 0.65 |
| between.digital.media_night_h.mean_sd | between_csds_media_night_h_mean_sd | soft | 0.25 | 0.65 |
| between.wearable.sleep_hours.mean_sd | between_csds_sleep_hours_mean_sd | soft | 0.25 | 0.65 |
| between.ema.stress.mean_sd | between_csds_stress_mean_sd | soft | 0.25 | 0.70 |
| between.ema.pa_proxy.mean_sd | between_csds_pa_hmm_mean_sd | soft | 0.25 | 0.70 |
| between.ema.depress_proxy.mean_sd | between_csds_depress_load_proxy_mean_sd | soft | 0.25 | 0.75 |
| **SOFT — BETWEEN-SUBJECT SHAPE (Q10 / Q90)** | | | | |
| between.ema.stress.mean_q10 | between_csds_stress_mean_q10 | soft | 0.20 | 0.55 |
| between.ema.stress.mean_q90 | between_csds_stress_mean_q90 | soft | 0.20 | 0.55 |
| between.ema.pa_proxy.mean_q10 | between_csds_pa_hmm_mean_q10 | soft | 0.20 | 0.80 |
| between.ema.pa_proxy.mean_q90 | between_csds_pa_hmm_mean_q90 | soft | 0.20 | 0.80 |
| between.ema.depress_proxy.mean_q10 | between_csds_depress_load_proxy_mean_q10 | soft | 0.20 | 1.05 |
| between.ema.depress_proxy.mean_q90 | between_csds_depress_load_proxy_mean_q90 | soft | 0.20 | 0.95 |
| **SOFT — STRUCTURAL SAFETY PRIORS (fixed corridors; no proportional band)** | | | | |
| ema.mania_load.mean | safety_mania_load_mean | soft | n/a | 0.85 |
| ema.ocd_load.mean | safety_ocd_load_mean | soft | n/a | 0.85 |
| ema.psychosis_load.mean | safety_psychosis_load_mean | soft | n/a | 0.85 |
| ema.suicidal_load.mean | safety_suicidal_load_mean | soft | n/a | 0.85 (CL1+3), 0.45 (CL4), 0.30 (CL2) |

**Supplementary Table S10.3. CSDS mechanistic parameters associated with GLOBEM cluster separation.** The table lists the 30 parameters with the largest between-cluster effects across the cluster-specific posterior particle ensembles, ranked by one-way ANOVA $\eta^{2}$. For each parameter, the maximum absolute $z$-shift indicates the largest standardized displacement of a cluster mean from the global particle mean. The dominant-cluster Cohen’s $d$compares the cluster showing the strongest deviation with all remaining clusters pooled; positive and negative values indicate higher and lower parameter values, respectively, in the dominant cluster. The accompanying descriptions summarise each parameter’s role in the CSDS.

| **Rank** | **CSDS parameter** | **Mechanistic domain** | **Role in the CSDS** | **η²** | **Maximum absolute z shift** | **Dominant-cluster Cohen's d vs rest** | **Dominant cluster (direction)** |
| --- | --- | --- | --- | --- | --- | --- | --- |
| 1 | wearable.steps_log_sd_max | Physical activity | Upper bound on the effective log-scale standard deviation of daily steps; caps day-to-day activity variability after state-dependent modulation. | 0.976 | 1.731 | 13.993 | C2 (higher) |
| 2 | wearable.steps_mod.sd_boost_cap | Physical activity | Caps state-dependent amplification of step variability, limiting the combined volatility boosts generated by active behavioural and psychiatric states. | 0.966 | 1.723 | 11.971 | C2 (higher) |
| 3 | wearable.steps_log_sd_min | Physical activity | Lower bound on the effective log-scale standard deviation of daily steps; permits stable, low-variability activity profiles while preventing variance collapse. | 0.962 | 1.716 | 10.853 | C2 (higher) |
| 4 | traits.anhedonia_prone | Trait vulnerability | Baseline susceptibility to anhedonic and depressive processes that enters downstream affective, behavioural, and recovery mechanisms. | 0.850 | 1.272 | -2.506 | C3 (lower) |
| 5 | traits.neuroticism | Trait vulnerability | Baseline emotional-reactivity trait contributing to negative bias and vulnerability across affective and strain dynamics. | 0.748 | 0.965 | 1.530 | C2 (higher) |
| 6 | ema.core.emotion_gain_base | Affective response / EMA | Baseline gain translating latent affective dynamics into EMA affect utilities; the effective gain is further modulated by resilience. | 0.742 | 1.068 | 1.810 | C3 (higher) |
| 7 | ema.pa.anhed_low_resource_amp | Affective response / EMA | Additional positive-affect debit when anhedonic or depressive liability coincides with low sleep, activity, or social resources. | 0.739 | 1.046 | -1.781 | C1 (lower) |
| 8 | traits.self_reg | Trait regulation | Baseline regulatory capacity influencing control, recovery, behavioural regulation, and vulnerability pathways. | 0.703 | 1.333 | 2.791 | C3 (higher) |
| 9 | ema.blip.hall_burst_amp | Psychosis / EMA | Amplitude of transient hallucination bursts generated from paranoia-burst activity and cognitive-disturbance gating in the EMA psychosis observation model. | 0.681 | 0.998 | 1.619 | C4 (higher) |
| 10 | oscillator.cross.dys_mult | Cross-domain coupling | Strength of the cross-domain gate by which digital dysregulation increases physiological vulnerability in the oscillator system. | 0.657 | 1.253 | -2.486 | C1 (lower) |
| 11 | traits.hopelessness | Trait vulnerability | Baseline hopelessness trait contributing to depressive persistence, social-disconnection colouring, and recovery-related pathways. | 0.636 | 1.393 | 3.034 | C2 (higher) |
| 12 | ema.trait.sleep_nominal_h | Sleep / EMA | Nominal sleep-duration reference used to calculate sleep deficit for EMA-related trait and observation effects. | 0.632 | 1.053 | 1.771 | C3 (higher) |
| 13 | traits.habit_social | Digital habit | Baseline social-platform habit trait influencing digital-channel preference and persistence (social media). | 0.627 | 1.052 | 1.798 | C1 (higher) |
| 14 | digitalDay.circadian_shaping_for_digital_domains_re.a01_psy | Digital dynamics | Weight of psychosis load in the A01 activation term used for circadian and state-dependent shaping of digital domains. | 0.619 | 1.142 | -2.029 | C3 (lower) |
| 15 | digitalDay.stable_satiation_fatigue.sat_input_c1_2 | Digital dynamics | Weight of the `iv_active` state in the digital satiation input, thereby modifying stable satiation–fatigue dynamics. | 0.606 | 1.271 | 2.567 | C1 (higher) |
| 16 | ema.psy.parano_psy_gate_coeff | Psychosis / EMA | Gain by which the psychosis gate and psychosis load drive paranoia utility in the EMA psychosis observation model. | 0.595 | 1.041 | -1.728 | C4 (lower) |
| 17 | ema.psy.insight_regime_coeff | Psychosis / EMA | Scales the regime-dependent psychosis-load term in insight utility, modulating the expression of insight across psychotic states. | 0.587 | 1.089 | 1.904 | C1 (higher) |
| 18 | digitalDay.daily_propensity_components.psy_checking_esc_c2 | Digital dynamics | Coefficient within the psychosis-related checking/escalation component of daily digital propensity. | 0.586 | 1.254 | -2.372 | C2 (lower) |
| 19 | ema.core.lonely_ela_coeff | Social-affective / EMA | Weight by which early-life adversity amplifies social-unsafety and loneliness under a current social deficit. | 0.585 | 1.297 | 2.614 | C3 (higher) |
| 20 | psychiatric.bpd.ext_dep_thresh | BPD dynamics | Externalising-load threshold above which a next-day depressive guilt/shame rebound is activated. | 0.582 | 1.166 | 2.066 | C2 (higher) |
| 21 | wearable.hrv_smoothing.alpha_min | Wearable physiology | Minimum value of the HRV smoothing coefficient, bounding the temporal smoothing applied during wearable-physiology rendering. | 0.577 | 1.154 | -2.051 | C4 (lower) |
| 22 | sleep.late_bed_base | Sleep dynamics | Baseline contribution to late-bedtime tendency before strain- and reward-sensitivity-dependent modifiers. | 0.575 | 1.283 | -2.623 | C1 (lower) |
| 23 | digitalDay.digital_oscillator_state_update.expr_c1_12 | Digital dynamics | Weight of psychosis load in the vulnerability term governing digital-oscillator dysregulation. | 0.573 | 1.059 | 1.761 | C3 (higher) |
| 24 | ema.sep.rum_cc_w | Rumination / EMA | Weight of low cognitive control in the rumination and perseveration driver. | 0.572 | 1.074 | -1.861 | C1 (lower) |
| 25 | ema.core.lonely_bpd_coeff | Social-affective / EMA | Attachment-sensitive social-hunger contribution to loneliness under a current social deficit. | 0.567 | 1.311 | 2.611 | C2 (higher) |
| 26 | strain.psych_strain.psy_ins_mix_span | Psychosis–strain coupling | Span above the lower floor for the psychosis–insight mixture contribution to psychological strain. | 0.567 | 0.951 | -1.498 | C2 (lower) |
| 27 | ema.prior.conc_base | Cognition / EMA | Baseline concentration input used to construct the EMA concentration channel before state-dependent modifiers. | 0.565 | 1.060 | 1.821 | C1 (higher) |
| 28 | ema.driver.pa_balance_collapse_negative_thresh | Positive affect / EMA | Negative-burden threshold required to activate the extreme positive-affect collapse term under severe low-resource states. | 0.564 | 1.087 | -1.834 | C2 (lower) |
| 29 | digitalDay.circadian_shaping_for_digital_domains_re.aut_social_damp_c1 | Digital dynamics | Weight of reward-sensitivity deficit in the `aut_social_damp` term that attenuates social-channel allocation during circadian shaping. | 0.560 | 1.224 | 2.367 | C1 (higher) |
| 30 | digitalDay.digital_oscillator_state_update.slowing_c2 | Digital dynamics | Weight of proximity to the current target or attractor (`near`) in slowing the digital-oscillator state update. | 0.560 | 1.242 | 2.328 | C2 (higher) |

**Supplementary Table S11.5 Interpretation of cluster-specific state taxonomies.** Individual subject states were aligned to an expandable cluster-specific registry when their fingerprints correlated with a registry state at Spearman rho > 0.50; unmatched states were added to the registry. Level and difference profiles are equal-subject-weighted observed state-conditional means of HMM features standardized using each participant's synthetic virtual-twin (VT) reference distribution. Accordingly, higher/lower and increasing/decreasing denote deviations from the participant-specific VT reference, not absolute levels or changes, and do not permit direct comparison of absolute burden between clusters. Weak depression-proxy contrasts within a cluster therefore do not imply low absolute depressive burden. Intensity terms summarise relative fingerprint magnitudes on this standardised scale. Difference features represent changes between consecutive available observations; for EMA variables, these observations were not necessarily on consecutive days. Descriptions are interpretive and non-diagnostic. State numbers are cluster-specific and do not establish cross-cluster homology.

| **Cluster** | **State** | **Description** | **Defining observed level profile** | **Observed dynamic signature** | **Persistence and dominant transition** |
| --- | --- | --- | --- | --- | --- |
| **CL1** | **S1** | Relatively high and rising night-time digital engagement with mildly higher and increasing stress | strongly higher night-time media use; markedly higher platform use; moderately lower depression proxy; mildly higher stress. | increasing night-time media use and platform use; mildly increasing stress. | moderate persistence (self-transition 0.61); most frequent non-self transition to S3 (0.16). |
| **CL1** | **S2** | Relatively low digital engagement with mildly higher but decreasing stress | markedly lower night-time media use; moderately lower platform use; mildly higher stress. | decreasing stress. | high persistence (self-transition 0.67); most frequent non-self transition to S1 (0.11). |
| **CL1** | **S3** | Relatively low and falling digital engagement with lower depression proxy and decreasing stress | markedly lower night-time media use; moderately lower platform use; moderately lower depression proxy. | decreasing night-time media use and platform use; mildly decreasing stress. | moderate persistence (self-transition 0.62); most frequent non-self transition to S1 (0.13). |
| **CL1** | **S4** | Relatively high and increasing night-time digital engagement with mildly higher stress | markedly higher night-time media use; moderately higher platform use; mildly higher stress; moderately lower depression proxy. | increasing night-time media use; mildly increasing platform use. | moderate persistence (self-transition 0.65); most frequent non-self transition to S3 (0.15). |
| **CL1** | **S5** | Very low digital engagement with lower positive affect but higher stress and depression proxy | strongly lower night-time media use; markedly lower platform use; moderately lower positive affect; moderately higher stress and depression proxy. | increasing stress and depression proxy. | moderate persistence (self-transition 0.62); most frequent non-self transition to S4 (0.15). |
| **CL1** | **S6** | Markedly lower depression proxy with relatively low digital engagement and stress | pronounced lower depression proxy; moderately lower platform use and stress; mildly lower night-time media use. | decreasing depression proxy; mildly decreasing night-time media use. | very high persistence (self-transition 0.74); most frequent non-self transition to S4 (0.07). |
| **CL1** | **S7** | Relatively high digital engagement with higher positive affect and stress but lower depression proxy | markedly higher platform and night-time media use; moderately higher positive affect and stress; moderately lower depression proxy; mildly lower step counts. | increasing platform and night-time media use; mildly decreasing depression proxy. | high persistence (self-transition 0.66); most frequent non-self transition to S3 (0.11). |
| **CL1** | **S8** | Relatively low digital engagement with higher and rising depression proxy | markedly lower platform use; moderately lower night-time media use; moderately higher depression proxy; mildly lower stress. | decreasing night-time media use and platform use; increasing depression proxy. | high persistence (self-transition 0.67); most frequent non-self transition to S4 (0.11). |
| **CL2** | **S1** | Relatively high and rising total and night-time digital engagement | strongly higher night-time media use; markedly higher platform use; mildly shorter sleep; mildly higher positive affect and depression proxy. | increasing night-time media use and platform use; mildly increasing depression proxy and decreasing stress. | moderate persistence (self-transition 0.62); most frequent non-self transition to S3 (0.15). |
| **CL2** | **S2** | Moderately high digital engagement with increasing night-time media use | markedly higher night-time media use; moderately higher platform use; mildly lower depression proxy. | increasing night-time media use; mildly increasing platform use and decreasing positive affect. | high persistence (self-transition 0.66); most frequent non-self transition to S3 (0.09). |
| **CL2** | **S3** | Relatively low and falling digital engagement with moderately higher depression proxy | markedly lower platform and night-time media use; moderately higher depression proxy; mildly lower positive affect and mildly higher stress. | decreasing night-time media use and platform use; mildly increasing positive affect. | moderate persistence (self-transition 0.63); most frequent non-self transition to S1 (0.11). |
| **CL2** | **S4** | Relatively low digital engagement with mildly higher stress and depression proxy | markedly lower night-time media and platform use; mildly higher stress, positive affect, and depression proxy. | mildly decreasing night-time media use; mildly increasing stress and positive affect. | moderate persistence (self-transition 0.65); most frequent non-self transition to S1 (0.11). |
| **CL2** | **S5** | Very low and falling digital engagement with mildly shorter sleep and rising depression proxy | markedly lower platform and night-time media use; mildly shorter sleep; mildly higher step counts; mildly lower stress and higher depression proxy. | decreasing platform and night-time media use; increasing depression proxy; mildly increasing stress. | moderate persistence (self-transition 0.65); most frequent non-self transition to S2 (0.08). |
| **CL2** | **S6** | Relatively high digital engagement with higher stress and lower depression proxy | markedly higher night-time media use; moderately higher platform use and stress; mildly longer sleep; mildly lower positive affect and depression proxy. | increasing platform and night-time media use and stress; decreasing depression proxy and positive affect. | high persistence (self-transition 0.65); most frequent non-self transition to S3 (0.09). |
| **CL2** | **S7** | Low-activity state with relatively low platform use but rising digital engagement | strongly lower step counts; moderately lower platform use; mildly higher night-time media use; mildly lower stress and depression proxy. | increasing platform and night-time media use; mildly increasing stress. | moderate persistence (self-transition 0.61); most frequent non-self transition to S3 (0.14). |
| **CL2** | **S8** | Relatively low and falling digital engagement with lower depression proxy, higher positive affect, and longer sleep | markedly lower platform and night-time media use and depression proxy; moderately longer sleep and higher positive affect; mildly higher step counts and lower stress. | decreasing platform and night-time media use; mildly increasing stress and positive affect. | moderate persistence (self-transition 0.61); most frequent non-self transition to S1 (0.14). |
| **CL2** | **S9** | Higher positive affect with very low depression proxy and falling digital engagement | strongly lower depression proxy; markedly lower night-time media use; markedly higher positive affect; moderately lower platform use, sleep duration, and step counts. | increasing depression proxy; decreasing platform and night-time media use and positive affect. | lower persistence (self-transition 0.59); most frequent non-self transition to S6 (0.21). |
| **CL2** | **S10** | Short-sleep state with relatively low depression proxy and stress | markedly lower depression proxy; moderately lower night-time media use, sleep duration, and stress; platform use near the VT reference. | decreasing positive affect and stress; mildly increasing depression proxy. | very high persistence (self-transition 0.71); most frequent non-self transition to S1 (0.16). |
| **CL3** | **S1** | Relatively low night-time digital use with higher and rising depression proxy but lower stress | markedly lower night-time media use; moderately higher depression proxy; moderately lower stress; mildly higher positive affect and lower platform use. | increasing depression proxy; mildly decreasing positive affect. | moderate persistence (self-transition 0.63); most frequent non-self transition to S2 (0.11). |
| **CL3** | **S2** | Higher and rising platform engagement with lower depression proxy | markedly higher platform use; mildly higher night-time media use and stress; moderately lower depression proxy; mildly lower positive affect. | increasing platform and night-time media use; mildly increasing depression proxy. | very high persistence (self-transition 0.70); most frequent non-self transition to S5 (0.13). |
| **CL3** | **S3** | Very low digital engagement with lower positive affect and rising stress | strongly lower night-time media use; markedly lower platform use; mildly longer sleep; mildly lower positive affect and depression proxy. | increasing stress; mildly increasing depression proxy. | moderate persistence (self-transition 0.61); most frequent non-self transition to S2 (0.20). |
| **CL3** | **S4** | Low night-time digital engagement with higher and strongly increasing stress | markedly lower night-time media use; moderately higher stress; mildly higher step counts and positive affect; mildly lower platform use. | strongly increasing stress; increasing depression proxy and mildly increasing positive affect. | lower persistence (self-transition 0.60); most frequent non-self transition to S8 (0.14). |
| **CL3** | **S5** | Relatively low and falling digital engagement with lower depression proxy | markedly lower platform and night-time media use; moderately lower depression proxy; mildly lower stress. | decreasing platform and night-time media use. | lower persistence (self-transition 0.59); most frequent non-self transition to S2 (0.14). |
| **CL3** | **S6** | Relatively high and rising digital engagement with lower and decreasing depression proxy | markedly higher platform and night-time media use; moderately lower depression proxy; mildly longer sleep. | increasing platform and night-time media use; decreasing depression proxy. | moderate persistence (self-transition 0.63); most frequent non-self transition to S5 (0.14). |
| **CL3** | **S7** | Low digital engagement with higher activity, shorter sleep, and lower depression proxy | markedly lower platform and night-time media use and depression proxy; moderately higher step counts; mildly shorter sleep. | mildly decreasing platform and night-time media use. | moderate persistence (self-transition 0.61); most frequent non-self transition to S2 (0.12). |
| **CL3** | **S8** | Very high and rising platform engagement with shorter sleep | strongly higher platform use; markedly higher night-time media use; moderately shorter sleep; mildly higher step counts and lower depression proxy. | increasing platform and night-time media use; mildly increasing positive affect and stress. | moderate persistence (self-transition 0.63); most frequent non-self transition to S5 (0.16). |
| **CL3** | **S9** | Pronounced digital engagement with lower stress, positive affect, and depression proxy | pronounced higher platform and night-time media use; markedly lower stress and positive affect; moderately lower depression proxy; moderately longer sleep. | increasing platform and night-time media use; decreasing depression proxy and mildly decreasing positive affect. | high persistence (self-transition 0.68); most frequent non-self transition to S5 (0.13). |
| **CL3** | **S10** | Very low and falling depression proxy with longer sleep and declining digital use | strongly lower depression proxy; moderately lower night-time media use; mildly longer sleep and higher stress; mildly lower platform use. | decreasing platform and night-time media use and depression proxy. | moderate persistence (self-transition 0.60); most frequent non-self transition to S2 (0.15). |
| **CL4** | **S1** | Relatively high and rising digital engagement with higher activity and stress | markedly higher platform and night-time media use; moderately higher step counts and stress; mildly higher positive affect. | increasing platform and night-time media use and stress; decreasing depression proxy. | moderate persistence (self-transition 0.64); most frequent non-self transition to S3 (0.17). |
| **CL4** | **S2** | Relatively high and rising digital engagement | markedly higher platform and night-time media use; other level features near the VT reference. | increasing platform and night-time media use. | high persistence (self-transition 0.67); most frequent non-self transition to S3 (0.15). |
| **CL4** | **S3** | Relatively low and falling digital engagement with mildly longer sleep | markedly lower platform and night-time media use; mildly longer sleep; mildly lower stress and positive affect. | decreasing platform and night-time media use; mildly decreasing stress and depression proxy. | moderate persistence (self-transition 0.60); most frequent non-self transition to S2 (0.10). |
| **CL4** | **S4** | Active low-digital state with higher and strongly increasing stress | markedly lower night-time media use; moderately lower platform use; moderately higher step counts and stress. | strongly increasing stress; mildly increasing positive affect. | high persistence (self-transition 0.66); most frequent non-self transition to S2 (0.16). |
| **CL4** | **S5** | Low night-time media use with mildly higher but decreasing stress | markedly lower night-time media use; mildly lower platform use; mildly higher stress. | decreasing stress; mildly increasing depression proxy. | moderate persistence (self-transition 0.62); most frequent non-self transition to S6 (0.07). |
| **CL4** | **S6** | Relatively low and falling digital engagement with higher depression proxy and lower positive affect | markedly lower night-time media and platform use; moderately higher depression proxy and step counts; mildly lower positive affect. | decreasing platform and night-time media use and positive affect; mildly increasing depression proxy. | high persistence (self-transition 0.66); most frequent non-self transition to S2 (0.09). |
| **CL4** | **S7** | Relatively low depression proxy and stress with low and falling digital engagement | markedly lower depression proxy, night-time media use, platform use, and stress; mildly higher positive affect. | decreasing depression proxy, platform use, and night-time media use; mildly decreasing stress. | moderate persistence (self-transition 0.62); most frequent non-self transition to S1 (0.11). |
| **CL4** | **S8** | Relatively high and rising digital engagement with shorter sleep and lower positive affect | markedly higher platform and night-time media use; mildly shorter sleep; mildly lower positive affect and higher depression proxy. | increasing platform and night-time media use; mildly increasing stress and depression proxy and decreasing positive affect. | high persistence (self-transition 0.66); most frequent non-self transition to S3 (0.16). |
| **CL4** | **S9** | Longer-sleep state with higher depression proxy and rising digital engagement | moderately longer sleep and higher depression proxy; moderately lower positive affect; mildly higher platform use and lower night-time media use. | increasing platform and night-time media use; decreasing stress. | high persistence (self-transition 0.69); most frequent non-self transition to S8 (0.17). |
| **CL4** | **S10** | Short-sleep, low night-time media state with higher and rising depression proxy and lower positive affect | markedly lower night-time media use and sleep duration; markedly higher depression proxy; moderately lower positive affect; mildly lower platform use. | strongly increasing depression proxy; decreasing night-time media use; mildly decreasing platform use and positive affect. | very high persistence (self-transition 0.70); most frequent non-self transition to S2 (0.09). |
| **CL4** | **S11** | Low-activity, low-digital state with higher positive affect and strongly rising depression proxy | markedly lower night-time media use and step counts; moderately lower platform use; moderately higher positive affect; depression proxy near the VT reference. | pronounced increase in depression proxy; decreasing positive affect; mildly increasing sleep duration. | very high persistence (self-transition 0.71); most frequent non-self transition to S2 (0.17). |

**Supplementary Table S11.6. State-specific occupancy and fingerprint concordance within each GLOBEM cluster. Occupancy** values are equal-subject-weighted means. Fingerprint values are median subject-level correlations. 95%-Cis are computed using 5000 subject-bootstrap iterations; valid n/N is shown after the semicolon. State-specific correlations may be based on fewer subjects when the corresponding observed state was not sufficiently represented. States are aligned within clusters only.

| **State** | **Observed occupancy** | **Synthetic occupancy** | **Overall fingerprint r** | **Level-feature r** | **Difference-feature r** |
| --- | --- | --- | --- | --- | --- |
| **Cluster 1** | | | | | |
| S1 | 0.205 (0.166–0.246) | 0.208 (0.184–0.232) | 0.842 (0.809–0.886); 46/50 | 0.894 (0.852–0.923); 46/50 | 0.702 (0.611–0.783); 45/50 |
| S2 | 0.113 (0.081–0.148) | 0.174 (0.139–0.209) | 0.849 (0.788–0.867); 31/50 | 0.917 (0.849–0.935); 31/50 | -0.014 (-0.173–0.142); 31/50 |
| S3 | 0.217 (0.172–0.262) | 0.167 (0.136–0.196) | 0.890 (0.821–0.918); 36/50 | 0.925 (0.875–0.941); 36/50 | 0.824 (0.694–0.848); 36/50 |
| S4 | 0.212 (0.170–0.255) | 0.157 (0.132–0.181) | 0.854 (0.790–0.890); 39/50 | 0.937 (0.876–0.953); 39/50 | 0.407 (0.080–0.455); 39/50 |
| S5 | 0.048 (0.025–0.074) | 0.066 (0.040–0.096) | 0.806 (0.776–0.904); 17/50 | 0.854 (0.768–0.937); 17/50 | 0.162 (0.078–0.551); 17/50 |
| S6 | 0.028 (0.012–0.046) | 0.045 (0.025–0.068) | 0.765 (0.504–0.899); 12/50 | 0.833 (0.603–0.909); 12/50 | 0.360 (-0.192–0.638); 11/50 |
| S7 | 0.080 (0.042–0.123) | 0.073 (0.039–0.109) | 0.869 (0.806–0.907); 15/50 | 0.919 (0.836–0.956); 15/50 | 0.409 (0.137–0.614); 15/50 |
| S8 | 0.096 (0.062–0.133) | 0.110 (0.078–0.143) | 0.833 (0.714–0.878); 23/50 | 0.898 (0.866–0.944); 23/50 | 0.379 (0.286–0.616); 21/50 |
| **Cluster 2** | | | | | |
| S1 | 0.209 (0.173–0.248) | 0.185 (0.161–0.209) | 0.817 (0.762–0.891); 45/50 | 0.903 (0.836–0.941); 45/50 | 0.681 (0.613–0.765); 45/50 |
| S2 | 0.153 (0.112–0.195) | 0.132 (0.100–0.163) | 0.842 (0.738–0.878); 30/50 | 0.915 (0.808–0.947); 30/50 | 0.373 (0.217–0.497); 30/50 |
| S3 | 0.175 (0.131–0.218) | 0.159 (0.123–0.195) | 0.877 (0.818–0.903); 32/50 | 0.899 (0.851–0.941); 32/50 | 0.695 (0.591–0.804); 32/50 |
| S4 | 0.134 (0.097–0.176) | 0.188 (0.161–0.215) | 0.797 (0.729–0.896); 39/50 | 0.840 (0.790–0.917); 39/50 | 0.279 (0.068–0.502); 39/50 |
| S5 | 0.084 (0.049–0.126) | 0.092 (0.061–0.125) | 0.814 (0.667–0.852); 20/50 | 0.828 (0.774–0.913); 20/50 | 0.485 (0.279–0.630); 20/50 |
| S6 | 0.088 (0.050–0.129) | 0.078 (0.047–0.113) | 0.835 (0.669–0.877); 17/50 | 0.872 (0.779–0.885); 17/50 | 0.412 (0.250–0.639); 17/50 |
| S7 | 0.038 (0.012–0.069) | 0.035 (0.014–0.060) | 0.731 (0.669–0.832); 7/50 | 0.787 (0.726–0.904); 7/50 | 0.491 (0.060–0.679); 7/50 |
| S8 | 0.087 (0.052–0.124) | 0.091 (0.061–0.123) | 0.858 (0.779–0.885); 20/50 | 0.900 (0.843–0.928); 20/50 | 0.609 (0.394–0.824); 20/50 |
| S9 | 0.012 (0.001–0.028) | 0.022 (0.003–0.049) | 0.681 (0.492–0.908); 4/50 | 0.880 (0.430–0.936); 4/50 | 0.253 (-0.097–0.566); 4/50 |
| S10 | 0.020 (0.004–0.043) | 0.017 (0.005–0.034) | 0.834 (0.615–0.971); 5/50 | 0.869 (0.741–0.994); 5/50 | 0.212 (0.017–0.276); 5/50 |
| **Cluster 3** | | | | | |
| S1 | 0.039 (0.018–0.063) | 0.079 (0.051–0.108) | 0.888 (0.713–0.927); 16/50 | 0.880 (0.821–0.948); 16/50 | 0.530 (0.443–0.645); 15/50 |
| S2 | 0.234 (0.165–0.305) | 0.157 (0.114–0.202) | 0.873 (0.849–0.903); 28/50 | 0.899 (0.886–0.929); 28/50 | 0.560 (0.474–0.669); 28/50 |
| S3 | 0.048 (0.023–0.077) | 0.086 (0.058–0.115) | 0.700 (0.583–0.804); 19/50 | 0.795 (0.693–0.878); 19/50 | 0.179 (0.044–0.436); 19/50 |
| S4 | 0.036 (0.015–0.062) | 0.080 (0.048–0.114) | 0.833 (0.793–0.889); 12/50 | 0.895 (0.844–0.947); 12/50 | 0.247 (-0.002–0.608); 12/50 |
| S5 | 0.196 (0.157–0.234) | 0.205 (0.186–0.224) | 0.847 (0.785–0.879); 42/50 | 0.903 (0.880–0.931); 42/50 | 0.506 (0.316–0.665); 42/50 |
| S6 | 0.104 (0.066–0.147) | 0.074 (0.049–0.100) | 0.843 (0.745–0.913); 22/50 | 0.905 (0.844–0.934); 22/50 | 0.432 (0.358–0.550); 22/50 |
| S7 | 0.080 (0.043–0.122) | 0.096 (0.068–0.125) | 0.830 (0.778–0.889); 21/50 | 0.903 (0.831–0.937); 21/50 | 0.345 (0.107–0.523); 21/50 |
| S8 | 0.140 (0.096–0.187) | 0.124 (0.093–0.158) | 0.855 (0.795–0.908); 31/50 | 0.914 (0.836–0.953); 31/50 | 0.613 (0.482–0.804); 31/50 |
| S9 | 0.032 (0.008–0.061) | 0.022 (0.005–0.041) | 0.879 (0.794–0.950); 6/50 | 0.906 (0.809–0.959); 6/50 | 0.736 (0.297–0.870); 6/50 |
| S10 | 0.091 (0.053–0.134) | 0.077 (0.048–0.108) | 0.856 (0.799–0.884); 16/50 | 0.900 (0.845–0.941); 16/50 | 0.646 (0.468–0.796); 16/50 |
| **Cluster 4** | | | | | |
| S1 | 0.153 (0.103–0.204) | 0.132 (0.095–0.171) | 0.857 (0.829–0.892); 31/50 | 0.895 (0.878–0.910); 31/50 | 0.725 (0.637–0.845); 31/50 |
| S2 | 0.177 (0.125–0.228) | 0.139 (0.104–0.175) | 0.767 (0.605–0.848); 29/50 | 0.846 (0.710–0.906); 29/50 | 0.465 (0.275–0.527); 29/50 |
| S3 | 0.216 (0.171–0.263) | 0.168 (0.141–0.194) | 0.856 (0.821–0.879); 39/50 | 0.895 (0.862–0.913); 39/50 | 0.667 (0.509–0.779); 39/50 |
| S4 | 0.042 (0.023–0.065) | 0.093 (0.064–0.123) | 0.760 (0.547–0.849); 17/50 | 0.860 (0.683–0.893); 17/50 | 0.178 (-0.174–0.539); 17/50 |
| S5 | 0.065 (0.042–0.090) | 0.092 (0.068–0.116) | 0.823 (0.640–0.878); 24/50 | 0.888 (0.767–0.933); 24/50 | 0.220 (0.151–0.384); 23/50 |
| S6 | 0.076 (0.034–0.130) | 0.065 (0.042–0.090) | 0.847 (0.677–0.884); 13/50 | 0.858 (0.763–0.903); 13/50 | 0.447 (0.338–0.617); 13/50 |
| S7 | 0.104 (0.072–0.140) | 0.131 (0.104–0.158) | 0.777 (0.680–0.861); 31/50 | 0.875 (0.796–0.939); 31/50 | 0.425 (0.180–0.600); 30/50 |
| S8 | 0.105 (0.066–0.148) | 0.101 (0.070–0.135) | 0.824 (0.613–0.895); 23/50 | 0.927 (0.785–0.947); 23/50 | 0.503 (0.386–0.682); 23/50 |
| S9 | 0.021 (0.002–0.051) | 0.018 (0.002–0.037) | 0.622 (0.271–0.860); 5/50 | 0.691 (0.118–0.948); 5/50 | 0.582 (0.438–0.824); 5/50 |
| S10 | 0.036 (0.016–0.059) | 0.045 (0.024–0.071) | 0.670 (0.573–0.769); 12/50 | 0.736 (0.556–0.848); 12/50 | 0.226 (0.043–0.623); 12/50 |
| S11 | 0.005 (0.000–0.015) | 0.017 (0.006–0.030) | 0.433 (0.389–0.866); 3/50 | 0.664 (0.553–0.775); 2/50 | 0.100 (-0.004–0.203); 2/50 |

**Supplementary Table S12.5. Monitoring-period-matched skill of the native AR-HMM over the causal monitoring-period-mean null on a fixed evaluation support common to all forecast horizons.** For each cluster and dense channel, the set of participant–evaluation-point keys jointly evaluable by the decoder and the monitoring-period-mean null was intersected across all five horizons, so that identical forecast origins (and hence identical causal monitoring-period lengths) were evaluated at every horizon. Monitoring-period-matched skill is defined as 1 − SSE_model_/SSE_null_ on the matched evaluation points, where positive values indicate lower squared error than the monitoring-period-mean null. n denotes the number of horizon-invariant matched evaluation points (constant across horizons by construction). Because origins and monitoring-period lengths are held fixed across horizons, any horizon dependence of skill cannot be attributed to shortening of the evaluable monitoring period; rising or sustained skill at longer horizons instead reflects captured subject-specific drift. Values are shown for the native AR-HMM under the synthetic data training regime. EMA channels are omitted because the sparse acquisition cadence precludes constructing a horizon-invariant evaluation support and these channels exhibit no horizon-dependent skill advantage for the analysis to adjudicate.

| **Cluster** | **Dense channel** | **Matched anchors, n** | **h = 1d** | **h = 3d** | **h = 7d** | **h = 14d** | **h = 21d** |
| --- | --- | --- | --- | --- | --- | --- | --- |
| **Cluster 1** | Total platform use | 1,274 | 0.018 | 0.016 | 0.040 | 0.074 | 0.095 |
|  | Night-time platform use | 1,253 | 0.220 | 0.126 | 0.132 | 0.167 | 0.171 |
|  | Sleep duration | 1,277 | 0.441 | 0.087 | −0.011 | −0.071 | −0.081 |
|  | Daily steps | 1,281 | 0.758 | 0.426 | 0.232 | 0.313 | 0.307 |
| **Cluster 2** | Total platform use | 1,482 | 0.054 | 0.022 | −0.001 | −0.008 | −0.032 |
|  | Night-time platform use | 1,438 | 0.329 | 0.182 | 0.173 | 0.162 | 0.174 |
|  | Sleep duration | 1,485 | 0.324 | −0.043 | −0.073 | 0.020 | 0.051 |
|  | Daily steps | 1,487 | 0.741 | 0.352 | 0.118 | 0.168 | 0.270 |
| **Cluster 3** | Total platform use | 1,311 | −0.009 | −0.009 | 0.044 | 0.081 | 0.088 |
|  | Night-time platform use | 1,277 | 0.107 | −0.013 | −0.020 | 0.036 | 0.054 |
|  | Sleep duration | 1,321 | 0.356 | 0.037 | 0.034 | −0.009 | −0.083 |
|  | Daily steps | 1,321 | 0.740 | 0.423 | 0.302 | 0.370 | 0.432 |
| **Cluster 4** | Total platform use | 1,334 | 0.009 | −0.007 | 0.021 | 0.009 | 0.023 |
|  | Night-time platform use | 1,305 | 0.067 | −0.031 | −0.055 | −0.041 | −0.041 |
|  | Sleep duration | 1,330 | 0.372 | 0.217 | 0.254 | 0.328 | 0.330 |
|  | Daily steps | 1,332 | 0.674 | 0.322 | 0.212 | 0.288 | 0.386 |

**Supplementary Table S12.6a.** Channel-specific pooled level-prediction performance of the native AR-HMM. Values are pooled total-variation prediction coefficients, $R_{pred,global}^{2}$, with participant-block bootstrap 95% confidence intervals.

**Panel A. Cluster 1**

| Channel | Training regime | Participants, n | h = 1d | h = 3d | h = 7d | h = 14d | h = 21d |
| --- | --- | --- | --- | --- | --- | --- | --- |
| Total platform use | Synthetic | 45-46 | 0.352 (0.197-0.458) | 0.340 (0.209-0.445) | 0.352 (0.236-0.462) | 0.336 (0.226-0.431) | 0.360 (0.251-0.441) |
| Total platform use | Hybrid | 45-46 | 0.366 (0.229-0.468) | 0.339 (0.184-0.438) | 0.342 (0.223-0.455) | 0.328 (0.204-0.419) | 0.349 (0.243-0.440) |
| Night-time platform use | Synthetic | 45-46 | 0.306 (0.230-0.375) | 0.192 (0.107-0.266) | 0.184 (0.103-0.258) | 0.194 (0.085-0.272) | 0.211 (0.101-0.293) |
| Night-time platform use | Hybrid | 45-46 | 0.309 (0.229-0.368) | 0.187 (0.095-0.263) | 0.176 (0.081-0.249) | 0.179 (0.068-0.251) | 0.192 (0.083-0.286) |
| Sleep duration | Synthetic | 46 | 0.760 (0.673-0.828) | 0.574 (0.404-0.690) | 0.501 (0.316-0.629) | 0.447 (0.210-0.589) | 0.412 (0.167-0.572) |
| Sleep duration | Hybrid | 46 | 0.771 (0.680-0.829) | 0.593 (0.414-0.697) | 0.523 (0.313-0.645) | 0.480 (0.258-0.614) | 0.440 (0.185-0.590) |
| Daily steps | Synthetic | 46 | 0.942 (0.915-0.956) | 0.846 (0.790-0.885) | 0.770 (0.681-0.819) | 0.766 (0.682-0.820) | 0.736 (0.627-0.794) |
| Daily steps | Hybrid | 46 | 0.944 (0.915-0.957) | 0.849 (0.791-0.885) | 0.770 (0.694-0.821) | 0.762 (0.664-0.813) | 0.732 (0.637-0.791) |
| Stress | Synthetic | 41-46 | 0.428 (0.288-0.553) | 0.430 (0.274-0.551) | 0.373 (0.229-0.481) | 0.350 (0.207-0.471) | 0.364 (0.226-0.475) |
| Stress | Hybrid | 41-46 | 0.426 (0.290-0.538) | 0.428 (0.282-0.554) | 0.370 (0.217-0.495) | 0.347 (0.185-0.466) | 0.362 (0.218-0.470) |
| Positive affect | Synthetic | 42-46 | 0.699 (0.555-0.794) | 0.697 (0.522-0.799) | 0.686 (0.506-0.794) | 0.663 (0.499-0.769) | 0.658 (0.449-0.790) |
| Positive affect | Hybrid | 42-46 | 0.697 (0.546-0.798) | 0.696 (0.529-0.802) | 0.685 (0.509-0.786) | 0.659 (0.498-0.768) | 0.653 (0.449-0.782) |
| Depression | Synthetic | 44-46 | 0.300 (0.128-0.410) | 0.322 (0.136-0.441) | 0.361 (0.160-0.484) | 0.396 (0.208-0.510) | 0.346 (0.174-0.481) |
| Depression | Hybrid | 44-46 | 0.292 (0.094-0.414) | 0.317 (0.133-0.451) | 0.356 (0.178-0.477) | 0.394 (0.201-0.509) | 0.345 (0.139-0.483) |

**Panel B. Cluster 2**

| Channel | Training regime | Participants, n | h = 1d | h = 3d | h = 7d | h = 14d | h = 21d |
| --- | --- | --- | --- | --- | --- | --- | --- |
| Total platform use | Synthetic | 47 | 0.333 (0.213-0.430) | 0.283 (0.165-0.378) | 0.256 (0.133-0.362) | 0.233 (0.104-0.350) | 0.224 (0.033-0.349) |
| Total platform use | Hybrid | 47 | 0.341 (0.238-0.433) | 0.281 (0.154-0.373) | 0.246 (0.104-0.341) | 0.226 (0.103-0.327) | 0.227 (0.093-0.336) |
| Night-time platform use | Synthetic | 47 | 0.390 (0.291-0.460) | 0.221 (0.132-0.294) | 0.188 (0.098-0.278) | 0.153 (0.074-0.233) | 0.149 (0.053-0.234) |
| Night-time platform use | Hybrid | 47 | 0.396 (0.305-0.464) | 0.225 (0.137-0.303) | 0.189 (0.096-0.276) | 0.152 (0.060-0.234) | 0.148 (0.056-0.222) |
| Sleep duration | Synthetic | 47 | 0.676 (0.593-0.740) | 0.459 (0.299-0.570) | 0.390 (0.207-0.549) | 0.372 (0.189-0.491) | 0.343 (0.164-0.474) |
| Sleep duration | Hybrid | 47 | 0.727 (0.658-0.772) | 0.507 (0.376-0.604) | 0.431 (0.267-0.543) | 0.397 (0.229-0.522) | 0.362 (0.203-0.495) |
| Daily steps | Synthetic | 47 | 0.952 (0.929-0.965) | 0.867 (0.798-0.902) | 0.791 (0.688-0.843) | 0.753 (0.619-0.817) | 0.720 (0.606-0.784) |
| Daily steps | Hybrid | 47 | 0.954 (0.930-0.967) | 0.870 (0.805-0.904) | 0.794 (0.702-0.851) | 0.754 (0.611-0.822) | 0.719 (0.611-0.781) |
| Stress | Synthetic | 46-47 | 0.362 (0.134-0.515) | 0.389 (0.178-0.538) | 0.396 (0.215-0.540) | 0.443 (0.283-0.570) | 0.428 (0.272-0.553) |
| Stress | Hybrid | 46-47 | 0.357 (0.106-0.527) | 0.390 (0.176-0.544) | 0.394 (0.223-0.539) | 0.435 (0.275-0.573) | 0.423 (0.260-0.547) |
| Positive affect | Synthetic | 46-47 | 0.471 (0.310-0.587) | 0.491 (0.328-0.603) | 0.509 (0.352-0.628) | 0.539 (0.419-0.637) | 0.495 (0.368-0.604) |
| Positive affect | Hybrid | 46-47 | 0.466 (0.287-0.578) | 0.490 (0.341-0.603) | 0.506 (0.373-0.626) | 0.535 (0.418-0.635) | 0.491 (0.363-0.604) |
| Depression | Synthetic | 46-47 | 0.479 (0.306-0.577) | 0.493 (0.324-0.599) | 0.470 (0.319-0.586) | 0.409 (0.213-0.521) | 0.378 (0.182-0.497) |
| Depression | Hybrid | 46-47 | 0.480 (0.338-0.600) | 0.493 (0.350-0.603) | 0.470 (0.318-0.586) | 0.408 (0.226-0.545) | 0.375 (0.215-0.497) |

**Panel C. Cluster 3**

| Channel | Training regime | Participants, n | h = 1d | h = 3d | h = 7d | h = 14d | h = 21d |
| --- | --- | --- | --- | --- | --- | --- | --- |
| Total platform use | Synthetic | 48 | 0.326 (0.229-0.395) | 0.317 (0.232-0.386) | 0.351 (0.257-0.428) | 0.352 (0.260-0.429) | 0.363 (0.270-0.440) |
| Total platform use | Hybrid | 48 | 0.333 (0.236-0.398) | 0.315 (0.225-0.378) | 0.345 (0.256-0.404) | 0.348 (0.260-0.432) | 0.356 (0.258-0.424) |
| Night-time platform use | Synthetic | 48 | 0.302 (0.230-0.361) | 0.154 (0.035-0.234) | 0.125 (-0.029-0.237) | 0.154 (-0.003-0.248) | 0.186 (0.057-0.281) |
| Night-time platform use | Hybrid | 48 | 0.304 (0.223-0.369) | 0.137 (-0.005-0.236) | 0.097 (-0.071-0.208) | 0.126 (-0.026-0.243) | 0.154 (-0.002-0.276) |
| Sleep duration | Synthetic | 48 | 0.690 (0.570-0.777) | 0.480 (0.296-0.621) | 0.411 (0.223-0.576) | 0.446 (0.253-0.581) | 0.460 (0.316-0.581) |
| Sleep duration | Hybrid | 48 | 0.716 (0.595-0.806) | 0.497 (0.320-0.628) | 0.416 (0.223-0.558) | 0.451 (0.276-0.582) | 0.471 (0.299-0.597) |
| Daily steps | Synthetic | 48 | 0.937 (0.900-0.956) | 0.841 (0.768-0.881) | 0.778 (0.685-0.839) | 0.754 (0.632-0.830) | 0.708 (0.545-0.796) |
| Daily steps | Hybrid | 48 | 0.938 (0.896-0.956) | 0.843 (0.776-0.886) | 0.777 (0.682-0.840) | 0.754 (0.617-0.836) | 0.706 (0.569-0.802) |
| Stress | Synthetic | 48 | 0.528 (0.415-0.623) | 0.517 (0.385-0.601) | 0.531 (0.407-0.615) | 0.535 (0.408-0.621) | 0.541 (0.416-0.622) |
| Stress | Hybrid | 48 | 0.526 (0.394-0.614) | 0.515 (0.391-0.607) | 0.529 (0.407-0.627) | 0.534 (0.409-0.627) | 0.540 (0.403-0.621) |
| Positive affect | Synthetic | 48 | 0.578 (0.464-0.666) | 0.580 (0.456-0.674) | 0.546 (0.410-0.644) | 0.594 (0.465-0.679) | 0.579 (0.426-0.675) |
| Positive affect | Hybrid | 48 | 0.575 (0.454-0.668) | 0.577 (0.447-0.667) | 0.540 (0.397-0.643) | 0.585 (0.456-0.677) | 0.573 (0.438-0.672) |
| Depression | Synthetic | 48 | 0.486 (0.280-0.625) | 0.487 (0.312-0.624) | 0.464 (0.234-0.618) | 0.408 (0.138-0.568) | 0.369 (0.111-0.536) |
| Depression | Hybrid | 48 | 0.486 (0.273-0.615) | 0.487 (0.298-0.619) | 0.465 (0.257-0.632) | 0.411 (0.108-0.578) | 0.372 (0.106-0.536) |

**Panel D. Cluster 4**

| Channel | Training regime | Participants, n | h = 1d | h = 3d | h = 7d | h = 14d | h = 21d |
| --- | --- | --- | --- | --- | --- | --- | --- |
| Total platform use | Synthetic | 49 | 0.357 (0.224-0.474) | 0.330 (0.189-0.448) | 0.331 (0.192-0.445) | 0.291 (0.150-0.413) | 0.279 (0.134-0.403) |
| Total platform use | Hybrid | 49 | 0.356 (0.218-0.457) | 0.328 (0.194-0.433) | 0.326 (0.177-0.445) | 0.289 (0.144-0.408) | 0.280 (0.137-0.394) |
| Night-time platform use | Synthetic | 49 | 0.142 (0.041-0.227) | 0.041 (-0.050-0.134) | -0.013 (-0.122-0.088) | 0.037 (-0.084-0.115) | 0.063 (-0.059-0.160) |
| Night-time platform use | Hybrid | 49 | 0.138 (0.044-0.238) | 0.027 (-0.067-0.112) | -0.048 (-0.175-0.055) | -0.042 (-0.227-0.091) | -0.012 (-0.154-0.102) |
| Sleep duration | Synthetic | 48-49 | 0.604 (0.456-0.703) | 0.449 (0.289-0.556) | 0.422 (0.247-0.527) | 0.451 (0.309-0.547) | 0.426 (0.271-0.541) |
| Sleep duration | Hybrid | 48-49 | 0.660 (0.542-0.742) | 0.467 (0.319-0.579) | 0.422 (0.262-0.537) | 0.446 (0.297-0.546) | 0.408 (0.269-0.524) |
| Daily steps | Synthetic | 49 | 0.935 (0.884-0.961) | 0.853 (0.768-0.905) | 0.787 (0.659-0.865) | 0.769 (0.641-0.849) | 0.752 (0.628-0.829) |
| Daily steps | Hybrid | 49 | 0.936 (0.889-0.962) | 0.856 (0.763-0.907) | 0.789 (0.645-0.867) | 0.767 (0.619-0.850) | 0.748 (0.611-0.824) |
| Stress | Synthetic | 47-49 | 0.252 (0.055-0.387) | 0.248 (0.073-0.367) | 0.269 (0.099-0.366) | 0.301 (0.127-0.418) | 0.304 (0.117-0.437) |
| Stress | Hybrid | 47-49 | 0.248 (0.059-0.378) | 0.246 (0.066-0.363) | 0.268 (0.112-0.368) | 0.302 (0.140-0.415) | 0.306 (0.110-0.435) |
| Positive affect | Synthetic | 48-49 | 0.259 (0.159-0.355) | 0.245 (0.149-0.330) | 0.262 (0.138-0.359) | 0.293 (0.161-0.401) | 0.272 (0.141-0.383) |
| Positive affect | Hybrid | 48-49 | 0.252 (0.150-0.334) | 0.240 (0.134-0.340) | 0.255 (0.141-0.366) | 0.286 (0.169-0.393) | 0.265 (0.127-0.373) |
| Depression | Synthetic | 48-49 | 0.295 (0.167-0.384) | 0.261 (0.133-0.356) | 0.210 (0.051-0.327) | 0.176 (0.003-0.305) | 0.160 (0.023-0.297) |
| Depression | Hybrid | 48-49 | 0.296 (0.165-0.395) | 0.263 (0.140-0.369) | 0.211 (0.062-0.339) | 0.174 (0.027-0.293) | 0.156 (-0.009-0.281) |

**Supplementary Table S12.6b.** Channel-specific pooled within-subject skill (1-SSE_model_/SSE_prefix-mean_) of the native AR-HMM vs. the causal participant-specific prefix-mean baseline (null). Within each cluster, channel and forecast horizon, model and baseline errors were calculated on identical participant-evaluation-point support. Values are pooled estimates with participant-block bootstrap 95%-CIs. Positive values indicate lower squared error than the null model.

**Panel A. Cluster 1**

| Channel | Training regime | Participants, n | h = 1d | h = 3d | h = 7d | h = 14d | h = 21d |
| --- | --- | --- | --- | --- | --- | --- | --- |
| Total platform use | Synthetic | 45-46 | 0.026 (-0.043-0.093) | 0.019 (-0.019-0.058) | 0.043 (0.013-0.071) | 0.073 (0.039-0.105) | 0.095 (0.054-0.139) |
| Total platform use | Hybrid | 45-46 | 0.046 (-0.013-0.107) | 0.018 (-0.018-0.054) | 0.029 (-0.003-0.059) | 0.062 (0.031-0.093) | 0.080 (0.037-0.123) |
| Night-time platform use | Synthetic | 45-46 | 0.210 (0.100-0.302) | 0.123 (0.056-0.186) | 0.129 (0.065-0.202) | 0.167 (0.095-0.240) | 0.171 (0.090-0.246) |
| Night-time platform use | Hybrid | 45-46 | 0.214 (0.108-0.306) | 0.117 (0.049-0.178) | 0.120 (0.057-0.184) | 0.151 (0.078-0.224) | 0.151 (0.073-0.221) |
| Sleep duration | Synthetic | 46 | 0.435 (0.338-0.516) | 0.078 (-0.074-0.215) | -0.023 (-0.251-0.178) | -0.078 (-0.347-0.138) | -0.081 (-0.402-0.169) |
| Sleep duration | Hybrid | 46 | 0.462 (0.367-0.544) | 0.119 (-0.009-0.235) | 0.024 (-0.171-0.181) | -0.013 (-0.230-0.152) | -0.028 (-0.299-0.173) |
| Daily steps | Synthetic | 46 | 0.750 (0.704-0.785) | 0.420 (0.333-0.491) | 0.237 (0.075-0.368) | 0.312 (0.203-0.408) | 0.307 (0.187-0.397) |
| Daily steps | Hybrid | 46 | 0.758 (0.712-0.795) | 0.433 (0.344-0.504) | 0.237 (0.071-0.363) | 0.302 (0.193-0.401) | 0.299 (0.184-0.392) |
| Stress | Synthetic | 41-46 | 0.297 (0.168-0.406) | 0.285 (0.141-0.404) | 0.246 (0.079-0.383) | 0.308 (0.155-0.438) | 0.286 (0.132-0.425) |
| Stress | Hybrid | 41-46 | 0.294 (0.168-0.408) | 0.282 (0.134-0.399) | 0.243 (0.071-0.381) | 0.306 (0.160-0.433) | 0.284 (0.131-0.424) |
| Positive affect | Synthetic | 42-46 | 0.174 (0.122-0.227) | 0.181 (0.127-0.235) | 0.208 (0.146-0.274) | 0.215 (0.156-0.279) | 0.202 (0.130-0.280) |
| Positive affect | Hybrid | 42-46 | 0.168 (0.115-0.221) | 0.178 (0.123-0.234) | 0.205 (0.145-0.269) | 0.204 (0.143-0.269) | 0.190 (0.117-0.267) |
| Depression | Synthetic | 44-46 | 0.217 (0.061-0.322) | 0.232 (0.100-0.331) | 0.301 (0.161-0.402) | 0.340 (0.195-0.439) | 0.375 (0.273-0.448) |
| Depression | Hybrid | 44-46 | 0.208 (0.053-0.312) | 0.226 (0.096-0.323) | 0.295 (0.153-0.395) | 0.338 (0.198-0.438) | 0.374 (0.279-0.449) |

**Panel B. Cluster 2**

| Channel | Training regime | Participants, n | h = 1d | h = 3d | h = 7d | h = 14d | h = 21d |
| --- | --- | --- | --- | --- | --- | --- | --- |
| Total platform use | Synthetic | 47 | 0.052 (-0.019-0.120) | 0.019 (-0.021-0.056) | 0.000 (-0.061-0.053) | -0.008 (-0.133-0.069) | -0.032 (-0.285-0.110) |
| Total platform use | Hybrid | 47 | 0.064 (-0.004-0.130) | 0.017 (-0.020-0.051) | -0.014 (-0.068-0.033) | -0.018 (-0.110-0.049) | -0.028 (-0.199-0.079) |
| Night-time platform use | Synthetic | 47 | 0.329 (0.176-0.469) | 0.184 (0.048-0.337) | 0.172 (0.047-0.304) | 0.158 (0.035-0.283) | 0.174 (0.021-0.318) |
| Night-time platform use | Hybrid | 47 | 0.335 (0.178-0.475) | 0.188 (0.051-0.341) | 0.173 (0.048-0.306) | 0.157 (0.042-0.286) | 0.173 (0.032-0.309) |
| Sleep duration | Synthetic | 47 | 0.326 (0.116-0.470) | -0.041 (-0.432-0.191) | -0.068 (-0.536-0.187) | 0.023 (-0.356-0.257) | 0.051 (-0.265-0.254) |
| Sleep duration | Hybrid | 47 | 0.430 (0.268-0.540) | 0.051 (-0.262-0.243) | 0.003 (-0.384-0.215) | 0.062 (-0.247-0.255) | 0.079 (-0.180-0.264) |
| Daily steps | Synthetic | 47 | 0.741 (0.683-0.787) | 0.352 (0.230-0.448) | 0.116 (-0.054-0.253) | 0.165 (0.008-0.303) | 0.270 (0.155-0.376) |
| Daily steps | Hybrid | 47 | 0.751 (0.699-0.792) | 0.369 (0.272-0.447) | 0.126 (-0.013-0.236) | 0.170 (0.037-0.289) | 0.267 (0.162-0.360) |
| Stress | Synthetic | 46-47 | 0.394 (0.251-0.505) | 0.361 (0.217-0.476) | 0.305 (0.224-0.381) | 0.350 (0.264-0.422) | 0.351 (0.261-0.437) |
| Stress | Hybrid | 46-47 | 0.389 (0.251-0.499) | 0.362 (0.216-0.475) | 0.303 (0.221-0.378) | 0.341 (0.258-0.413) | 0.344 (0.254-0.435) |
| Positive affect | Synthetic | 46-47 | 0.228 (0.133-0.319) | 0.219 (0.128-0.309) | 0.210 (0.116-0.300) | 0.244 (0.152-0.330) | 0.231 (0.135-0.320) |
| Positive affect | Hybrid | 46-47 | 0.220 (0.130-0.305) | 0.218 (0.136-0.305) | 0.206 (0.114-0.294) | 0.237 (0.154-0.316) | 0.225 (0.137-0.311) |
| Depression | Synthetic | 46-47 | 0.354 (0.223-0.451) | 0.354 (0.218-0.451) | 0.304 (0.171-0.403) | 0.319 (0.194-0.407) | 0.358 (0.264-0.433) |
| Depression | Hybrid | 46-47 | 0.355 (0.216-0.452) | 0.354 (0.223-0.449) | 0.305 (0.169-0.398) | 0.318 (0.190-0.403) | 0.355 (0.257-0.429) |

**Panel C. Cluster 3**

| Channel | Training regime | Participants, n | h = 1d | h = 3d | h = 7d | h = 14d | h = 21d |
| --- | --- | --- | --- | --- | --- | --- | --- |
| Total platform use | Synthetic | 48 | -0.009 (-0.063-0.037) | -0.011 (-0.053-0.030) | 0.045 (0.006-0.081) | 0.081 (0.042-0.121) | 0.088 (0.039-0.140) |
| Total platform use | Hybrid | 48 | 0.002 (-0.040-0.044) | -0.014 (-0.053-0.022) | 0.037 (-0.002-0.073) | 0.076 (0.031-0.117) | 0.079 (0.025-0.131) |
| Night-time platform use | Synthetic | 48 | 0.120 (0.038-0.191) | -0.015 (-0.118-0.052) | -0.020 (-0.146-0.067) | 0.026 (-0.067-0.098) | 0.054 (-0.039-0.131) |
| Night-time platform use | Hybrid | 48 | 0.123 (0.051-0.189) | -0.035 (-0.137-0.036) | -0.053 (-0.180-0.038) | -0.006 (-0.119-0.078) | 0.016 (-0.095-0.108) |
| Sleep duration | Synthetic | 48 | 0.356 (0.273-0.436) | 0.037 (-0.087-0.142) | 0.034 (-0.118-0.168) | -0.009 (-0.193-0.151) | -0.083 (-0.252-0.084) |
| Sleep duration | Hybrid | 48 | 0.409 (0.327-0.496) | 0.068 (-0.053-0.161) | 0.043 (-0.105-0.160) | 0.001 (-0.181-0.151) | -0.061 (-0.231-0.094) |
| Daily steps | Synthetic | 48 | 0.740 (0.650-0.804) | 0.423 (0.295-0.525) | 0.302 (0.181-0.394) | 0.370 (0.271-0.442) | 0.432 (0.365-0.486) |
| Daily steps | Hybrid | 48 | 0.745 (0.655-0.810) | 0.430 (0.300-0.528) | 0.300 (0.178-0.397) | 0.370 (0.273-0.445) | 0.428 (0.362-0.482) |
| Stress | Synthetic | 48 | 0.232 (0.103-0.342) | 0.219 (0.093-0.327) | 0.264 (0.175-0.346) | 0.276 (0.187-0.352) | 0.214 (0.057-0.337) |
| Stress | Hybrid | 48 | 0.229 (0.102-0.336) | 0.216 (0.085-0.323) | 0.261 (0.174-0.342) | 0.275 (0.185-0.353) | 0.211 (0.057-0.334) |
| Positive affect | Synthetic | 48 | 0.258 (0.185-0.333) | 0.240 (0.170-0.318) | 0.255 (0.195-0.315) | 0.277 (0.201-0.355) | 0.280 (0.203-0.356) |
| Positive affect | Hybrid | 48 | 0.252 (0.181-0.326) | 0.233 (0.165-0.308) | 0.244 (0.185-0.306) | 0.262 (0.187-0.337) | 0.269 (0.195-0.341) |
| Depression | Synthetic | 48 | 0.219 (0.043-0.347) | 0.237 (0.042-0.364) | 0.203 (-0.020-0.344) | 0.186 (-0.087-0.409) | 0.228 (0.014-0.406) |
| Depression | Hybrid | 48 | 0.218 (0.040-0.346) | 0.237 (0.044-0.364) | 0.204 (-0.014-0.342) | 0.189 (-0.069-0.401) | 0.231 (0.018-0.408) |

**Panel D. Cluster 4**

| Channel | Training regime | Participants, n | h = 1d | h = 3d | h = 7d | h = 14d | h = 21d |
| --- | --- | --- | --- | --- | --- | --- | --- |
| Total platform use | Synthetic | 49 | 0.012 (-0.044-0.069) | -0.007 (-0.068-0.051) | 0.023 (-0.033-0.074) | 0.009 (-0.072-0.075) | 0.023 (-0.043-0.080) |
| Total platform use | Hybrid | 49 | 0.011 (-0.043-0.063) | -0.011 (-0.069-0.047) | 0.016 (-0.034-0.063) | 0.007 (-0.066-0.071) | 0.025 (-0.038-0.079) |
| Night-time platform use | Synthetic | 49 | 0.062 (-0.012-0.137) | -0.029 (-0.100-0.030) | -0.064 (-0.166-0.019) | -0.051 (-0.167-0.038) | -0.041 (-0.185-0.071) |
| Night-time platform use | Hybrid | 49 | 0.058 (-0.016-0.134) | -0.044 (-0.119-0.021) | -0.102 (-0.236-0.007) | -0.137 (-0.318-0.001) | -0.124 (-0.325-0.031) |
| Sleep duration | Synthetic | 48-49 | 0.365 (0.216-0.477) | 0.202 (0.045-0.335) | 0.240 (0.061-0.384) | 0.320 (0.172-0.438) | 0.330 (0.189-0.452) |
| Sleep duration | Hybrid | 48-49 | 0.455 (0.356-0.540) | 0.228 (0.081-0.350) | 0.240 (0.069-0.379) | 0.314 (0.159-0.436) | 0.310 (0.161-0.438) |
| Daily steps | Synthetic | 49 | 0.658 (0.585-0.732) | 0.328 (0.207-0.431) | 0.222 (0.083-0.353) | 0.291 (0.169-0.409) | 0.386 (0.287-0.480) |
| Daily steps | Hybrid | 49 | 0.663 (0.593-0.734) | 0.339 (0.222-0.441) | 0.228 (0.097-0.355) | 0.285 (0.168-0.406) | 0.377 (0.276-0.476) |
| Stress | Synthetic | 47-49 | 0.366 (0.235-0.457) | 0.341 (0.215-0.434) | 0.262 (0.134-0.354) | 0.316 (0.152-0.429) | 0.372 (0.193-0.486) |
| Stress | Hybrid | 47-49 | 0.362 (0.234-0.452) | 0.339 (0.213-0.431) | 0.261 (0.137-0.355) | 0.317 (0.160-0.434) | 0.373 (0.205-0.488) |
| Positive affect | Synthetic | 48-49 | 0.231 (0.175-0.290) | 0.235 (0.183-0.285) | 0.259 (0.202-0.311) | 0.256 (0.199-0.312) | 0.267 (0.191-0.336) |
| Positive affect | Hybrid | 48-49 | 0.224 (0.166-0.284) | 0.229 (0.174-0.282) | 0.251 (0.195-0.302) | 0.249 (0.193-0.302) | 0.259 (0.183-0.330) |
| Depression | Synthetic | 48-49 | 0.254 (0.110-0.380) | 0.247 (0.113-0.363) | 0.235 (0.091-0.351) | 0.242 (0.064-0.374) | 0.250 (0.141-0.359) |
| Depression | Hybrid | 48-49 | 0.256 (0.115-0.381) | 0.250 (0.116-0.364) | 0.235 (0.090-0.352) | 0.240 (0.072-0.369) | 0.247 (0.138-0.356) |

**Supplementary Table S12.7a.** Rolling-origin directional classification of future deviations. Values are percentages with participant-block bootstrap 95%-CIs; 50% denotes chance-level balanced accuracy.

**Panel A. Cluster 1**

| Channel | Training regime | Participants, n | h = 1d | h = 3d | h = 7d | h = 14d | h = 21d |
| --- | --- | --- | --- | --- | --- | --- | --- |
| Total platform use | Synthetic | 45-46 | 60.5% (57.1-63.6%) | 56.0% (52.7-59.3%) | 59.1% (55.5-62.5%) | 63.2% (59.2-66.8%) | 62.9% (58.9-66.5%) |
| Total platform use | Hybrid | 45-46 | 60.9% (57.6-63.7%) | 56.7% (53.3-60.1%) | 58.8% (55.2-62.1%) | 61.6% (57.7-65.2%) | 62.3% (57.8-66.2%) |
| Night-time platform use | Synthetic | 45-46 | 65.2% (61.7-68.9%) | 61.0% (56.3-64.8%) | 59.9% (55.3-63.5%) | 62.9% (58.4-66.9%) | 62.3% (57.2-66.8%) |
| Night-time platform use | Hybrid | 45-46 | 64.5% (60.6-68.2%) | 60.6% (55.8-64.5%) | 60.0% (56.3-64.0%) | 61.6% (57.8-66.1%) | 60.5% (55.5-64.9%) |
| Sleep duration | Synthetic | 46 | 75.8% (72.2-79.0%) | 56.8% (50.8-61.8%) | 53.0% (45.7-59.4%) | 53.1% (47.2-58.7%) | 50.8% (45.2-57.2%) |
| Sleep duration | Hybrid | 46 | 76.4% (72.9-80.2%) | 59.7% (54.3-65.0%) | 54.6% (47.1-61.2%) | 54.5% (47.1-60.5%) | 54.3% (48.0-60.2%) |
| Daily steps | Synthetic | 46 | 86.1% (83.4-88.2%) | 72.3% (66.3-77.5%) | 67.6% (60.5-74.5%) | 69.5% (62.7-75.1%) | 66.1% (58.0-73.3%) |
| Daily steps | Hybrid | 46 | 86.3% (83.7-88.2%) | 72.4% (66.7-77.8%) | 67.4% (60.1-73.9%) | 68.6% (61.8-74.7%) | 65.5% (56.9-72.7%) |
| Stress | Synthetic | 41-46 | 72.7% (64.0-79.9%) | 70.3% (62.8-77.3%) | 68.5% (60.1-76.2%) | 72.4% (62.6-80.1%) | 73.8% (63.6-82.1%) |
| Stress | Hybrid | 41-46 | 72.7% (65.2-80.3%) | 70.3% (62.7-76.9%) | 68.5% (59.8-76.7%) | 72.4% (62.9-80.7%) | 73.8% (65.2-82.0%) |
| Positive affect | Synthetic | 42-46 | 66.9% (59.1-72.6%) | 65.7% (58.2-72.7%) | 68.4% (60.4-75.7%) | 73.5% (65.5-80.9%) | 73.0% (64.9-79.0%) |
| Positive affect | Hybrid | 42-46 | 68.0% (60.4-75.2%) | 67.3% (59.4-73.7%) | 70.5% (63.1-76.6%) | 72.7% (64.2-80.0%) | 72.5% (63.6-79.3%) |
| Depression | Synthetic | 44-46 | 75.3% (65.2-84.0%) | 72.4% (63.1-80.9%) | 77.8% (68.4-86.5%) | 80.0% (71.2-87.6%) | 74.6% (66.3-81.6%) |
| Depression | Hybrid | 44-46 | 75.3% (65.3-86.2%) | 72.4% (63.1-80.8%) | 77.8% (68.8-87.7%) | 80.0% (71.1-88.2%) | 74.6% (66.6-81.1%) |

**Panel B. Cluster 2**

| Channel | Training regime | Participants, n | h = 1d | h = 3d | h = 7d | h = 14d | h = 21d |
| --- | --- | --- | --- | --- | --- | --- | --- |
| Total platform use | Synthetic | 47 | 58.5% (55.6-61.0%) | 56.9% (53.9-59.4%) | 54.9% (51.3-58.2%) | 58.4% (55.0-61.5%) | 61.2% (57.5-65.0%) |
| Total platform use | Hybrid | 47 | 59.3% (56.8-62.0%) | 56.7% (53.8-59.4%) | 54.9% (51.9-58.3%) | 56.6% (53.4-59.7%) | 58.0% (54.0-61.9%) |
| Night-time platform use | Synthetic | 47 | 66.2% (62.5-70.2%) | 62.6% (57.7-67.3%) | 62.8% (58.0-67.2%) | 62.4% (57.0-67.2%) | 63.3% (58.1-67.7%) |
| Night-time platform use | Hybrid | 47 | 66.5% (62.1-70.7%) | 62.4% (57.4-67.2%) | 61.9% (57.1-67.0%) | 62.2% (57.2-67.0%) | 62.4% (56.9-67.6%) |
| Sleep duration | Synthetic | 47 | 75.1% (71.5-78.7%) | 61.6% (56.6-66.3%) | 59.9% (54.7-65.0%) | 58.5% (53.1-63.8%) | 59.3% (52.8-65.7%) |
| Sleep duration | Hybrid | 47 | 77.1% (73.7-80.7%) | 64.1% (59.8-67.7%) | 61.0% (56.9-64.9%) | 59.6% (54.3-64.5%) | 59.8% (53.8-65.7%) |
| Daily steps | Synthetic | 47 | 82.8% (80.1-85.1%) | 66.9% (61.3-72.2%) | 62.5% (55.6-68.6%) | 65.3% (59.0-71.6%) | 69.8% (63.4-75.7%) |
| Daily steps | Hybrid | 47 | 83.7% (80.5-86.2%) | 67.6% (61.9-71.9%) | 62.7% (56.9-68.5%) | 65.4% (58.8-70.7%) | 70.8% (63.8-76.4%) |
| Stress | Synthetic | 46-47 | 73.5% (64.9-80.8%) | 72.7% (63.9-81.2%) | 69.6% (60.9-77.8%) | 74.3% (66.2-81.7%) | 74.0% (65.1-82.7%) |
| Stress | Hybrid | 46-47 | 73.5% (65.4-81.4%) | 72.7% (62.5-81.9%) | 69.6% (60.4-79.2%) | 74.3% (65.8-81.2%) | 74.0% (64.7-81.9%) |
| Positive affect | Synthetic | 46-47 | 66.5% (59.3-72.3%) | 65.5% (58.7-71.8%) | 66.5% (59.5-72.7%) | 67.9% (62.2-73.9%) | 67.4% (60.1-74.0%) |
| Positive affect | Hybrid | 46-47 | 64.7% (58.2-71.2%) | 65.2% (57.9-71.1%) | 66.6% (59.5-73.2%) | 65.9% (59.0-72.2%) | 65.4% (58.3-72.2%) |
| Depression | Synthetic | 46-47 | 71.2% (61.9-79.6%) | 72.0% (62.0-80.7%) | 72.4% (63.5-80.7%) | 72.4% (61.5-83.4%) | 70.9% (61.7-78.7%) |
| Depression | Hybrid | 46-47 | 71.2% (61.4-79.2%) | 72.0% (63.5-81.2%) | 72.8% (62.5-81.3%) | 72.8% (62.0-82.7%) | 70.4% (61.3-78.5%) |

**Panel C. Cluster 3**

| Channel | Training regime | Participants, n | h = 1d | h = 3d | h = 7d | h = 14d | h = 21d |
| --- | --- | --- | --- | --- | --- | --- | --- |
| Total platform use | Synthetic | 48 | 56.8% (53.6-60.0%) | 54.3% (50.9-57.8%) | 57.3% (53.6-61.3%) | 57.8% (54.0-61.4%) | 56.7% (53.0-60.2%) |
| Total platform use | Hybrid | 48 | 56.6% (53.5-59.7%) | 54.8% (51.4-58.3%) | 57.0% (53.3-60.7%) | 57.6% (53.1-61.5%) | 57.4% (53.9-60.8%) |
| Night-time platform use | Synthetic | 48 | 59.9% (56.4-63.4%) | 54.3% (51.5-57.5%) | 53.8% (50.3-57.9%) | 54.4% (50.8-58.4%) | 53.8% (50.1-57.7%) |
| Night-time platform use | Hybrid | 48 | 59.8% (56.2-63.3%) | 53.7% (50.6-57.3%) | 53.8% (50.3-57.8%) | 54.1% (50.0-58.9%) | 54.2% (50.0-58.0%) |
| Sleep duration | Synthetic | 48 | 73.1% (68.6-77.1%) | 60.5% (55.5-65.4%) | 59.0% (53.8-65.4%) | 56.9% (49.7-63.5%) | 57.8% (51.2-64.9%) |
| Sleep duration | Hybrid | 48 | 74.2% (69.9-77.9%) | 62.4% (57.3-67.7%) | 58.8% (52.8-64.2%) | 56.8% (50.2-63.6%) | 56.3% (49.2-63.0%) |
| Daily steps | Synthetic | 48 | 81.9% (78.8-84.7%) | 67.7% (62.8-72.2%) | 66.4% (61.4-71.5%) | 71.8% (66.7-77.0%) | 77.0% (70.0-82.0%) |
| Daily steps | Hybrid | 48 | 82.3% (79.0-85.0%) | 69.0% (64.6-73.2%) | 66.9% (61.2-72.0%) | 71.9% (65.3-76.9%) | 77.0% (70.3-81.9%) |
| Stress | Synthetic | 48 | 70.7% (63.3-77.2%) | 71.1% (64.3-78.3%) | 75.6% (68.0-82.2%) | 78.2% (70.7-86.2%) | 78.0% (70.4-85.0%) |
| Stress | Hybrid | 48 | 70.7% (63.2-77.0%) | 71.1% (63.9-78.6%) | 75.6% (68.6-82.6%) | 78.2% (70.5-85.9%) | 78.0% (69.5-84.8%) |
| Positive affect | Synthetic | 48 | 70.0% (62.9-76.2%) | 68.6% (61.3-75.2%) | 71.5% (65.3-76.7%) | 73.5% (66.9-78.9%) | 74.7% (67.9-79.9%) |
| Positive affect | Hybrid | 48 | 70.2% (62.9-76.5%) | 68.7% (61.8-75.2%) | 71.3% (65.2-76.2%) | 71.6% (65.0-77.6%) | 73.4% (65.9-78.8%) |
| Depression | Synthetic | 48 | 55.0% (46.7-65.0%) | 54.4% (45.9-63.2%) | 54.5% (45.7-65.4%) | 56.6% (44.9-69.7%) | 65.3% (55.6-76.0%) |
| Depression | Hybrid | 48 | 55.0% (46.8-64.3%) | 54.4% (45.4-65.3%) | 54.5% (44.1-64.3%) | 56.6% (46.0-68.8%) | 65.3% (55.3-76.8%) |

**Panel D. Cluster 4**

| Channel | Training regime | Participants, n | h = 1d | h = 3d | h = 7d | h = 14d | h = 21d |
| --- | --- | --- | --- | --- | --- | --- | --- |
| Total platform use | Synthetic | 49 | 58.9% (55.6-62.1%) | 56.6% (53.7-59.6%) | 57.2% (53.7-60.6%) | 57.4% (54.5-60.5%) | 57.2% (53.5-60.7%) |
| Total platform use | Hybrid | 49 | 58.1% (54.4-61.5%) | 54.0% (51.0-57.1%) | 54.9% (51.3-58.4%) | 53.6% (49.5-57.4%) | 55.3% (51.4-59.1%) |
| Night-time platform use | Synthetic | 49 | 63.7% (60.2-67.0%) | 54.5% (52.2-57.4%) | 53.2% (50.9-56.2%) | 52.1% (50.5-53.8%) | 52.1% (50.6-54.6%) |
| Night-time platform use | Hybrid | 49 | 62.4% (59.0-65.4%) | 53.9% (51.4-56.6%) | 52.6% (50.5-55.7%) | 51.6% (50.2-53.4%) | 51.6% (50.1-53.5%) |
| Sleep duration | Synthetic | 48-49 | 72.9% (67.4-77.4%) | 65.3% (59.0-70.9%) | 68.2% (61.8-74.3%) | 68.5% (60.7-74.8%) | 66.6% (58.9-74.1%) |
| Sleep duration | Hybrid | 48-49 | 73.7% (69.1-78.0%) | 66.4% (60.4-71.3%) | 67.1% (61.3-73.1%) | 67.1% (59.9-74.2%) | 66.1% (58.2-72.7%) |
| Daily steps | Synthetic | 49 | 81.7% (77.7-85.1%) | 65.6% (59.9-71.1%) | 63.1% (56.9-68.8%) | 64.7% (58.3-70.9%) | 70.9% (63.1-76.1%) |
| Daily steps | Hybrid | 49 | 81.9% (77.9-84.8%) | 66.2% (61.0-71.4%) | 62.8% (57.0-68.4%) | 64.4% (58.2-70.7%) | 70.9% (63.6-77.3%) |
| Stress | Synthetic | 47-49 | 75.0% (67.8-81.6%) | 72.6% (64.5-79.8%) | 71.2% (62.6-79.0%) | 69.4% (59.5-78.4%) | 72.9% (61.9-82.5%) |
| Stress | Hybrid | 47-49 | 74.3% (67.5-81.2%) | 72.5% (64.1-80.1%) | 71.2% (62.3-78.1%) | 71.1% (62.9-79.3%) | 72.9% (61.3-82.7%) |
| Positive affect | Synthetic | 48-49 | 70.1% (65.3-74.3%) | 70.9% (65.1-75.2%) | 74.8% (69.0-79.1%) | 74.4% (67.5-79.8%) | 77.2% (70.1-83.4%) |
| Positive affect | Hybrid | 48-49 | 71.1% (65.9-76.2%) | 72.5% (67.7-77.6%) | 75.5% (70.3-80.5%) | 75.7% (68.9-82.0%) | 77.2% (69.5-83.4%) |
| Depression | Synthetic | 48-49 | 63.9% (54.1-72.9%) | 64.3% (55.2-73.5%) | 63.5% (54.3-71.7%) | 65.3% (58.0-72.9%) | 65.6% (58.9-71.9%) |
| Depression | Hybrid | 48-49 | 63.9% (54.0-73.2%) | 64.3% (54.8-72.7%) | 63.5% (55.4-72.0%) | 65.3% (57.4-72.5%) | 65.6% (59.6-72.2%) |

**Supplementary Table S12.7b.** Rolling-origin correspondence of future deviations from the causal history mean. Values are pooled Spearman correlations with participant-block bootstrap 95%-CIs.

**Panel A. Cluster 1**

| Channel | Training regime | Participants, n | h = 1d | h = 3d | h = 7d | h = 14d | h = 21d |
| --- | --- | --- | --- | --- | --- | --- | --- |
| Total platform use | Synthetic | 45-46 | 0.292 (0.209-0.365) | 0.212 (0.143-0.282) | 0.263 (0.201-0.317) | 0.336 (0.270-0.396) | 0.362 (0.285-0.427) |
| Total platform use | Hybrid | 45-46 | 0.301 (0.221-0.370) | 0.202 (0.118-0.271) | 0.239 (0.175-0.300) | 0.320 (0.243-0.382) | 0.339 (0.255-0.418) |
| Night-time platform use | Synthetic | 45-46 | 0.443 (0.344-0.541) | 0.359 (0.247-0.461) | 0.379 (0.297-0.451) | 0.425 (0.315-0.502) | 0.434 (0.308-0.526) |
| Night-time platform use | Hybrid | 45-46 | 0.440 (0.332-0.531) | 0.339 (0.243-0.435) | 0.359 (0.268-0.445) | 0.393 (0.286-0.474) | 0.404 (0.275-0.502) |
| Sleep duration | Synthetic | 46 | 0.661 (0.584-0.740) | 0.322 (0.160-0.477) | 0.254 (0.063-0.429) | 0.188 (-0.017-0.370) | 0.137 (-0.053-0.340) |
| Sleep duration | Hybrid | 46 | 0.675 (0.607-0.750) | 0.334 (0.193-0.471) | 0.251 (0.081-0.415) | 0.212 (0.045-0.395) | 0.178 (0.007-0.348) |
| Daily steps | Synthetic | 46 | 0.854 (0.812-0.883) | 0.641 (0.542-0.714) | 0.527 (0.380-0.646) | 0.568 (0.428-0.679) | 0.539 (0.364-0.662) |
| Daily steps | Hybrid | 46 | 0.859 (0.822-0.887) | 0.647 (0.554-0.720) | 0.516 (0.380-0.627) | 0.550 (0.400-0.666) | 0.525 (0.347-0.659) |
| Stress | Synthetic | 41-46 | 0.534 (0.417-0.625) | 0.508 (0.389-0.609) | 0.478 (0.336-0.610) | 0.566 (0.414-0.663) | 0.531 (0.377-0.661) |
| Stress | Hybrid | 41-46 | 0.527 (0.417-0.615) | 0.500 (0.361-0.603) | 0.471 (0.333-0.591) | 0.563 (0.403-0.674) | 0.526 (0.359-0.653) |
| Positive affect | Synthetic | 42-46 | 0.382 (0.265-0.466) | 0.389 (0.282-0.475) | 0.447 (0.333-0.536) | 0.530 (0.434-0.609) | 0.494 (0.360-0.579) |
| Positive affect | Hybrid | 42-46 | 0.390 (0.274-0.475) | 0.397 (0.290-0.484) | 0.453 (0.330-0.541) | 0.522 (0.412-0.609) | 0.479 (0.357-0.566) |
| Depression | Synthetic | 44-46 | 0.624 (0.504-0.728) | 0.583 (0.454-0.687) | 0.623 (0.502-0.714) | 0.634 (0.486-0.735) | 0.609 (0.507-0.688) |
| Depression | Hybrid | 44-46 | 0.615 (0.500-0.715) | 0.580 (0.454-0.705) | 0.617 (0.482-0.731) | 0.632 (0.469-0.732) | 0.609 (0.492-0.687) |

**Panel B. Cluster 2**

| Channel | Training regime | Participants, n | h = 1d | h = 3d | h = 7d | h = 14d | h = 21d |
| --- | --- | --- | --- | --- | --- | --- | --- |
| Total platform use | Synthetic | 47 | 0.297 (0.227-0.352) | 0.211 (0.152-0.265) | 0.209 (0.129-0.288) | 0.234 (0.144-0.313) | 0.283 (0.154-0.384) |
| Total platform use | Hybrid | 47 | 0.295 (0.225-0.348) | 0.202 (0.143-0.263) | 0.171 (0.098-0.247) | 0.192 (0.110-0.286) | 0.227 (0.085-0.330) |
| Night-time platform use | Synthetic | 47 | 0.516 (0.412-0.622) | 0.405 (0.285-0.544) | 0.400 (0.278-0.513) | 0.395 (0.280-0.517) | 0.411 (0.260-0.533) |
| Night-time platform use | Hybrid | 47 | 0.513 (0.409-0.616) | 0.400 (0.262-0.529) | 0.389 (0.252-0.507) | 0.383 (0.250-0.499) | 0.398 (0.268-0.524) |
| Sleep duration | Synthetic | 47 | 0.631 (0.548-0.704) | 0.365 (0.265-0.456) | 0.333 (0.208-0.449) | 0.347 (0.227-0.468) | 0.342 (0.185-0.480) |
| Sleep duration | Hybrid | 47 | 0.680 (0.605-0.755) | 0.421 (0.335-0.510) | 0.371 (0.272-0.476) | 0.362 (0.240-0.479) | 0.344 (0.188-0.463) |
| Daily steps | Synthetic | 47 | 0.839 (0.804-0.865) | 0.549 (0.464-0.631) | 0.356 (0.223-0.483) | 0.422 (0.275-0.542) | 0.523 (0.398-0.614) |
| Daily steps | Hybrid | 47 | 0.845 (0.810-0.873) | 0.570 (0.490-0.645) | 0.364 (0.221-0.484) | 0.426 (0.297-0.536) | 0.528 (0.398-0.617) |
| Stress | Synthetic | 46-47 | 0.609 (0.493-0.704) | 0.594 (0.482-0.692) | 0.557 (0.456-0.649) | 0.610 (0.512-0.691) | 0.598 (0.464-0.688) |
| Stress | Hybrid | 46-47 | 0.608 (0.481-0.707) | 0.593 (0.465-0.685) | 0.547 (0.438-0.639) | 0.601 (0.507-0.676) | 0.593 (0.467-0.684) |
| Positive affect | Synthetic | 46-47 | 0.471 (0.346-0.571) | 0.458 (0.324-0.556) | 0.467 (0.348-0.570) | 0.487 (0.361-0.584) | 0.492 (0.370-0.604) |
| Positive affect | Hybrid | 46-47 | 0.455 (0.331-0.561) | 0.453 (0.324-0.560) | 0.468 (0.344-0.569) | 0.487 (0.374-0.580) | 0.491 (0.376-0.598) |
| Depression | Synthetic | 46-47 | 0.486 (0.350-0.608) | 0.479 (0.330-0.593) | 0.492 (0.337-0.620) | 0.525 (0.384-0.649) | 0.604 (0.473-0.701) |
| Depression | Hybrid | 46-47 | 0.488 (0.355-0.610) | 0.482 (0.339-0.595) | 0.493 (0.347-0.608) | 0.522 (0.376-0.644) | 0.598 (0.466-0.691) |

**Panel C. Cluster 3**

| Channel | Training regime | Participants, n | h = 1d | h = 3d | h = 7d | h = 14d | h = 21d |
| --- | --- | --- | --- | --- | --- | --- | --- |
| Total platform use | Synthetic | 48 | 0.196 (0.130-0.258) | 0.138 (0.059-0.217) | 0.201 (0.096-0.281) | 0.221 (0.139-0.309) | 0.242 (0.162-0.318) |
| Total platform use | Hybrid | 48 | 0.191 (0.120-0.253) | 0.128 (0.052-0.203) | 0.181 (0.087-0.262) | 0.216 (0.126-0.299) | 0.228 (0.148-0.313) |
| Night-time platform use | Synthetic | 48 | 0.360 (0.272-0.443) | 0.277 (0.187-0.353) | 0.305 (0.203-0.389) | 0.318 (0.210-0.411) | 0.348 (0.242-0.442) |
| Night-time platform use | Hybrid | 48 | 0.365 (0.274-0.439) | 0.243 (0.124-0.331) | 0.228 (0.117-0.337) | 0.259 (0.142-0.370) | 0.290 (0.162-0.398) |
| Sleep duration | Synthetic | 48 | 0.608 (0.524-0.672) | 0.310 (0.217-0.417) | 0.295 (0.178-0.417) | 0.244 (0.091-0.379) | 0.223 (0.073-0.380) |
| Sleep duration | Hybrid | 48 | 0.653 (0.575-0.721) | 0.331 (0.220-0.424) | 0.292 (0.161-0.415) | 0.243 (0.110-0.404) | 0.226 (0.070-0.382) |
| Daily steps | Synthetic | 48 | 0.862 (0.827-0.890) | 0.625 (0.527-0.712) | 0.527 (0.410-0.630) | 0.606 (0.484-0.687) | 0.696 (0.596-0.761) |
| Daily steps | Hybrid | 48 | 0.864 (0.828-0.891) | 0.630 (0.536-0.704) | 0.525 (0.397-0.624) | 0.607 (0.490-0.706) | 0.696 (0.591-0.767) |
| Stress | Synthetic | 48 | 0.462 (0.348-0.563) | 0.454 (0.340-0.555) | 0.509 (0.394-0.601) | 0.531 (0.397-0.632) | 0.512 (0.380-0.645) |
| Stress | Hybrid | 48 | 0.459 (0.331-0.557) | 0.451 (0.327-0.547) | 0.506 (0.371-0.598) | 0.530 (0.407-0.647) | 0.511 (0.353-0.636) |
| Positive affect | Synthetic | 48 | 0.513 (0.411-0.602) | 0.487 (0.382-0.575) | 0.538 (0.441-0.603) | 0.531 (0.427-0.611) | 0.541 (0.419-0.644) |
| Positive affect | Hybrid | 48 | 0.503 (0.395-0.588) | 0.478 (0.363-0.565) | 0.528 (0.436-0.593) | 0.520 (0.424-0.599) | 0.530 (0.426-0.609) |
| Depression | Synthetic | 48 | 0.440 (0.286-0.590) | 0.431 (0.256-0.577) | 0.464 (0.310-0.592) | 0.467 (0.261-0.625) | 0.516 (0.332-0.660) |
| Depression | Hybrid | 48 | 0.434 (0.290-0.575) | 0.429 (0.236-0.580) | 0.465 (0.301-0.598) | 0.466 (0.273-0.630) | 0.512 (0.350-0.658) |

**Panel D. Cluster 4**

| Channel | Training regime | Participants, n | h = 1d | h = 3d | h = 7d | h = 14d | h = 21d |
| --- | --- | --- | --- | --- | --- | --- | --- |
| Total platform use | Synthetic | 49 | 0.265 (0.202-0.326) | 0.156 (0.061-0.240) | 0.213 (0.140-0.291) | 0.199 (0.112-0.285) | 0.218 (0.132-0.290) |
| Total platform use | Hybrid | 49 | 0.250 (0.186-0.311) | 0.137 (0.054-0.220) | 0.187 (0.108-0.265) | 0.179 (0.079-0.266) | 0.208 (0.127-0.292) |
| Night-time platform use | Synthetic | 49 | 0.378 (0.297-0.460) | 0.282 (0.202-0.358) | 0.320 (0.225-0.409) | 0.288 (0.194-0.365) | 0.264 (0.166-0.354) |
| Night-time platform use | Hybrid | 49 | 0.369 (0.295-0.442) | 0.245 (0.152-0.323) | 0.299 (0.202-0.402) | 0.253 (0.166-0.348) | 0.246 (0.144-0.346) |
| Sleep duration | Synthetic | 48-49 | 0.599 (0.491-0.693) | 0.447 (0.287-0.578) | 0.469 (0.298-0.613) | 0.522 (0.343-0.653) | 0.522 (0.362-0.644) |
| Sleep duration | Hybrid | 48-49 | 0.652 (0.574-0.722) | 0.466 (0.323-0.588) | 0.465 (0.299-0.608) | 0.518 (0.360-0.649) | 0.509 (0.325-0.630) |
| Daily steps | Synthetic | 49 | 0.821 (0.771-0.863) | 0.543 (0.423-0.642) | 0.449 (0.337-0.556) | 0.515 (0.385-0.612) | 0.634 (0.548-0.694) |
| Daily steps | Hybrid | 49 | 0.827 (0.772-0.868) | 0.555 (0.439-0.642) | 0.449 (0.309-0.563) | 0.510 (0.397-0.610) | 0.628 (0.538-0.686) |
| Stress | Synthetic | 47-49 | 0.559 (0.438-0.661) | 0.528 (0.402-0.634) | 0.459 (0.335-0.578) | 0.507 (0.361-0.636) | 0.573 (0.404-0.725) |
| Stress | Hybrid | 47-49 | 0.556 (0.427-0.670) | 0.527 (0.398-0.629) | 0.460 (0.329-0.580) | 0.508 (0.343-0.632) | 0.576 (0.392-0.717) |
| Positive affect | Synthetic | 48-49 | 0.486 (0.393-0.552) | 0.497 (0.416-0.562) | 0.540 (0.452-0.601) | 0.582 (0.486-0.648) | 0.597 (0.476-0.676) |
| Positive affect | Hybrid | 48-49 | 0.478 (0.391-0.546) | 0.487 (0.399-0.545) | 0.533 (0.448-0.598) | 0.582 (0.495-0.646) | 0.592 (0.483-0.664) |
| Depression | Synthetic | 48-49 | 0.462 (0.313-0.594) | 0.467 (0.308-0.609) | 0.498 (0.364-0.626) | 0.484 (0.324-0.617) | 0.514 (0.400-0.629) |
| Depression | Hybrid | 48-49 | 0.462 (0.306-0.592) | 0.470 (0.299-0.610) | 0.501 (0.345-0.608) | 0.486 (0.338-0.629) | 0.510 (0.380-0.615) |

**Supplementary Table S12.8a.** Between-participant correspondence of longitudinal deviation slopes. Values are Spearman correlations between observed and predicted slopes with participant-block bootstrap 95% confidence intervals.

**Panel A. Cluster 1**

| Channel | Training regime | Participants, n | h = 1d | h = 3d | h = 7d | h = 14d | h = 21d |
| --- | --- | --- | --- | --- | --- | --- | --- |
| Total platform use | Synthetic | 45-46 | 0.805 (0.572-0.958) | 0.459 (0.116-0.756) | 0.667 (0.472-0.795) | 0.398 (0.067-0.661) | 0.352 (0.078-0.623) |
| Total platform use | Hybrid | 45-46 | 0.797 (0.518-0.949) | 0.402 (0.054-0.687) | 0.384 (0.074-0.670) | 0.349 (0.005-0.616) | 0.303 (-0.018-0.573) |
| Night-time platform use | Synthetic | 45-46 | 0.853 (0.698-0.940) | 0.380 (0.073-0.689) | 0.674 (0.475-0.817) | 0.427 (0.048-0.687) | 0.225 (-0.104-0.542) |
| Night-time platform use | Hybrid | 45-46 | 0.872 (0.736-0.944) | 0.361 (0.046-0.659) | 0.485 (0.241-0.683) | 0.500 (0.225-0.729) | 0.312 (-0.012-0.606) |
| Sleep duration | Synthetic | 45-46 | 0.804 (0.637-0.910) | 0.491 (0.216-0.710) | 0.410 (0.077-0.661) | 0.336 (0.018-0.600) | 0.339 (-0.006-0.634) |
| Sleep duration | Hybrid | 45-46 | 0.842 (0.691-0.936) | 0.517 (0.261-0.727) | 0.465 (0.131-0.713) | 0.285 (-0.023-0.545) | 0.215 (-0.102-0.528) |
| Daily steps | Synthetic | 46 | 0.931 (0.836-0.971) | 0.545 (0.259-0.778) | 0.138 (-0.207-0.445) | 0.609 (0.295-0.848) | 0.309 (-0.016-0.632) |
| Daily steps | Hybrid | 46 | 0.936 (0.854-0.972) | 0.572 (0.291-0.773) | 0.069 (-0.262-0.375) | 0.523 (0.171-0.813) | 0.195 (-0.180-0.500) |
| Stress | Synthetic | 34-40 | 0.306 (-0.027-0.586) | 0.377 (0.018-0.668) | 0.429 (0.087-0.724) | 0.507 (0.095-0.789) | 0.414 (0.118-0.672) |
| Stress | Hybrid | 34-40 | 0.242 (-0.114-0.532) | 0.315 (-0.048-0.642) | 0.277 (-0.130-0.625) | 0.564 (0.193-0.812) | 0.326 (-0.012-0.612) |
| Positive affect | Synthetic | 39-41 | 0.286 (-0.018-0.537) | 0.150 (-0.145-0.442) | 0.399 (0.096-0.672) | 0.501 (0.178-0.745) | 0.219 (-0.182-0.507) |
| Positive affect | Hybrid | 39-41 | 0.233 (-0.057-0.490) | 0.150 (-0.205-0.447) | 0.342 (-0.052-0.613) | 0.343 (-0.012-0.693) | 0.084 (-0.292-0.417) |
| Depression | Synthetic | 34-39 | 0.236 (-0.183-0.560) | 0.308 (-0.069-0.578) | 0.282 (-0.102-0.574) | 0.407 (0.029-0.725) | 0.301 (-0.023-0.635) |
| Depression | Hybrid | 34-39 | 0.262 (-0.105-0.576) | 0.149 (-0.219-0.471) | 0.086 (-0.293-0.432) | 0.552 (0.249-0.772) | 0.375 (0.025-0.643) |

**Panel B. Cluster 2**

| Channel | Training regime | Participants, n | h = 1d | h = 3d | h = 7d | h = 14d | h = 21d |
| --- | --- | --- | --- | --- | --- | --- | --- |
| Total platform use | Synthetic | 46 | 0.924 (0.853-0.963) | 0.826 (0.682-0.912) | 0.746 (0.510-0.867) | 0.598 (0.370-0.749) | 0.367 (0.083-0.637) |
| Total platform use | Hybrid | 46 | 0.934 (0.850-0.968) | 0.809 (0.652-0.898) | 0.613 (0.335-0.797) | 0.420 (0.147-0.624) | 0.161 (-0.192-0.452) |
| Night-time platform use | Synthetic | 46 | 0.923 (0.839-0.961) | 0.654 (0.387-0.823) | 0.603 (0.320-0.779) | 0.480 (0.194-0.702) | 0.347 (0.048-0.610) |
| Night-time platform use | Hybrid | 46 | 0.920 (0.834-0.958) | 0.626 (0.356-0.824) | 0.554 (0.277-0.763) | 0.433 (0.136-0.676) | 0.337 (0.006-0.615) |
| Sleep duration | Synthetic | 46 | 0.752 (0.491-0.918) | 0.406 (0.115-0.642) | 0.417 (0.102-0.657) | 0.536 (0.309-0.725) | 0.385 (0.084-0.632) |
| Sleep duration | Hybrid | 46 | 0.880 (0.778-0.933) | 0.420 (0.128-0.654) | 0.488 (0.190-0.725) | 0.468 (0.231-0.675) | 0.288 (-0.055-0.585) |
| Daily steps | Synthetic | 46 | 0.885 (0.695-0.971) | 0.728 (0.486-0.864) | 0.594 (0.298-0.801) | 0.545 (0.230-0.774) | 0.397 (0.098-0.614) |
| Daily steps | Hybrid | 46 | 0.891 (0.708-0.975) | 0.746 (0.525-0.889) | 0.596 (0.307-0.800) | 0.532 (0.245-0.731) | 0.339 (0.062-0.602) |
| Stress | Synthetic | 42-43 | 0.418 (0.111-0.649) | 0.379 (0.025-0.609) | 0.256 (-0.083-0.528) | 0.485 (0.181-0.742) | 0.534 (0.227-0.754) |
| Stress | Hybrid | 42-43 | 0.433 (0.125-0.649) | 0.299 (-0.034-0.595) | 0.304 (0.004-0.557) | 0.525 (0.252-0.739) | 0.482 (0.149-0.705) |
| Positive affect | Synthetic | 46 | 0.313 (0.018-0.571) | 0.293 (-0.008-0.564) | 0.399 (0.098-0.614) | 0.502 (0.248-0.719) | 0.539 (0.304-0.713) |
| Positive affect | Hybrid | 46 | 0.288 (0.006-0.605) | 0.321 (0.045-0.557) | 0.357 (0.059-0.603) | 0.449 (0.149-0.673) | 0.522 (0.280-0.697) |
| Depression | Synthetic | 44-46 | 0.315 (0.045-0.546) | 0.291 (-0.038-0.587) | 0.265 (-0.035-0.541) | 0.462 (0.179-0.681) | 0.600 (0.313-0.774) |
| Depression | Hybrid | 44-46 | 0.279 (-0.023-0.553) | 0.276 (-0.052-0.571) | 0.245 (-0.063-0.552) | 0.415 (0.099-0.657) | 0.527 (0.230-0.736) |

**Panel C. Cluster 3**

| Channel | Training regime | Participants, n | h = 1d | h = 3d | h = 7d | h = 14d | h = 21d |
| --- | --- | --- | --- | --- | --- | --- | --- |
| Total platform use | Synthetic | 48 | 0.726 (0.493-0.883) | 0.576 (0.325-0.763) | 0.515 (0.278-0.688) | 0.221 (-0.070-0.502) | 0.322 (0.009-0.596) |
| Total platform use | Hybrid | 48 | 0.736 (0.496-0.891) | 0.576 (0.315-0.782) | 0.502 (0.263-0.710) | 0.186 (-0.078-0.458) | 0.331 (0.046-0.592) |
| Night-time platform use | Synthetic | 48 | 0.900 (0.821-0.949) | 0.480 (0.187-0.759) | 0.395 (0.105-0.636) | 0.336 (0.058-0.591) | 0.338 (0.063-0.589) |
| Night-time platform use | Hybrid | 48 | 0.900 (0.827-0.939) | 0.416 (0.124-0.666) | 0.237 (-0.069-0.524) | 0.263 (-0.041-0.548) | 0.308 (0.028-0.569) |
| Sleep duration | Synthetic | 48 | 0.847 (0.681-0.945) | 0.525 (0.245-0.747) | 0.702 (0.469-0.849) | 0.571 (0.299-0.778) | 0.429 (0.150-0.665) |
| Sleep duration | Hybrid | 48 | 0.903 (0.811-0.956) | 0.556 (0.319-0.763) | 0.702 (0.472-0.857) | 0.523 (0.203-0.738) | 0.382 (0.074-0.644) |
| Daily steps | Synthetic | 48 | 0.928 (0.821-0.979) | 0.723 (0.536-0.856) | 0.420 (0.099-0.673) | 0.417 (0.112-0.649) | 0.379 (0.073-0.613) |
| Daily steps | Hybrid | 48 | 0.923 (0.805-0.979) | 0.698 (0.463-0.839) | 0.403 (0.087-0.683) | 0.385 (0.093-0.644) | 0.321 (0.040-0.563) |
| Stress | Synthetic | 38-46 | 0.379 (0.123-0.631) | 0.437 (0.175-0.639) | 0.581 (0.366-0.761) | 0.483 (0.204-0.714) | 0.172 (-0.182-0.449) |
| Stress | Hybrid | 38-46 | 0.339 (0.046-0.593) | 0.277 (0.026-0.512) | 0.443 (0.160-0.669) | 0.409 (0.129-0.641) | 0.088 (-0.260-0.361) |
| Positive affect | Synthetic | 46-48 | 0.229 (-0.053-0.517) | 0.208 (-0.085-0.494) | 0.357 (0.054-0.604) | 0.466 (0.196-0.664) | 0.442 (0.150-0.641) |
| Positive affect | Hybrid | 46-48 | 0.226 (-0.081-0.488) | 0.168 (-0.114-0.418) | 0.303 (0.026-0.568) | 0.330 (0.005-0.593) | 0.391 (0.120-0.618) |
| Depression | Synthetic | 40-45 | 0.513 (0.232-0.719) | 0.432 (0.090-0.694) | 0.567 (0.254-0.788) | 0.478 (0.105-0.763) | 0.245 (-0.100-0.540) |
| Depression | Hybrid | 40-45 | 0.422 (0.086-0.682) | 0.327 (-0.010-0.600) | 0.545 (0.282-0.751) | 0.417 (0.001-0.723) | 0.302 (-0.069-0.566) |

**Panel D. Cluster 4**

| Channel | Training regime | Participants, n | h = 1d | h = 3d | h = 7d | h = 14d | h = 21d |
| --- | --- | --- | --- | --- | --- | --- | --- |
| Total platform use | Synthetic | 49 | 0.877 (0.769-0.923) | 0.662 (0.435-0.819) | 0.656 (0.422-0.794) | 0.552 (0.337-0.697) | 0.099 (-0.166-0.379) |
| Total platform use | Hybrid | 49 | 0.888 (0.780-0.932) | 0.624 (0.367-0.803) | 0.478 (0.227-0.664) | 0.377 (0.145-0.573) | 0.072 (-0.181-0.330) |
| Night-time platform use | Synthetic | 49 | 0.912 (0.804-0.957) | 0.703 (0.526-0.835) | 0.580 (0.345-0.776) | 0.269 (-0.041-0.556) | 0.126 (-0.164-0.393) |
| Night-time platform use | Hybrid | 49 | 0.904 (0.800-0.958) | 0.647 (0.421-0.830) | 0.557 (0.261-0.733) | 0.298 (-0.024-0.567) | 0.138 (-0.151-0.373) |
| Sleep duration | Synthetic | 48-49 | 0.766 (0.594-0.872) | 0.264 (-0.045-0.515) | 0.380 (0.058-0.593) | 0.573 (0.372-0.727) | 0.379 (0.103-0.607) |
| Sleep duration | Hybrid | 48-49 | 0.852 (0.738-0.915) | 0.370 (0.069-0.616) | 0.424 (0.141-0.645) | 0.474 (0.226-0.675) | 0.259 (-0.049-0.510) |
| Daily steps | Synthetic | 48-49 | 0.978 (0.943-0.990) | 0.790 (0.601-0.888) | 0.488 (0.236-0.690) | 0.428 (0.139-0.666) | 0.397 (0.097-0.626) |
| Daily steps | Hybrid | 48-49 | 0.975 (0.934-0.988) | 0.789 (0.616-0.896) | 0.495 (0.227-0.716) | 0.347 (0.023-0.586) | 0.310 (0.009-0.574) |
| Stress | Synthetic | 39-45 | 0.534 (0.305-0.709) | 0.351 (0.097-0.584) | 0.224 (-0.150-0.548) | 0.257 (-0.055-0.554) | 0.244 (-0.124-0.532) |
| Stress | Hybrid | 39-45 | 0.405 (0.100-0.662) | 0.215 (-0.099-0.510) | 0.179 (-0.148-0.507) | 0.214 (-0.084-0.475) | 0.092 (-0.247-0.400) |
| Positive affect | Synthetic | 47-49 | 0.288 (0.000-0.554) | 0.113 (-0.196-0.362) | 0.324 (0.016-0.565) | 0.399 (0.102-0.643) | 0.219 (-0.103-0.488) |
| Positive affect | Hybrid | 47-49 | 0.258 (-0.022-0.497) | 0.148 (-0.152-0.381) | 0.334 (0.069-0.559) | 0.410 (0.084-0.654) | 0.191 (-0.114-0.474) |
| Depression | Synthetic | 45-47 | 0.273 (-0.016-0.544) | 0.455 (0.146-0.661) | 0.507 (0.213-0.726) | 0.581 (0.293-0.790) | 0.471 (0.226-0.662) |
| Depression | Hybrid | 45-47 | 0.198 (-0.096-0.487) | 0.410 (0.102-0.649) | 0.516 (0.246-0.739) | 0.582 (0.294-0.796) | 0.341 (0.035-0.593) |

**Supplementary Table S12.8b.** Agreement in participant-level longitudinal slope direction. Values are percentages with participant-block bootstrapped 95%-CIs.

**Panel A. Cluster 1**

| Channel | Training regime | Participants, n | h = 1d | h = 3d | h = 7d | h = 14d | h = 21d |
| --- | --- | --- | --- | --- | --- | --- | --- |
| Total platform use | Synthetic | 45-46 | 87.0% (78.3-95.7%) | 73.9% (60.9-84.8%) | 69.6% (56.5-82.6%) | 67.4% (52.2-80.4%) | 66.7% (53.3-80.0%) |
| Total platform use | Hybrid | 45-46 | 89.1% (80.4-97.8%) | 69.6% (56.5-82.6%) | 63.0% (50.0-76.1%) | 65.2% (52.2-78.3%) | 64.4% (48.9-77.8%) |
| Night-time platform use | Synthetic | 45-46 | 82.6% (71.7-93.5%) | 67.4% (54.3-80.4%) | 76.1% (63.0-89.1%) | 71.7% (58.7-84.8%) | 62.2% (46.7-77.8%) |
| Night-time platform use | Hybrid | 45-46 | 82.6% (71.7-93.5%) | 71.7% (58.7-84.8%) | 65.2% (52.2-78.3%) | 71.7% (58.7-82.6%) | 64.4% (48.9-80.0%) |
| Sleep duration | Synthetic | 45-46 | 78.3% (67.4-89.1%) | 67.4% (54.3-80.4%) | 67.4% (54.3-80.4%) | 63.0% (50.0-76.1%) | 62.2% (48.9-77.8%) |
| Sleep duration | Hybrid | 45-46 | 80.4% (67.4-91.3%) | 73.9% (60.9-87.0%) | 71.7% (56.5-84.8%) | 63.0% (50.0-73.9%) | 62.2% (48.9-77.8%) |
| Daily steps | Synthetic | 46 | 84.8% (71.7-93.5%) | 69.6% (56.5-82.6%) | 54.3% (41.3-69.6%) | 76.1% (63.0-87.0%) | 65.2% (52.2-78.3%) |
| Daily steps | Hybrid | 46 | 89.1% (80.4-97.8%) | 71.7% (56.5-82.6%) | 50.0% (34.8-65.2%) | 71.7% (58.7-84.8%) | 58.7% (43.5-71.7%) |
| Stress | Synthetic | 34-40 | 42.5% (25.0-57.5%) | 55.6% (38.9-72.2%) | 52.6% (36.8-68.4%) | 52.8% (38.9-69.4%) | 50.0% (32.4-64.7%) |
| Stress | Hybrid | 34-40 | 45.0% (30.0-60.0%) | 52.8% (36.1-69.4%) | 47.4% (31.6-63.2%) | 58.3% (41.7-75.0%) | 55.9% (38.2-70.6%) |
| Positive affect | Synthetic | 39-41 | 48.8% (34.1-63.4%) | 61.0% (43.9-75.6%) | 75.6% (61.0-87.8%) | 68.3% (53.7-82.9%) | 38.5% (23.1-53.8%) |
| Positive affect | Hybrid | 39-41 | 48.8% (34.1-63.4%) | 58.5% (43.9-73.2%) | 56.1% (41.5-73.2%) | 53.7% (39.0-68.3%) | 41.0% (25.6-56.4%) |
| Depression | Synthetic | 34-39 | 35.1% (21.6-51.4%) | 48.7% (33.3-64.1%) | 38.9% (22.2-55.6%) | 44.1% (29.4-58.8%) | 45.9% (32.4-59.5%) |
| Depression | Hybrid | 34-39 | 37.8% (21.6-54.1%) | 35.9% (20.5-51.3%) | 30.6% (16.7-44.4%) | 44.1% (26.5-61.8%) | 54.1% (37.8-70.3%) |

**Panel B. Cluster 2**

| Channel | Training regime | Participants, n | h = 1d | h = 3d | h = 7d | h = 14d | h = 21d |
| --- | --- | --- | --- | --- | --- | --- | --- |
| Total platform use | Synthetic | 46 | 91.3% (82.6-97.8%) | 80.4% (67.4-91.3%) | 82.6% (71.7-93.5%) | 60.9% (45.7-76.1%) | 58.7% (43.5-73.9%) |
| Total platform use | Hybrid | 46 | 89.1% (80.4-95.7%) | 84.8% (76.1-93.5%) | 73.9% (60.9-87.0%) | 58.7% (45.7-73.9%) | 56.5% (41.3-69.6%) |
| Night-time platform use | Synthetic | 46 | 87.0% (76.1-95.7%) | 76.1% (65.2-91.3%) | 69.6% (56.5-82.6%) | 65.2% (52.2-78.3%) | 71.7% (58.7-82.6%) |
| Night-time platform use | Hybrid | 46 | 87.0% (76.1-95.7%) | 71.7% (58.7-84.8%) | 69.6% (56.5-80.4%) | 58.7% (43.5-73.9%) | 63.0% (50.0-78.3%) |
| Sleep duration | Synthetic | 46 | 84.8% (73.9-93.5%) | 69.6% (56.5-84.8%) | 67.4% (54.3-80.4%) | 63.0% (50.0-76.1%) | 58.7% (45.7-73.9%) |
| Sleep duration | Hybrid | 46 | 84.8% (73.9-93.5%) | 73.9% (60.9-87.0%) | 71.7% (58.7-82.6%) | 69.6% (56.5-82.6%) | 58.7% (45.7-71.7%) |
| Daily steps | Synthetic | 46 | 91.3% (82.6-97.8%) | 78.3% (67.4-89.1%) | 73.9% (60.9-84.8%) | 65.2% (52.2-78.3%) | 54.3% (39.1-69.6%) |
| Daily steps | Hybrid | 46 | 91.3% (82.6-97.8%) | 76.1% (63.0-87.0%) | 73.9% (60.9-84.8%) | 67.4% (54.3-80.4%) | 56.5% (43.5-71.7%) |
| Stress | Synthetic | 42-43 | 51.2% (34.9-67.4%) | 53.5% (37.2-69.8%) | 53.5% (39.5-67.4%) | 62.8% (48.8-76.7%) | 57.1% (42.9-71.4%) |
| Stress | Hybrid | 42-43 | 53.5% (37.2-69.8%) | 48.8% (32.6-62.8%) | 55.8% (39.5-72.1%) | 72.1% (58.1-83.7%) | 57.1% (42.9-71.4%) |
| Positive affect | Synthetic | 46 | 73.9% (60.9-87.0%) | 58.7% (43.5-71.7%) | 65.2% (50.0-78.3%) | 71.7% (58.7-82.6%) | 73.9% (60.9-87.0%) |
| Positive affect | Hybrid | 46 | 73.9% (60.9-87.0%) | 63.0% (50.0-78.3%) | 65.2% (50.0-78.3%) | 69.6% (56.5-82.6%) | 73.9% (60.9-87.0%) |
| Depression | Synthetic | 44-46 | 61.4% (47.7-75.0%) | 56.8% (40.9-70.5%) | 53.3% (37.8-68.9%) | 64.4% (51.1-77.8%) | 69.6% (56.5-82.6%) |
| Depression | Hybrid | 44-46 | 61.4% (45.5-75.0%) | 59.1% (45.5-72.7%) | 53.3% (37.8-66.7%) | 62.2% (46.7-75.6%) | 67.4% (54.3-80.4%) |

**Panel C. Cluster 3**

| Channel | Training regime | Participants, n | h = 1d | h = 3d | h = 7d | h = 14d | h = 21d |
| --- | --- | --- | --- | --- | --- | --- | --- |
| Total platform use | Synthetic | 48 | 83.3% (72.9-93.8%) | 77.1% (64.6-87.5%) | 72.9% (58.3-83.3%) | 66.7% (52.1-77.1%) | 58.3% (43.8-72.9%) |
| Total platform use | Hybrid | 48 | 81.2% (68.8-91.7%) | 68.8% (54.2-81.2%) | 68.8% (56.2-81.2%) | 64.6% (52.1-77.1%) | 58.3% (45.8-72.9%) |
| Night-time platform use | Synthetic | 48 | 89.6% (81.2-97.9%) | 72.9% (60.4-85.4%) | 56.2% (41.7-68.8%) | 66.7% (54.2-79.2%) | 64.6% (52.1-77.1%) |
| Night-time platform use | Hybrid | 48 | 83.3% (70.8-93.8%) | 58.3% (43.8-70.8%) | 47.9% (33.3-62.5%) | 56.2% (41.7-70.8%) | 62.5% (50.0-75.0%) |
| Sleep duration | Synthetic | 48 | 87.5% (77.1-95.8%) | 72.9% (60.4-85.4%) | 72.9% (60.4-85.4%) | 77.1% (64.6-89.6%) | 62.5% (47.9-75.0%) |
| Sleep duration | Hybrid | 48 | 91.7% (83.3-97.9%) | 75.0% (60.4-85.4%) | 72.9% (60.4-85.4%) | 75.0% (62.5-87.5%) | 64.6% (50.0-77.1%) |
| Daily steps | Synthetic | 48 | 87.5% (77.1-95.8%) | 68.8% (54.2-81.2%) | 56.2% (43.8-70.8%) | 58.3% (43.8-70.8%) | 64.6% (50.0-77.1%) |
| Daily steps | Hybrid | 48 | 87.5% (77.1-95.8%) | 70.8% (58.3-83.3%) | 58.3% (43.8-70.8%) | 58.3% (43.8-72.9%) | 58.3% (43.8-72.9%) |
| Stress | Synthetic | 38-46 | 55.6% (42.2-68.9%) | 58.7% (43.5-71.7%) | 71.1% (57.8-84.4%) | 62.2% (46.7-75.6%) | 55.3% (42.1-71.1%) |
| Stress | Hybrid | 38-46 | 55.6% (40.0-68.9%) | 54.3% (39.1-67.4%) | 62.2% (48.9-75.6%) | 51.1% (37.8-66.7%) | 42.1% (26.3-57.9%) |
| Positive affect | Synthetic | 46-48 | 58.3% (43.8-70.8%) | 54.2% (39.6-68.8%) | 60.4% (45.8-72.9%) | 56.2% (39.6-68.8%) | 58.7% (43.5-71.7%) |
| Positive affect | Hybrid | 46-48 | 56.2% (43.8-68.8%) | 47.9% (35.4-60.4%) | 54.2% (37.5-68.8%) | 47.9% (33.3-62.5%) | 60.9% (45.7-76.1%) |
| Depression | Synthetic | 40-45 | 46.5% (32.6-60.5%) | 39.5% (25.6-55.8%) | 40.9% (27.3-56.8%) | 40.0% (25.0-55.0%) | 42.2% (28.9-57.8%) |
| Depression | Hybrid | 40-45 | 39.5% (25.6-53.5%) | 34.9% (20.9-48.8%) | 40.9% (27.3-54.5%) | 35.0% (20.0-50.0%) | 46.7% (31.1-62.2%) |

**Panel D. Cluster 4**

| Channel | Training regime | Participants, n | h = 1d | h = 3d | h = 7d | h = 14d | h = 21d |
| --- | --- | --- | --- | --- | --- | --- | --- |
| Total platform use | Synthetic | 49 | 87.8% (79.6-95.9%) | 73.5% (61.2-85.7%) | 65.3% (53.1-77.6%) | 73.5% (59.2-85.7%) | 55.1% (40.8-67.3%) |
| Total platform use | Hybrid | 49 | 85.7% (75.5-93.9%) | 69.4% (55.1-81.6%) | 69.4% (55.1-81.6%) | 65.3% (51.0-77.6%) | 55.1% (40.8-69.4%) |
| Night-time platform use | Synthetic | 49 | 89.8% (81.6-98.0%) | 71.4% (57.1-83.7%) | 69.4% (57.1-83.7%) | 65.3% (53.1-77.6%) | 63.3% (49.0-77.6%) |
| Night-time platform use | Hybrid | 49 | 87.8% (77.6-95.9%) | 79.6% (69.4-89.8%) | 69.4% (55.1-81.6%) | 65.3% (51.0-77.6%) | 59.2% (44.9-73.5%) |
| Sleep duration | Synthetic | 48-49 | 75.5% (63.3-87.8%) | 59.2% (44.9-73.5%) | 71.4% (57.1-83.7%) | 75.5% (63.3-87.8%) | 66.7% (54.2-79.2%) |
| Sleep duration | Hybrid | 48-49 | 85.7% (77.6-93.9%) | 61.2% (46.9-75.5%) | 71.4% (59.2-83.7%) | 73.5% (61.2-85.7%) | 58.3% (43.8-72.9%) |
| Daily steps | Synthetic | 48-49 | 89.8% (79.6-98.0%) | 73.5% (61.2-85.7%) | 63.3% (49.0-77.6%) | 59.2% (44.9-73.5%) | 64.6% (52.1-77.1%) |
| Daily steps | Hybrid | 48-49 | 89.8% (79.6-98.0%) | 71.4% (59.2-83.7%) | 67.3% (55.1-79.6%) | 53.1% (36.7-67.3%) | 60.4% (45.8-72.9%) |
| Stress | Synthetic | 39-45 | 62.2% (46.7-75.6%) | 55.6% (42.2-68.9%) | 48.8% (34.9-65.1%) | 51.2% (37.2-67.4%) | 53.8% (38.5-71.8%) |
| Stress | Hybrid | 39-45 | 55.6% (42.2-68.9%) | 44.4% (31.1-57.8%) | 44.2% (30.2-60.5%) | 53.5% (37.2-67.4%) | 46.2% (30.8-61.5%) |
| Positive affect | Synthetic | 47-49 | 63.3% (49.0-75.5%) | 46.9% (34.7-61.2%) | 61.2% (46.9-75.5%) | 54.2% (39.6-66.7%) | 53.2% (40.4-66.0%) |
| Positive affect | Hybrid | 47-49 | 57.1% (42.9-71.4%) | 51.0% (38.8-65.3%) | 61.2% (49.0-73.5%) | 56.2% (41.7-68.8%) | 55.3% (38.3-68.1%) |
| Depression | Synthetic | 45-47 | 55.6% (40.0-68.9%) | 62.2% (46.7-77.8%) | 60.9% (47.8-73.9%) | 57.8% (42.2-71.1%) | 68.1% (55.3-80.9%) |
| Depression | Hybrid | 45-47 | 51.1% (37.8-66.7%) | 66.7% (51.1-80.0%) | 63.0% (47.8-78.3%) | 60.0% (46.7-75.6%) | 55.3% (40.4-68.1%) |

**Supplementary Table S12.11a.** Channel-specific complete-case repeated-measures analyses of participant-level within-subject skill relative to the causal prefix-mean baseline. The 3 x 2 x 5 models included three decoders, two training regimes and five forecast horizons. For every channel, participant-anchor support was matched across decoders, regimes, the prefix-mean baseline and persistence separately within each horizon; complete participants across the full factorial design were then retained. Null models defined the common evaluation support but were not included as factorial levels. Holm-adjusted P values and partial eta-squared are reported.

**Panel A. Cluster 1**

| Channel | Effect | Participants, n | F (df1, df2) | Holm p | Partial eta2 |
| --- | --- | --- | --- | --- | --- |
| Total platform use | Decoder | 45 | 8.83 (2,88) | 0.051 | 0.167 |
| Total platform use | Training regime | 45 | 0.01 (1,44) | 1.000 | 0.000 |
| Total platform use | Forecast horizon | 45 | 5.31 (4,176) | 0.072 | 0.108 |
| Total platform use | Decoder x training regime | 45 | 3.02 (2,88) | 1.000 | 0.064 |
| Total platform use | Decoder x forecast horizon | 45 | 1.16 (8,352) | 1.000 | 0.026 |
| Total platform use | Training regime x forecast horizon | 45 | 9.63 (4,176) | **<0.001** | 0.180 |
| Total platform use | Decoder x training regime x forecast horizon | 45 | 2.75 (8,352) | 0.755 | 0.059 |
| Night-time platform use | Decoder | 45 | 3.78 (2,88) | 1.000 | 0.079 |
| Night-time platform use | Training regime | 45 | 0.72 (1,44) | 1.000 | 0.016 |
| Night-time platform use | Forecast horizon | 45 | 0.74 (4,176) | 1.000 | 0.017 |
| Night-time platform use | Decoder x training regime | 45 | 2.90 (2,88) | 1.000 | 0.062 |
| Night-time platform use | Decoder x forecast horizon | 45 | 8.45 (8,352) | **<0.001** | 0.161 |
| Night-time platform use | Training regime x forecast horizon | 45 | 9.43 (4,176) | **<0.001** | 0.177 |
| Night-time platform use | Decoder x training regime x forecast horizon | 45 | 9.10 (8,352) | **<0.001** | 0.171 |
| Sleep duration | Decoder | 46 | 0.34 (2,90) | 1.000 | 0.007 |
| Sleep duration | Training regime | 46 | 9.24 (1,45) | 0.536 | 0.170 |
| Sleep duration | Forecast horizon | 46 | 2.32 (4,180) | 1.000 | 0.049 |
| Sleep duration | Decoder x training regime | 46 | 7.05 (2,90) | 0.212 | 0.135 |
| Sleep duration | Decoder x forecast horizon | 46 | 7.76 (8,360) | **<0.001** | 0.147 |
| Sleep duration | Training regime x forecast horizon | 46 | 1.13 (4,180) | 1.000 | 0.025 |
| Sleep duration | Decoder x training regime x forecast horizon | 46 | 2.05 (8,360) | 1.000 | 0.044 |
| Daily steps | Decoder | 46 | 8.12 (2,90) | 0.088 | 0.153 |
| Daily steps | Training regime | 46 | 2.90 (1,45) | 1.000 | 0.060 |
| Daily steps | Forecast horizon | 46 | 3.65 (4,180) | 0.881 | 0.075 |
| Daily steps | Decoder x training regime | 46 | 2.63 (2,90) | 1.000 | 0.055 |
| Daily steps | Decoder x forecast horizon | 46 | 8.13 (8,360) | **<0.001** | 0.153 |
| Daily steps | Training regime x forecast horizon | 46 | 1.78 (4,180) | 1.000 | 0.038 |
| Daily steps | Decoder x training regime x forecast horizon | 46 | 1.93 (8,360) | 1.000 | 0.041 |
| Stress | Decoder | 39 | 0.97 (2,76) | 1.000 | 0.025 |
| Stress | Training regime | 39 | 0.32 (1,38) | 1.000 | 0.008 |
| Stress | Forecast horizon | 39 | 0.47 (4,152) | 1.000 | 0.012 |
| Stress | Decoder x training regime | 39 | 0.30 (2,76) | 1.000 | 0.008 |
| Stress | Decoder x forecast horizon | 39 | 4.30 (8,304) | **0.010** | 0.102 |
| Stress | Training regime x forecast horizon | 39 | 1.16 (4,152) | 1.000 | 0.030 |
| Stress | Decoder x training regime x forecast horizon | 39 | 1.09 (8,304) | 1.000 | 0.028 |
| Positive affect | Decoder | 41 | 4.76 (2,80) | 1.000 | 0.106 |
| Positive affect | Training regime | 41 | 5.36 (1,40) | 1.000 | 0.118 |
| Positive affect | Forecast horizon | 41 | 1.42 (4,160) | 1.000 | 0.034 |
| Positive affect | Decoder x training regime | 41 | 0.37 (2,80) | 1.000 | 0.009 |
| Positive affect | Decoder x forecast horizon | 41 | 1.35 (8,320) | 1.000 | 0.033 |
| Positive affect | Training regime x forecast horizon | 41 | 1.78 (4,160) | 1.000 | 0.043 |
| Positive affect | Decoder x training regime x forecast horizon | 41 | 0.39 (8,320) | 1.000 | 0.010 |
| Depression | Decoder | 32 | 0.24 (2,62) | 1.000 | 0.008 |
| Depression | Training regime | 32 | 1.09 (1,31) | 1.000 | 0.034 |
| Depression | Forecast horizon | 32 | 1.23 (4,124) | 1.000 | 0.038 |
| Depression | Decoder x training regime | 32 | 0.18 (2,62) | 1.000 | 0.006 |
| Depression | Decoder x forecast horizon | 32 | 0.90 (8,248) | 1.000 | 0.028 |
| Depression | Training regime x forecast horizon | 32 | 0.99 (4,124) | 1.000 | 0.031 |
| Depression | Decoder x training regime x forecast horizon | 32 | 0.83 (8,248) | 1.000 | 0.026 |

**Panel B. Cluster 2**

| Channel | Effect | Participants, n | F (df1, df2) | Holm p | Partial eta2 |
| --- | --- | --- | --- | --- | --- |
| Total platform use | Decoder | 47 | 6.48 (2,92) | 0.331 | 0.123 |
| Total platform use | Training regime | 47 | 0.93 (1,46) | 1.000 | 0.020 |
| Total platform use | Forecast horizon | 47 | 0.58 (4,184) | 1.000 | 0.012 |
| Total platform use | Decoder x training regime | 47 | 4.19 (2,92) | 1.000 | 0.084 |
| Total platform use | Decoder x forecast horizon | 47 | 3.36 (8,368) | 0.147 | 0.068 |
| Total platform use | Training regime x forecast horizon | 47 | 7.17 (4,184) | **0.004** | 0.135 |
| Total platform use | Decoder x training regime x forecast horizon | 47 | 5.11 (8,368) | **<0.001** | 0.100 |
| Night-time platform use | Decoder | 47 | 7.34 (2,92) | 0.164 | 0.138 |
| Night-time platform use | Training regime | 47 | 4.08 (1,46) | 1.000 | 0.082 |
| Night-time platform use | Forecast horizon | 47 | 1.53 (4,184) | 1.000 | 0.032 |
| Night-time platform use | Decoder x training regime | 47 | 5.62 (2,92) | 0.659 | 0.109 |
| Night-time platform use | Decoder x forecast horizon | 47 | 11.16 (8,368) | **<0.001** | 0.195 |
| Night-time platform use | Training regime x forecast horizon | 47 | 8.82 (4,184) | **<0.001** | 0.161 |
| Night-time platform use | Decoder x training regime x forecast horizon | 47 | 6.77 (8,368) | **<0.001** | 0.128 |
| Sleep duration | Decoder | 47 | 0.69 (2,92) | 1.000 | 0.015 |
| Sleep duration | Training regime | 47 | 3.76 (1,46) | 1.000 | 0.076 |
| Sleep duration | Forecast horizon | 47 | 1.62 (4,184) | 1.000 | 0.034 |
| Sleep duration | Decoder x training regime | 47 | 2.51 (2,92) | 1.000 | 0.052 |
| Sleep duration | Decoder x forecast horizon | 47 | 5.88 (8,368) | **<0.001** | 0.113 |
| Sleep duration | Training regime x forecast horizon | 47 | 4.20 (4,184) | 0.392 | 0.084 |
| Sleep duration | Decoder x training regime x forecast horizon | 47 | 3.16 (8,368) | 0.258 | 0.064 |
| Daily steps | Decoder | 47 | 18.29 (2,92) | **<0.001** | 0.284 |
| Daily steps | Training regime | 47 | 8.26 (1,46) | 0.781 | 0.152 |
| Daily steps | Forecast horizon | 47 | 3.94 (4,184) | 0.578 | 0.079 |
| Daily steps | Decoder x training regime | 47 | 8.93 (2,92) | **0.046** | 0.163 |
| Daily steps | Decoder x forecast horizon | 47 | 25.41 (8,368) | **<0.001** | 0.356 |
| Daily steps | Training regime x forecast horizon | 47 | 11.61 (4,184) | **<0.001** | 0.202 |
| Daily steps | Decoder x training regime x forecast horizon | 47 | 10.83 (8,368) | **<0.001** | 0.191 |
| Stress | Decoder | 46 | 0.52 (2,90) | 1.000 | 0.011 |
| Stress | Training regime | 46 | 11.05 (1,45) | 0.258 | 0.197 |
| Stress | Forecast horizon | 46 | 0.61 (4,180) | 1.000 | 0.013 |
| Stress | Decoder x training regime | 46 | 4.72 (2,90) | 1.000 | 0.095 |
| Stress | Decoder x forecast horizon | 46 | 0.71 (8,360) | 1.000 | 0.016 |
| Stress | Training regime x forecast horizon | 46 | 0.56 (4,180) | 1.000 | 0.012 |
| Stress | Decoder x training regime x forecast horizon | 46 | 1.66 (8,360) | 1.000 | 0.036 |
| Positive affect | Decoder | 46 | 1.44 (2,90) | 1.000 | 0.031 |
| Positive affect | Training regime | 46 | 0.00 (1,45) | 1.000 | 0.000 |
| Positive affect | Forecast horizon | 46 | 0.28 (4,180) | 1.000 | 0.006 |
| Positive affect | Decoder x training regime | 46 | 0.92 (2,90) | 1.000 | 0.020 |
| Positive affect | Decoder x forecast horizon | 46 | 0.84 (8,360) | 1.000 | 0.018 |
| Positive affect | Training regime x forecast horizon | 46 | 0.27 (4,180) | 1.000 | 0.006 |
| Positive affect | Decoder x training regime x forecast horizon | 46 | 0.35 (8,360) | 1.000 | 0.008 |
| Depression | Decoder | 46 | 0.11 (2,90) | 1.000 | 0.002 |
| Depression | Training regime | 46 | 0.20 (1,45) | 1.000 | 0.004 |
| Depression | Forecast horizon | 46 | 1.63 (4,180) | 1.000 | 0.035 |
| Depression | Decoder x training regime | 46 | 0.14 (2,90) | 1.000 | 0.003 |
| Depression | Decoder x forecast horizon | 46 | 3.38 (8,360) | 0.141 | 0.070 |
| Depression | Training regime x forecast horizon | 46 | 0.35 (4,180) | 1.000 | 0.008 |
| Depression | Decoder x training regime x forecast horizon | 46 | 1.10 (8,360) | 1.000 | 0.024 |

**Panel C. Cluster 3**

| Channel | Effect | Participants, n | F (df1, df2) | Holm p | Partial eta2 |
| --- | --- | --- | --- | --- | --- |
| Total platform use | Decoder | 44 | 17.55 (2,86) | **<0.001** | 0.290 |
| Total platform use | Training regime | 44 | 5.07 (1,43) | 1.000 | 0.105 |
| Total platform use | Forecast horizon | 44 | 5.49 (4,172) | 0.054 | 0.113 |
| Total platform use | Decoder x training regime | 44 | 6.68 (2,86) | 0.289 | 0.134 |
| Total platform use | Decoder x forecast horizon | 44 | 2.11 (8,344) | 1.000 | 0.047 |
| Total platform use | Training regime x forecast horizon | 44 | 3.80 (4,172) | 0.712 | 0.081 |
| Total platform use | Decoder x training regime x forecast horizon | 44 | 0.55 (8,344) | 1.000 | 0.013 |
| Night-time platform use | Decoder | 44 | 5.77 (2,86) | 0.594 | 0.118 |
| Night-time platform use | Training regime | 44 | 10.47 (1,43) | 0.331 | 0.196 |
| Night-time platform use | Forecast horizon | 44 | 2.47 (4,172) | 1.000 | 0.054 |
| Night-time platform use | Decoder x training regime | 44 | 13.56 (2,86) | **0.001** | 0.240 |
| Night-time platform use | Decoder x forecast horizon | 44 | 4.73 (8,344) | **0.003** | 0.099 |
| Night-time platform use | Training regime x forecast horizon | 44 | 2.54 (4,172) | 1.000 | 0.056 |
| Night-time platform use | Decoder x training regime x forecast horizon | 44 | 1.05 (8,344) | 1.000 | 0.024 |
| Sleep duration | Decoder | 44 | 5.39 (2,86) | 0.794 | 0.111 |
| Sleep duration | Training regime | 44 | 16.20 (1,43) | **0.036** | 0.274 |
| Sleep duration | Forecast horizon | 44 | 1.17 (4,172) | 1.000 | 0.026 |
| Sleep duration | Decoder x training regime | 44 | 13.63 (2,86) | **0.001** | 0.241 |
| Sleep duration | Decoder x forecast horizon | 44 | 4.18 (8,344) | **0.014** | 0.089 |
| Sleep duration | Training regime x forecast horizon | 44 | 1.28 (4,172) | 1.000 | 0.029 |
| Sleep duration | Decoder x training regime x forecast horizon | 44 | 0.76 (8,344) | 1.000 | 0.017 |
| Daily steps | Decoder | 44 | 57.84 (2,86) | **<0.001** | 0.574 |
| Daily steps | Training regime | 44 | 1.60 (1,43) | 1.000 | 0.036 |
| Daily steps | Forecast horizon | 44 | 12.00 (4,172) | **<0.001** | 0.218 |
| Daily steps | Decoder x training regime | 44 | 0.86 (2,86) | 1.000 | 0.020 |
| Daily steps | Decoder x forecast horizon | 44 | 70.21 (8,344) | **<0.001** | 0.620 |
| Daily steps | Training regime x forecast horizon | 44 | 11.47 (4,172) | **<0.001** | 0.211 |
| Daily steps | Decoder x training regime x forecast horizon | 44 | 13.43 (8,344) | **<0.001** | 0.238 |
| Stress | Decoder | 41 | 4.73 (2,80) | 1.000 | 0.106 |
| Stress | Training regime | 41 | 0.98 (1,40) | 1.000 | 0.024 |
| Stress | Forecast horizon | 41 | 0.83 (4,160) | 1.000 | 0.020 |
| Stress | Decoder x training regime | 41 | 3.57 (2,80) | 1.000 | 0.082 |
| Stress | Decoder x forecast horizon | 41 | 0.35 (8,320) | 1.000 | 0.009 |
| Stress | Training regime x forecast horizon | 41 | 0.51 (4,160) | 1.000 | 0.013 |
| Stress | Decoder x training regime x forecast horizon | 41 | 1.24 (8,320) | 1.000 | 0.030 |
| Positive affect | Decoder | 45 | 7.77 (2,88) | 0.119 | 0.150 |
| Positive affect | Training regime | 45 | 6.97 (1,44) | 1.000 | 0.137 |
| Positive affect | Forecast horizon | 45 | 0.58 (4,176) | 1.000 | 0.013 |
| Positive affect | Decoder x training regime | 45 | 2.52 (2,88) | 1.000 | 0.054 |
| Positive affect | Decoder x forecast horizon | 45 | 0.52 (8,352) | 1.000 | 0.012 |
| Positive affect | Training regime x forecast horizon | 45 | 0.63 (4,176) | 1.000 | 0.014 |
| Positive affect | Decoder x training regime x forecast horizon | 45 | 0.98 (8,352) | 1.000 | 0.022 |
| Depression | Decoder | 30 | 1.87 (2,58) | 1.000 | 0.061 |
| Depression | Training regime | 30 | 2.31 (1,29) | 1.000 | 0.074 |
| Depression | Forecast horizon | 30 | 2.44 (4,116) | 1.000 | 0.078 |
| Depression | Decoder x training regime | 30 | 2.09 (2,58) | 1.000 | 0.067 |
| Depression | Decoder x forecast horizon | 30 | 1.18 (8,232) | 1.000 | 0.039 |
| Depression | Training regime x forecast horizon | 30 | 2.26 (4,116) | 1.000 | 0.072 |
| Depression | Decoder x training regime x forecast horizon | 30 | 1.92 (8,232) | 1.000 | 0.062 |

**Panel D. Cluster 4**

| Channel | Effect | Participants, n | F (df1, df2) | Holm p | Partial eta2 |
| --- | --- | --- | --- | --- | --- |
| Total platform use | Decoder | 48 | 8.75 (2,94) | 0.051 | 0.157 |
| Total platform use | Training regime | 48 | 3.08 (1,47) | 1.000 | 0.062 |
| Total platform use | Forecast horizon | 48 | 0.49 (4,188) | 1.000 | 0.010 |
| Total platform use | Decoder x training regime | 48 | 4.26 (2,94) | 1.000 | 0.083 |
| Total platform use | Decoder x forecast horizon | 48 | 2.96 (8,376) | 0.436 | 0.059 |
| Total platform use | Training regime x forecast horizon | 48 | 4.56 (4,188) | 0.227 | 0.088 |
| Total platform use | Decoder x training regime x forecast horizon | 48 | 8.09 (8,376) | **<0.001** | 0.147 |
| Night-time platform use | Decoder | 48 | 13.77 (2,94) | **<0.001** | 0.227 |
| Night-time platform use | Training regime | 48 | 7.19 (1,47) | 1.000 | 0.133 |
| Night-time platform use | Forecast horizon | 48 | 2.13 (4,188) | 1.000 | 0.043 |
| Night-time platform use | Decoder x training regime | 48 | 11.06 (2,94) | **0.008** | 0.190 |
| Night-time platform use | Decoder x forecast horizon | 48 | 12.29 (8,376) | **<0.001** | 0.207 |
| Night-time platform use | Training regime x forecast horizon | 48 | 3.88 (4,188) | 0.633 | 0.076 |
| Night-time platform use | Decoder x training regime x forecast horizon | 48 | 2.57 (8,376) | 1.000 | 0.052 |
| Sleep duration | Decoder | 47 | 7.36 (2,92) | 0.163 | 0.138 |
| Sleep duration | Training regime | 47 | 9.57 (1,46) | 0.461 | 0.172 |
| Sleep duration | Forecast horizon | 47 | 0.73 (4,184) | 1.000 | 0.016 |
| Sleep duration | Decoder x training regime | 47 | 5.60 (2,92) | 0.667 | 0.108 |
| Sleep duration | Decoder x forecast horizon | 47 | 6.17 (8,368) | **<0.001** | 0.118 |
| Sleep duration | Training regime x forecast horizon | 47 | 4.26 (4,184) | 0.356 | 0.085 |
| Sleep duration | Decoder x training regime x forecast horizon | 47 | 0.57 (8,368) | 1.000 | 0.012 |
| Daily steps | Decoder | 48 | 21.56 (2,94) | **<0.001** | 0.314 |
| Daily steps | Training regime | 48 | 3.65 (1,47) | 1.000 | 0.072 |
| Daily steps | Forecast horizon | 48 | 3.05 (4,188) | 1.000 | 0.061 |
| Daily steps | Decoder x training regime | 48 | 3.41 (2,94) | 1.000 | 0.068 |
| Daily steps | Decoder x forecast horizon | 48 | 18.83 (8,376) | **<0.001** | 0.286 |
| Daily steps | Training regime x forecast horizon | 48 | 4.36 (4,188) | 0.305 | 0.085 |
| Daily steps | Decoder x training regime x forecast horizon | 48 | 8.31 (8,376) | **<0.001** | 0.150 |
| Stress | Decoder | 39 | 0.11 (2,76) | 1.000 | 0.003 |
| Stress | Training regime | 39 | 0.06 (1,38) | 1.000 | 0.002 |
| Stress | Forecast horizon | 39 | 0.51 (4,152) | 1.000 | 0.013 |
| Stress | Decoder x training regime | 39 | 0.92 (2,76) | 1.000 | 0.024 |
| Stress | Decoder x forecast horizon | 39 | 2.61 (8,304) | 1.000 | 0.064 |
| Stress | Training regime x forecast horizon | 39 | 0.46 (4,152) | 1.000 | 0.012 |
| Stress | Decoder x training regime x forecast horizon | 39 | 1.45 (8,304) | 1.000 | 0.037 |
| Positive affect | Decoder | 47 | 4.72 (2,92) | 1.000 | 0.093 |
| Positive affect | Training regime | 47 | 25.18 (1,46) | **0.001** | 0.354 |
| Positive affect | Forecast horizon | 47 | 1.96 (4,184) | 1.000 | 0.041 |
| Positive affect | Decoder x training regime | 47 | 1.66 (2,92) | 1.000 | 0.035 |
| Positive affect | Decoder x forecast horizon | 47 | 1.27 (8,368) | 1.000 | 0.027 |
| Positive affect | Training regime x forecast horizon | 47 | 0.95 (4,184) | 1.000 | 0.020 |
| Positive affect | Decoder x training regime x forecast horizon | 47 | 1.96 (8,368) | 1.000 | 0.041 |
| Depression | Decoder | 48 | 1.85 (2,94) | 1.000 | 0.038 |
| Depression | Training regime | 48 | 0.68 (1,47) | 1.000 | 0.014 |
| Depression | Forecast horizon | 48 | 1.43 (4,188) | 1.000 | 0.029 |
| Depression | Decoder x training regime | 48 | 1.49 (2,94) | 1.000 | 0.031 |
| Depression | Decoder x forecast horizon | 48 | 0.63 (8,376) | 1.000 | 0.013 |
| Depression | Training regime x forecast horizon | 48 | 2.31 (4,188) | 1.000 | 0.047 |
| Depression | Decoder x training regime x forecast horizon | 48 | 1.04 (8,376) | 1.000 | 0.022 |

**Supplementary Table S12.11b.** Holm-significant factor-level post hoc comparisons of participant-level within-subject skill relative to the causal prefix-mean baseline. Only decoder and training-regime contrasts with Holm-adjusted P < 0.05 are shown. Mean paired differences are reported with parametric 95%-CIs; positive differences favour the first level named. Model-versus-prefix-mean and model-versus-persistence benchmark contrasts remain available in the forecasting codebook and are not reproduced here.

| Cluster | Channel | Contrast family | Context | Horizon, d | Comparison | n | Mean difference (95% CI) | dz | Holm p |
| --- | --- | --- | --- | --- | --- | --- | --- | --- | --- |
| 1 | Sleep duration | Decoder comparison | Hybrid | 1 | Sticky HMM vs Native AR-HMM | 46 | -0.357 (-0.475, -0.238) | -0.892 | <0.001 |
| 1 | Daily steps | Decoder comparison | Hybrid | 1 | Sticky HMM vs Native AR-HMM | 46 | -0.729 (-0.997, -0.462) | -0.809 | 0.004 |
| 1 | Daily steps | Decoder comparison | Hybrid | 1 | Sticky HMM vs Sticky HMM + AR head | 46 | -0.717 (-0.988, -0.446) | -0.786 | 0.007 |
| 2 | Total platform use | Training-regime comparison | Sticky HMM + AR head | 14 | Synthetic vs Hybrid | 47 | 0.023 (0.014, 0.032) | 0.731 | 0.020 |
| 2 | Sleep duration | Decoder comparison | Hybrid | 1 | Sticky HMM vs Sticky HMM + AR head | 47 | -0.298 (-0.380, -0.216) | -1.069 | <0.001 |
| 2 | Daily steps | Decoder comparison | Hybrid | 1 | Sticky HMM vs Native AR-HMM | 47 | -0.805 (-1.054, -0.557) | -0.951 | <0.001 |
| 2 | Daily steps | Decoder comparison | Hybrid | 1 | Sticky HMM vs Sticky HMM + AR head | 47 | -0.804 (-1.056, -0.552) | -0.936 | <0.001 |
| 2 | Daily steps | Decoder comparison | Synthetic | 1 | Sticky HMM vs Native AR-HMM | 47 | -1.566 (-2.212, -0.920) | -0.711 | 0.031 |
| 2 | Daily steps | Decoder comparison | Synthetic | 1 | Sticky HMM vs Sticky HMM + AR head | 47 | -1.553 (-2.200, -0.905) | -0.704 | 0.037 |
| 3 | Night-time platform use | Decoder comparison | Synthetic | 1 | Sticky HMM vs Native AR-HMM | 44 | -0.555 (-0.759, -0.351) | -0.827 | 0.005 |
| 3 | Night-time platform use | Decoder comparison | Synthetic | 1 | Sticky HMM vs Sticky HMM + AR head | 44 | -0.494 (-0.695, -0.294) | -0.751 | 0.025 |
| 3 | Sleep duration | Decoder comparison | Hybrid | 1 | Sticky HMM vs Native AR-HMM | 44 | -0.372 (-0.507, -0.238) | -0.842 | 0.004 |
| 3 | Sleep duration | Decoder comparison | Synthetic | 1 | Sticky HMM vs Native AR-HMM | 44 | -1.444 (-1.988, -0.900) | -0.807 | 0.008 |
| 3 | Sleep duration | Decoder comparison | Synthetic | 1 | Sticky HMM vs Sticky HMM + AR head | 44 | -1.198 (-1.684, -0.712) | -0.749 | 0.026 |
| 3 | Daily steps | Decoder comparison | Hybrid | 1 | Sticky HMM vs Native AR-HMM | 44 | -0.659 (-0.748, -0.569) | -2.239 | <0.001 |
| 3 | Daily steps | Decoder comparison | Hybrid | 1 | Sticky HMM vs Sticky HMM + AR head | 44 | -0.647 (-0.737, -0.557) | -2.182 | <0.001 |
| 3 | Daily steps | Decoder comparison | Synthetic | 1 | Sticky HMM vs Native AR-HMM | 44 | -0.907 (-1.110, -0.703) | -1.357 | <0.001 |
| 3 | Daily steps | Decoder comparison | Synthetic | 1 | Sticky HMM vs Sticky HMM + AR head | 44 | -0.898 (-1.098, -0.698) | -1.366 | <0.001 |
| 3 | Daily steps | Decoder comparison | Hybrid | 3 | Sticky HMM vs Native AR-HMM | 44 | -0.236 (-0.323, -0.150) | -0.829 | 0.005 |
| 3 | Daily steps | Decoder comparison | Hybrid | 3 | Sticky HMM vs Sticky HMM + AR head | 44 | -0.247 (-0.329, -0.165) | -0.918 | <0.001 |
| 3 | Daily steps | Decoder comparison | Synthetic | 3 | Sticky HMM vs Native AR-HMM | 44 | -0.389 (-0.549, -0.229) | -0.740 | 0.032 |
| 3 | Daily steps | Decoder comparison | Synthetic | 3 | Sticky HMM vs Sticky HMM + AR head | 44 | -0.394 (-0.560, -0.228) | -0.723 | 0.046 |
| 3 | Daily steps | Decoder comparison | Hybrid | 21 | Sticky HMM vs Native AR-HMM | 44 | -0.191 (-0.267, -0.115) | -0.764 | 0.019 |
| 3 | Daily steps | Decoder comparison | Hybrid | 21 | Sticky HMM vs Sticky HMM + AR head | 44 | -0.181 (-0.247, -0.115) | -0.839 | 0.004 |
| 3 | Daily steps | Training-regime comparison | Sticky HMM | 21 | Synthetic vs Hybrid | 44 | 0.166 (0.098, 0.234) | 0.740 | 0.032 |
| 4 | Total platform use | Decoder comparison | Hybrid | 1 | Sticky HMM + AR head vs Native AR-HMM | 48 | 0.051 (0.031, 0.071) | 0.731 | 0.016 |
| 4 | Total platform use | Decoder comparison | Synthetic | 1 | Sticky HMM + AR head vs Native AR-HMM | 48 | 0.053 (0.032, 0.073) | 0.743 | 0.012 |
| 4 | Total platform use | Decoder comparison | Hybrid | 3 | Sticky HMM + AR head vs Native AR-HMM | 48 | 0.083 (0.050, 0.115) | 0.741 | 0.013 |
| 4 | Total platform use | Decoder comparison | Synthetic | 3 | Sticky HMM + AR head vs Native AR-HMM | 48 | 0.087 (0.051, 0.124) | 0.696 | 0.036 |
| 4 | Sleep duration | Decoder comparison | Hybrid | 3 | Sticky HMM + AR head vs Native AR-HMM | 47 | -0.236 (-0.326, -0.145) | -0.762 | 0.010 |
| 4 | Sleep duration | Decoder comparison | Synthetic | 3 | Sticky HMM + AR head vs Native AR-HMM | 47 | -0.286 (-0.393, -0.179) | -0.783 | 0.006 |
| 4 | Daily steps | Decoder comparison | Hybrid | 1 | Sticky HMM vs Native AR-HMM | 48 | -0.622 (-0.726, -0.518) | -1.732 | <0.001 |
| 4 | Daily steps | Decoder comparison | Hybrid | 1 | Sticky HMM vs Sticky HMM + AR head | 48 | -0.643 (-0.749, -0.537) | -1.754 | <0.001 |
| 4 | Daily steps | Decoder comparison | Synthetic | 1 | Sticky HMM vs Native AR-HMM | 48 | -1.080 (-1.435, -0.726) | -0.885 | <0.001 |
| 4 | Daily steps | Decoder comparison | Synthetic | 1 | Sticky HMM vs Sticky HMM + AR head | 48 | -1.096 (-1.461, -0.732) | -0.874 | <0.001 |

**Supplementary Table S12.12.** Channel-specific oracle gains in the 2 AR-HMM forecast-attenuation audit (n=13 participants). Oracle gain was defined as 100 x (MSE_standard_ – MSE_oracle_) / MSE_standard_; positive values therefore indicate lower mean-squared prediction error than the standard AR-HMM forecast. Participant-specific gains were summarized across the primary 7-, 14-, and 21-day horizons and across particle-grouped split repetitions before cohort aggregation. Percentile 95% confidence intervals were estimated from 1,000 participant-level bootstrap resamples. The combined oracle supplied both the future Viterbi state path and the realized preceding target value; its gain is not expected to equal the sum of the two single-oracle gains. Mechanism classifications used the prespecified 15-percentage-point meaningful-gain threshold and 10-percentage-point dominance threshold. Ordinal EMA targets are not shown because the continuous future-lag oracle was not applicable to their forecasting model.

| **Target channel** | **Participants, n** | **Future-state oracle gain, median % (95% CI)** | **Future-lag oracle gain, median % (95% CI)** | **Combined state-and-lag oracle gain, median % (95% CI)** | **Mechanism classification** |
| --- | --- | --- | --- | --- | --- |
| Night-time platform use | 13 | 34.7 (29.2-44.0) | 12.6 (6.5-21.5) | 47.5 (37.2-51.8) | Mixed state-AR (state-led) |
| Total platform use | 13 | 34.7 (29.5-38.2) | 14.4 (7.7-25.9) | 43.3 (40.5-50.8) | State-uncertainty dominant |
| Sleep duration | 13 | 5.6 (1.4-5.9) | 4.2 (2.7-8.9) | 13.9 (12.2-17.0) | Oracle-resistant residual limitation |
| Daily steps | 13 | 0.5 (-0.5-3.5) | 60.8 (50.7-67.5) | 61.9 (51.5-67.7) | Recursive-AR dominant |

**Supplementary Table S13a. Group-level split-half tests of hard level and ABC-SMC-weighted soft virtual-twin target specifications.** Signed relative displacement was calculated as first-half minus full-series anchor, divided by the absolute full-series anchor; negative values therefore indicate lower first-half anchors. Displacement in corridor widths uses the full-series corridor width as denominator. For soft targets, ABC-SMC weights were applied within participant before group-level testing; Wilcoxon signed-rank and exact binomial sign tests used participant-level summaries as the independent observations. p-values are unadjusted diagnostic statistics.

**Panel A. Hard level target specifications**

| **Sample** | **Participants, n** | **Paired target rows, n** | **Median participant signed relative shift** | **Median participant shift in corridor widths** | **Corridor breaches, n/N (%)** | **Wilcoxon p** | **Exact sign-test p** |
| --- | --- | --- | --- | --- | --- | --- | --- |
| All clusters | 80 | 640 | -0.44% | -0.014 | 98/640 (15.3%) | 0.952 | 0.434 |
| Cluster 1 | 20 | 160 | 0.42% | 0.014 | 16/160 (10.0%) | 0.668 | 0.824 |
| Cluster 2 | 20 | 160 | -1.05% | -0.027 | 31/160 (19.4%) | 0.225 | 0.115 |
| Cluster 3 | 20 | 160 | 0.31% | 0.008 | 21/160 (13.1%) | 0.211 | 1.000 |
| Cluster 4 | 20 | 160 | -0.32% | -0.019 | 30/160 (18.8%) | 0.588 | 0.824 |

**Panel B. ABC-SMC-weighted empirical soft target specifications**

| **Target scope** | **Sample** | **Participants, n** | **Cumulative ABC-SMC weight per participant** | **Median weighted signed shift in corridor widths** | **Weighted breach rate** | **Wilcoxon p** | **Exact sign-test p** |
| --- | --- | --- | --- | --- | --- | --- | --- |
| All scored empirical soft targets | All clusters | 80 | 4.75 | -0.164 | 22.1% | **<0.001** | **<0.001** |
| All scored empirical soft targets | Cluster 1 | 20 | 4.75 | -0.167 | 21.4% | **<0.001** | **<0.001** |
| All scored empirical soft targets | Cluster 2 | 20 | 4.75 | -0.188 | 24.6% | **<0.001** | **<0.001** |
| All scored empirical soft targets | Cluster 3 | 20 | 4.75 | -0.128 | 16.6% | **<0.001** | **0.003** |
| All scored empirical soft targets | Cluster 4 | 20 | 4.75 | -0.163 | 26.0% | **0.001** | **0.003** |
| Threshold/extreme-rate targets | All clusters | 80 | 0.85 | 0.000 | 27.1% | 0.820 | 0.807 |
| Threshold/extreme-rate targets | Cluster 1 | 20 | 0.85 | -0.027 | 32.4% | 0.443 | 0.302 |
| Threshold/extreme-rate targets | Cluster 2 | 20 | 0.85 | -0.019 | 25.3% | 0.926 | 1.000 |
| Threshold/extreme-rate targets | Cluster 3 | 20 | 0.85 | 0.000 | 10.0% | 0.162 | 0.581 |
| Threshold/extreme-rate targets | Cluster 4 | 20 | 0.85 | 0.142 | 40.6% | 0.205 | 0.359 |
| Within-subject dispersion targets | All clusters | 80 | 3.20 | -0.258 | 25.1% | **<0.001** | **<0.001** |
| Within-subject dispersion targets | Cluster 1 | 20 | 3.20 | -0.245 | 23.1% | **<0.001** | **<0.001** |
| Within-subject dispersion targets | Cluster 2 | 20 | 3.20 | -0.265 | 29.8% | **<0.001** | **<0.001** |
| Within-subject dispersion targets | Cluster 3 | 20 | 3.20 | -0.128 | 20.9% | **0.001** | **0.003** |
| Within-subject dispersion targets | Cluster 4 | 20 | 3.20 | -0.279 | 26.7% | **<0.001** | **<0.001** |
| Simple autocorrelation targets | All clusters | 80 | 0.70 | -0.046 | 2.5% | **<0.001** | **0.005** |
| Simple autocorrelation targets | Cluster 1 | 20 | 0.70 | -0.019 | 0.0% | 0.563 | 0.824 |
| Simple autocorrelation targets | Cluster 2 | 20 | 0.70 | -0.051 | 0.0% | **0.032** | **0.041** |
| Simple autocorrelation targets | Cluster 3 | 20 | 0.70 | -0.076 | 5.0% | **0.007** | **0.041** |
| Simple autocorrelation targets | Cluster 4 | 20 | 0.70 | -0.044 | 5.0% | 0.287 | 0.503 |

**Supplementary Table S13b.** Channel-specific split-half stability of hard level virtual-twin target specifications. Panel A presents medians and percentile 95% confidence intervals from 5,000 participant-level bootstrap resamples stratified by cluster. A corridor breach occurred when the full-series anchor lay outside the corresponding first-half corridor. Panel B reports unadjusted diagnostic group-level tests within each cluster; each cluster contained 20 participants. The Wilcoxon and sign tests evaluate systematic signed anchor displacement and do not test the absolute magnitude of target movement.

**Panel A. Complete balanced sample (n=80)**

| **Hard level target** | **Absolute relative shift, median % (95% CI)** | **Signed relative shift, median % (95% CI)** | **Jaccard overlap, median (95% CI)** | **Corridor breaches, n/N (%) and 95% CI** | **Wilcoxon p** | **Exact sign-test p** |
| --- | --- | --- | --- | --- | --- | --- |
| Total platform use | 4.4 (3.7-5.1) | -0.3 (-1.6-0.9) | 0.641 (0.592-0.692) | 5/80 (6.2%) 95% CI 1.2-11.2% | 0.714 | 0.738 |
| Night-time platform use | 6.1 (5.2-8.0) | -0.3 (-2.3-3.9) | 0.531 (0.433-0.580) | 24/80 (30.0%) 95% CI 20.0-40.0% | 0.668 | 0.911 |
| Night-time platform share | 4.2 (3.5-6.1) | -0.4 (-1.3-0.8) | 0.763 (0.694-0.826) | 6/80 (7.5%) 95% CI 2.5-13.8% | 0.948 | 0.434 |
| Sleep duration | 2.3 (1.9-2.7) | -0.1 (-0.8-2.0) | 0.799 (0.762-0.825) | 2/80 (2.5%) 95% CI 0.0-6.2% | 0.177 | 0.911 |
| Daily steps | 9.0 (7.0-11.7) | -2.3 (-4.2-2.2) | 0.389 (0.290-0.484) | 35/80 (43.8%) 95% CI 32.5-54.6% | 0.510 | 0.314 |
| Stress | 4.5 (3.6-5.6) | -0.2 (-2.8-1.1) | 0.730 (0.635-0.770) | 4/80 (5.0%) 95% CI 1.2-10.0% | 0.413 | 0.731 |
| Positive affect | 7.1 (5.6-8.2) | 1.0 (-2.0-2.9) | 0.723 (0.646-0.769) | 8/80 (10.0%) 95% CI 3.8-16.2% | 0.790 | 0.911 |
| Depression | 14.2 (11.6-22.4) | 0.0 (-5.7-0.0) | 0.699 (0.610-0.798) | 14/80 (17.5%) 95% CI 10.0-25.0% | 0.586 | 0.336 |

**Panel B. Cluster-specific group-level diagnostic tests**

| **Cluster** | **Hard level target** | **Participants, n** | **Median signed relative shift** | **Corridor breaches, n/N (%)** | **Wilcoxon p** | **Exact sign-test p** |
| --- | --- | --- | --- | --- | --- | --- |
| Cluster 1 | Total platform use | 20 | -1.69% | 0/20 (0.0%) | 0.779 | 0.503 |
| Cluster 1 | Night-time platform use | 20 | 3.30% | 7/20 (35.0%) | 0.380 | 0.503 |
| Cluster 1 | Night-time platform share | 20 | 1.56% | 0/20 (0.0%) | 0.121 | 0.503 |
| Cluster 1 | Sleep duration | 20 | 0.77% | 1/20 (5.0%) | 0.444 | 0.824 |
| Cluster 1 | Daily steps | 20 | -2.40% | 6/20 (30.0%) | 0.538 | 0.824 |
| Cluster 1 | Stress | 20 | -4.33% | 1/20 (5.0%) | 0.131 | 0.167 |
| Cluster 1 | Positive affect | 20 | 1.29% | 1/20 (5.0%) | 0.401 | 1.000 |
| Cluster 1 | Depression | 20 | 0.00% | 0/20 (0.0%) | 0.286 | 0.424 |
| Cluster 2 | Total platform use | 20 | 2.16% | 2/20 (10.0%) | 0.198 | 0.503 |
| Cluster 2 | Night-time platform use | 20 | 2.15% | 7/20 (35.0%) | 0.444 | 1.000 |
| Cluster 2 | Night-time platform share | 20 | -0.79% | 4/20 (20.0%) | 0.668 | 0.503 |
| Cluster 2 | Sleep duration | 20 | 0.92% | 1/20 (5.0%) | 0.514 | 1.000 |
| Cluster 2 | Daily steps | 20 | -4.81% | 8/20 (40.0%) | **0.022** | 0.115 |
| Cluster 2 | Stress | 20 | -0.05% | 0/20 (0.0%) | 0.360 | 1.000 |
| Cluster 2 | Positive affect | 20 | -4.17% | 1/20 (5.0%) | 0.162 | 0.115 |
| Cluster 2 | Depression | 20 | -3.19% | 8/20 (40.0%) | 0.198 | 0.503 |
| Cluster 3 | Total platform use | 20 | -0.56% | 1/20 (5.0%) | 0.723 | 0.503 |
| Cluster 3 | Night-time platform use | 20 | -0.40% | 6/20 (30.0%) | 1.000 | 1.000 |
| Cluster 3 | Night-time platform share | 20 | -0.63% | 1/20 (5.0%) | 0.867 | 0.824 |
| Cluster 3 | Sleep duration | 20 | -0.32% | 0/20 (0.0%) | 0.490 | 0.824 |
| Cluster 3 | Daily steps | 20 | -1.02% | 10/20 (50.0%) | 0.380 | 0.503 |
| Cluster 3 | Stress | 20 | 1.51% | 1/20 (5.0%) | 0.142 | 0.629 |
| Cluster 3 | Positive affect | 20 | 2.37% | 2/20 (10.0%) | 0.151 | 0.115 |
| Cluster 3 | Depression | 20 | 4.76% | 0/20 (0.0%) | 0.118 | 0.118 |
| Cluster 4 | Total platform use | 20 | -0.26% | 2/20 (10.0%) | 0.985 | 1.000 |
| Cluster 4 | Night-time platform use | 20 | -1.98% | 4/20 (20.0%) | 0.287 | 0.263 |
| Cluster 4 | Night-time platform share | 20 | -1.74% | 1/20 (5.0%) | 0.173 | 0.263 |
| Cluster 4 | Sleep duration | 20 | -0.30% | 0/20 (0.0%) | 0.588 | 0.824 |
| Cluster 4 | Daily steps | 20 | 5.96% | 11/20 (55.0%) | 0.185 | 0.503 |
| Cluster 4 | Stress | 20 | -0.30% | 2/20 (10.0%) | 0.563 | 1.000 |
| Cluster 4 | Positive affect | 20 | 1.63% | 4/20 (20.0%) | 0.422 | 0.824 |
| Cluster 4 | Depression | 20 | -9.27% | 6/20 (30.0%) | 0.341 | 0.115 |

**Supplementary Table S13c. Posterior and downstream functional robustness of split-half virtual twins in clusters 1 and 2.** Split-half VTs were calibrated from target specifications derived only from the first temporal half of each participant’s observed series and compared with matched full-timeseries VTs. C2 values are mean ± SD across observed subjects. C3 causal skill is the channel-balanced skill relative to the causal participant-specific prefix mean. Retained skill is split-half skill divided by full-timeseries skill.

**Panel A. ABC-SMC particle-cloud comparison**

| **Cluster** | **Subjects** | **Posterior median r** | **Posterior IQR r** | **Median \|shift\| (SD)** | **>1 SD** | **>2 SD** | **Cross/within ratio** | **Split/full IQR ratio** |
| --- | --- | --- | --- | --- | --- | --- | --- | --- |
| CL1 | 20 | 0.989 | 0.969 | 0.650 | 31.1% | 4.7% | 1.397 | 1.050 |
| CL2 | 20 | 0.988 | 0.977 | 0.720 | 35.1% | 5.9% | 1.420 | 1.133 |

**Panel B. Synthetic-to-observed decoding correspondence between AR-HMM models**

| **Metric** | **CL1 split-half** | **CL1 full-series** | **CL2 split-half** | **CL2 full-series** |
| --- | --- | --- | --- | --- |
| Occupancy correlation | 0.790 ± 0.120 | 0.784 ± 0.131 | 0.842 ± 0.144 | 0.840 ± 0.119 |
| Occupancy TV distance | 0.219 ± 0.092 | 0.215 ± 0.096 | 0.189 ± 0.099 | 0.216 ± 0.097 |
| Transition correlation | 0.940 ± 0.034 | 0.942 ± 0.035 | 0.948 ± 0.028 | 0.951 ± 0.029 |
| Transition JSD | 0.068 ± 0.030 | 0.066 ± 0.039 | 0.069 ± 0.035 | 0.069 ± 0.048 |
| Fingerprint correlation | 0.740 ± 0.112 | 0.799 ± 0.088 | 0.707 ± 0.131 | 0.739 ± 0.129 |
| Fingerprint Frobenius distance | 0.428 ± 0.135 | 0.411 ± 0.164 | 0.407 ± 0.135 | 0.410 ± 0.164 |

**Panel C. Causal forecasting skill and preservation (Synthetic-only training regime) of AR-HMM models**

| **Horizon** | **CL1 split** | **CL1 full** | **CL1 retained** | **CL2 split** | **CL2 full** | **CL2 retained** | **Mean retained** |
| --- | --- | --- | --- | --- | --- | --- | --- |
| 1 d | 0.475 | 0.479 | 99.2% | 0.565 | 0.577 | 97.8% | 98.5% |
| 3 d | 0.411 | 0.429 | 95.8% | 0.497 | 0.518 | 96.1% | 96.0% |
| 7 d | 0.396 | 0.439 | 90.2% | 0.449 | 0.489 | 91.9% | 91.1% |
| 14 d | 0.385 | 0.452 | 85.3% | 0.419 | 0.492 | 85.3% | 85.3% |
| 21 d | 0.394 | 0.477 | 82.7% | 0.399 | 0.495 | 80.7% | 81.7% |

**Notes.** Particle-cloud comparisons included 20 matched participants per cluster, with 50 accepted particles generated per VT and 1,644 parameters common to the split-half and full-series models. Decoding included all 20 participants in each cluster. For forecasting, quality control retained the same participants in the split-half and full-series analyses: 19/20 in CL1 and 18/20 in CL2. Hybrid-online forecasting results showed a comparable preservation pattern.

### Supplementary Figures

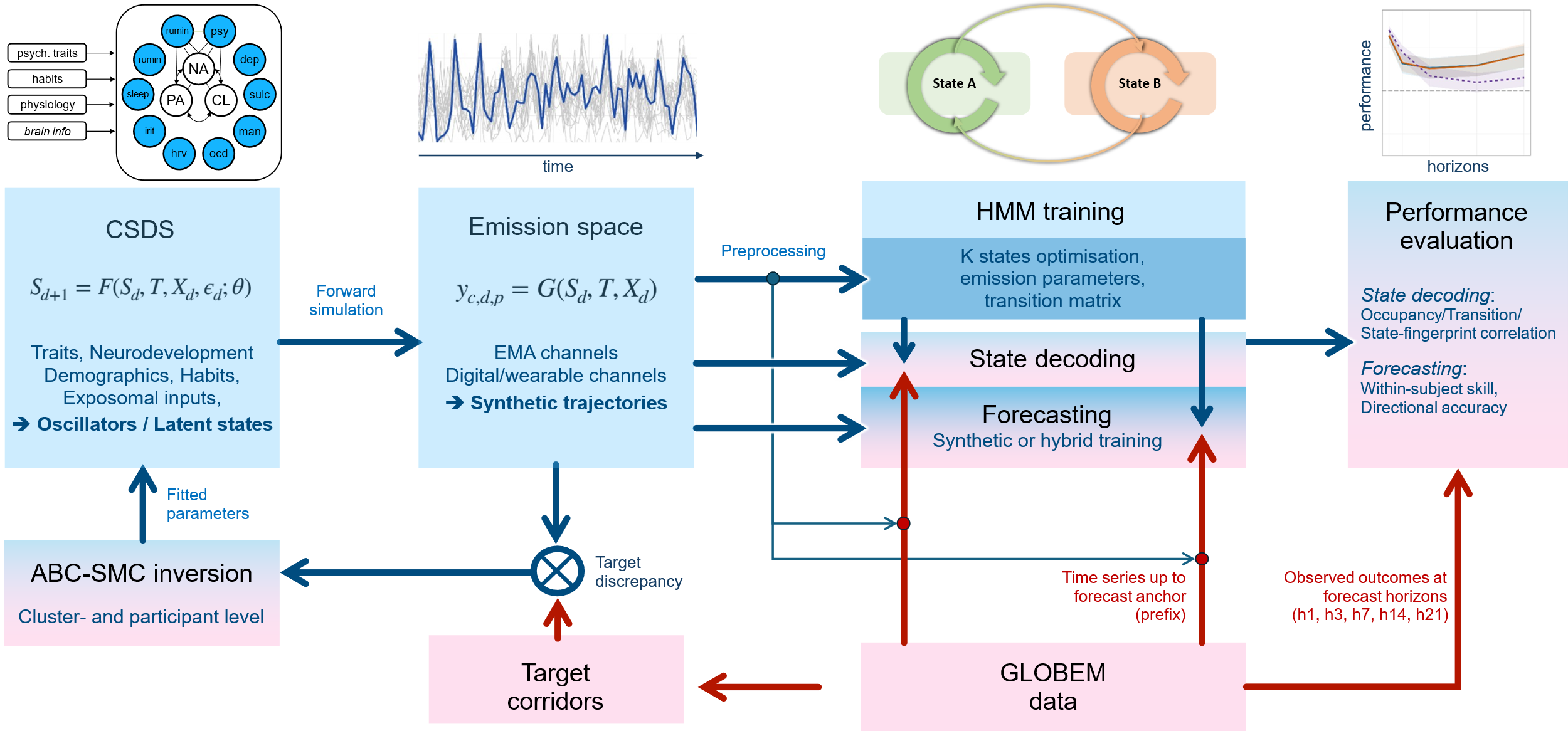

**Supplementary Figure S1: Overview of the study design and computational workflow.** The Coupled Stochastic Dynamical System (CSDS) generates latent mental-health, behavioural and physiological states from person-level traits, neurodevelopmental and demographic factors, habits and stochastic exposomal inputs. These latent dynamics are mapped through the emission model onto observable ecological momentary assessment (EMA), digital-behavioural and wearable-derived time series. Empirical GLOBEM data are used to define phenotype target corridors, against which simulated emissions are evaluated during approximate Bayesian computation with sequential Monte Carlo-style proposal refinement (ABC-SMC); accepted parameter configurations are returned to the CSDS to generate cluster- or participant-specific synthetic trajectories. Preprocessed synthetic trajectories are subsequently used to train participant-specific Hidden Markov Models (HMMs), including optimisation of the number of latent states, emission parameters and transition structure. The trained models are applied to observed GLOBEM time series for state decoding and forecasting. Forecasts are generated under synthetic or hybrid regimes using only observations available up to each forecast anchor (causal prefix), and are evaluated against subsequently observed data at horizons of 1, 3, 7, 14 and 21 days. State-decoding performance is quantified from concordance of state occupancy, transition structure and state fingerprints between synthetic and observed data; forecasting performance is assessed using within-subject skill and directional accuracy. Blue pathways denote model generation and model-based analysis, whereas red pathways indicate uses of empirical GLOBEM data for model inversion, causal-prefix forecasting and evaluation.

**Supplementary Figures S10.1a–d (next pages). Representative trajectory ensembles from top-ranked -level CSDS particles.**
For each empirical , 20 independent simulations were generated from the top-ranked accepted -level particle. Grey lines show individual stochastic realizations; the blue line shows one representative realization. Panels display the principal digital, wearable and EMA-derived channels used in the validation framework. The lower summary panel compares run-level means with the empirical target corridors. These figures illustrate that accepted -level solutions generate distinct -typical dynamic regimes while also revealing residual calibration mismatch for selected channels.

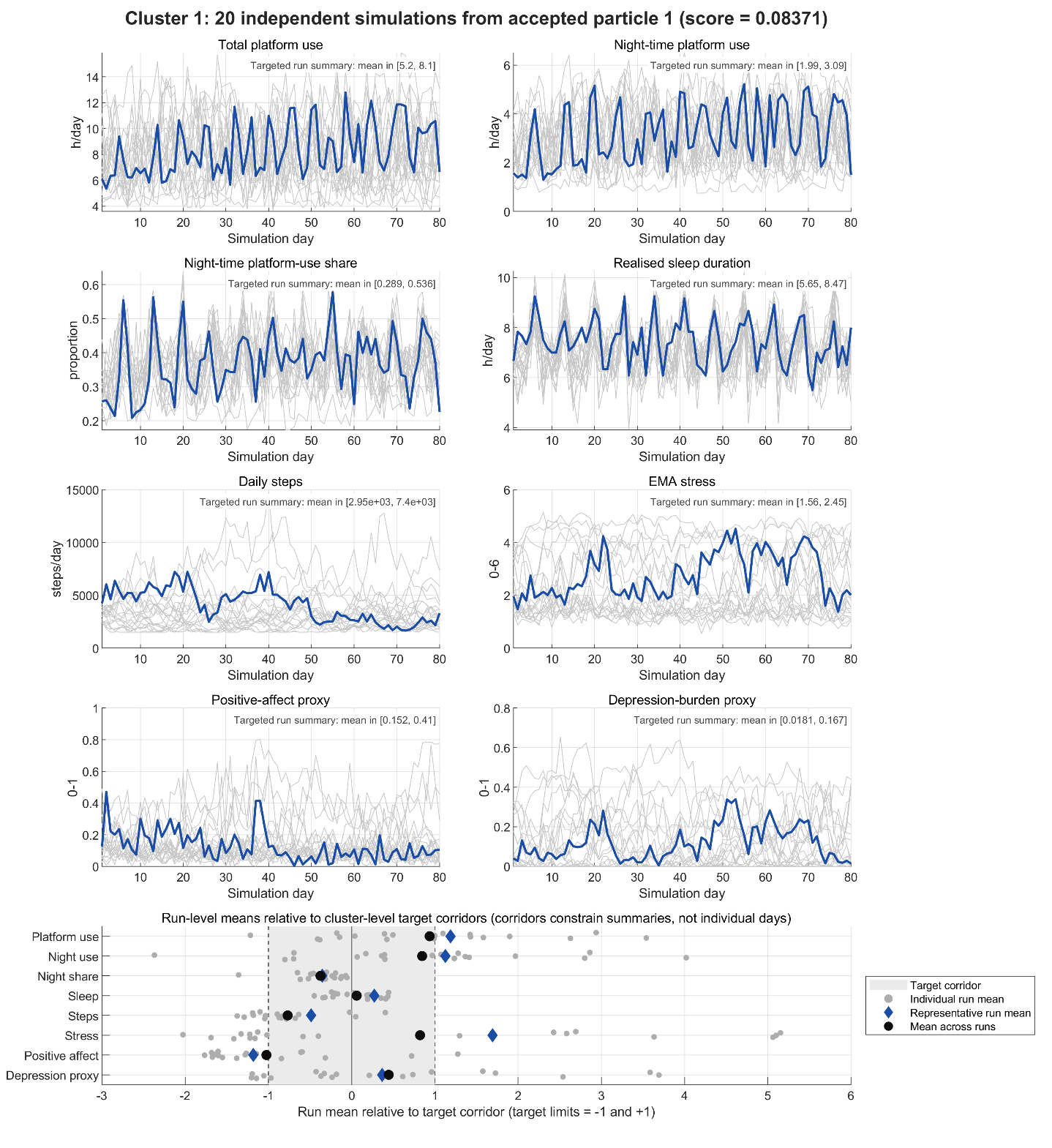

**Supplementary Figure S10.1a**: Representative trajectory ensembles from the top-ranked -level particle of GLOBEM cluster 1.

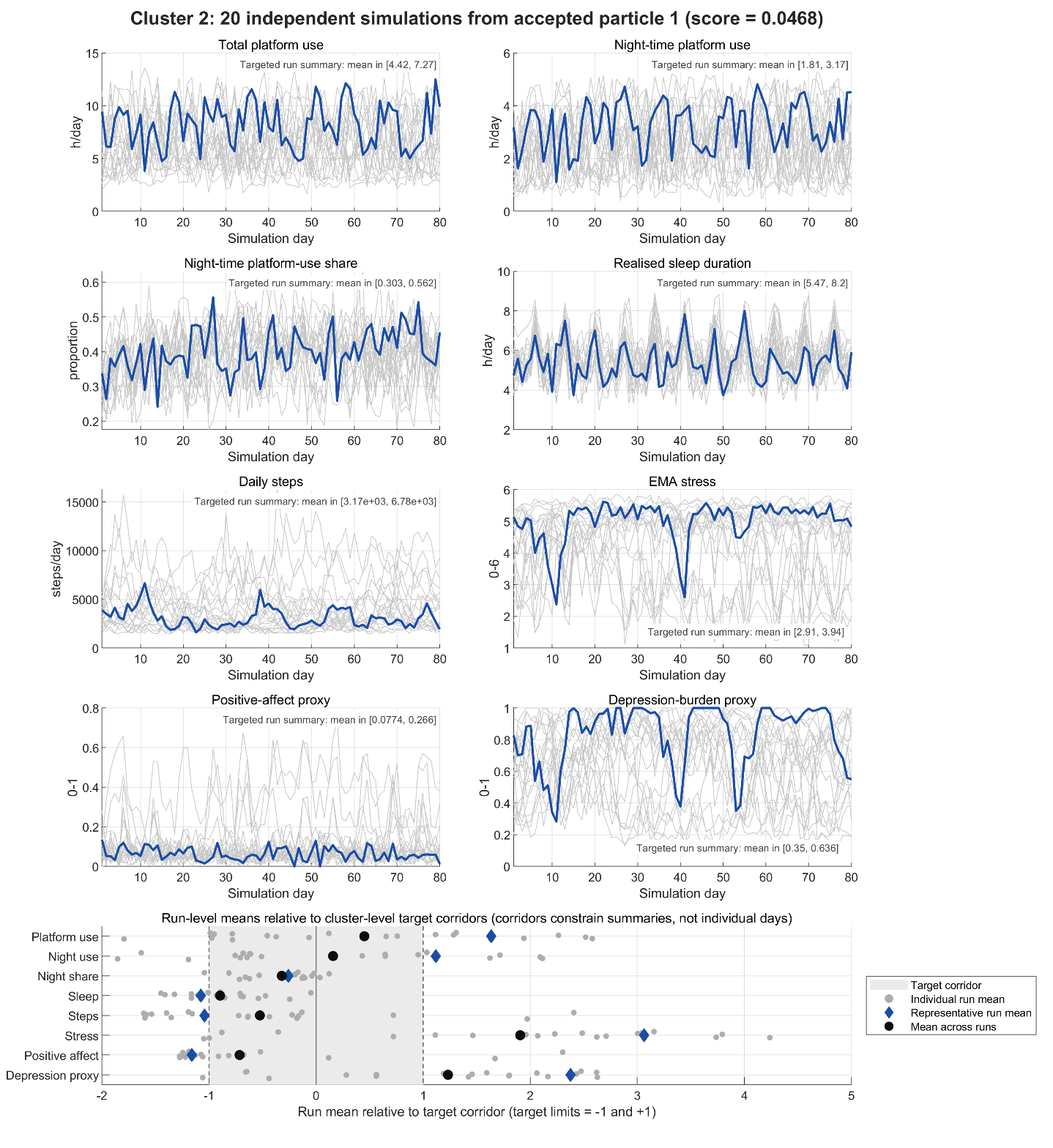

**Supplementary Figure S10.1b**: Representative trajectory ensembles from the top-ranked -level particle of GLOBEM cluster 2.

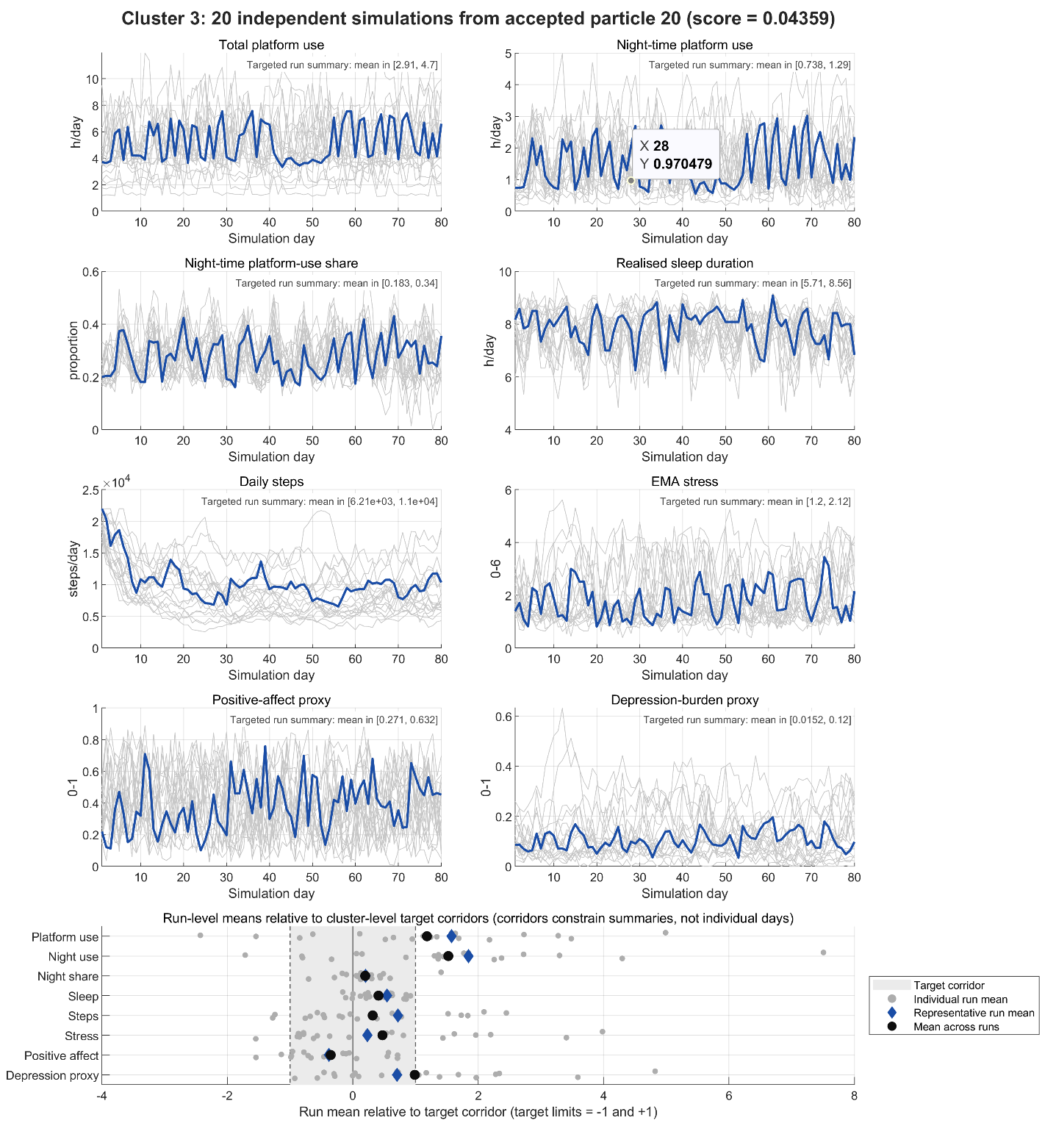

**Supplementary Figure S10.1c**: Representative trajectory ensembles from the top-ranked -level particle of GLOBEM cluster 3.

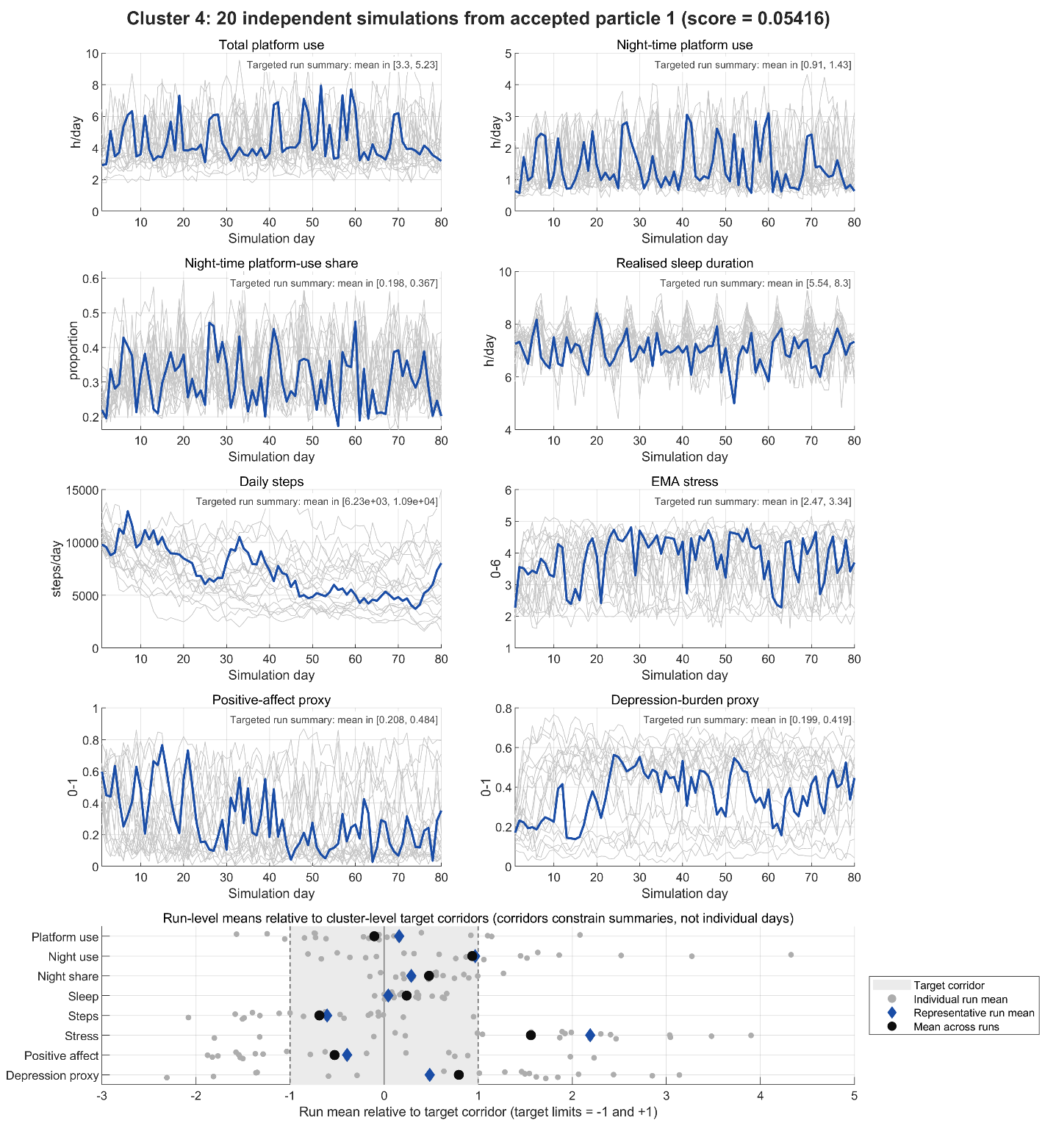

**Supplementary Figure S10.1d**: Representative trajectory ensembles from the top-ranked -level particle of GLOBEM cluster 4.

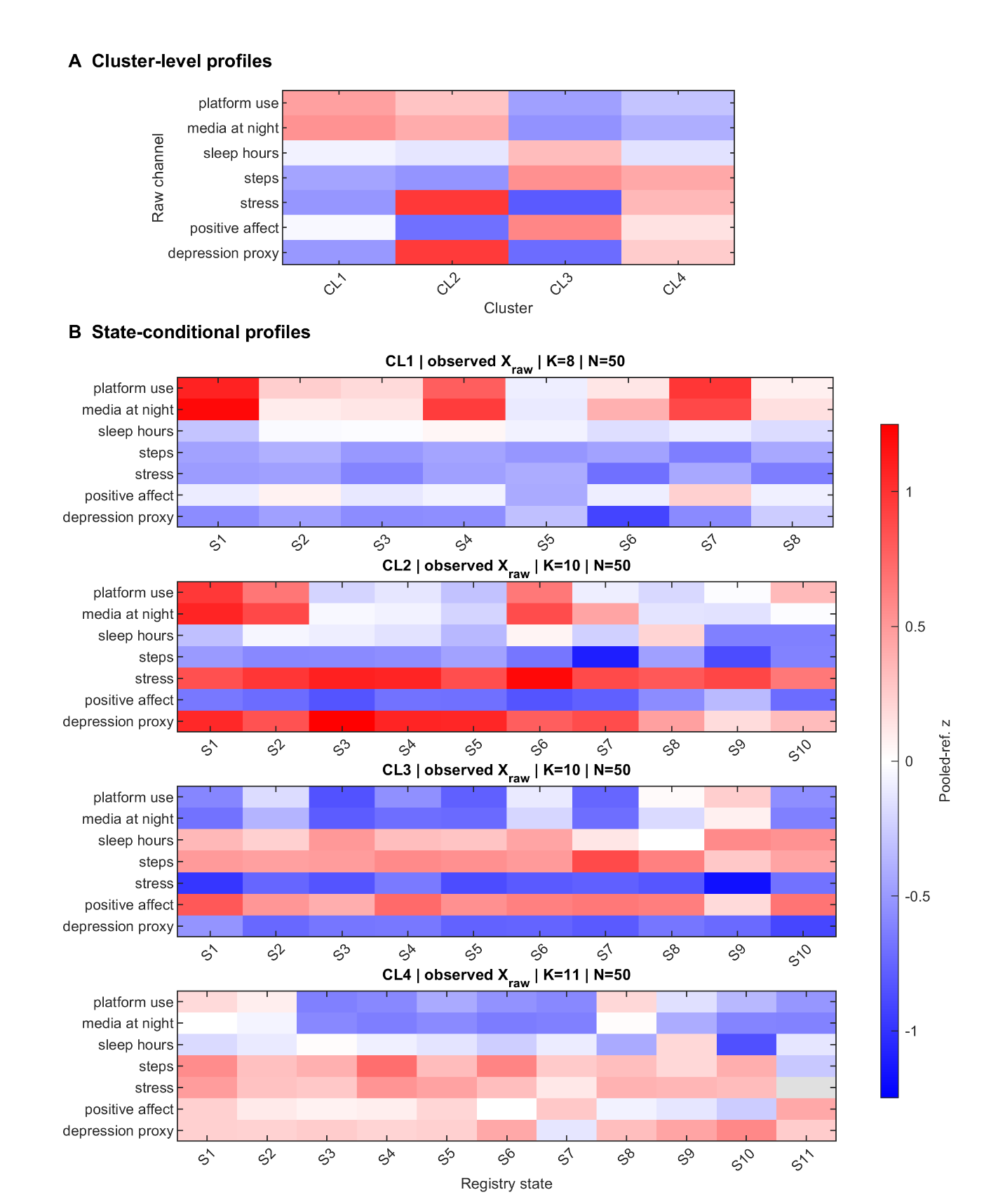

**Supplementary Figure S11.5a. Observed raw-channel levels across GLOBEM clusters and registry states.** **(A)** Cluster-level profiles of the seven observed raw channels. For each participant, channel means were calculated across observations assigned to supported aligned registry states and were then averaged across participants. **(B)** State-conditional profiles for each cluster-specific registry state. Raw-channel means were first calculated separately within each participant and state and were subsequently averaged across participants. Values in both panels are feature-wise z-scores relative to a common equal-subject-weighted pooled four-cluster $X_{raw}$ mean and standard deviation. Positive and negative values therefore indicate levels above or below the pooled reference for the respective channel, while preserving between-cluster differences.
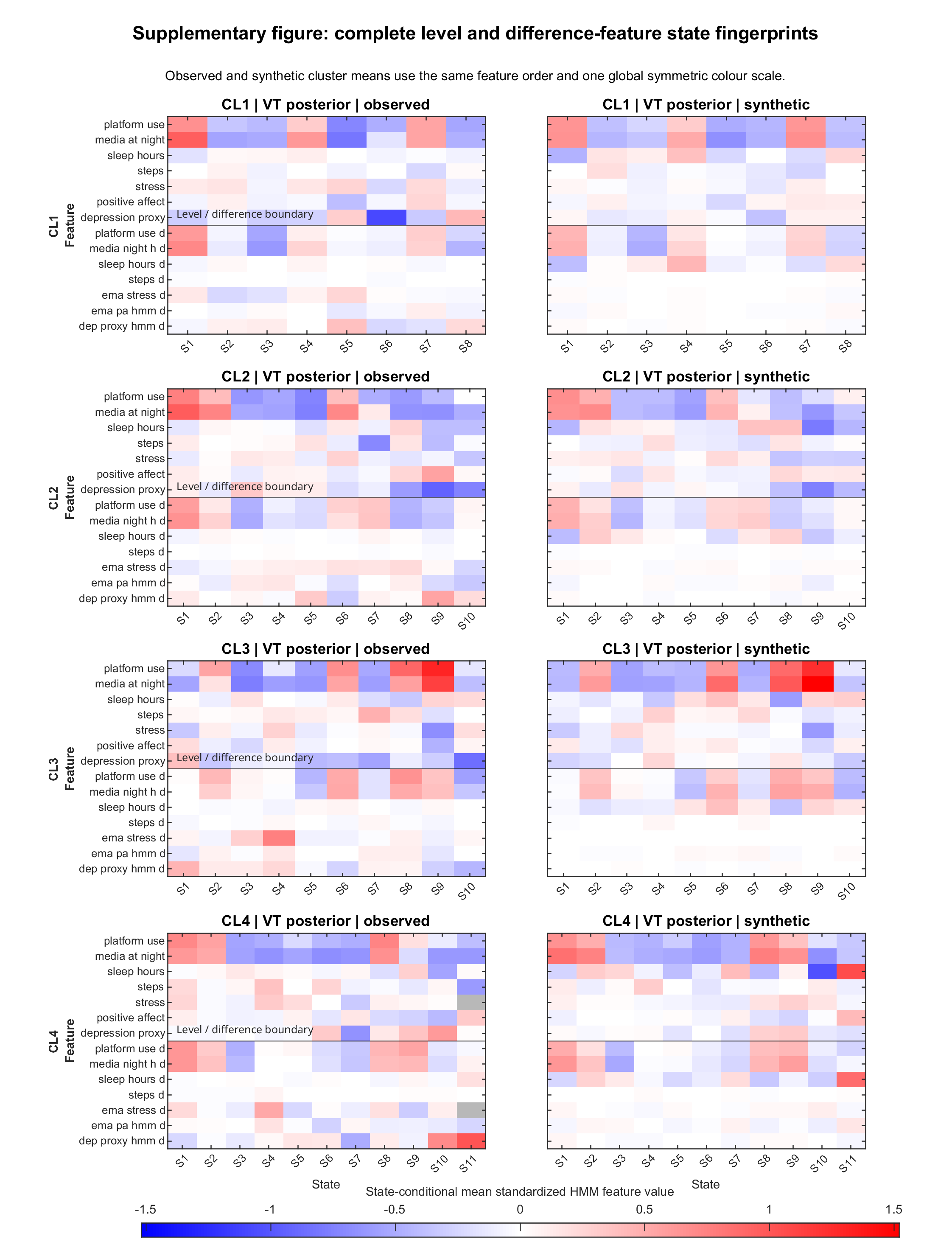

**Supplementary Figure S11.5b. Complete observed and synthetic sticky-HMM state fingerprints.** Equal-subject-weighted state-conditional mean fingerprints are shown for the seven level features and the corresponding seven difference features in each GLOBEM. Observed and synthetic panels use the same feature ordering and a common global symmetric colour scale. Difference features are denoted by the suffix _d and represent the change-feature component of the HMM input space. States were aligned within s only; identically numbered states do not imply cross- homology.

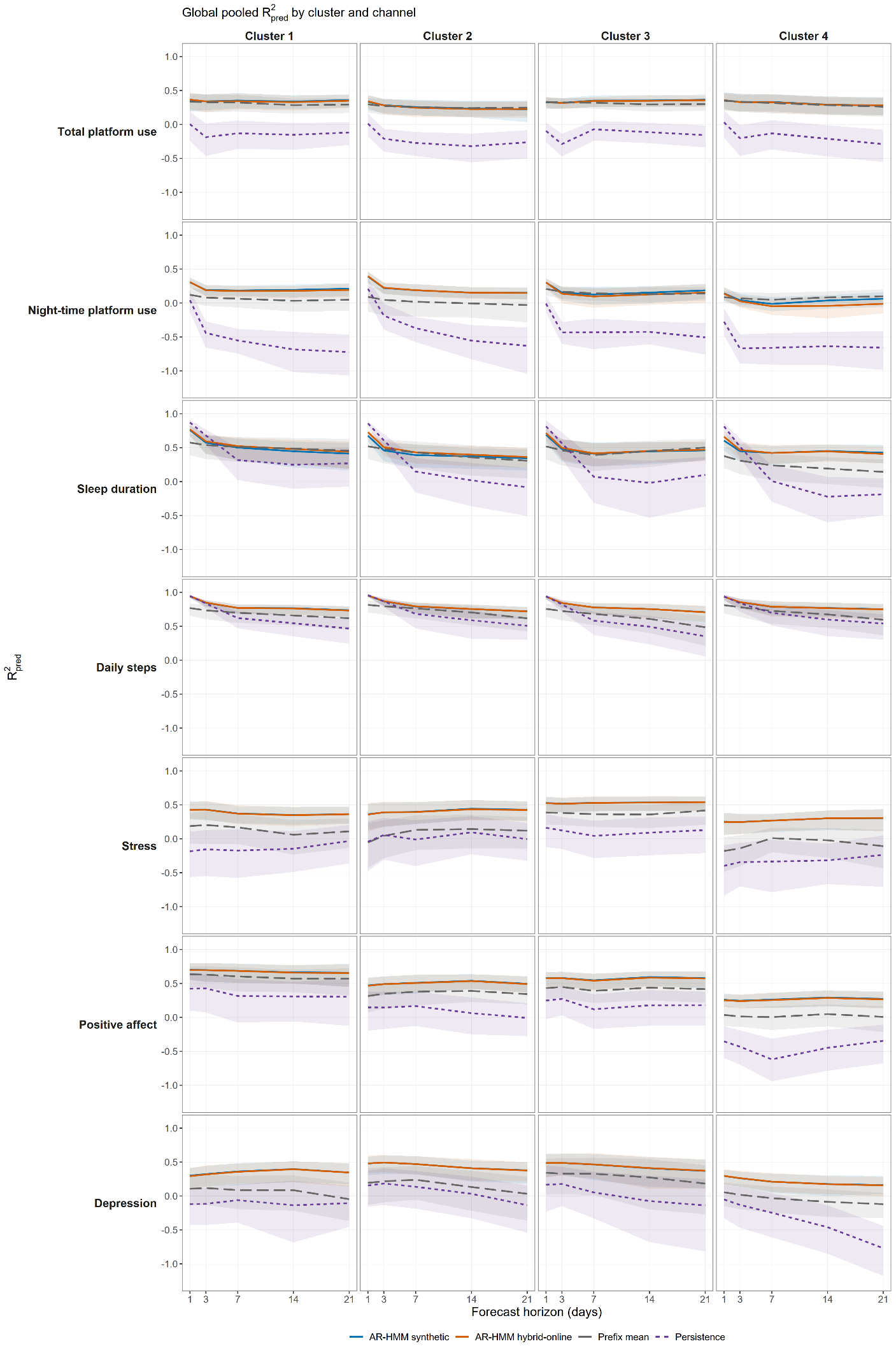

**Supplementary Figure S12.6a. Global pooled prediction performance by cluster and channel.** Horizon-specific global pooled $R_{pred, global}^{2}$for synthetic and hybrid native AR-HMM forecasts, the causal prefix-mean reference, and persistence. Rows represent the seven behavioural channels and columns the four clusters; shaded bands denote participant-block bootstrap 95% Cis.

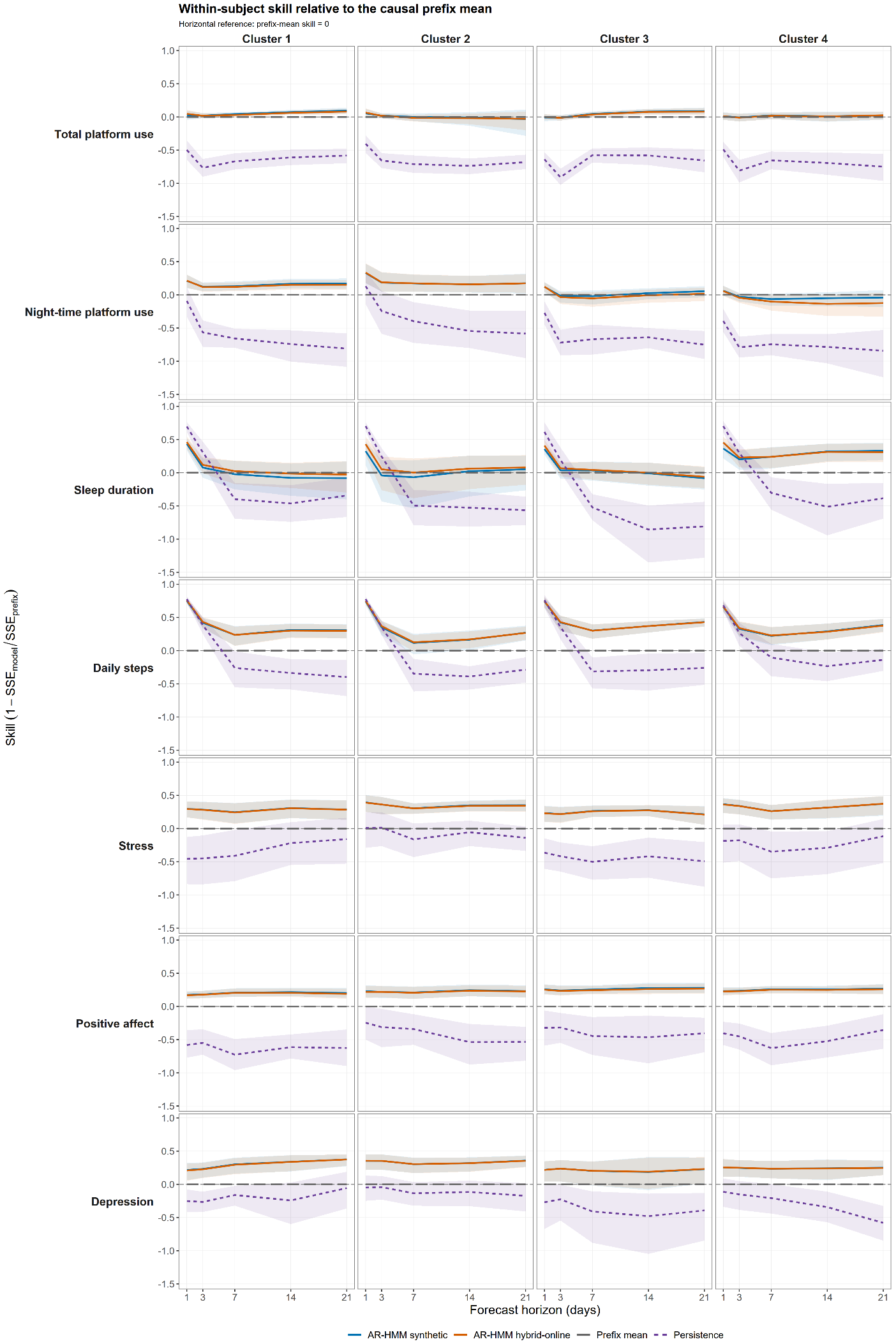

**Supplementary Figure S12.6b**. Within-subject forecasting skill by cluster and channel. Horizon-specific skill of synthetic and hybrid native AR-HMM forecasts relative to the causal participant-specific prefix mean, with persistence shown as an additional comparator. Skill was calculated as $1-SSE_{model}/SSE_{prefix}$; zero denotes equal error to the prefix-mean reference. Shaded bands denote participant-block bootstrap 95% CIs.

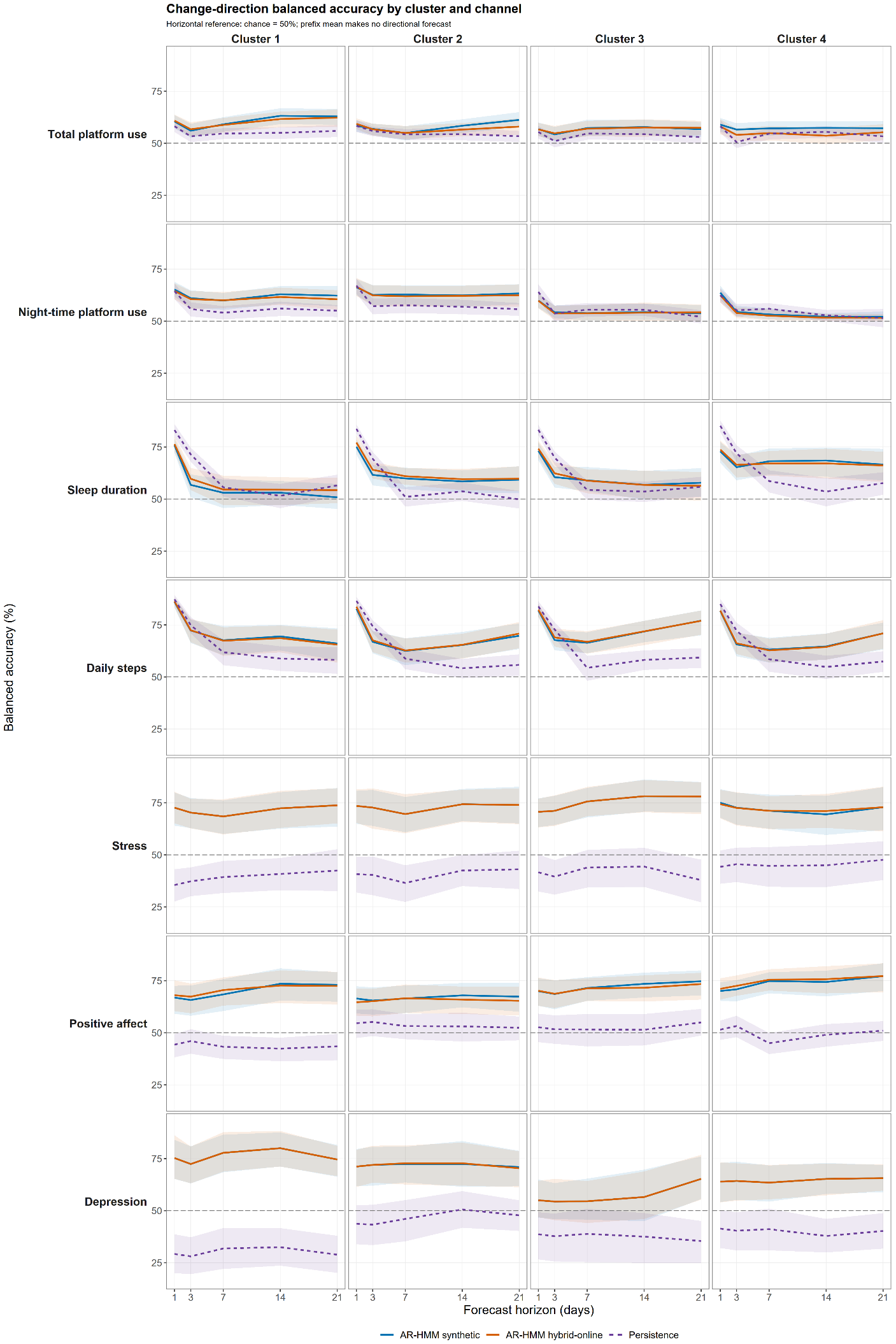

**Supplementary Figure S12.7**. Change-direction prediction by cluster and channel. Horizon-specific balanced accuracy for synthetic and hybrid native AR-HMM forecasts and persistence across the seven channels and four clusters. The horizontal reference line denotes chance performance at 50%; shaded bands denote participant-block bootstrap 95% CIs.

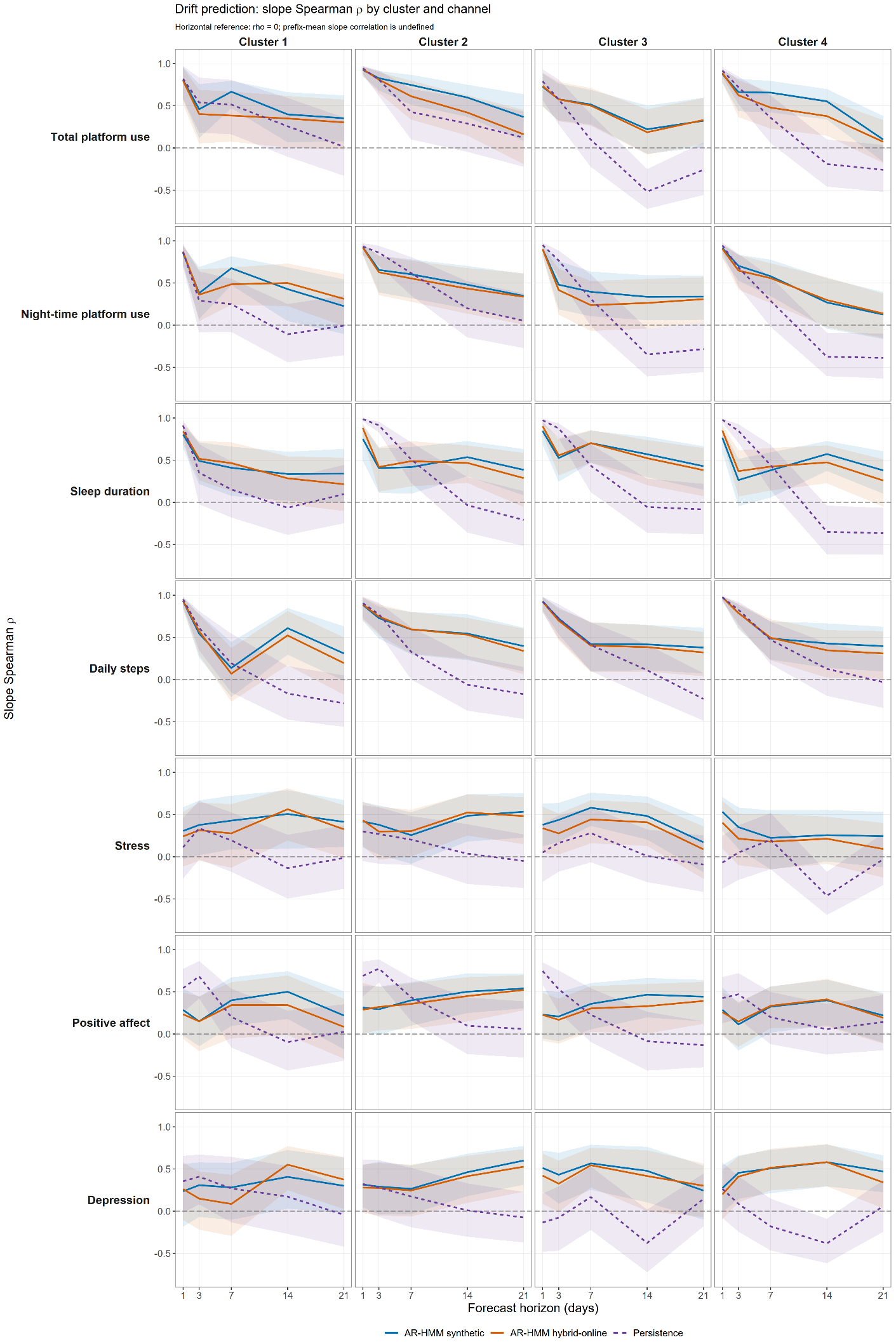

**Supplementary Figure S12.8**. Drift prediction by cluster and channel. Horizon-specific Spearman correlations between observed and predicted participant-level temporal slopes for synthetic and hybrid native AR-HMM forecasts and persistence. The horizontal reference line denotes ρ=0; shaded bands denote participant-block bootstrap 95% CIs.

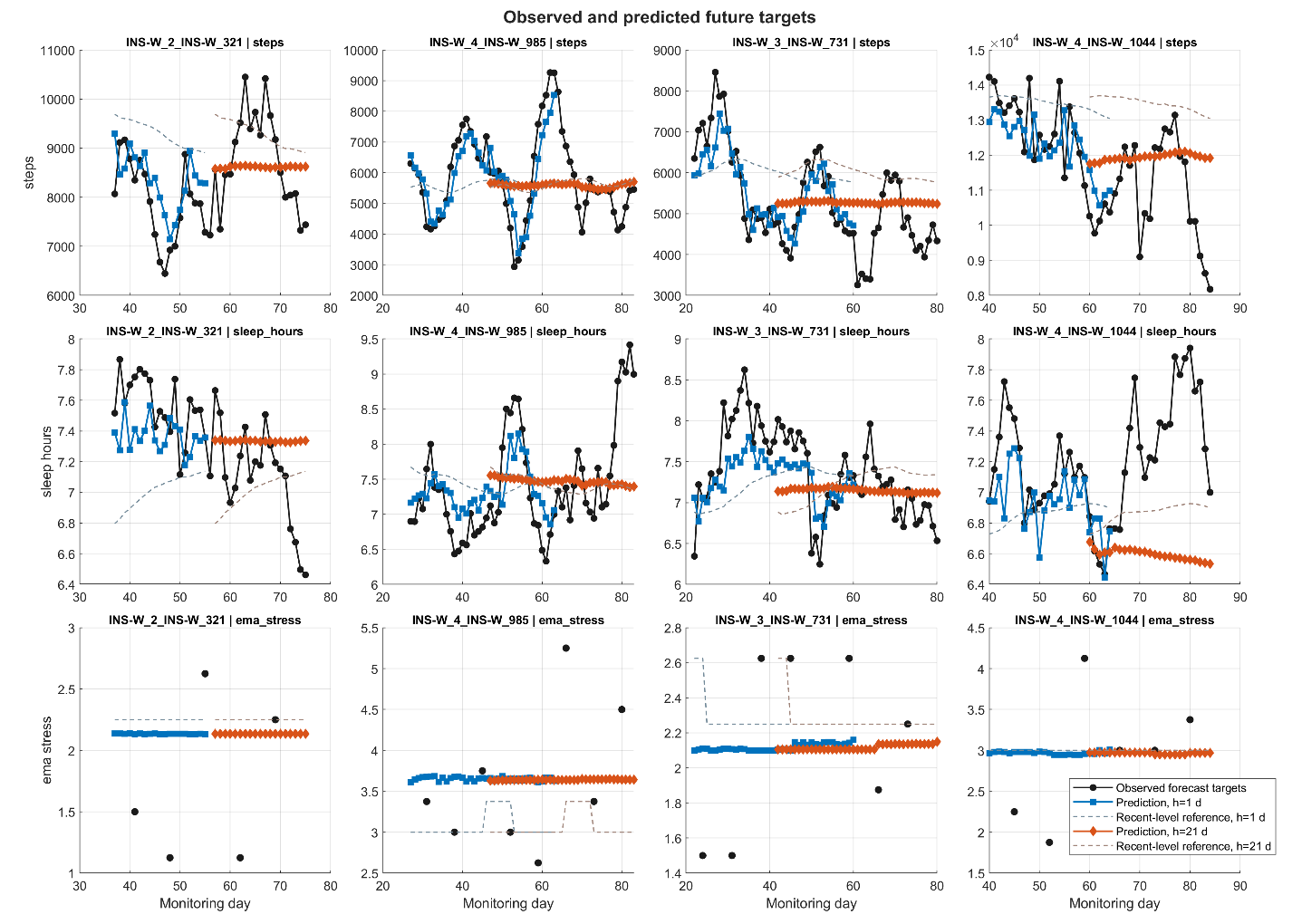

**Supplementary Figure S12.10a. Observed future targets vs. AR-HMM predictions for the h=1 and h=21 horizons in the step, sleep and EMA stress channels of the 4 centroid-close GLOBEM participants**. Black lines indicate observed targets while blue and orange lines indicate the h=1 and h=21 predictions, respectively. Dashed lines show the recent-level references for the respective time horizons.

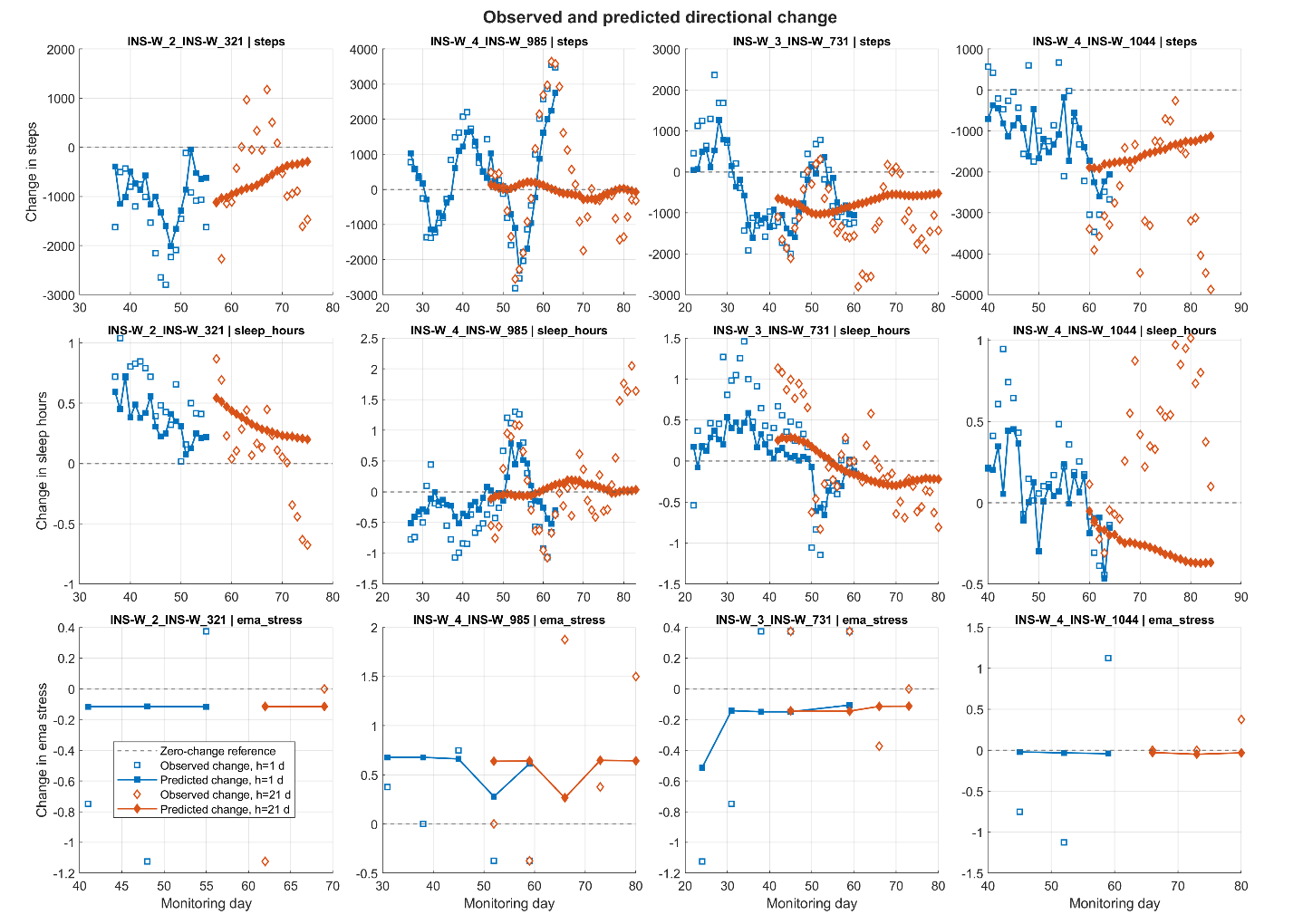
**Supplementary Figure S12.10b. Observed directional change vs. AR-HMM directional change predictions for the h=1 and h=21 horizons in the step, sleep and EMA stress channels of the 4 centroid-close GLOBEM participants**. Empty squares and diamonds indicate observed change targets while fille counterparts show the AR-HMM’s h=1 and h=21 change predictions, respectively.

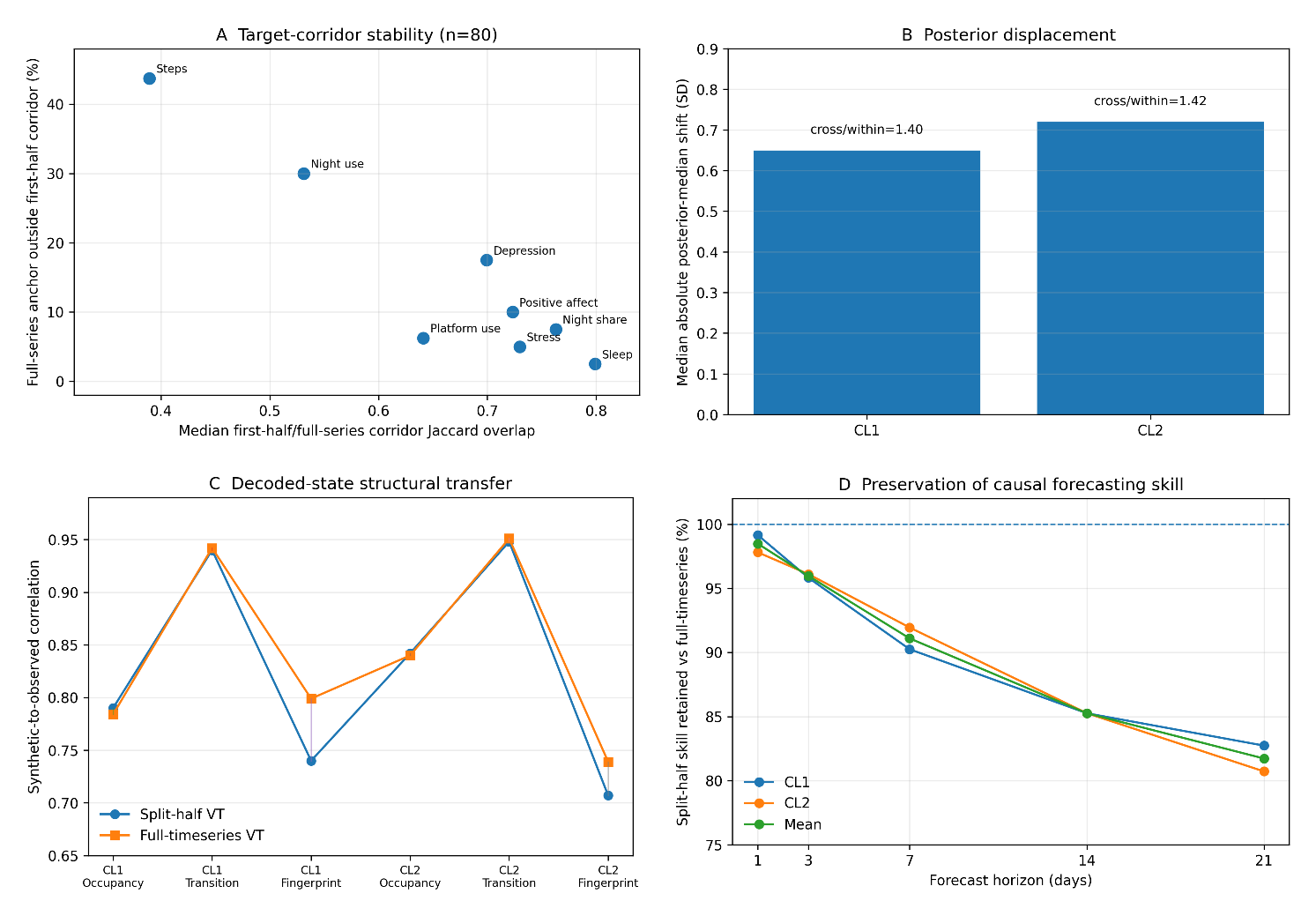

**Supplementary Figure 13. Split-half validation across calibration targets, posterior parameterization, decoded dynamics, and causal forecasting**. **Panel A**, hard-target corridor stability across the balanced 80-participant sample: higher Jaccard overlap and lower breach rate indicate greater stability. **Panel B**, median posterior displacement between matched split-half and full-series particle clouds; bar annotations show the median cross-cloud/within-cloud nearest-neighbour distance ratio. **Panel C**, synthetic-to-observed correlations for state occupancy, transition structure, and multivariate state fingerprints under split-half and full-series calibration. **Panel D**, percentage of full-series synthetic causal forecasting skill retained by split-half VTs at 1-, 3-, 7-, 14-, and 21-day horizons. The green mean line is the unweighted mean of the CL1 and CL2 retention ratios. Together, the panels show that measurable target and posterior perturbation coexisted with near-preservation of decoded transition/occupancy structure and most downstream forecasting skill.
